# Urinary steroid metabolome shows adrenal, gonadal, and neuroactive steroid dysregulation in adolescents with depression

**DOI:** 10.1101/2025.07.10.25331252

**Authors:** Lars Dinkelbach, Stefan A. Wudy, Michaela F. Hartmann, Lars Libuda, Manuel Föcker, Johannes Hebebrand, Anke Hinney, Ute Nöthlings, Ute Alexy, Corinna Grasemann, Raphael Hirtz

## Abstract

**Context and Objective:** Steroid hormone profiles in affective disorders suggest hypothalamic– pituitary–adrenal (HPA) axis dysregulation and may reveal novel therapeutic targets. However, most existing studies focus narrowly on glucocorticoids. This study aims to comprehensively characterize alterations in the steroid metabolome of adolescents with depressive symptoms.

**Design, Setting, and Patients:** This cross-sectional study analyzed the urinary excretion of 39 steroid metabolites (via gas chromatography-mass spectrometry) from 75 adolescent psychiatric patients with depressive symptoms (63 females, age 15.6 ± 1.3 years) and 75 healthy controls (64 females, age 15.3 ± 1.3 years), matched for age, sex, and pubertal status.

**Results:** Patients exhibited significantly elevated excretion rates (µg/24h) of corticosterone metabolites (median = 608.4, interquartile range (IQR): 342.4 - 1208.2 vs. controls: median = 321.0, IQR: 243.9 - 443.8), dehydroepiandrosterone (DHEA) metabolites (median = 1253.8, IQR: 569.8 - 2796.2 vs. median = 519.5, IQR: 254.0 - 1028.7), androgen metabolites (median = 6721.0, IQR: 4185.6 - 9395.8 vs. median = 3680.4, IQR: 2510.8 - 5419.0), and individual progesterone and glucocorticoid metabolites, while estradiol excretion was lower (median = 4.0, IQR: 2.9 - 5.8 vs. median = 5.8, IQR: 4.3 - 7.7). Analyses of enzyme activities via multivariate machine learning identified the tetrahydrated urinary metabolite ratio of 11-deoxycorticosterone (TH-DOC) to corticosterone metabolites as a biomarker to distinguish patients from controls (AUC = 0.800, 95%-CI [0.702 – 0.882]).

**Conclusions:** Elevated excretion rates of ACTH-dependent hormones indicate chronic stress in adolescents with depressive symptoms. The TH-DOC-to-corticosterone metabolite ratio may help identify at-risk patients or guide personalized therapies.

## Background

Unipolar depression is the single most important factor contributing to overall disease burden in adolescence and early adulthood ^1,2^. Although the pathophysiology of depression is multifaceted, involving genetic, neurobiological, environmental, and cognitive factors, growing evidence supports the role of hypothalamic-pituitary-adrenal (HPA) axis dysregulation in the development and persistence of depressive symptoms and may serve as a potential target for novel therapeutic interventions ^3,4^.

Numerous studies have investigated the relationship between steroid hormones in blood and the occurrence of major depressive disorders (MDD) in adults. A common finding is the upregulation of the HPA axis, characterized by elevated levels of cortisol and adrenocorticotropic hormone (ACTH), blunted suppression of the HPA axis in response to dexamethasone, and changes in specific enzymes involved in steroid metabolism, e.g. the 11-β-hydroxysteroid dehydrogenase activity (11β-HSD) ^3–6^.

To date, most studies of the steroid metabolism in MDD have focused on blood measurements of single steroids (e.g., cortisol) or a distinct set of steroids and their precursors, without addressing the complex interrelationships between steroids and their regulation by common enzymes and regulatory pathways. In a recent study by our group, comprehensive serum steroid profiles of 261 adolescents with depression confirmed increased cortisol levels but also suggested a broader elevation of adrenal steroids, including progesterone, 17-hydroxyprogesterone, 11-deoxycortisol, cortisone, and corticosterone ^7^. Further analyses revealed a distinct pattern of HPA-axis dysregulation, indicated by an increased corticosterone to 11-deoxycorticosterone ratio, which accurately discriminated between psychiatric patients and controls ^7^.

The latter finding also suggests changes at the level of neuroactive steroids, specifically tetrahydro-11-deoxycorticosterone (TH-DOC), a metabolite of 11-deoxycorticosterone. TH-DOC as well as the 3α reduced progesterone derivatives 5α-pregnane-3α-ol-20-one (allopregnanolone, aPo) and 5β-pregnane-3α-ol-20-one (Pregnanolone, Po), and the testosterone metabolite 5α-androstane-3α,17β-diol (3α-androstanediol, 5αAD-3α,17β) have been shown to be potent positive allosteric modulators of central γ-aminobutyric acid A (GABA_A_) receptors ^8^, with potential inhibitory, anticonvulsive, anxiolytic, and antidepressant effects ^9^. However, while allopregnanolone has been successfully developed as a medication for post-partum depression ^10–12^, the contribution of most neuroactive steroids to adolescent MDD has not yet been investigated.

While we already conducted a LC-MS/MS analysis of serum steroids ^7^ the analysis of urine samples by gas chromatography-mass spectrometry (GC-MS) allows for a more comprehensive assessment including anabolic and catabolic steroid metabolism ^13^, as well as a larger spectrum of analytes such as neuroactive steroids. Urine recovery is non-invasive and may circumvent stress-related confounding as with blood collection.

The objective of this cross-sectional observational study was to compare 39 urinary steroid metabolites (measured with GC-MS, for a complete list of metabolites and abbreviations used see Fig. 1) between 75 adolescent psychiatric patients (morning spot urine samples) with at least mild depressive symptoms (BDI-II ≥ 14, 77.3% diagnosed with MDD) and 75 healthy controls (drawn from a population based cohort, 24-hour urine samples) matched for age, sex, and pubertal status. Additionally, metabolite ratios, serving as proxies for enzyme activities, were evaluated as biomarkers to distinguish patients from controls.

**Figure 1.**
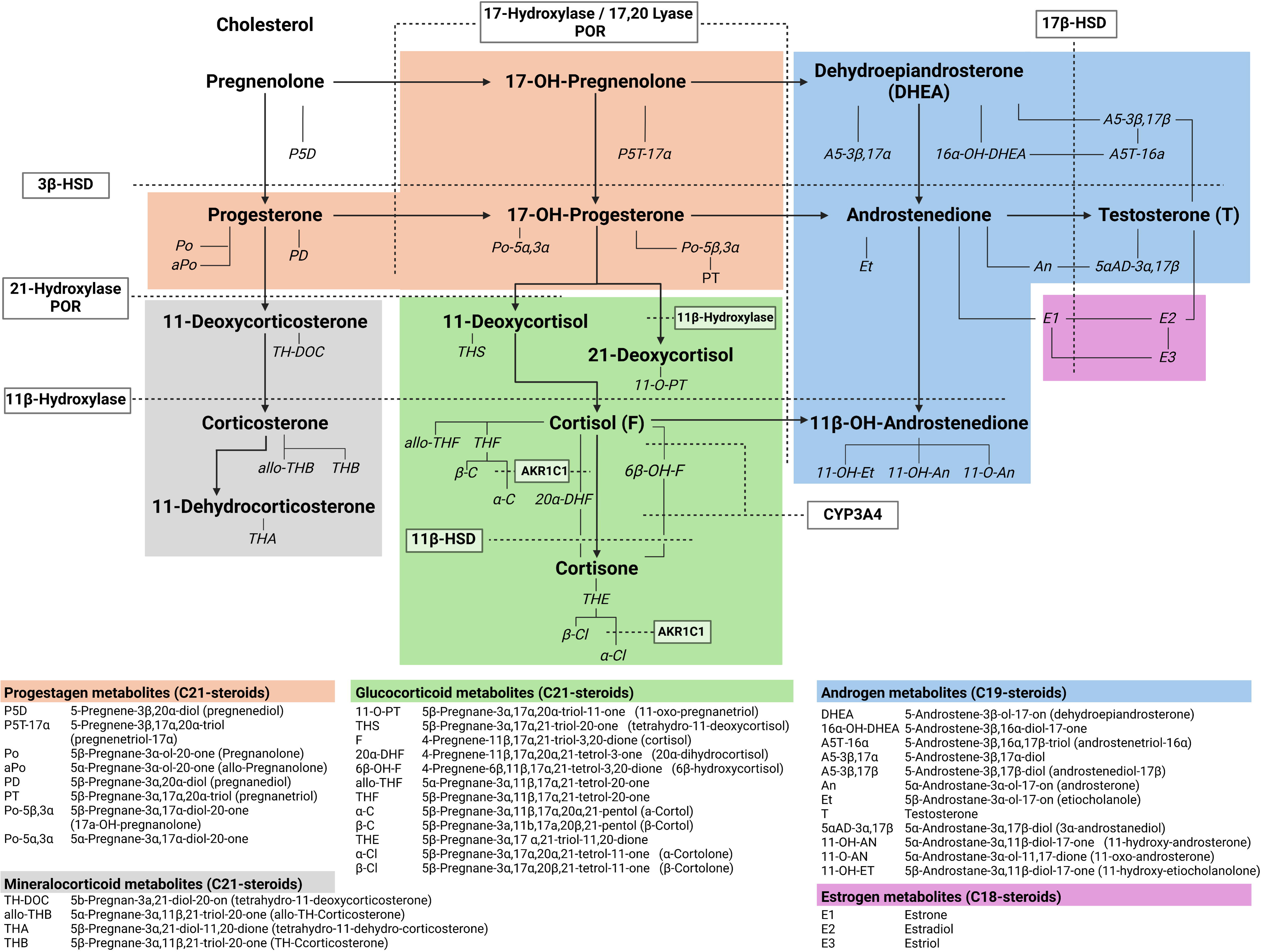
Urinary steroid metabolism pathway. This figure depicts how the steroid metabolites utilized in the current study and selected enzymes are embedded in the steroid metabolism and is not intended to give a detailed overview of the steroid biosynthesis. A more detailed description of steroid biosynthesis, which also served as the basis for this figure, can be found in the article by Ackermann, Groessl, Pruijm, Ponte, Escher, d’Uscio, Guessous, Ehret, Pechère-Bertschi and Martin ^52^. Steroids primarily found in serum are presented in bold. Urinary steroid metabolites are shown in italics. Enzymes are highlighted with a frame, the respective reactions catalyzed by the given enzymes are illustrated with dashed lines.

## Materials and methods

### Psychiatric sample

The present study utilized data derived from the baseline visit of a double-blind randomized controlled trial assessing the efficacy of vitamin D3 supplementation on depressive symptoms. The study primarily targeted patients with 25-hydroxyvitamin D3 deficiency (≤ 30nmol/l) but also included patients with sufficient vitamin D levels during the recruitment stage. Eligible participants were either adolescent inpatients or daycare patients with at least mild depressive symptoms (Inclusion criteria: age 11.0 to 18.9 years, Beck Depression Inventory-II (BDI-II) total score ≥ 14). The study was conducted between June 2016 and May 2018 at the Department of Child and Adolescent Psychiatry, Psychosomatics, and Psychotherapy at LVR-Klinikum Essen ^14,15^. In addition, patients were selected from the observational cross-sectional ’Nutrition and Mental Health study’ (September 2018 - May 2020) conducted at the same clinic which aimed to examine the relationship between nutrition and mental health. Exclusion criteria for both studies were intellectual disability or a concomitant severe somatic disease. Since the protocol for the baseline visit of both studies was identical, data were pooled.

Baseline assessments included comprehensive data collection on medical history (including medication use), anthropometrics (height and weight), and pubertal status (Tanner staging by medical staff). Additionally, morning spot-urine samples were collected and stored at -80°C. Depressive symptoms were assessed via self-report with the Beck Depression Inventory II (BDI-II ^16,17^), which employs 21 Likert-scaled items (0 to 3) to assess the severity of depressive symptoms according to the criteria of the Diagnostic and Statistical Manual of Mental Disorders-IV (DSM-IV). Total depressive symptoms scores were classified as mild (14 - 19), moderate (20 - 28), or severe (> 28, Tab 1.) ^16^. Psychiatric diagnoses were established by either semi-structured interviews with the Kiddie Schedule for Affective Disorders and Schizophrenia for School Aged Children, Present-Lifetime version (K-SADS-PL) according to DSM-IV criteria (N = 68 cases, 90.7%) or, in case of missing K-SADS-PL evaluations, by clinical assessment according to the International Statistical Classification of Diseases and Related Health Problems 10 (ICD-10) (N = 7 cases, 9.3%).

Since the present study aimed to validate and expand upon findings from our previous study on the serum steroid profile in depressed adolescents ^7^, particularly concerning TH-DOC and further neuroactive steroids, patients with the highest 11-corticosterone to 11-deoxycorticosterone ratio were chosen for detailed urinary steroid metabolome analysis. Given the resource-intensive nature of the urinary analyses, the present study was limited to 75 adolescents from this clinical sample. All clinical subjects were of normal weight (z-BMI: -1.96 to +1.96 SDS).

### Control Cohort

Data from control subjects were retrieved from the Dortmund Nutritional and Anthropometric Longitudinally Designed (DONALD) study, a longitudinal observational study investigating diet, growth, and metabolism of healthy children from infancy to adulthood ^18^. Data collection included comprehensive information on the participants’ health, medication, anthropometric measures, pubertal status (Tanner staging via medical examination), and 24-hour urine samples. The period over which samples were collected was documented, and the volumes of the samples were adjusted to account for minor deviations from an exact 24-hour collection period. 24-hour urine samples were immediately frozen at home after each micturition (at -12°C to -18°C degrees) and stored at -18°C to -22°C. For aliquot preparation, samples were thawed overnight, stirred, and refrozen at -18°C to - 22°C until analysis.

Control subjects were matched on the individual level as closely as possible to patients by Tanner stage, age, and sex. All controls were of normal weight (z-BMI: -1.96 to +1.96 SDS) and free of endocrine or psychiatric diagnoses.

### Steroid mass spectrometry

Steroids in 24-hour urine specimens were quantified using gas chromatography-mass spectrometry (GC-MS) analysis, which was performed at the Steroid Research & Mass Spectrometry Unit, Pediatric Endocrinology & Diabetology, Justus Liebig University Giessen. The analytical technique is described in detail elsewhere ^13,19^. In brief, free and conjugated steroids were extracted by solid phase extraction from 2,5mL of urine. Conjugates were enzymatically hydrolyzed followed by recovery of the hydrolyzed steroids by a second solid phase extraction step. Known amounts of internal standards (5α-androstane-3α,17α-diol and stigmasterol) were added to each extract before formation of methyloxime-trimethylsilyl ethers. GC was carried out on an Optima-1 MS fused silica column (Macherey-Nagel, Düren, Germany) housed in an Agilent Technologies 6890 series GC equipped with an Agilent 7683 series injector that was directly interfaced to an Agilent Technologies 5975 inert XL mass selective detector. Carrier gas was helium.

### Data processing

A total of 39 primary metabolites were analyzed (for an overview, see Fig. 1). Twelve metabolites yielded results below the lower limit of detection (LLD) in some subjects (see Supplemental Tab. S1). Values below the LLD were imputed using the R package zComposition (version 1.5.0-3 ^20^) which allows for a robust imputation of left-censored data. To compare the psychiatric sample (spot urine) with controls (24-hour urine), steroid concentrations (µg/l) were converted to estimated 24-hour steroid excretion rates (µg/24h) with the following formula:

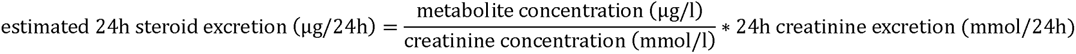

For the psychiatric sample, urinary 24-hour creatinine excretion rates for each patient were derived from normative data, normalized for height and sex ^21^. For control subjects, 24-hour creatinine excretion rates were calculated based on the measured creatinine concentration and the collection volume. The 24-hour creatinine excretion rates did not differ between patients and controls (Tab. 1). The BMI (kg/m²) for all subjects was z-transformed based on reference tables for German children and adolescents, stratified by age and sex (RefCurv version 0.4.4, ^22^).

**Table 1.**
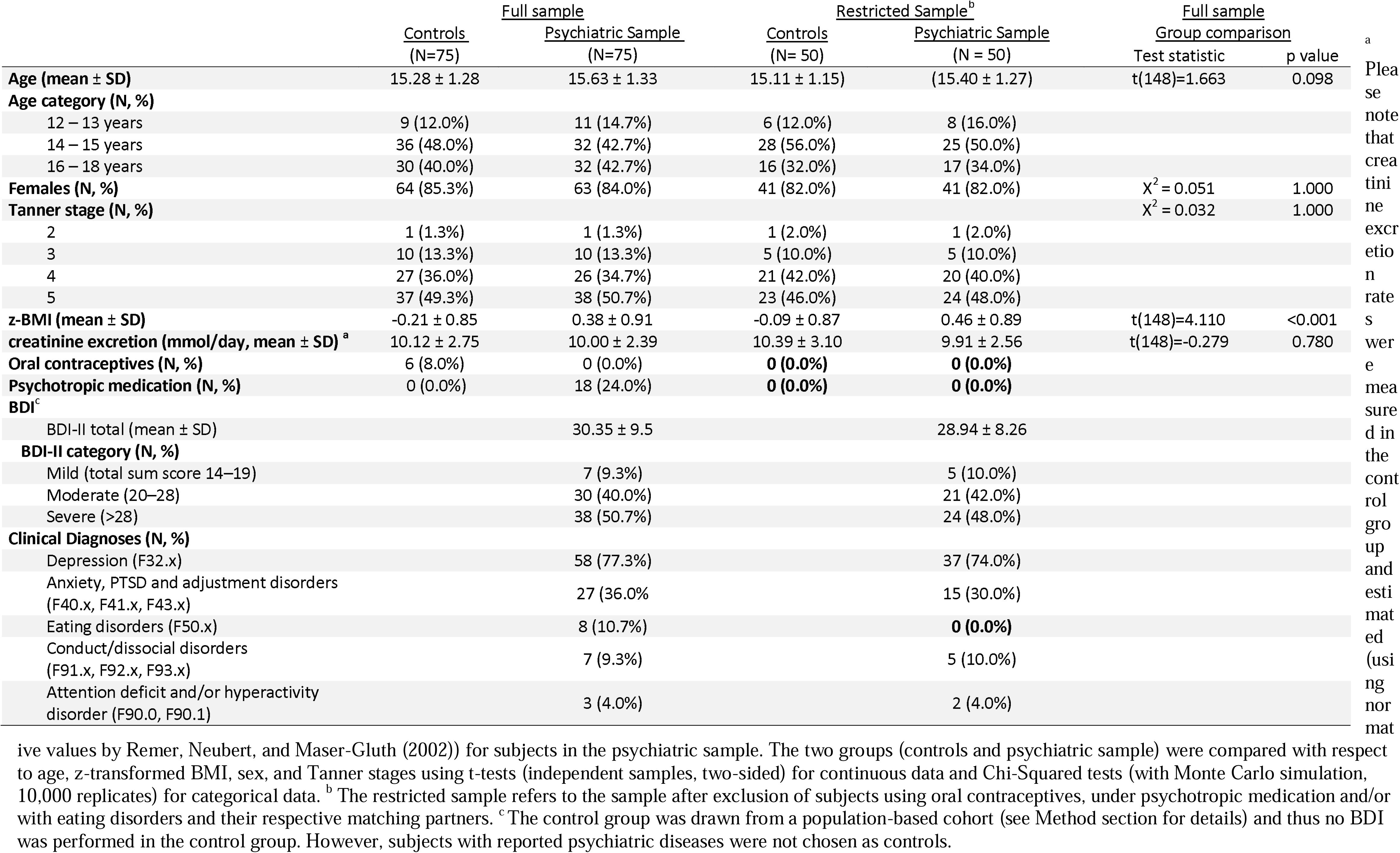
Sample characteristics.

### Statistical analyses

#### Group comparisons

Individual metabolites, combined metabolites (i.e. the sum of selected metabolites, see Supplemental Tab. 2 for an overview and formulas used), and ratios of selected metabolites (Tab. 2) were compared between the psychiatric sample and controls. Given the right-skewed distribution with severe outliers of many metabolites (see Supplemental Fig. S1-21 for boxplots of metabolites, sums and ratios), and considering that many metabolites did not follow a normal distribution even after log_10_- or ln-transformation (as identified by Shapiro-Wilk tests and QQ plots), rank-based analysis were performed using the R package Rfit (version 0.24.6, ^23^). Despite efforts to match controls to patients, a perfect match was not achieved for age (at the individual level) and z-transformed BMI (Tab. 1). Therefore, rank-based linear regression models for each outcome parameter (individual metabolites, combined metabolites, ratios) were calculated, including age and BMI (z-transformed based on reference tables) as independent variables. To facilitate interpretation of effect sizes in the absence of established estimates for rank-based linear regression models, unadjusted Wilcoxon rank sum tests were conducted (using the R package stats). Effect sizes were estimated as r-values based on the Wilcoxon test statistic (by the R package rcompanion, version 2.4.35 ^24^). These r-values were subsequently converted to Cohen’s d ^25^, and classified as small, medium, or large according to the following criteria: |d| ≥ 0.2, |d| ≥ 0.5 and |d| ≥ 0.8, respectively ^26^. The p-values for the primary outcome (group effect for metabolites, sums, and ratios) were corrected for multiple comparisons using the false discovery rate (FDR, two-tailed, R package stats ^27^) and considered significant at *p* < 0.05. Considering the assumption for power calculations (details are given in the Supplemental Material), analyses had sufficient power (1 – beta-error=0.8) to detect a small to medium effect size of d > 0.47. All data analyses were conducted with IBM SPSS Statistics 29.0 (International Business Machines Corporation, Armonk, USA) or with R version 4.3.2.

**Table 2.**
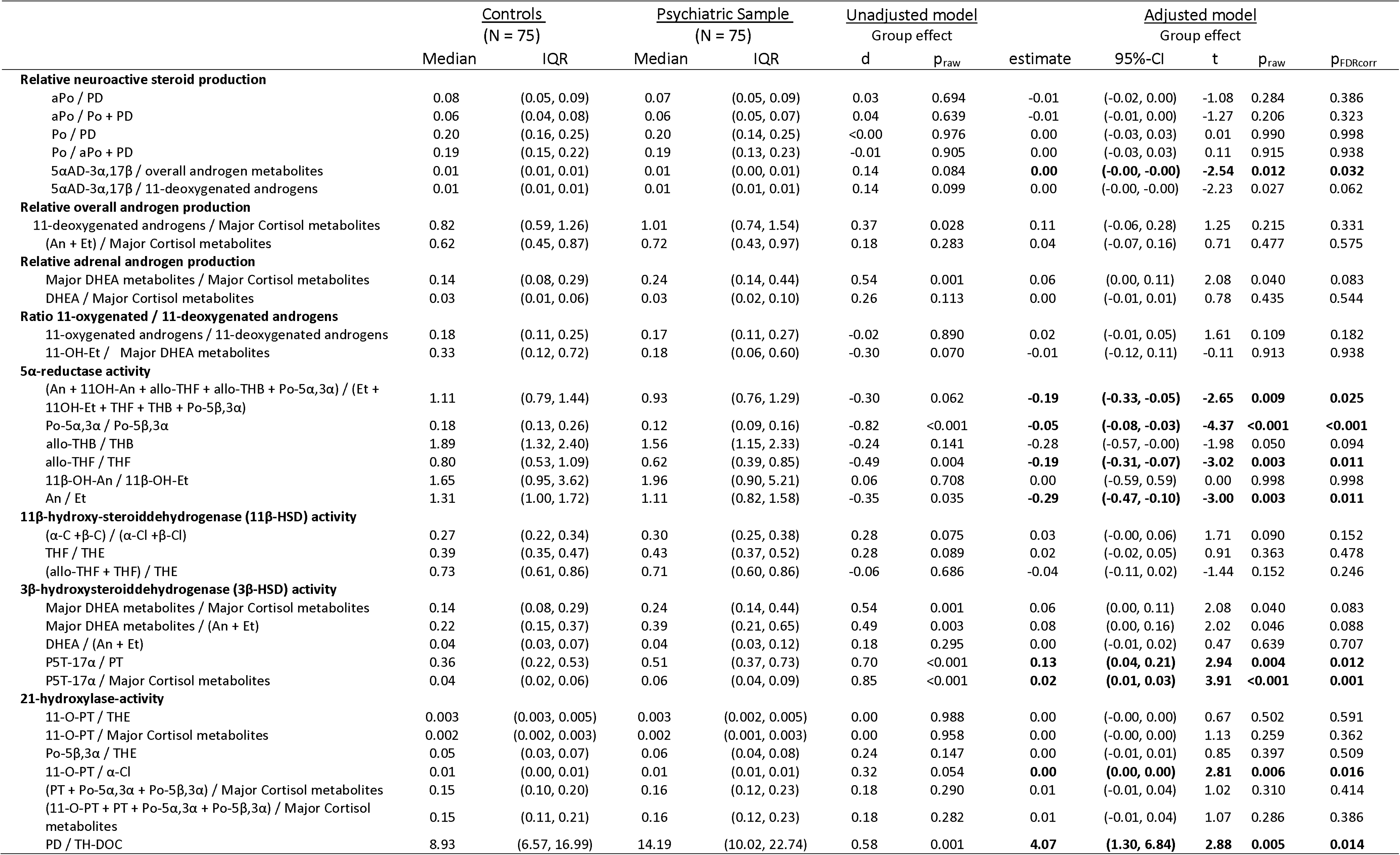
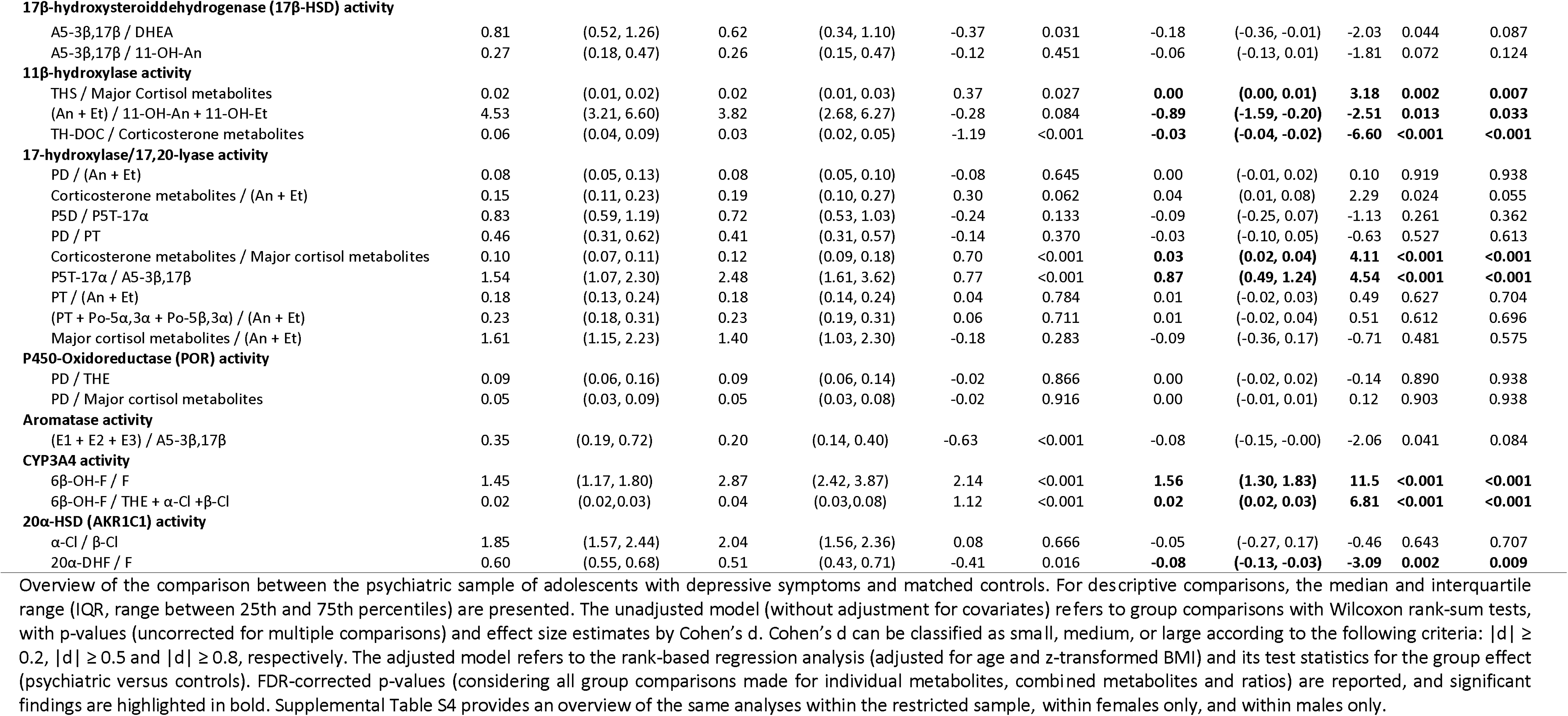
Full sample: Group comparisons of metabolite ratios as measures of enzyme activity.

### Classification

The accuracy of the urine metabolome to discriminate between the psychiatric patients and controls was assessed by steroid ratios, as they are less susceptible to sampling conditions, particularly diurnal steroid fluctuations ^28^. To determine if analyzing multiple ratios simultaneously enhances biomarker performance and to identify particularly informative single ratios, multivariate receiver operating characteristic (ROC) curves were constructed using a multivariate random forest classifier across six models, ranging from two to all available ratios. The ratios of the best performing multivariate model (based on the AUC) were then followed in univariate ROC analyses. To ensure robustness of ROC analyses, cross validation was performed. For multivariate ROC analyses, Monte-Carlo cross validation with balanced sub-sampling was performed (for details, see Supplemental Material). For univariate ROC analysis, a two-fold cross-validation was performed by dividing the sample into a test (N = 80) and verification sample (N = 70) while preserving the 1:1 matching of patients and controls. For univariate ROC curves, an optimal cut-off point was identified as the point on the ROC curve closest to the top-left corner. This point was defined by the minimum value for (1 - sensitivities)² + (1 - specificities)². The AUC was converted to Cohen’s d according to Rice and Harris ^29^. ROC analyses were conducted using the MetaboAnalyst R package (version 4.0.0 ^30^).

### Sensitivity analyses

To exclude an effect of oral contraceptives, psychotropic medication (antidepressants, neuroleptics, or stimulants) or a concomitant eating disorder on steroid metabolism, a restricted sample (n = 50 in each group) was defined. In additional sensitivity analyses, primary analyses were reconducted in females and males only to consider sex-specific effects. Robustness of the main findings of the ROC analyses was further evaluated in two subsamples: (1) those with at least mild depressive symptoms (BDI – II ≥ 14) *and* a confirmed clinical diagnosis of depression, and (2) those with values above the LLD. Each of these sensitivity analyses was conducted while maintaining the 1:1 matching on the individual level. In addition, cross-validation of multivariate and univariate ROC analyses was performed (see Supplemental Methods for details), yielding to similar results as in the primary analyses (Tab. 3). To determine whether storage time (i.e., time from sample collection to sample analysis) influenced our results, Spearman’s rank correlation coefficients were calculated between storage time and metabolite excretion rates across the entire sample. Here, a negative correlation was found between 6β-OH-F and storage time (ρ = -0.320, p < 0.001). Since none of the other metabolites tested exhibited a significant association with storage time (Supplemental Tab. S5), and the long-term stability of most urinary analytes under similar storage conditions ^31^, this finding is of unknown significance. Although group differences overshadowed this finding (see Supplemental Fig. 22), 6β-OH-F-associated ratios were excluded from multivariate ROC analyses to avoid introducing potential (albeit minor) bias.

**Table 3.** Univariate ROC analyses of selected ratios.

|  | Test sample (N = 80) |  |  |  |  |  | Verification sample (N = 70) |  | Cross validation |
| --- | --- | --- | --- | --- | --- | --- | --- | --- | --- |
|  | AUC | 95%-CI | Cohen's d | Optimal cut-off | Sensitivity | Specificity | AUC | 95%-CI | p-value<sup>a</sup> |
| **11β-hydroxylase activity (11β-hydroxylase 3)**<br>TH-DOC / Corticosterone metabolites | 0.800 | (0.702,0.882) | 1.19 | 0.0432<sup>b</sup> | 0.750 | 0.725 | 0.789 | (0.658,0.890) | 0.858 |
| **17-hydroxylase/17,20-lyase activity (17-hydroxylase 5)**<br>Corticosterone metabolites / Major cortisol metabolites | 0.698 | (0.571,0.797) | 0.73 | 0.109 | 0.600 | 0.825 | 0.688 | (0.565,0.810) | 0.914 |
| **3β-HSD activity (3β-HSD 5)**<br>P5T-17α / Major Cortisol metabolites | 0.755 | (0.649,0.858) | 0.98 | 0.0449 | 0.800 | 0.625 | 0.693 | (0.558,0.810) | 0.499 |
| **5α-reductase activity (5α-reductase 2)**<br>Po-5α,3α / Po-5β,3α | 0.736 | (0.618,0.844) | 0.89 | 0.120 | 0.600 | 0.775 | 0.716 | (0.585,0.837) | 0.741 |
| **20α-HSD (AKR1C1) activity (AKR1C1 2)**<br>20α-DHF / F | 0.639 | (0.526,0.767) | 0.50 | 0.569 | 0.675 | 0.650 | 0.581 | (0.422,0.725) | 0.529 |
| **17-hydroxylase/17,20-lyase activity (17-hydroxylase 6)**<br>P5T-17α / A5-3β,17β | 0.690 | (0.584,0.801) | 0.70 | 2.350 | 0.575 | 0.750 | 0.727 | (0.584,0.826) | 0.710 |
| **3β-HSD activity (3β-HSD 4)**<br>P5T-17α / PT | 0.707 | (0.581,0.812) | 0.77 | 0.386 | 0.800 | 0.550 | 0.695 | (0.560,0.803) | 0.911 |
| **Aromatase**<br>(E1 + E2 + E3) / A5-3β,17β | 0.679 | (0.547,0.782) | 0.66 | 0.228 | 0.625 | 0.700 | 0.670 | (0.520,0.794) | 0.969 |
| **17-hydroxylase/17,20-lyase activity (17-hydroxylase 2)**<br>Corticosterone metabolites / (An + Et) | 0.509 | (0.399,0.650) | 0.03 | 0.173 | 0.600 | 0.575 | 0.659 | (0.535,0.785) | 0.114 |
| **21-Hydroxylase activity (21-hydroxylase 7)**<br>PD / TH-DOC | 0.661 | (0.540,0.773) | 0.61 | 16.0 | 0.525 | 0.700 | 0.658 | (0.525,0.792) | 0.966 |

Overview of the test properties of the top ten ratios identified by multivariate ROC analysis (ranked by their average importance). For two-fold cross-validation, the sample was split in a test and in a validation sample. <sup>a</sup> DeLong's test for the comparison of two ROC curves (Null hypothesis: The true difference between both AUC is equal to zero) were calculated. <sup>b</sup> For the TH-DOC/corticosterone metabolite ratio, an additional optimal cut-off was found (0.0399, Sensitivity = 0.725, Specificity = 0.750). However, in the analysis in the full-sample, only the optimal cut-off of 0.0432 was replicated.

### Ethics

All studies included in the current analysis have been approved by the local ethics committee of the Medical Faculty of the University Duisburg-Essen (15-6363-BO) or of the University of Bonn (project identification of the most recent version: 185/20). Both studies were conducted in accordance with the Declaration of Helsinki. Written informed consent was obtained from all parents (or the respective legal guardians) to participate in the study.

## Results

### Sample Characteristics

To compare the psychiatric sample (spot urine) with controls (24-hour urine), steroid concentrations (µg/l) were converted to estimated 24-hour steroid excretion rates (µg/24h) under consideration of 24-hour creatinine excretion rates (see Method section for details). The psychiatric sample and matched controls did not differ significantly in terms of 24-hour creatinine excretion rates, age, sex, and Tanner stages (for demographic and clinical characteristics, see Tab. 1). However, there was a significant difference in z-transformed body mass index (BMI; psychiatric sample: 0.38 (± 0.91), control group: -0.21 (± 0.85)).

### Group Comparison of Steroid Urine Metabolites

Descriptively, the urinary steroid metabolome of psychiatric subjects showed increased daily excretion rates for most steroid metabolites compared to controls, except for the 11-deoxycorticosterone metabolite TH-DOC and estrogen metabolites, which exhibited decreased excretion rates (Fig. 2; for details, see Supplemental Tab. S2). In rank-based linear regression models, considering age and z-transformed BMI as covariates and after correction for multiple comparisons, 25 of the 46 individual and combined metabolites exhibited significant differences between patients and controls.

**Figure 2.**
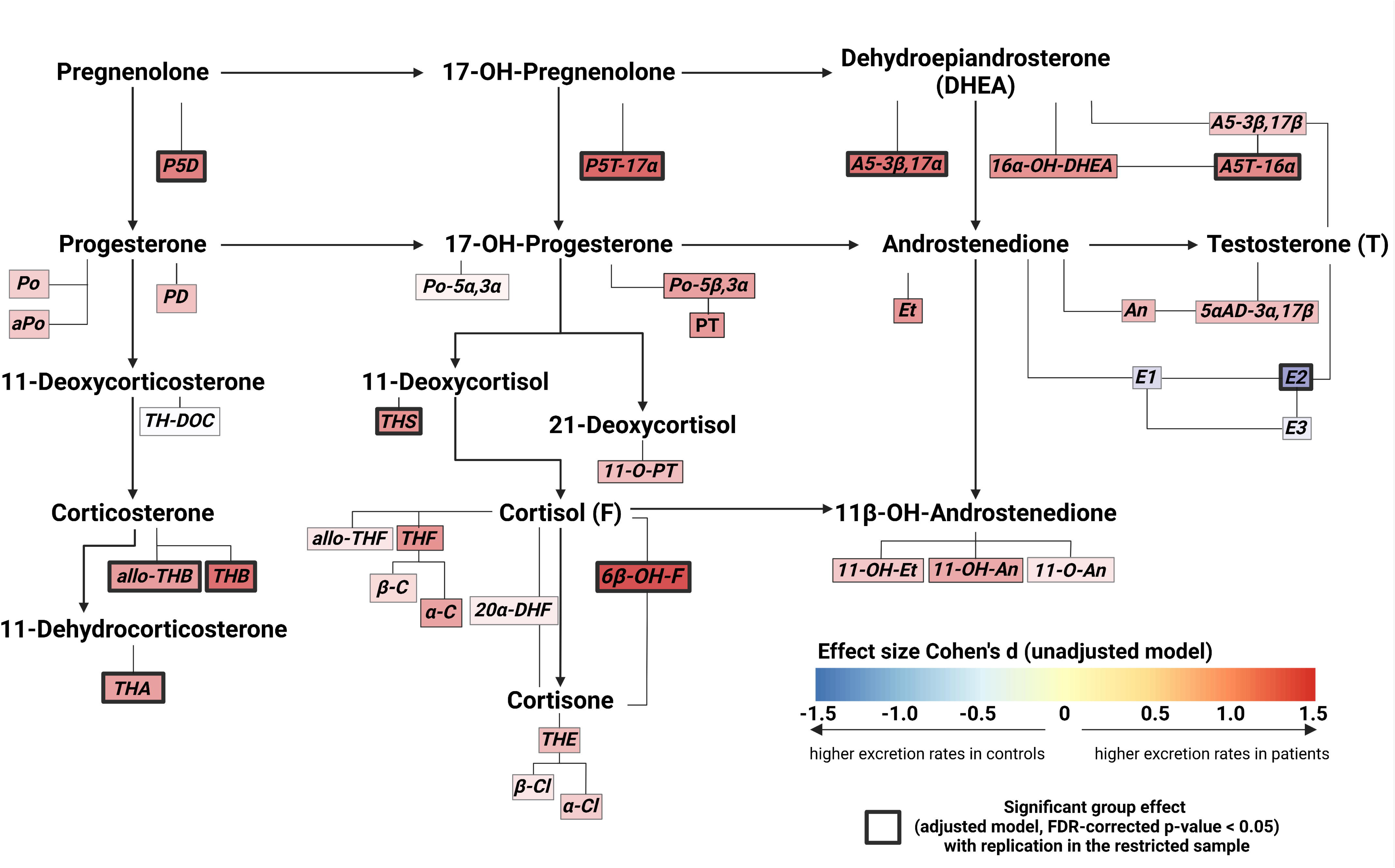
Illustration of the group effects (controls versus psychiatric sample) of daily excretion rates (µg/24h) of single steroid metabolites. The heatscale underlying each metabolite represents effect size estimate (Cohen’s d) of the unadjusted model (Wilcoxon rank sum tests). The effect size estimate d can be classified as small, medium, or large according to the following criteria: |d| ≥ 0.2, |d| ≥ 0.5 and |d| ≥ 0.8, respectively ^25,26^. Please note that the effect sizes for the urinary free daily excretion rates of cortisol (F), dehydroepiandrosterone (DHEA) and testosterone (T) are not illustrated. Comparisons with a (false discovery rate, FDR) corrected p-value < 0.05 for the group effect in the adjusted analysis (rank-based regression analyses, adjusted for the confounders age and z-transformed BMI, see Supplemental Tab. S2) which could be replicated in the sensitivity analysis with the restricted sample (see Supplemental Tab. S3) are highlighted with a bold frame. A full list of the abbreviations used for the metabolites is given in Fig. 1.

Notably, all corticosterone metabolites (allo-THB, THA, and THB) and most progestogen metabolites (P5D, P5T-17α, PT, and Po-5β,3α) demonstrated significant group effects (Supplemental Tab. S2, Supplemental Fig. 1 and 2). With regard to metabolites of glucocorticoids and their precursors, patients exhibited a Cushing-like constellation with elevated excretion rates of cortisol (THF, α-C and 6β-OH-F) and 11-deoxycortisol (THS) metabolites. Additionally, excretion rates were increased for free cortisol (F) and the 21-deoxycortisol metabolite 11-O-PT (Supplemental Tab. S2). Combined cortisol metabolites did not differ significantly between groups (major cortisol metabolites allo-THF + THF + THE: p_FDR-corrected_ = 0.073, all cortisol metabolites: p_FDR-corrected=_ 0.121).

Patients also exhibited higher daily excretion rates for combined total androgen metabolites, 11-oxygenated and 11-deoxygenated androgens, and total DHEA metabolites (Supplemental Tab. 2). Significant effects for individual metabolites included the DHEA metabolites 16α-OH-DHEA, A5-3β,17α, and A5T-16α, the androstenedione metabolite etiocholanolone (Et), as well as the 11-oxygenated metabolites 11-OH-Et and 11-OH-An. Regarding estrogens, estradiol (E2) excretion rates were significantly lower in patients compared to controls.

Sensitivity analyses: A restricted subsample was created by excluding all subjects with oral contraception (N = 6, control sample only) and/or patients with psychotropic medication (N = 18, psychiatric sample only) and/or with a concomitant eating disorder (N = 8, psychiatric sample only) and their respective 1:1 matched partner, leading to a restricted sample with N = 50 patients and N = 50 controls. Reanalysis of group comparisons using the restricted sample confirmed 11 out of the 25 previously significant findings (Supplemental Tab. 3). Notably, androgens were the main contributors to differences from the primary analyses: The effect estimates for combined androgens (DHEA metabolites, 11-oxygenated androgens, 11-deoxygenated androgens and overall androgen metabolites), the DHEA-metabolite 16α-OH-DHEA, the androstenedione metabolite etiocholanolone (Et), as well as the 11-oxygenated metabolites 11-OH-Et and 11-OH-An were smaller in the restricted sample, suggesting confounding. Similarly, the significant group effects seen for urinary free cortisol (F) and for the glucocorticoid metabolites α-C, THF, and 11-OPT as well as for the progestagene metabolites PT and Po-5β,3α were not replicated in the restricted sample. To assess sex-specific effects, analyses were conducted separately for females and males. All eleven individual and combined metabolites that exhibited a group effect in the restricted sample also demonstrated a significant group effect in females-only analyses. Of these, only seven showed a significant group effect in males-only analyses. However, due to the small sample size in the males-only group (Tab. 1), it is unclear whether these findings reflect female-specific effects or are attributable to reduced statistical power.

### Ratios as Indicators of Enzyme Activity

Rank-based regression analysis demonstrated significant differences between the total control group and the total psychiatric sample in 17 out of 54 enzyme activity ratios, indicating different metabolic processing in the psychiatric sample (see Tab. 2 for all formulas and ratios analyzed, for boxplots see Supplemental Fig. S7-S21). These ratios include:

5α-reductase activity: Ratio of all 5α reduced metabolites (sum of An, 11OH-An, allo-THF, allo-THB, and Po-5α,3α) to their precursors (sum of Et, 11OH-Et, THF, THB, and Po-5β,3α), the ratio of Po-5α,3α to Po-5β,3α, the ratio of allo-THF to THF and the ratio of An to Et.

Relative neuroactive steroid production: Ratio of 5αAD-3α,17β to the sum of all androgen metabolites.

3β-HSD activity: Ratio of P5T-17α to both the sum of major cortisol metabolites and to PT.

21-Hydroxylase activity: Ratios of 11-O-PT to α-Cl and of PD to TH-DOC.

11β-Hydroxylase activity: Ratio of THS to the sum of major cortisol metabolites, ratio of the sum of An and Et to their 11-hydroxylased counterparts (sum of 11-OH-An and 11-OH-Et) and the ratio of TH-DOC to the sum of corticosterone metabolites.

17-Hydroxylase/17,20-lyase activity: Ratios of the sum of corticosterone metabolites to the sum of major cortisol metabolites, and of P5T-17α to A5-3β,17β.

CYP3A4 activity: Ratios of 6β-OH-F to F and 6β-OH-F to the combined THE + α-Cl + β-Cl levels.

AKR1C1 activity: Ratio of 20α-DHF to F.

Sensitivity Analyses: In both the restricted sample and the females-only sample, findings for 15 out of 17 ratios from the primary analyses were confirmed. Exceptions included the ratios of 5αAD-3α,17β to the sum of androgen metabolites and 11-OPT to α-Cl (Supplemental Tab. S4). Notably, males-only but not females-only analyses showed significant group effects for the majority of the remaining 21-hydroxylase activity ratios (see Supplemental Tab. S4 and Supplemental Fig. S14), suggesting a male-specific effect.

### Biomarker Performance: ROC Curves

Multivariate ROC analysis with random forest classifier utilizing ratios of steroid metabolites were performed to discriminate between the psychiatric sample and matched controls. The multivariate ROC analysis incorporating 26 ratios yielded the highest discriminatory performance (area under the curve [AUC] = 0.827, 95% CI [0.757 - 0.907]; Cohen’s d = 1.33, average accuracy 74.3%). However, since the model with ten ratios showed a similarly strong performance (AUC = 0.820, 95% CI [0.755 - 0.893]; Cohen’s d = 1.29, average accuracy 74.4%), these ten ratios were further analyzed (Fig. 3 and Tab. 3). In univariate ROC analysis, nine of these ten ratios significantly performed above random classification (AUC = 0.5), exhibiting effect sizes (Cohen’s d) from 0.03 to 1.19.

**Figure 3.**
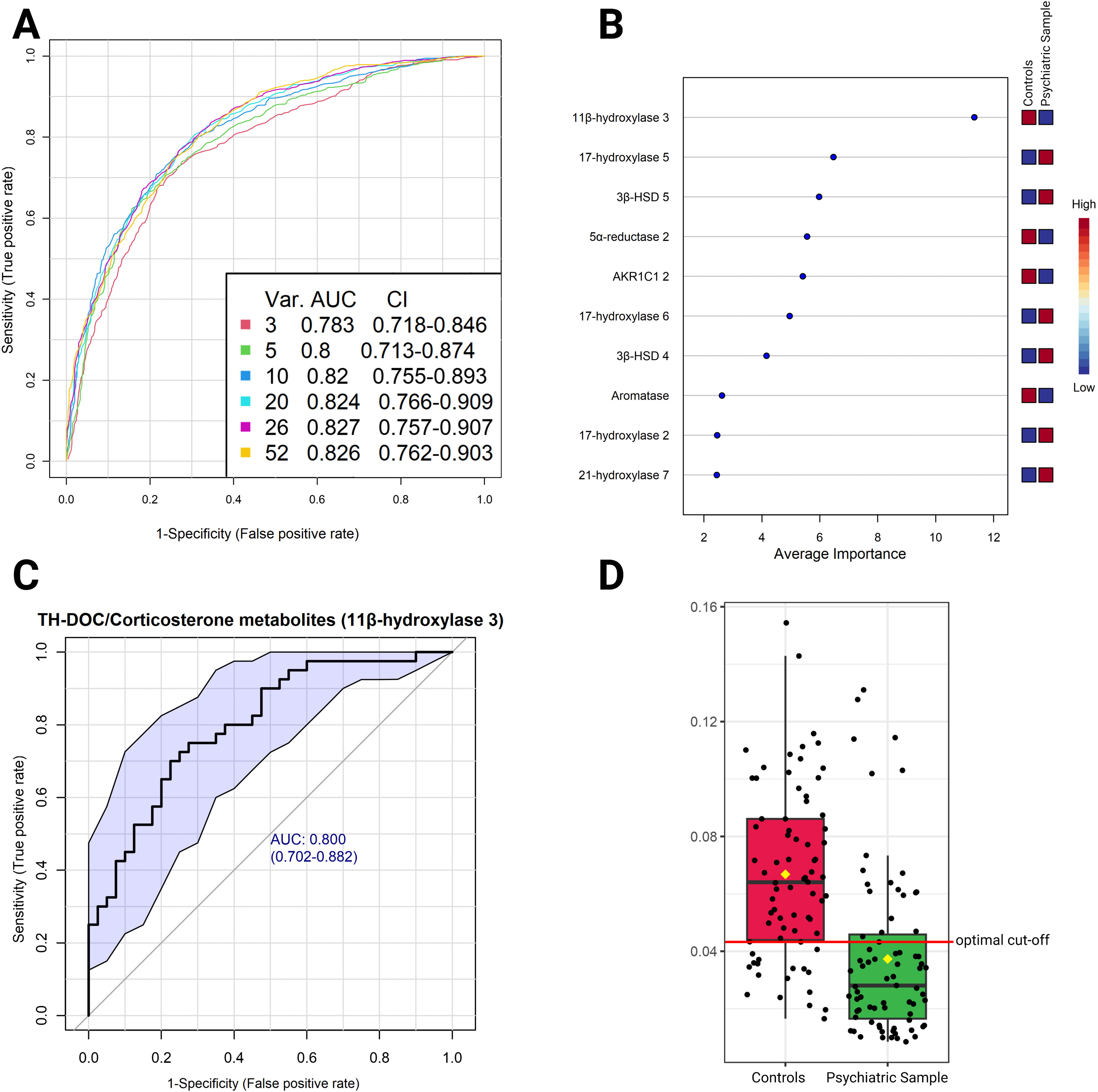
Receiver operatic characteristic (ROC) curves of the performance of urinary metabolites to distinguish between the control group and the psychiatric sample. **A:** Curves of multivariate ROC analyses using a random forest classifier. The area under the curve (AUC) as well as the 95% confidence intervals (CI) for models based on three up to 52 ratios are provided. With more than ten ratios, the incorporation of supplementary ratios into the model yielded only marginal enhancements in model performance. **B:** The ten ratios contributing to the model with ten outcome variables (AUC = 0.820, 95%-CI [0.755 - 0.893]), arranged according to their average importance. Red tiles (in comparison to blue tiles) indicate that high values of the respective outcome led to a classification into the respective group (control group / psychiatric sample). For reasons of presentability, ratios were abbreviated according to their order of appearance in Tab. 2 (e.g. 17-hydroxylase 5 = Corticosterone metabolites / Major Cortisol metabolites). **C:** Univariate ROC analysis (test-sample only) of the TH-DOC / Corticosterone metabolites ratio (as an indicator for 11β-hydroxysteroiddehydrogenase activity). **D:** Boxplot (full sample) comparing TH-DOC / Corticosterone metabolites ratio between groups. ROC-analyses identified an optimal cut-off point at a ratio of 0.0432.

Sensitivity Analyses: Reconducting ROC analyses in the restricted sample, as well as in a subsample comprising individuals with a psychiatric diagnosis of MDD, confirmed the results (Supplemental Fig. S24).

## Discussion

Previous studies investigating the HPA-axis in children and adolescents with depression have focused on individual steroids (e.g., cortisol) or selected subsets of steroids. In contrast, the present study provides a comprehensive analysis of the urinary steroid metabolome (measured by GC-MS) in depressed adolescents, including key neuroactive steroids. With this approach, widespread changes in the steroid metabolome were detected, most likely due to a marked ACTH-overdrive. Thus, these findings confirm the hypothesized changes in the HPA-axis of adolescents with depression and are in line with a previous study on the steroid constellation in blood of depressed adolescents of our working group ^7^, which used a subset of the same psychiatric sample.

### Increased HPA-Axis Activity in Adolescents with Depressive Symptoms

The analysis of the urinary steroid metabolome indicates dysfunction at the adrenal level, suggestive of chronic ACTH-overdrive in adolescent depression. This conclusion is supported by several key findings:

First, we observed increased excretion rates of all steroid precursors and their metabolites, with significant group differences for pregnenolone and 17-OH-pregnenolone, all of which are regulated by ACTH ^7^.

Second, in line with elevated precursor levels, we found increased levels of corticosterone and its metabolites. Corticosterone is synthesized in both the zona glomerulosa, under the influence of angiotensin and potassium, and the zona fasciculata, alongside glucocorticoids, under the control of ACTH ^32^. The increase in mineralocorticoid synthesis in response to chronic ACTH overdrive is consistent with observations in patients with pituitary Cushing’s syndrome ^13,33^. This finding is also supported by previous research on serum adrenal steroids in MDD, including our recent study on the serum steroid metabolome in adolescents ^7^.

Third, in contrast to most of the HPA-axis metabolites studied here, we observed reduced levels of estradiol in the psychiatric sample. Since stress-induced activation of the HPA axis can interfere with the hypothalamic–pituitary–gonadal (HPG) axis, particularly under chronic stress ^34^, these findings indicate persistent ACTH-overdrive in depressed adolescents. However, caution is warranted in interpreting these results, as we were unable to control for menstrual cycle stages.

Fourth, there was a general trend towards higher glucocorticoid levels in depressed adolescents compared to controls. These findings align with our previous study on serum adrenal metabolites ^7^ and with observations of steroid hormone alterations in pituitary Cushing’s disease ^13,33^.

Fifth, individual DHEA metabolites such as androstenediol-17α (A5-3β,17α) and androstenetriol-16α (A5T-16α) were elevated in the psychiatric sample compared to controls in primary and sensitivity analyses. Given that DHEA levels increase in response to ACTH-mediated stress ^35^, these findings further implicate HPA axis dysfunction in adolescent MDD. DHEA is a negative allosteric modulator of central GABA_A_ receptors, thereby reducing GABA**_A_**-induced inhibition ^36^. In adolescents, high levels of DHEA have been associated with onset of depression and higher burden of anxiety symptoms ^37,38^.

Notably, this is the first study to investigate the role of neuroactive steroids other than DHEA for depression in adolescents. Neuroactive steroids as allopregnanolone (aPo), pregnanolone (Po), 3α-androstanediol (5αAD-3α,17β) or tetrahydro-11-deoxycorticosterone (TH-DOC) directly interact with receptors of the CNS (mainly inhibitory GABA_A_ receptors) and thus can alter CNS excitability. Descriptively, we found increased urinary excretion rates of allopregnanolone, pregnanolone and 3α-androstanediol in adolescents with depressive symptoms compared to controls. However, neither the excretion rates of these neuroactive steroids nor their ratios showed a significant group effect in primary and sensitivity analyses, leaving the findings inconclusive.

### Ratios of Urinary Steroid Metabolites Show Biomarker Potential

To reduce the complexity of a comprehensive steroid metabolome analysis and observer-related confounding ^39,40^, we applied an unsupervised random-forest classifier to analyze whether combining multiple metabolite ratios (as indicators for enzyme activity) can be utilized to identify depressed adolescents compared to controls. The best performing classifier was based on 26 ratios and provided strong diagnostic properties with an AUC of 0.827 (95%-CI [0.757 - 0.907]). Notably, the seven top performing ratios also exhibited significant group effects after FDR correction and adjustment for several covariates in rank-based linear regression models (Fig. 3, Tab. 3), adding further validity to their use as diagnostic biomarkers.

In the current study, the ratio of the 11-deoxycorticosterone metabolite TH-DOC to the sum of corticosterone metabolites emerged as one of the best-performing biomarkers to differentiate between patients and controls (AUC = 0.800, 95%-CI: [0.702 - 0.882]). In line, our previous study on serum steroid profiles in depressed adolescents identified the 11-deoxycorticosterone / corticosterone ratio as a valuable biomarker ^7^ and hypothesized that lower serum 11-deoxycorticosterone levels may result from increased TH-DOC synthesis. However, the urine metabolome of depressive adolescents described in the present study does not support this hypothesis, as TH-DOC excretion rates were descriptively reduced in patients compared to controls.

The present study, along with previous research, provides evidence of increased glucocorticoid and mineralocorticoid synthesis in adolescent MDD. This implies the potential to modify disease-related steroid imbalance in MDD: the 11β-hydroxylase inhibitor Metyrapone increases urinary TH-DOC levels and decreases corticosterone levels in depressed adults ^41^. TH-DOC has direct GABA_A_-ergic inhibitory and potentially antidepressant effects while corticosterone maintains the stress response via central MR-receptors ^8,42–44^. However, while Metyrapone has shown augmenting effects when combined with conventional antidepressants or as monotherapy in treatment-resistant depression ^45^, a more recent randomized trial failed to demonstrate additional benefit ^46^. Nonetheless, considering the broad inclusion criteria and the focus on adults in the study byMcAllister-Williams, Anderson, Finkelmeyer, Gallagher, Grunze, Haddad, Hughes, Lloyd, Mamasoula and McColl ^46^ the ratio of TH-DOC to the sum of corticosterone metabolites could be a promising surrogate maker to identify candidates for more individualized Metyrapone-trials in adolescents.

### The Role of 5α-Reductase Inhibition in Adolescent Depression

Notably, several ratios consistently indicated a reduced 5α-reductase activity in the psychiatric sample, with significant group differences in progestagene metabolism (Ratio of Po-5α,3α to Po- 5β,3α), glucocorticoid metabolism (ratio of allo-THF to THF), and androgen metabolism (ratio of An to Et). This suggests a widespread association of 5α-reductase activity with depression. In line with this conclusion, 5α-reductase inhibitors, utilized for the treatment of androgenetic alopecia or benign prostatic hyperplasia, are known to induce neuropsychiatric side effects, in particular depression and self-harming behavior ^47,48^. Prior research on 5α-reductase inhibitors and depression mainly involved older men. The current study suggests reduced 5α-reductase activity in adolescents, suggesting a broader role for 5α-reductase metabolism in the development of depressive symptoms. This is supported by prior findings of reduced 5α-reductase activity, as estimated through urinary steroid metabolite ratios measured via immunoassays, in 19 depressed women compared to 15 controls ^6^.

One widely proposed mechanism for how 5α-reductase inhibitors contribute to affective disorders involves decreasing neuroactive steroid levels, particularly allopregnanolone ^47,49^. However, this hypothesis is not supported by our findings, as patients descriptively exhibited increased excretion rates of allopregnanolone (aPo). Alternative mechanisms, such as interactions with dopaminergic pathways or epigenetic modifications ^47^, warrant further investigation. The consistent observation of reduced 5α-reductase activity in our study highlights its potential as a promising target for future research on adolescent depression.

### Limitations

This study is subject to several limitations that should be addressed in future studies.

First, the psychiatric sample provided morning spot urine samples, while the control cohort collected 24-hour urine samples. To account for this, metabolite concentrations were standardized in relation to creatinine excretion rates, which were comparable between both groups. Additionally, metabolite ratios as proxies for enzyme-activities were compared between groups and used in ROC analyses since they are less susceptible to sampling conditions ^28^. In our study, relatively few glucocorticoid metabolites exhibited significant group differences. This finding may relate to diurnal fluctuations of urinary steroid metabolites, particularly glucocorticoids, peaking midday to afternoon ^50^, and sampling in the psychiatric cohort in the early morning. Supporting this conclusion, a recent study investigating intraindividual comparisons of steroid excretion rates between spot and 24-hour urine samples reported low correlations for glucocorticoid metabolites but high correlations for androgen and corticosterone metabolites ^28^.

Second, while urine collection is non-invasive and unlikely to alter steroid metabolism, differences in sampling conditions—such as recent hospital admission for psychiatric patients compared to at-home collection for controls—may have led to perceived stress, potentially biasing the results. However, as previously discussed, there was no evidence of an effect of hospitalization on (serum) cortisol levels when comparing inpatients and outpatients in the present cohort ^51^.

Third, while the study is powered to detect medium to large group effects, it lacks power to fully analyze confounders and interactions. This limitation is particularly relevant for sex-specific effects, given the small number of males in the control (n = 11) and psychiatric (n = 12) sample.

Fourth, the psychiatric sample comprised of a heterogenous group of psychiatric patients with frequent psychiatric comorbidities (Tab. 1) and potential additional characteristics, such as experienced adverse life events, which may contribute to the manifestation and severity of depressive symptoms. Thus, even though all patients exhibited at least mild depressive symptoms according to established criteria (BDI–II ≥ 14), and key findings regarding biomarker potential were replicated in a subsample with an additional clinical diagnosis of depression, the specificity of our results to depression remains to be determined in future studies.

Fifth, the control group was drawn from a population-based cohort, and information on underlying diseases relied on self-reports. Although individuals reporting psychiatric conditions were excluded, systematic screening (e.g. using the BDI-II) for psychiatric disorders could enhance the selection of appropriate controls.

## Conclusions

Here, we comprehensively describe the urinary steroid metabolome of adolescents with depressive symptoms, including the analysis of neuroactive steroids. This study confirms and extends key findings of our previous study on serum steroid profiles in an overlapping cohort ^7^: Patients were characterized by a distinct pattern of HPA-axis upregulation, presumably reflecting a chronic ACTH-overdrive. Analyses of enzyme activities revealed a reduction in 5α-reductase activity, indicating its potential role in the etiology of adolescent MDD. The ratio of TH-DOC to the sum of corticosterone metabolites exhibited the most promising biomarker performance to identify patients. Future studies are needed to address the methodological limitations of the current study and to evaluate the potential of our findings to guide individualized therapeutic approaches in this highly relevant disease entity.

## Data Availability

Anonymized data may be available upon reasonable request to the corresponding author.

## Acknowledgments

This work was supported by a fellowship of the University Medicine Essen clinician scientist Academy (UMEA) to LD which was supported by the German Research Foundation (Deutsche Forschungsgemeinschaft; FU 356/12-2). The DONALD study was funded by the Ministry of Innovation,Science and Research of North Rhine-Westphalia, Germany. The manuscript’s language style was enhanced through the utilization of artificial intelligence (ChatGPT). Figures were created with BioRender.com.

## Author contributions

RH, CG, SAW and LD conceptualized the study. LD and RH designed the statistical analysis. LD conducted statistical analyses. LD wrote the first draft of the manuscript. Steroid analyses were carried out in the Steroid Research & Mass Spectrometry Unit of the Division of Pediatric Endocrinology & Diabetology, Justus Liebig University Giessen (Head SAW) under the responsibility of MFH and SAW. SAW, MFH, LL, MF, JH, AH, UN, UA, CG, and RH critically revised the manuscript.

## Abbreviations

ACTH: Adrenocorticotropic hormone
AKR1C1: Aldo-keto reductase family 1 member C1 or 20α-hydroxysteroid dehydrogenase
AUC: area under the curve
BDI-II: Beck Depression Inventory II
BMI: body mass index
CYP3A4: Cytochrome P450 3A4
DONALD study: Dortmund Nutritional and Anthropometric Longitudinally Designed study
DSM-IV: Diagnostic and Statistical Manual of Mental Disorders-IV
FDR: false discovery rate
GABA_A_ receptor: γ-aminobutyric acid A receptor
GC-MS: gas chromatography-mass spectrometry
HPA: hypothalamic-pituitary-adrenal
3β-HSD: 3β-hydroxysteroiddehydrogenase
11β-HSD: 11β-hydroxysteroiddehydrogenase
17β-HSD: 17β-hydroxysteroiddehydrogenase
ICD-10: International Statistical Classification of Diseases and Related Health Problems 10
K-SADS-PL: Kiddie Schedule for Affective Disorders and Schizophrenia for School Aged Children, Present-Lifetime version
LC-MS: liquid chromatography-tandem mass spectrometry
LLD: lower limit of detection
MDD: major depressive disorder
ROC: receiver operating characteristic.
For a complete list of abbreviations used for single steroid metabolites, see Fig. 1.

