## Supplemental Material for "Urinary steroid metabolome shows adrenal, gonadal, and neuroactive steroid dysregulation in adolescents with depression"

### Urinary steroid metabolomics reveals dysregulation of adrenal, gonadal, and neuroactive steroids in adolescents with depressive symptoms

### Lars Dinkelbach, Stefan A. Wudy, Michaela F. Hartmann, Lars Libuda, Manuel Föcker, Johannes Hebebrand, Anke Hinney, Ute Nöthlings, Ute Alexy, Corinna Grasemann, and Raphael Hirtz

Supplemental Methods

Power Analysis

Cross-validation in ROC analyses

Boxplots

Supplemental Figure 1 – Progestagen (C21-steroids) urine metabolites.

Supplemental Figure 2 – Mineralocorticoid (C21-steroids) urine metabolites.

Supplemental Figure 3 – Glucocorticoid (C21-steroids) urine metabolites.

Supplemental Figure 4 – Androgen (C19-steroids) urine metabolites.

Supplemental Figure 5 – Estrogen (C18-steroids) urine metabolites.

Supplemental Figure 6 – Summarized steroid urine metabolites.

Supplemental Figure 7 – Ratios: relative neuroactive steroid production

Supplemental Figure 8 – Ratios: relative overall androgen production.

Supplemental Figure 9 – Ratios: relative adrenal androgen production.

Supplemental Figure 10 – 11-oxygenated in relation to 11-deoxygenated androgens.

Supplemental Figure 11 – Ratios: 5α-reductase.

Supplemental Figure 12 – Ratios:11β-hydroxy-steroiddehydrogenase (11β-HSD).

Supplemental Figure 13 – Ratios: 3β-hydroxysteroiddehydrogenase (3β-HSD).

Supplemental Figure 14 – Ratios: 21-hydroxylase.

Supplemental Figure 15 – Ratios: 17β-hydroxysteroiddehydrogenase.

Supplemental Figure 16 – Ratios: 11β-hydroxylase.

Supplemental Figure 17 – Ratios: 17-hydroxylase/17,20-lyase.

Supplemental Figure 18 – Ratios: P450-Oxidoreductase (POR).

Supplemental Figure 19 – Ratio: aromatase.

Supplemental Figure 20 – Ratio: CYP3A4.

Supplemental Figure 21 – Ratio: 20α-HSD (AKR1C1).

Sensitivity Analysis – Storage Time

Supplemental Figures 22 – Scatter plot of 6β-OH-F and storage time.

ROC curves

Supplemental Figures 23 – ROC curves of selected ratios.

Supplemental Figures 24 –	Sensitivity analyses: ROC curves of the

TH-DOC / Corticosterone ratio

Supplemental Figures 25 –	ROC curve of the 6β-OH-F / F ratio

Supplemental References

**Supplemental Methods**

**Power Analysis**

There is no established approach for power analyses of the robust rank-based analysis of variance. Therefore, power analyses of the unadjusted Wilcoxon rank sum tests were performed using G*Power 3.1.9.7 (1). Power analyses for the Wilcoxon rank sum tests rely on assumptions about the underlying distributions, which are difficult to define due to the exploratory nature of this study and the large number of examined metabolites. However, for estimation purposes, normal distributions and an uncorrected alpha error of 0.05 (two-sided) were assumed. Under these assumptions, analyses had power (1 – beta-error = 0.8) to detect a small to medium effect size of ∣d∣ > 0.47.

**Cross-validation in ROC analyses**

For multivariate ROC analyses, Monte-Carlo cross validation with balanced sub-sampling was performed. In each cross-validation cycle, two-thirds of the total sample size were used to assess the relative importance of each variable. In six separate models, the three, five, 10, 20, 26, and 52 (full model) ratios ranking highest were then integrated into random forest classification models, which were evaluated in the remaining one-third of the sample. For univariate ROC analysis, a two-fold cross-validation was performed by dividing the sample into a test (N = 80) and verification sample (N = 70) while preserving the 1:1 matching of patients and controls. The univariate ROC curves were compared between these subsamples using DeLong’s test (R package pROC, version 1.18.5, (2)).

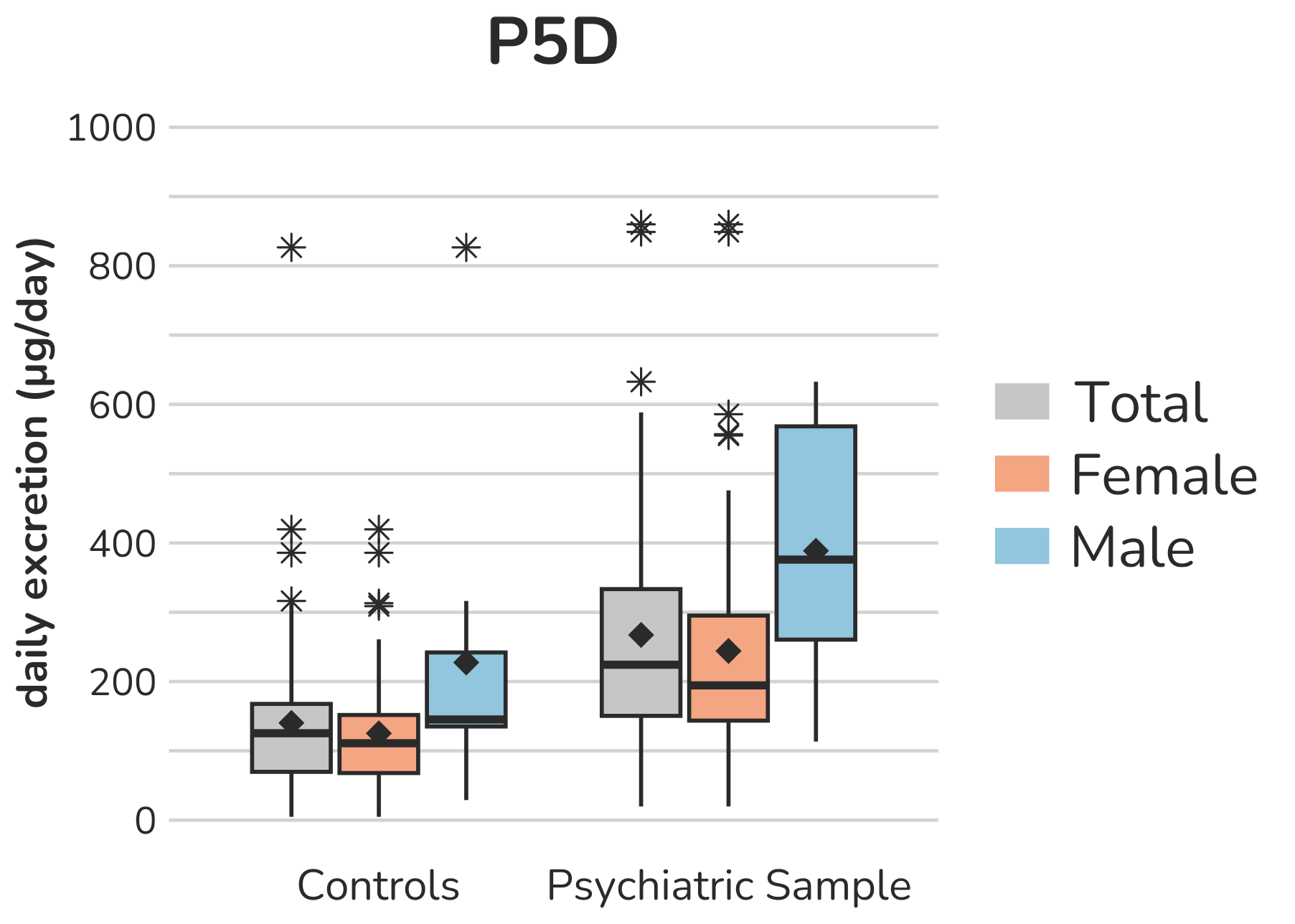

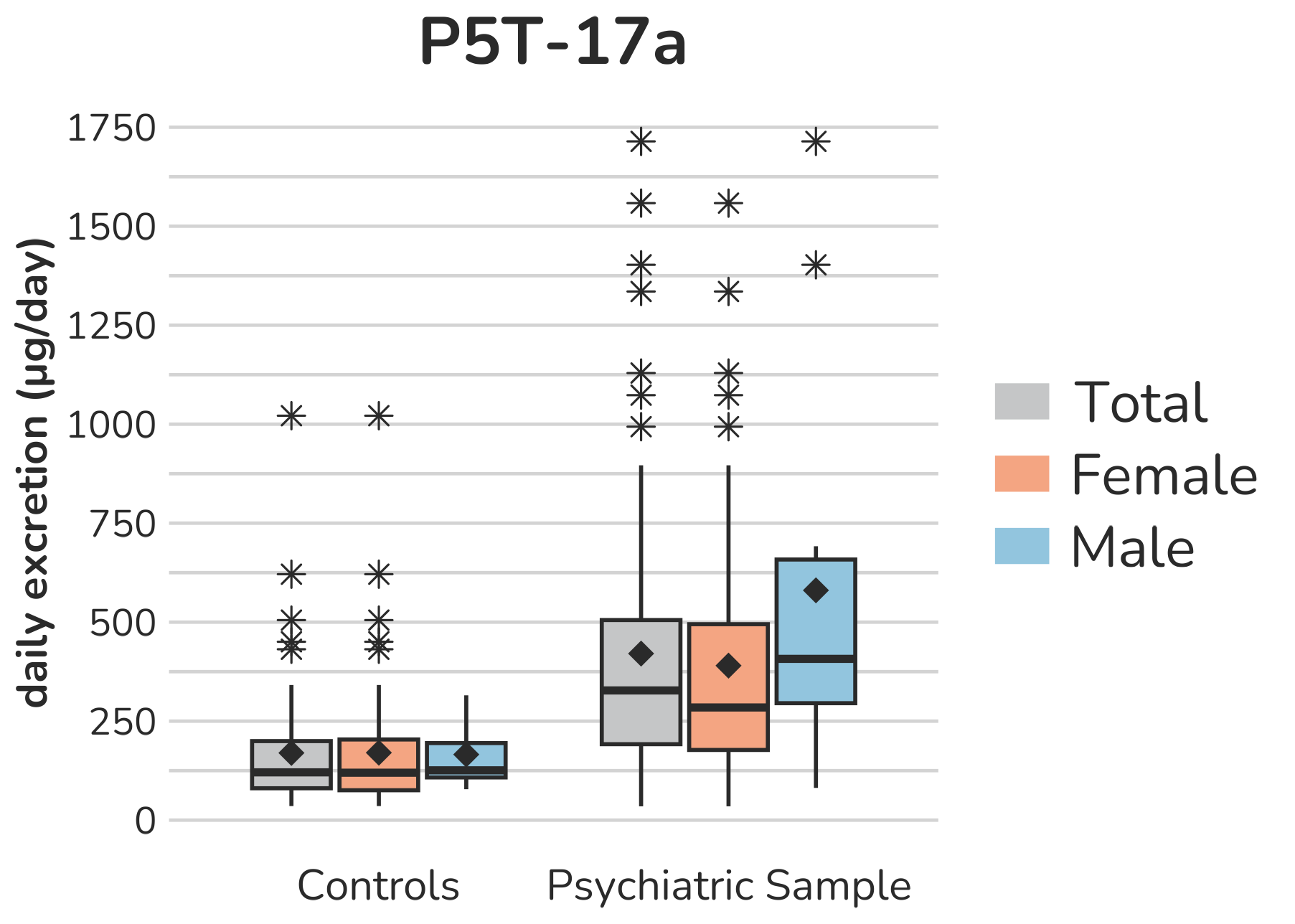

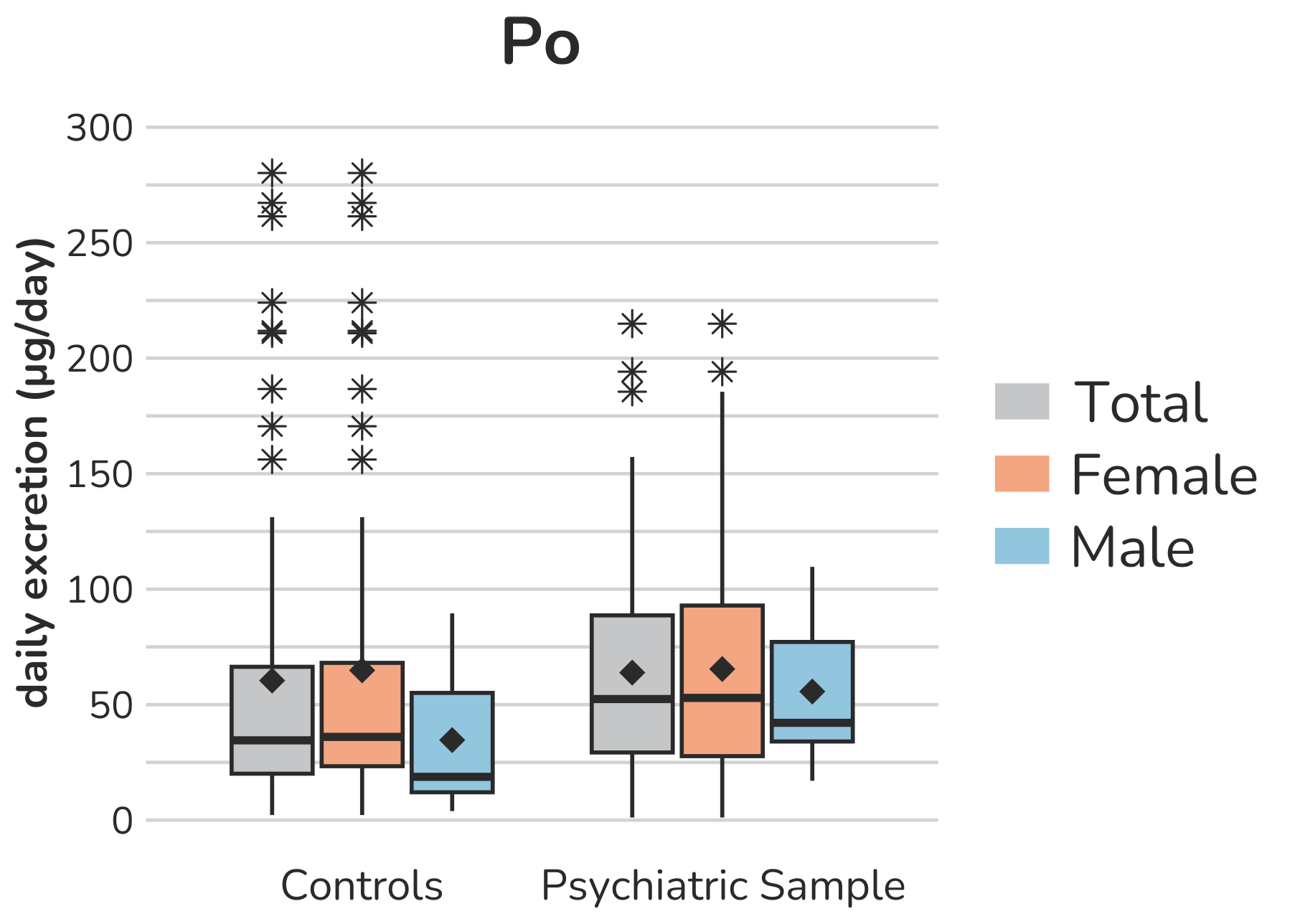

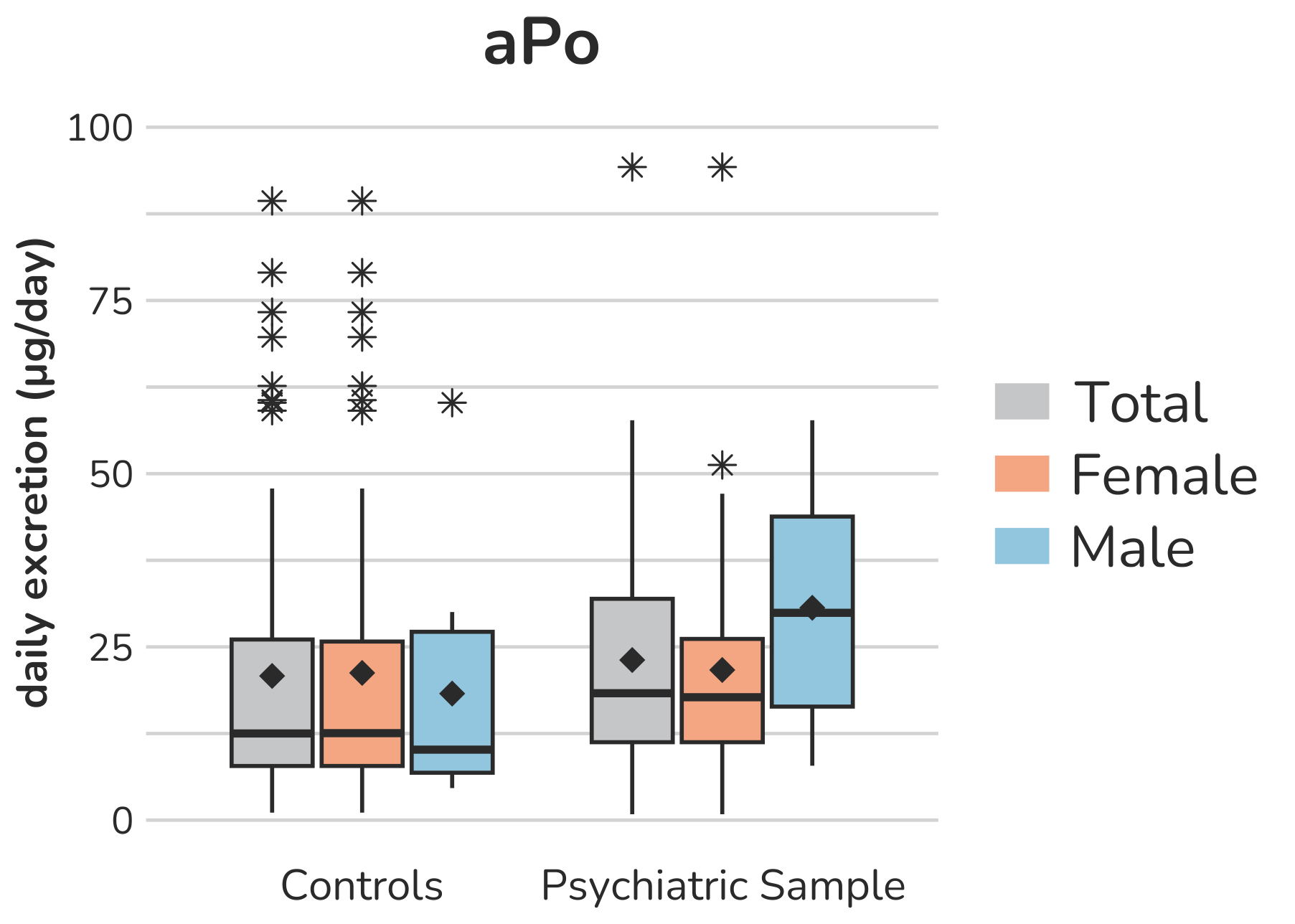

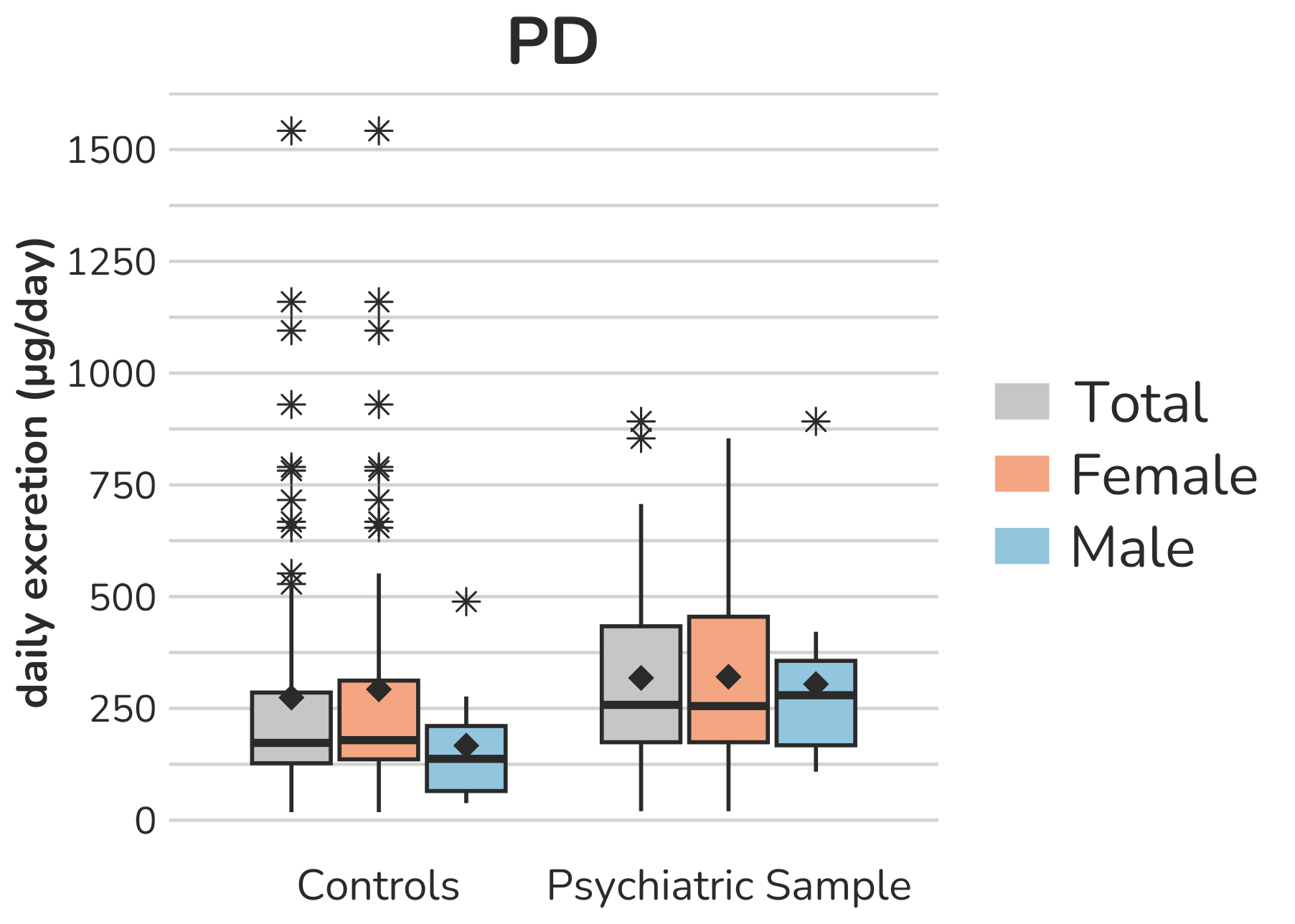

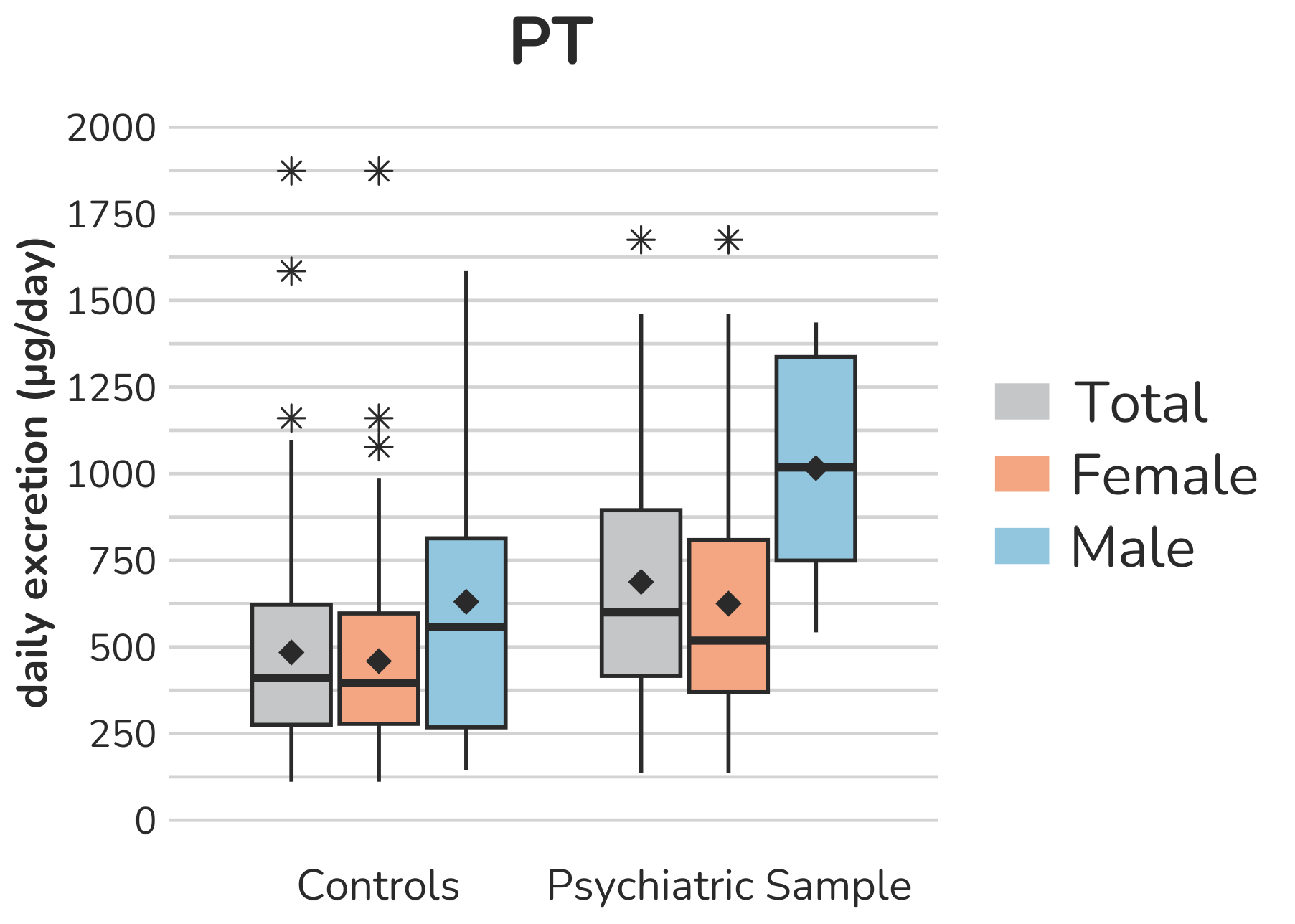

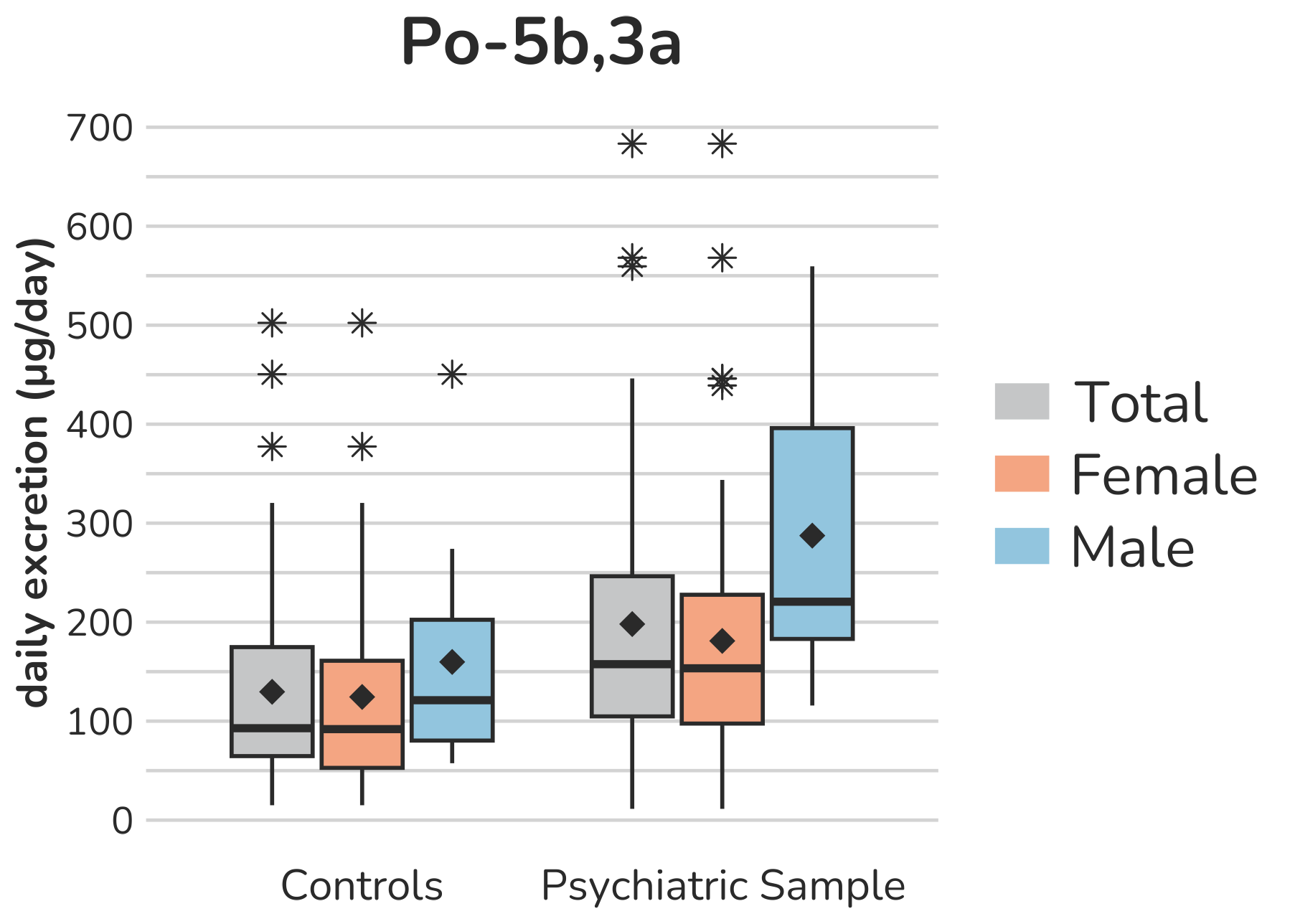

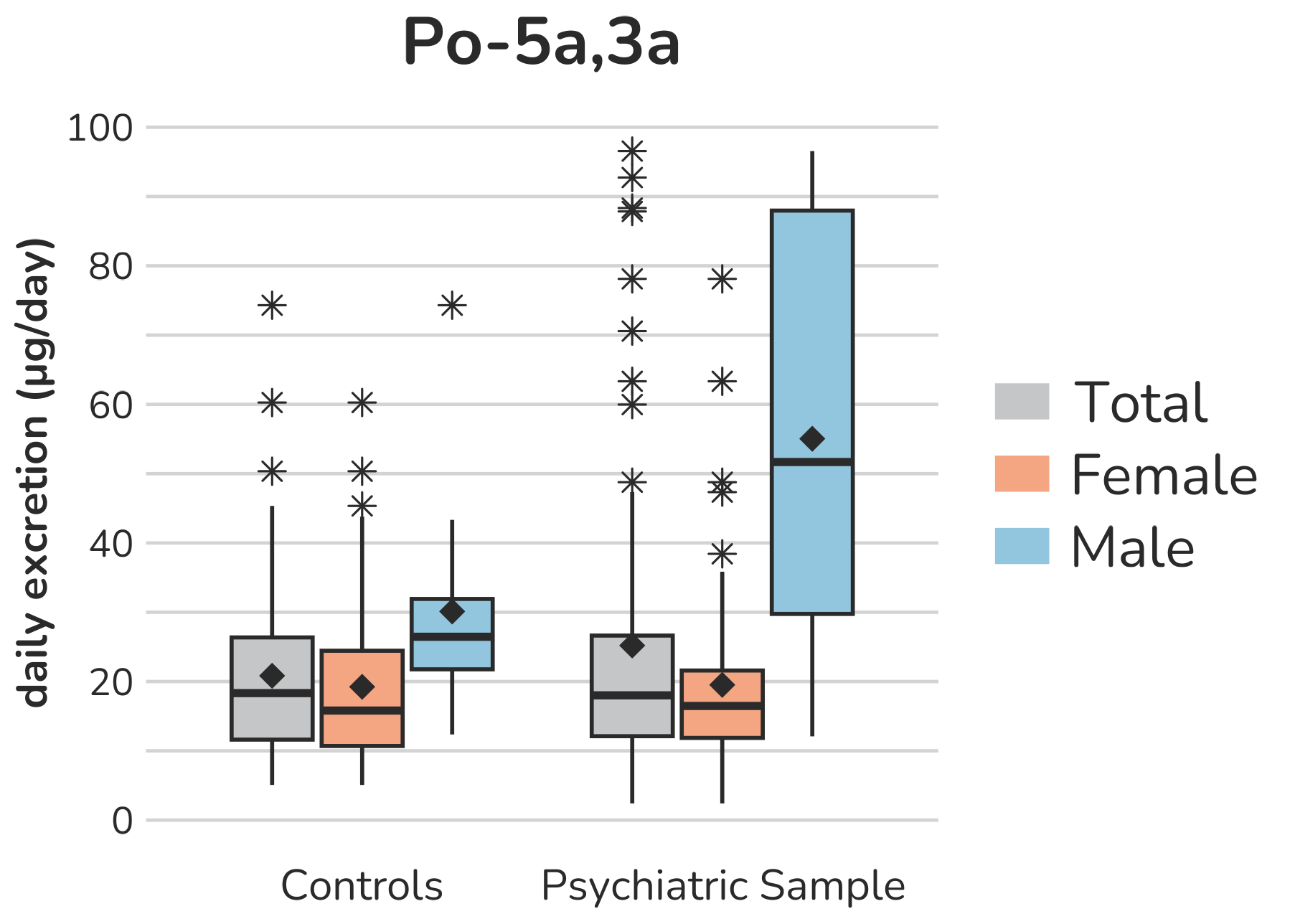

**Supplemental Figure S1 – Boxplots of progestagen (C21-steroids) urine metabolites.** Each box gives the 25th and 75th percentile (lower and upper boundary of the box), the median (horizontal line within each box), and the mean (rhombus within each box). The whiskers are defined by 1.5 times the interquartile range. For P5T-17a one subject (male, control group) with a daily excretion of 2963.7µg is excluded from the figure to enhance the visualization of the remaining values. Aside from this, all outliers are displayed (illustrated by *).

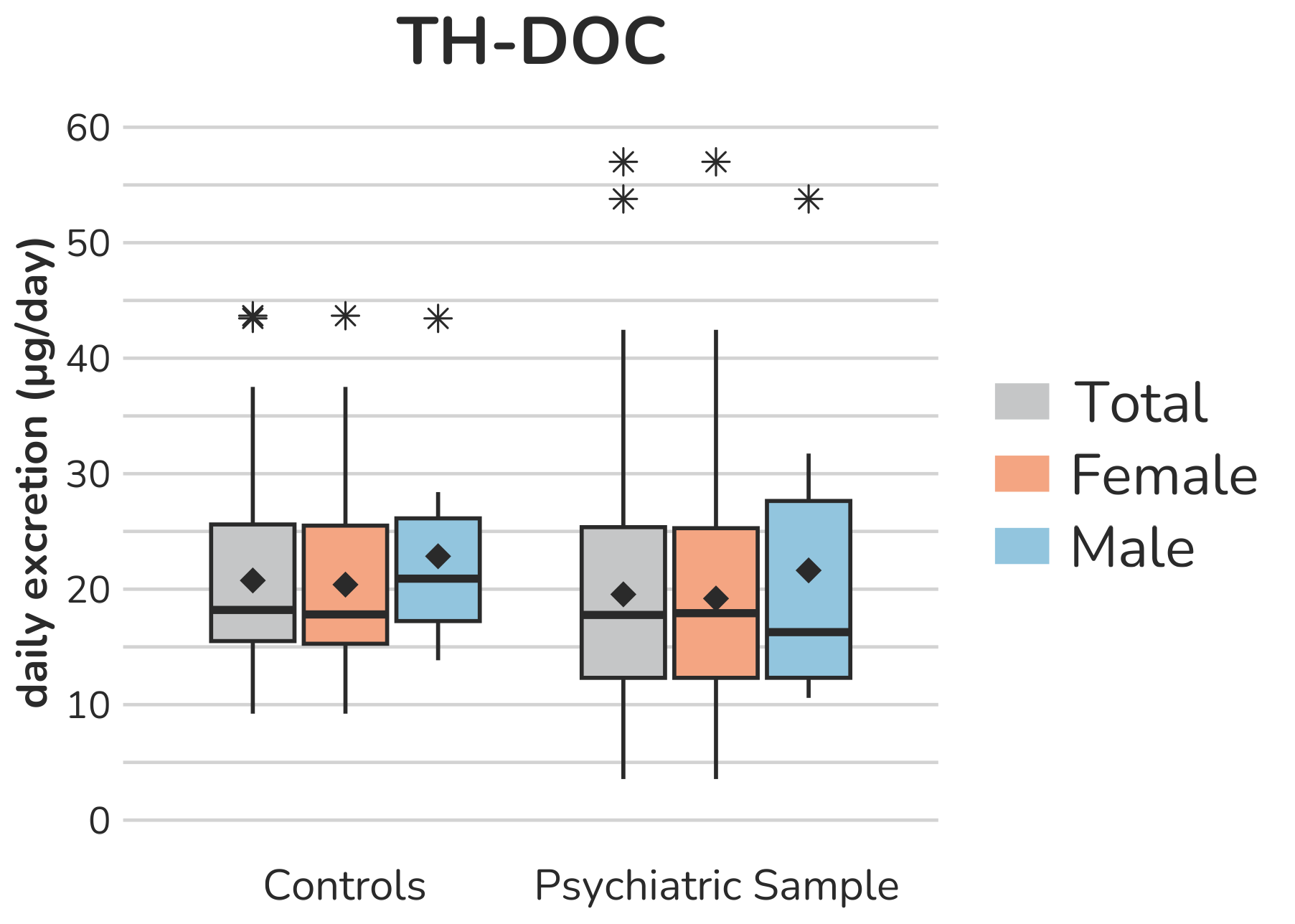

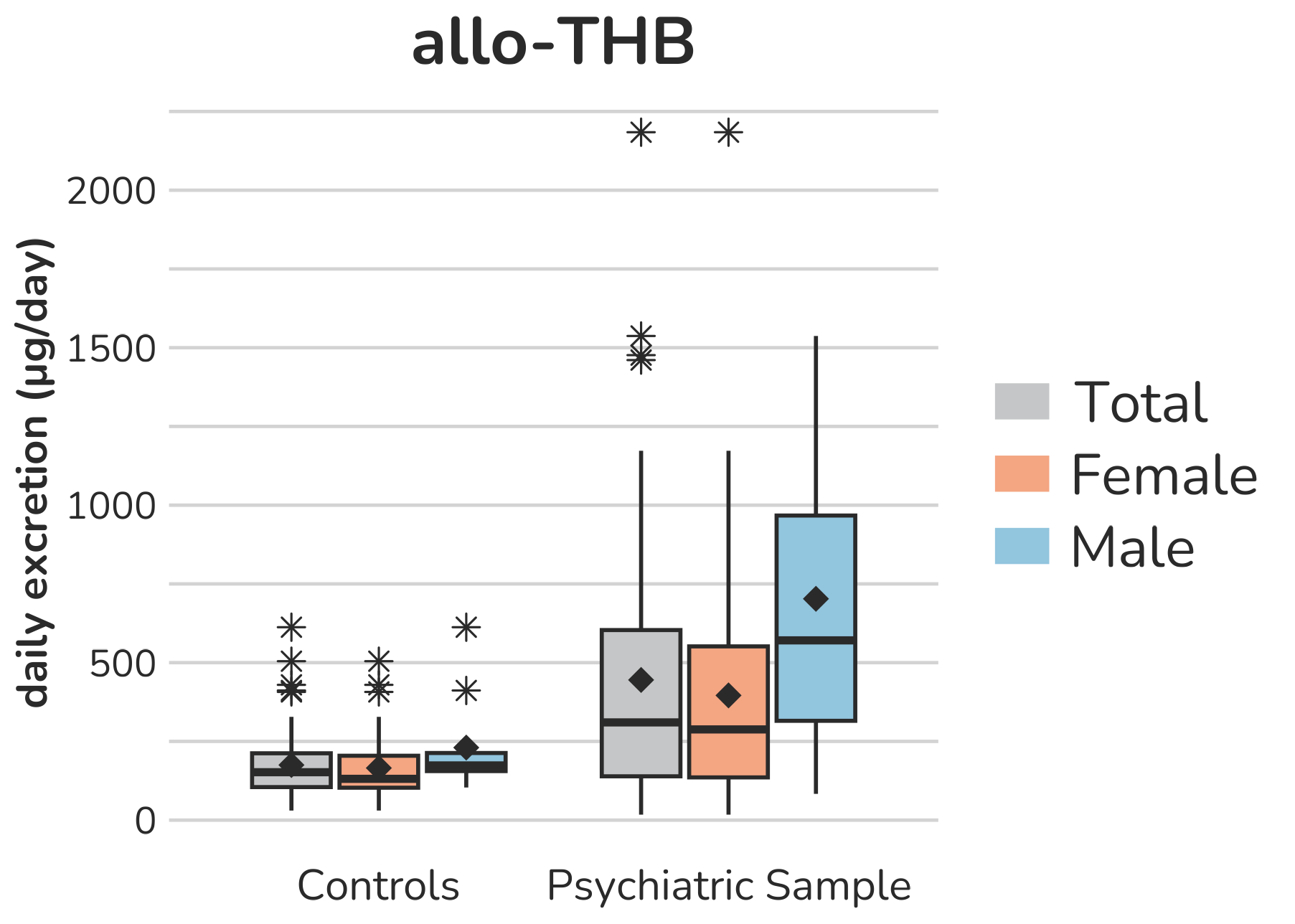

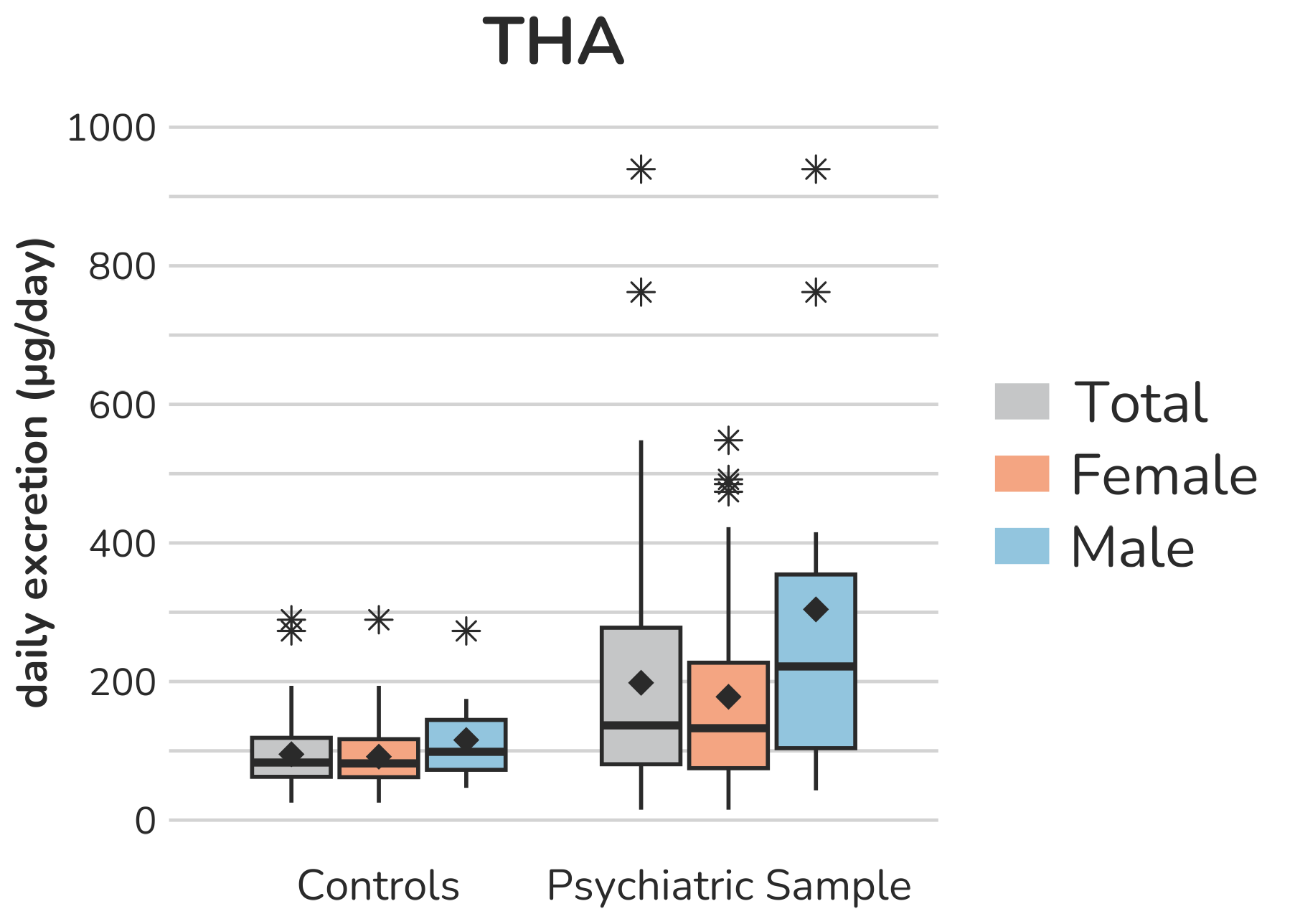

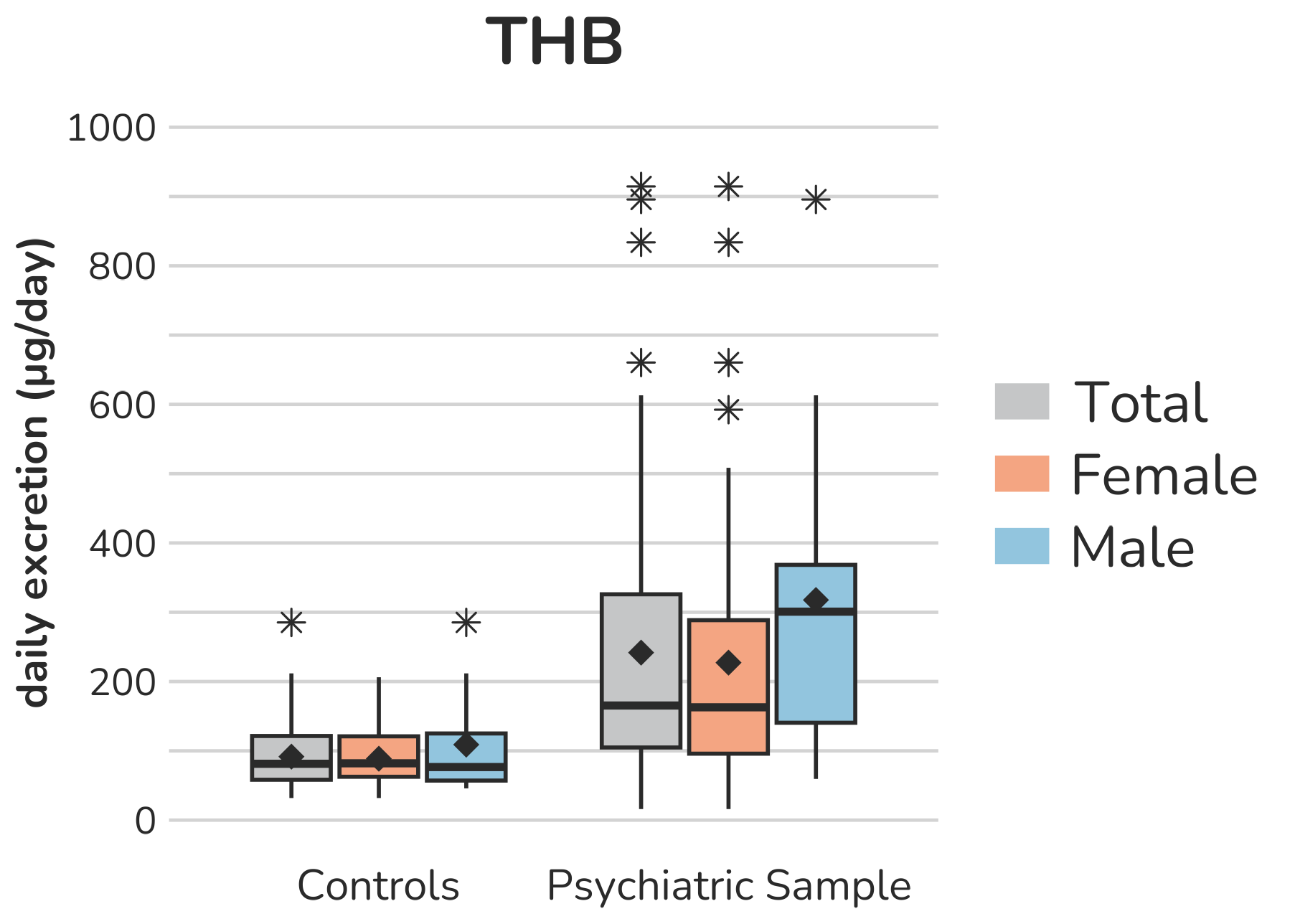

**Supplemental Figure S2 – Boxplots of mineralocorticoid (C21-steroids) urine metabolites.** Each box gives the 25th and 75th percentile (lower and upper boundary of the box), the median (horizontal line within each box), and the mean (rhombus within each box). The whiskers are defined by 1.5 times the interquartile range. For TH-DOC one subject (male, psychiatric sample) with a daily excretion of 142.2µg is excluded from the figure to enhance the visualization of the remaining values. Aside from this, all outliers are displayed (illustrated by *).

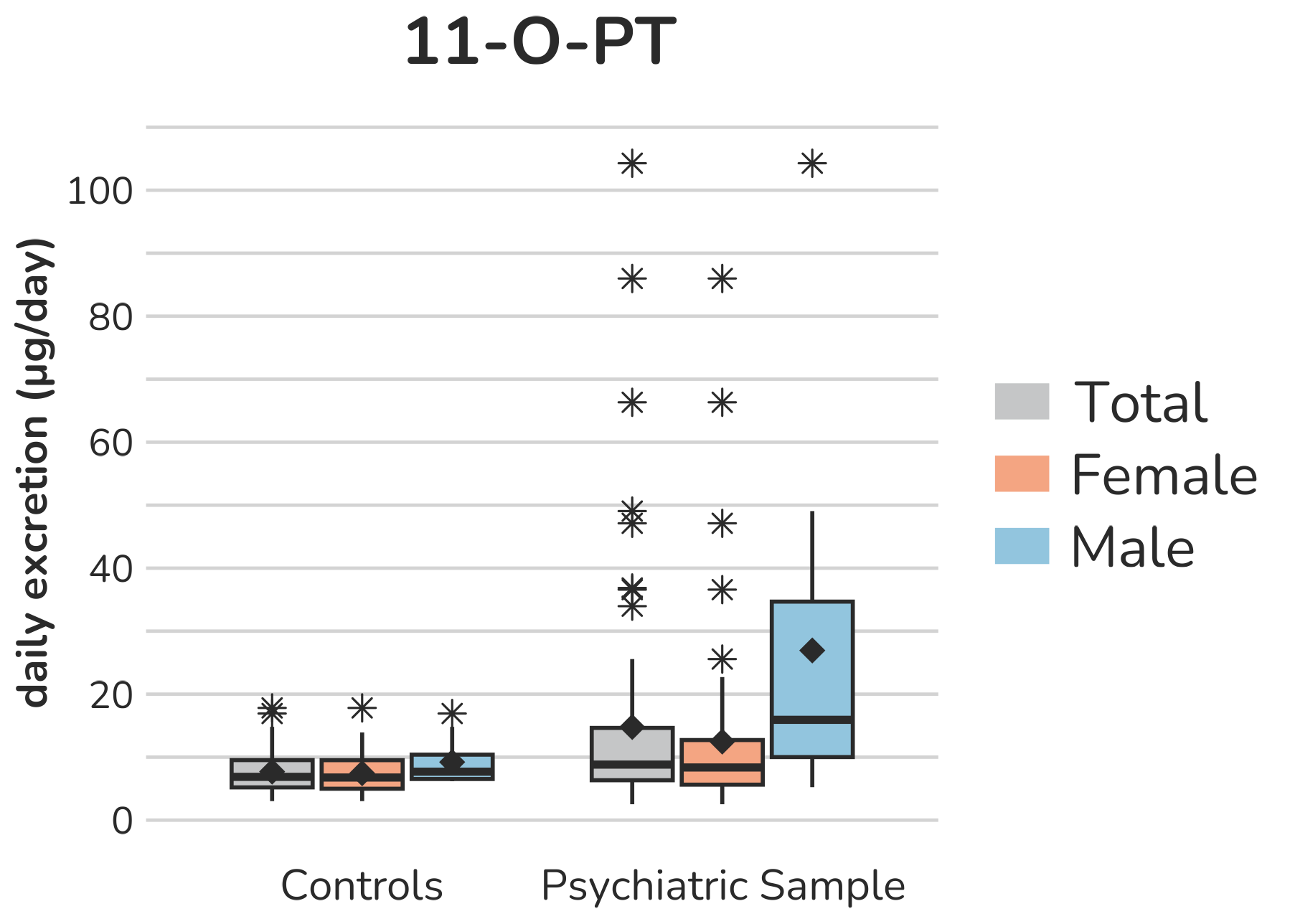

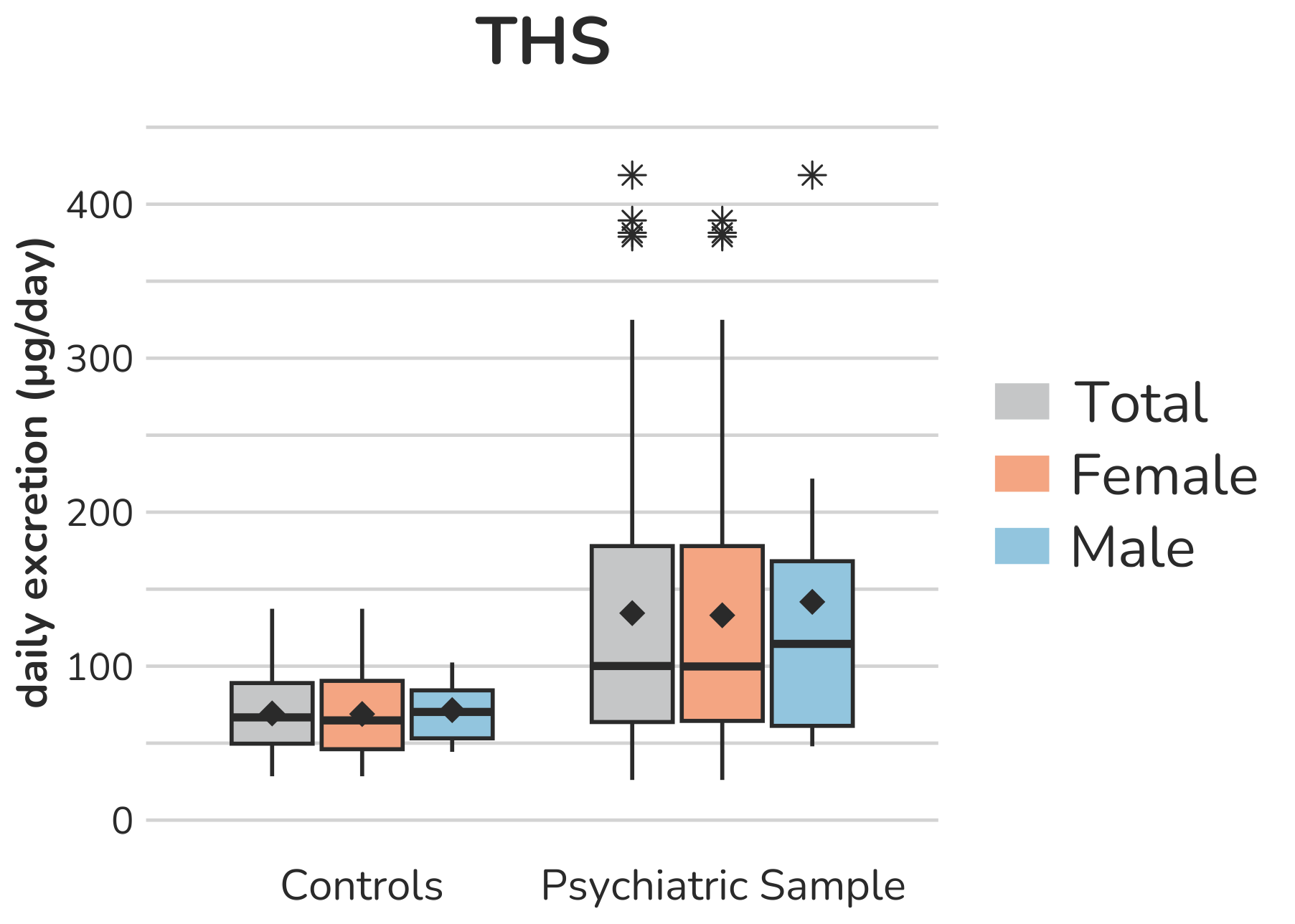

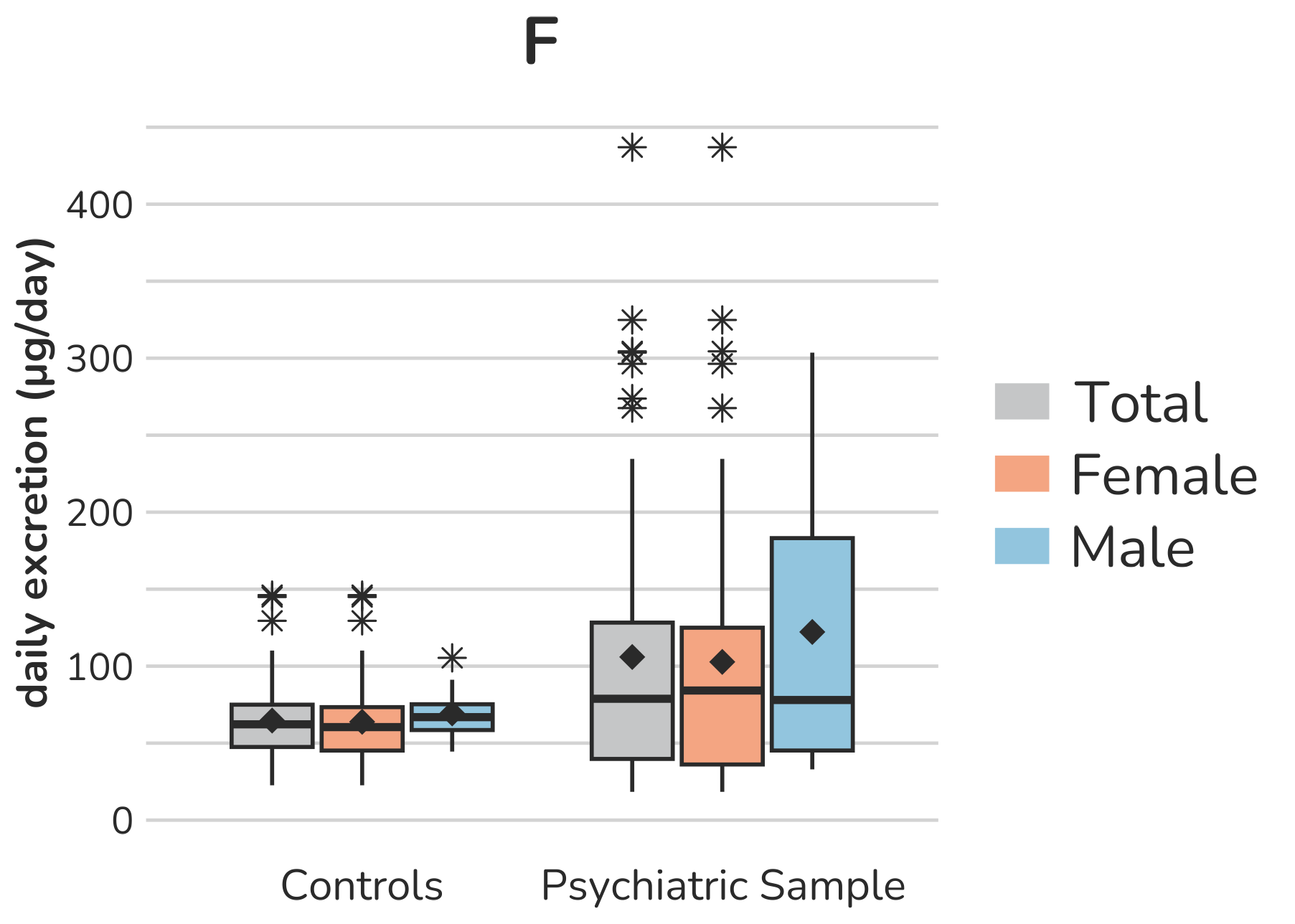

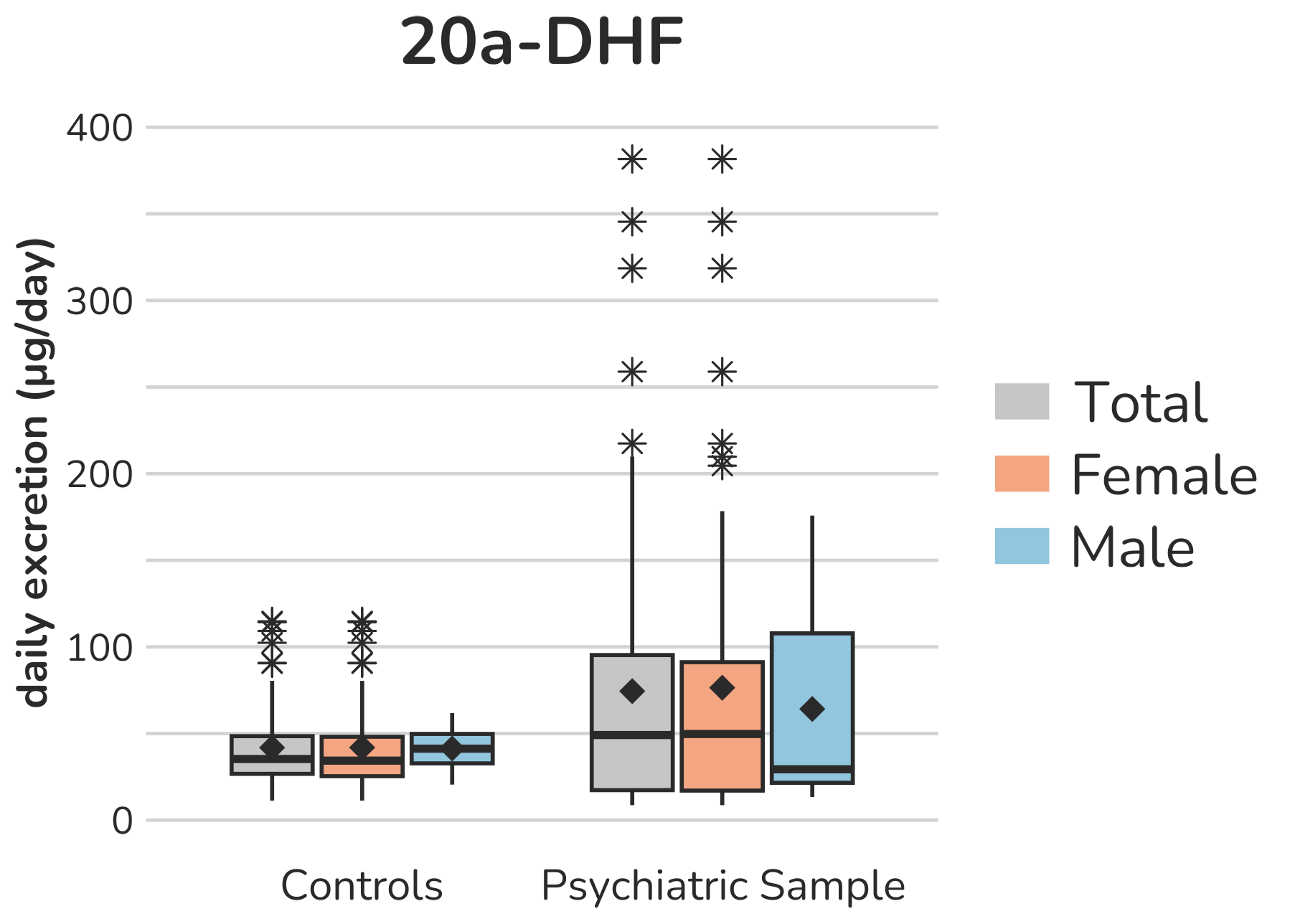

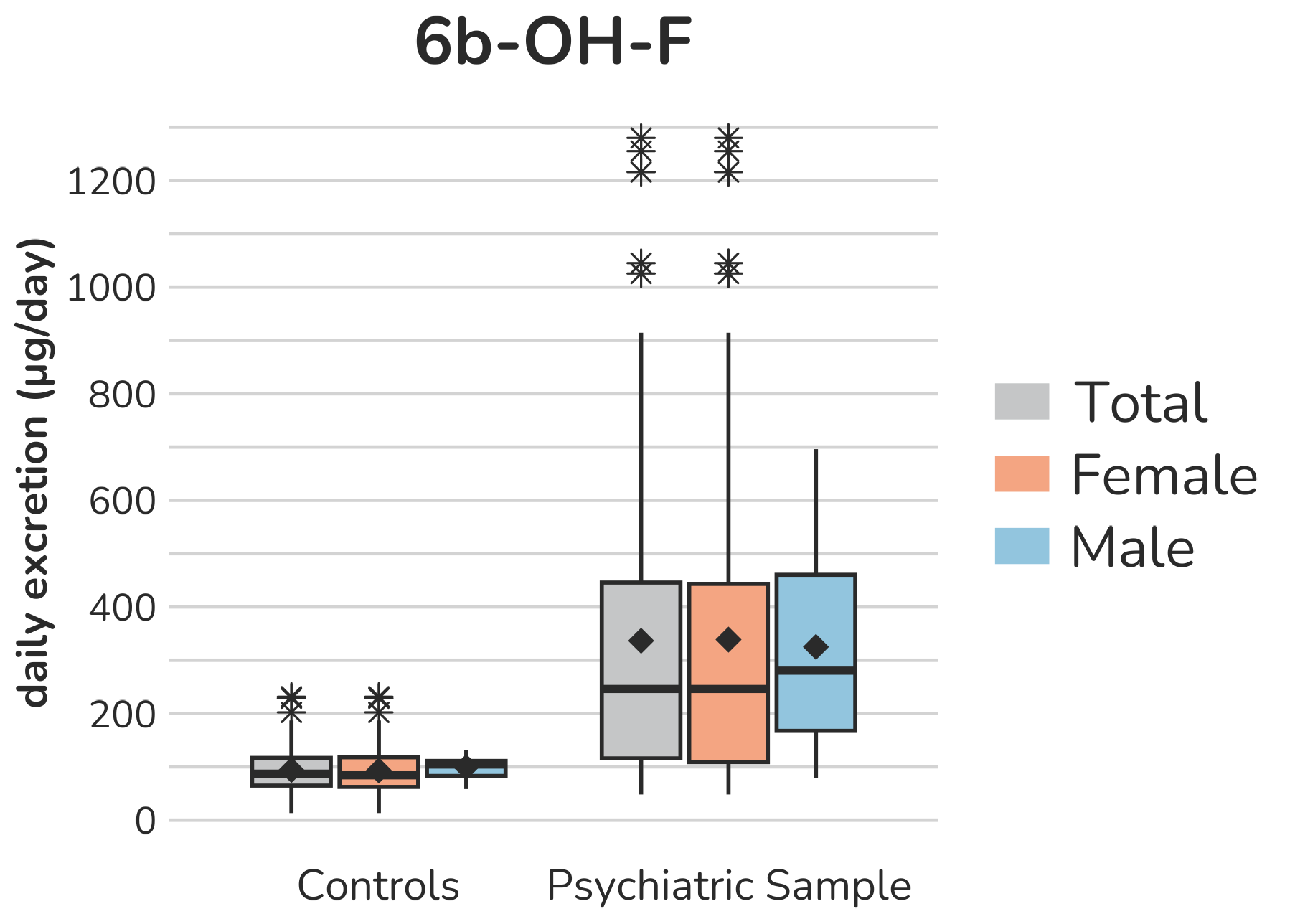

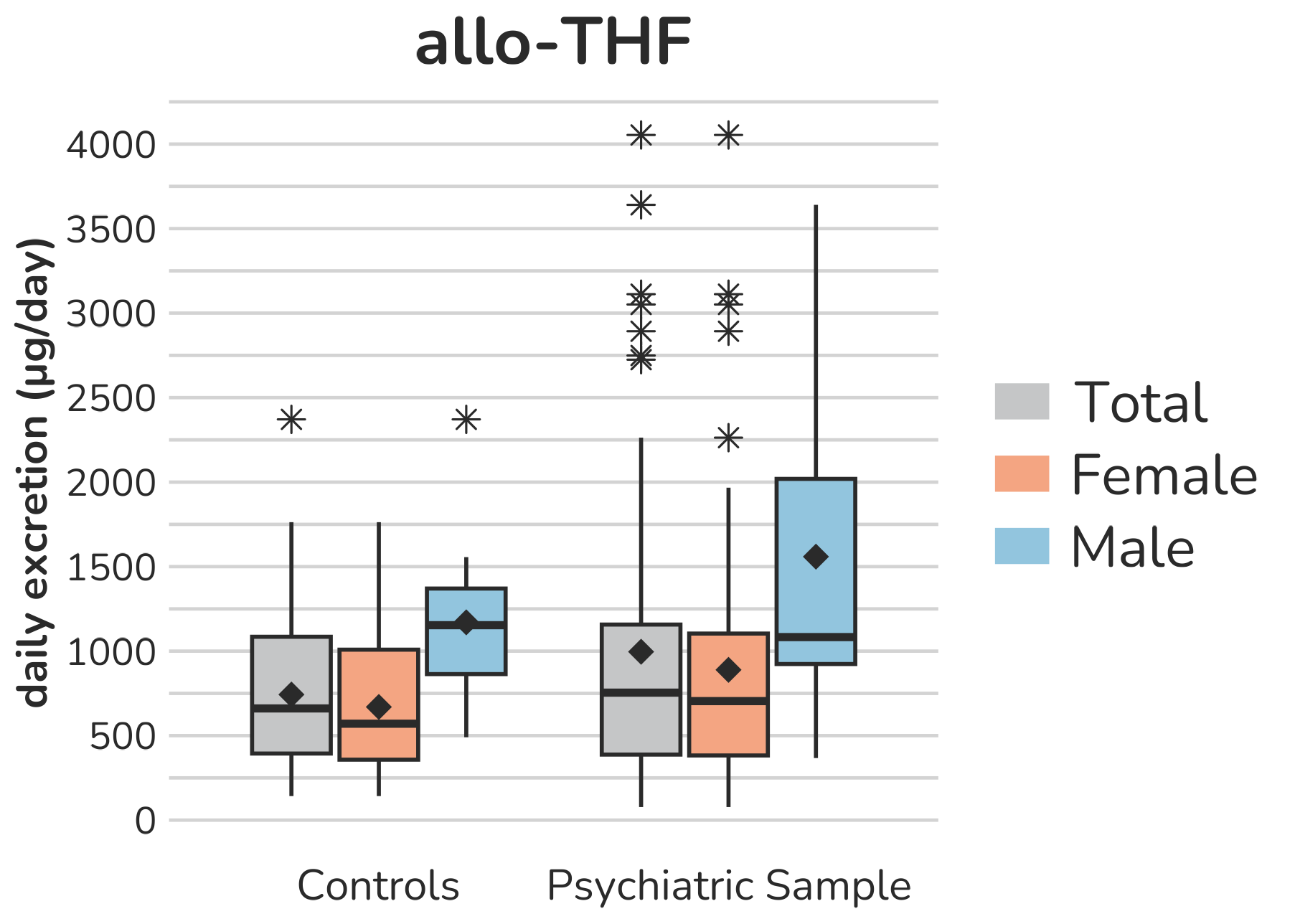

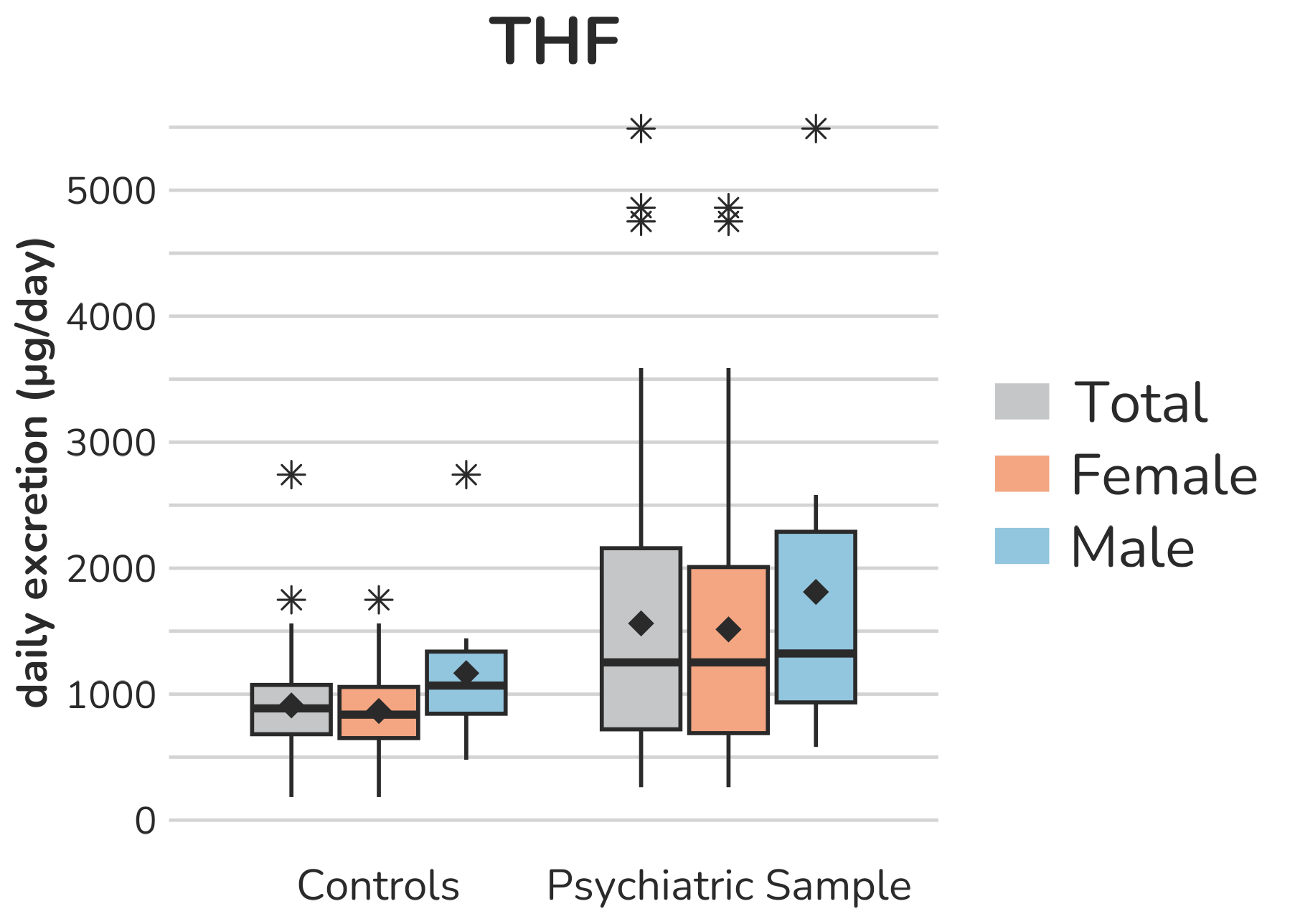

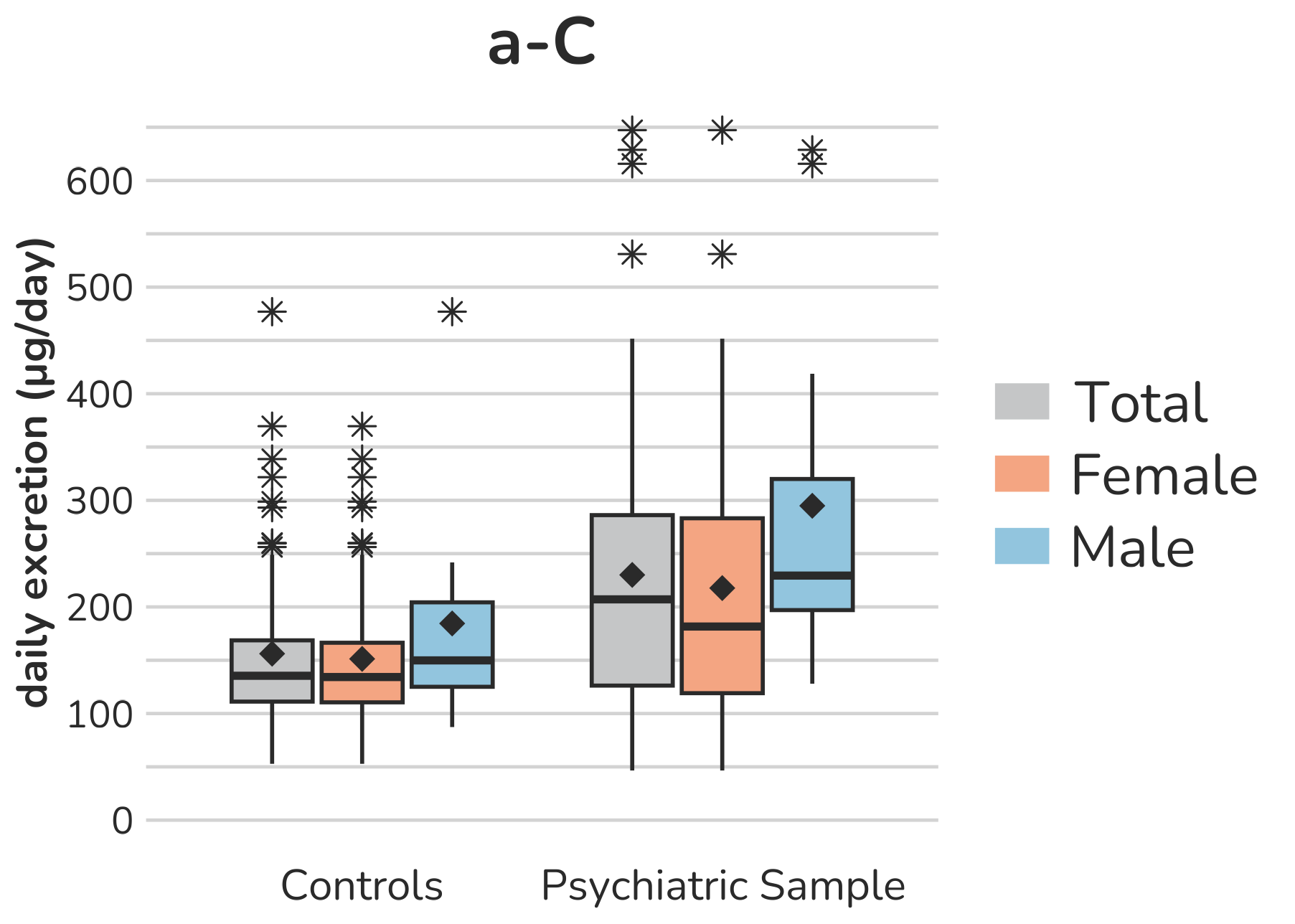

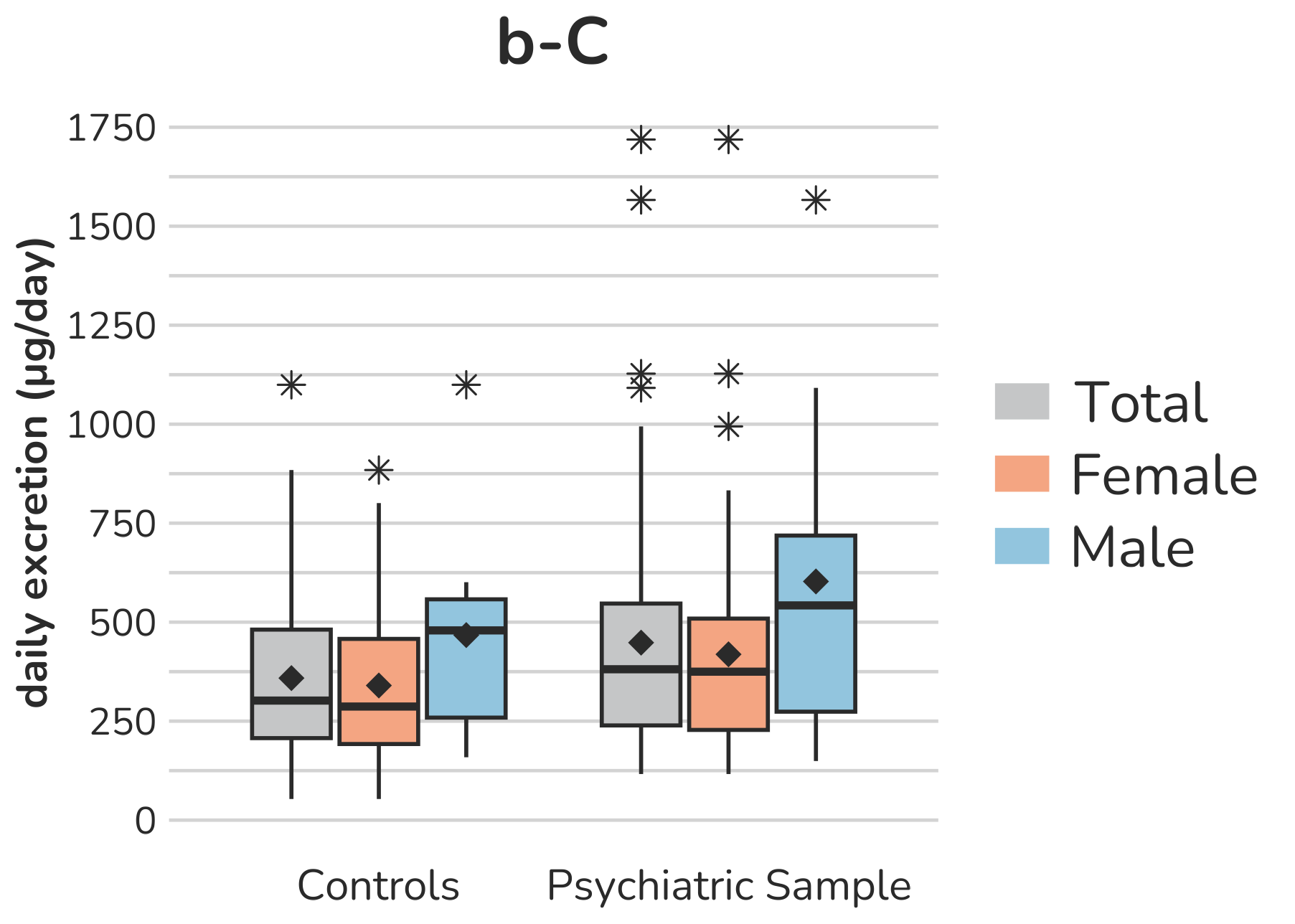

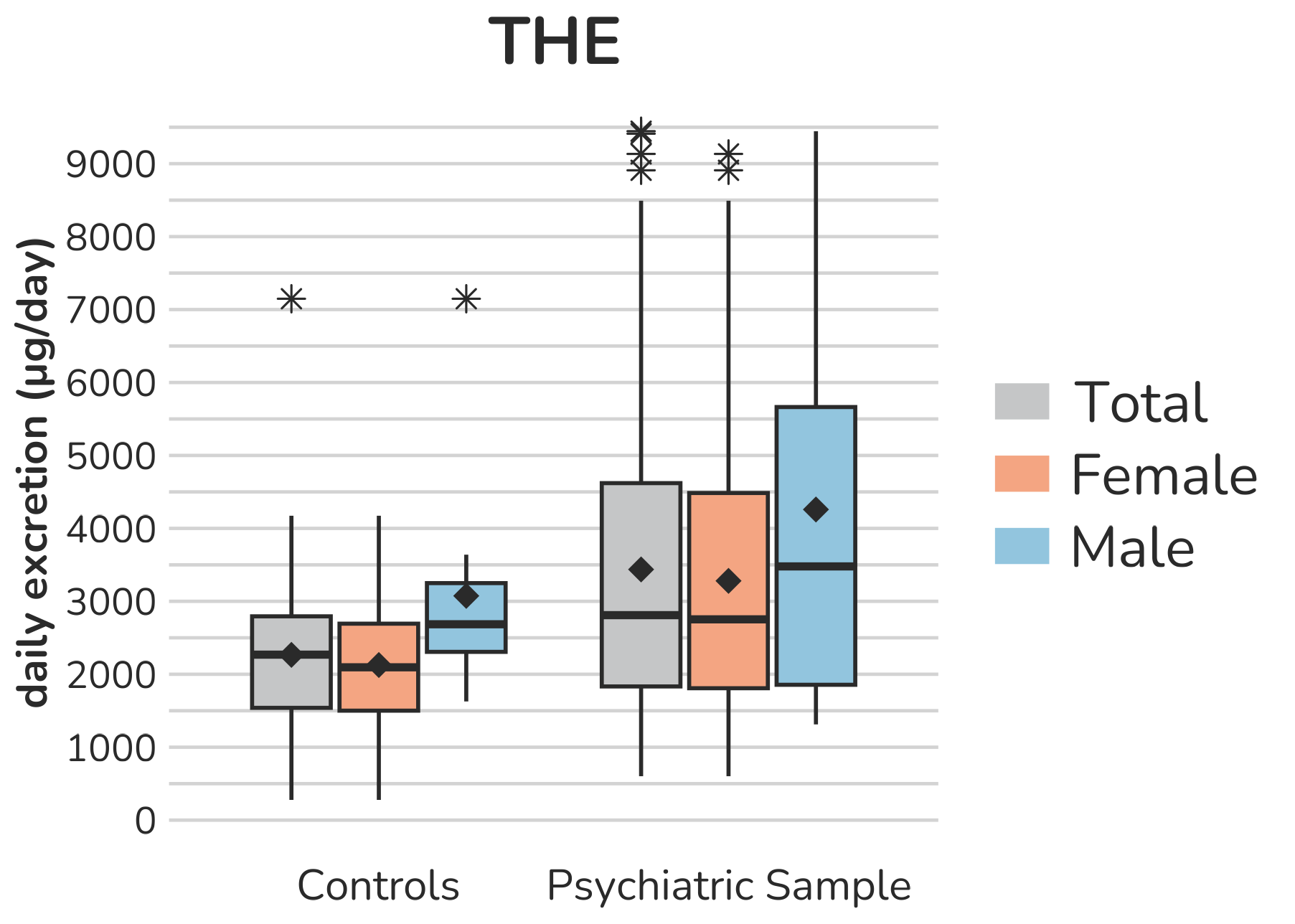

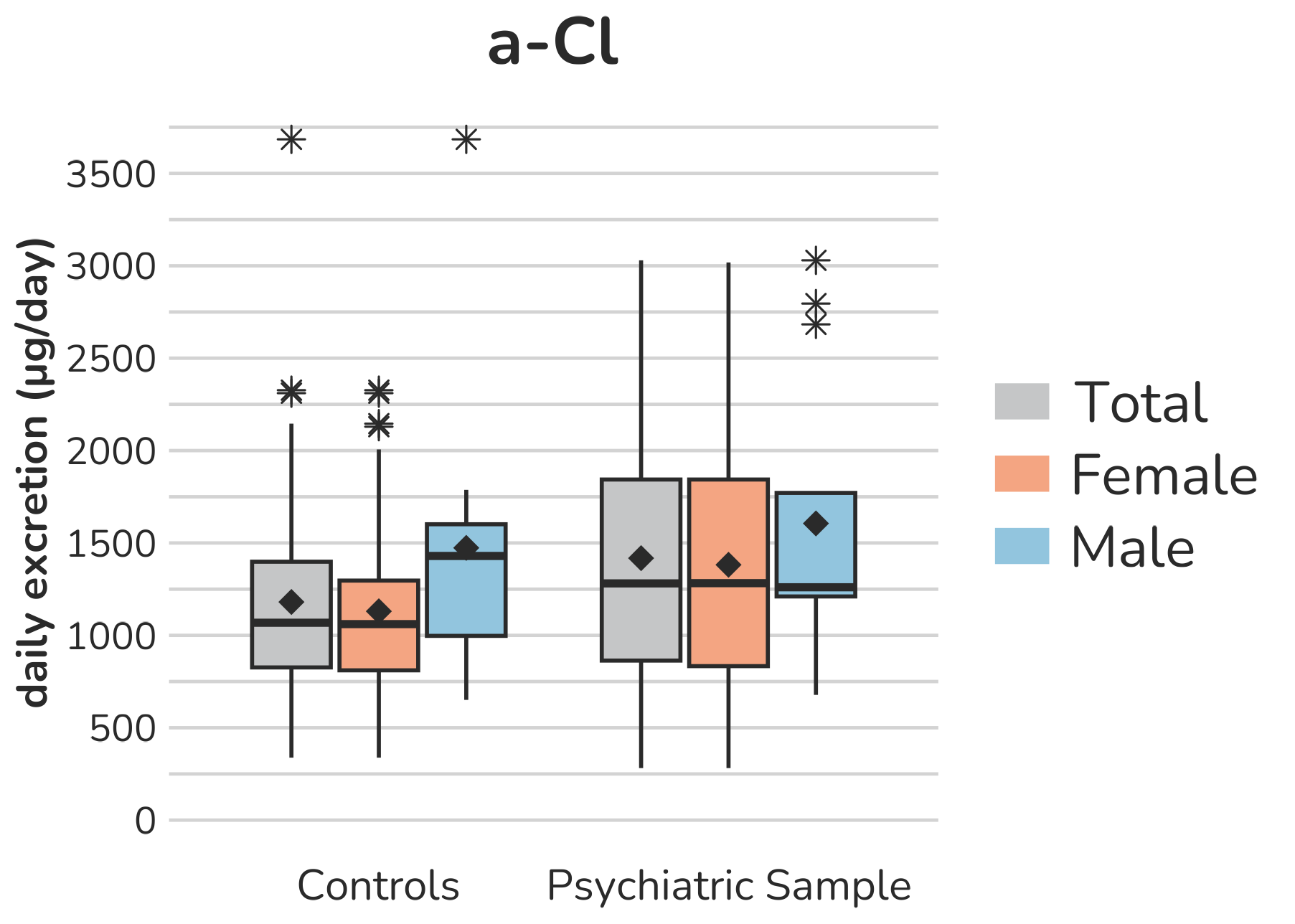

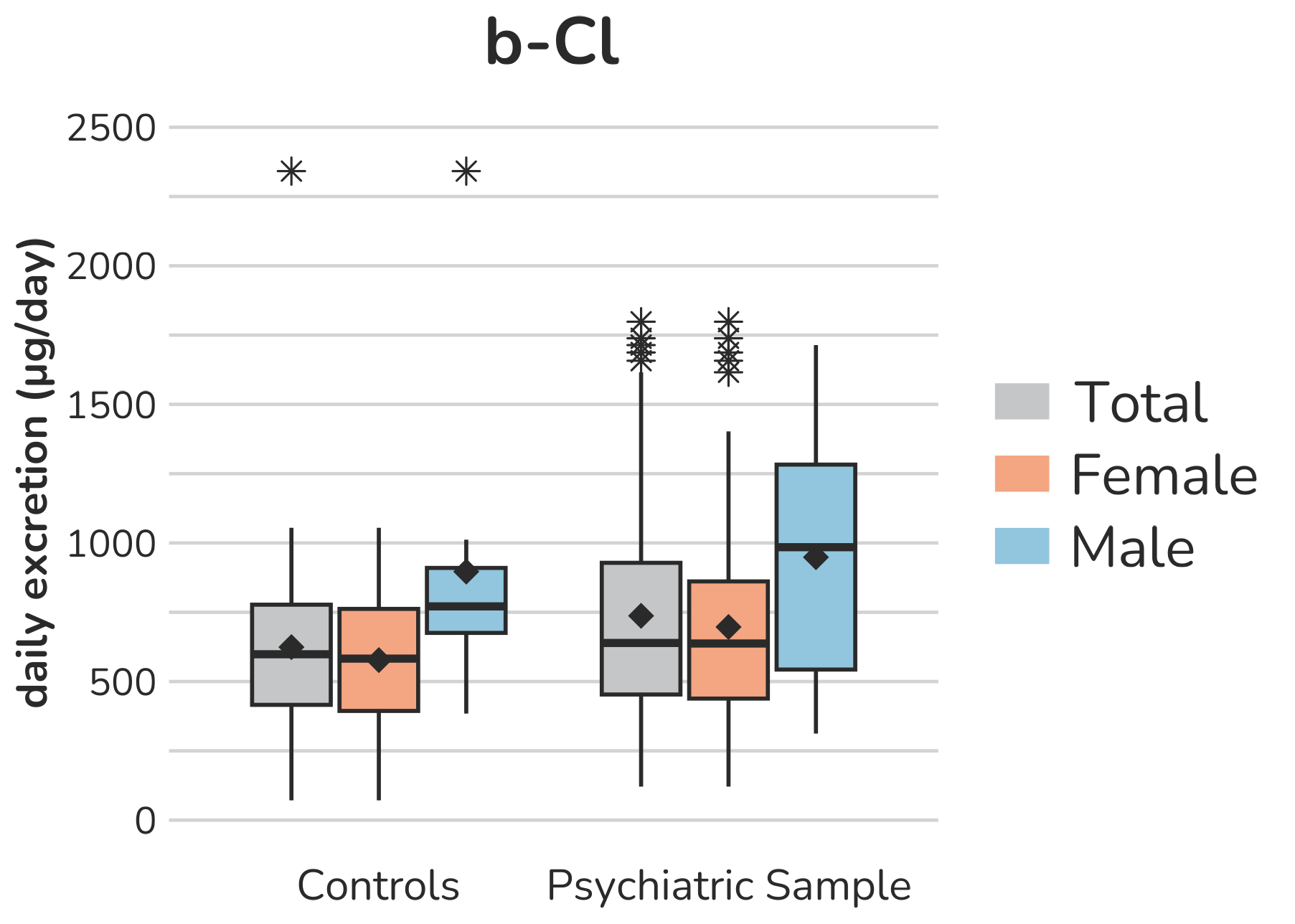

**Supplemental Figure S3 – Boxplots of glucocorticoid (C21-steroids) urine metabolites.** Each box gives the 25th and 75th percentile (lower and upper boundary of the box), the median (horizontal line within each box), and the mean (rhombus within each box). The whiskers are defined by 1.5 times the interquartile range. For F, two subject (both female, psychiatric sample) with a daily excretion of 806.6µg and 874.1µg were excluded from the figure to enhance the visualization of the remaining values. Aside from this, all outliers are displayed (illustrated by *).

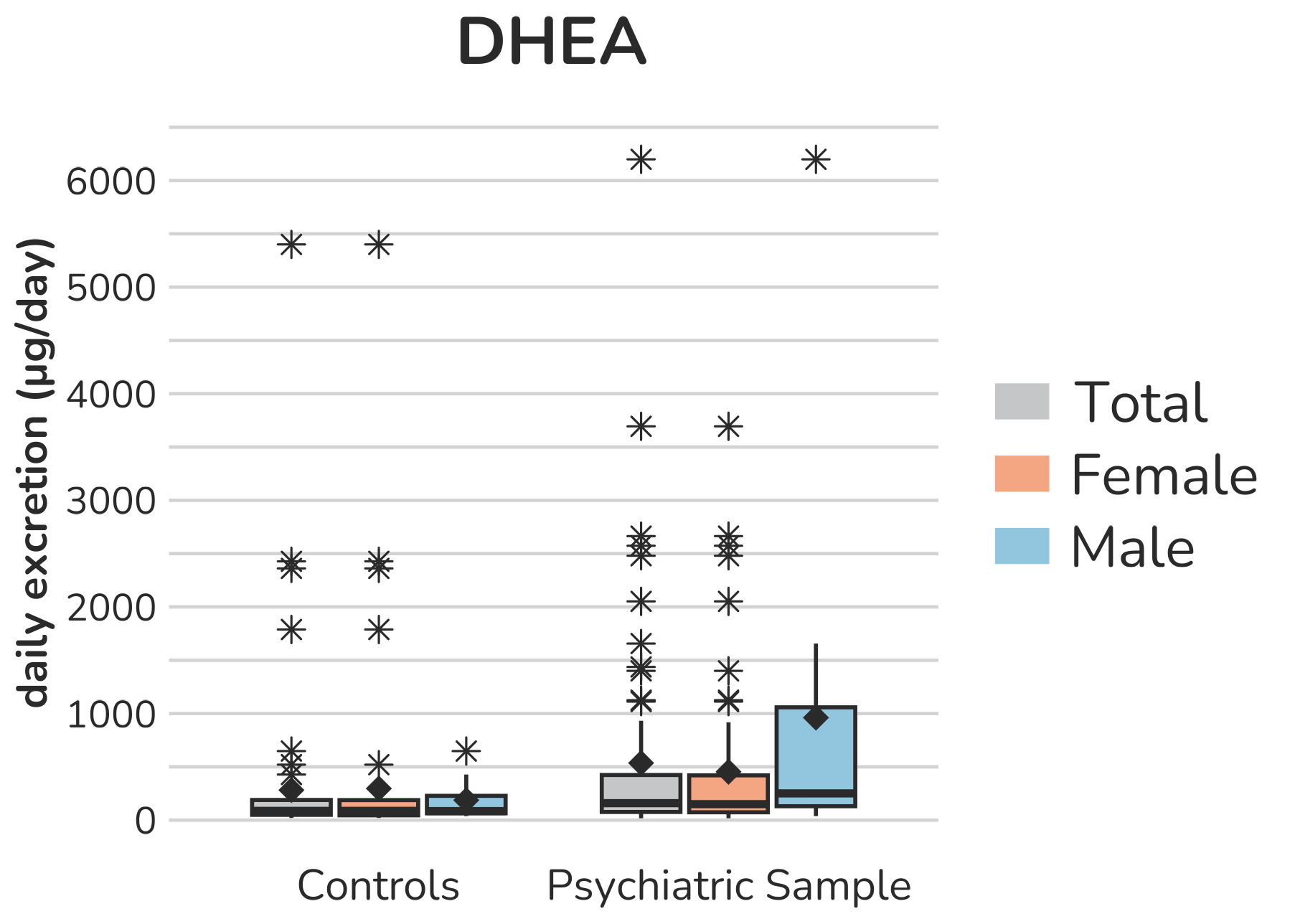

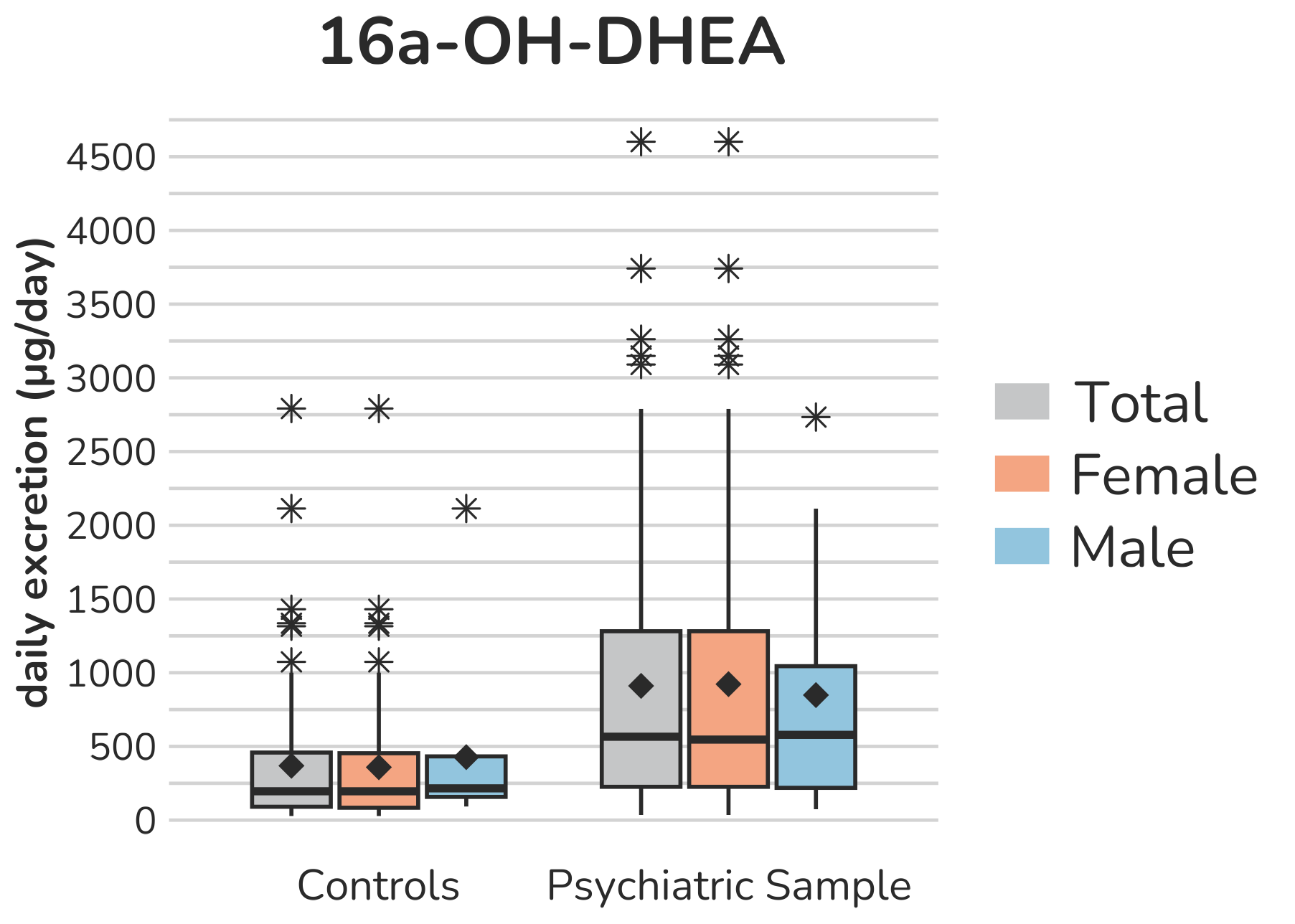

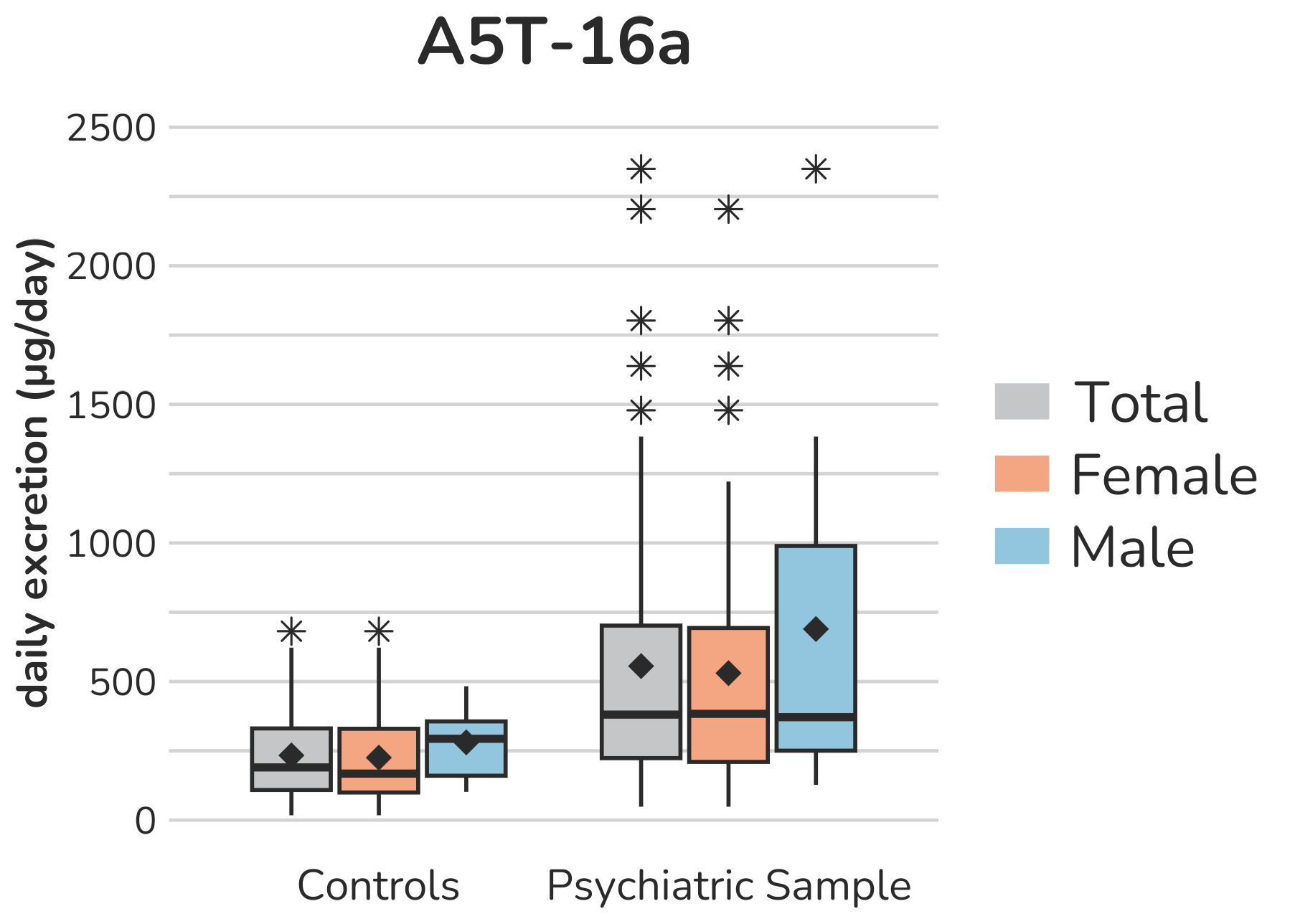

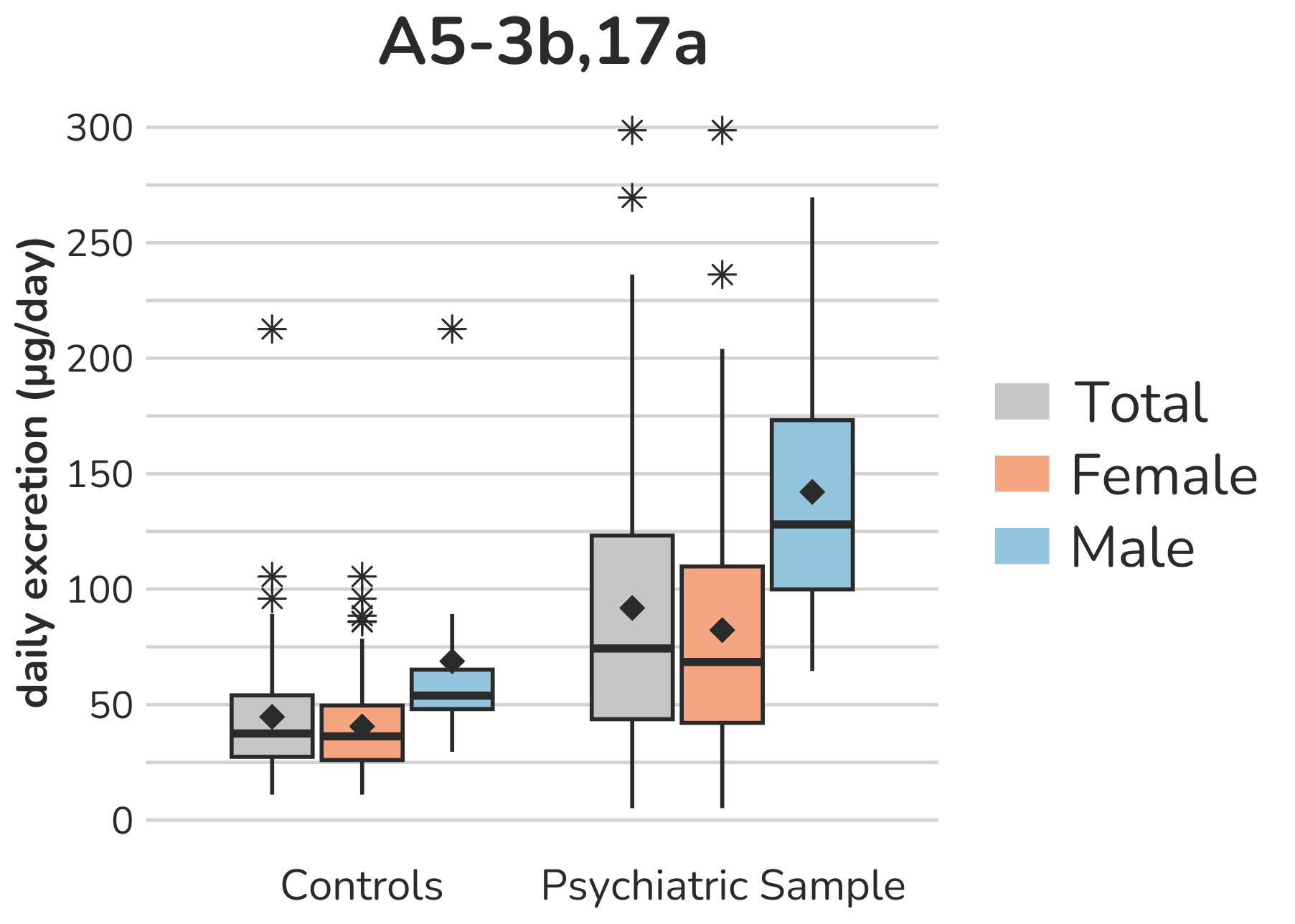

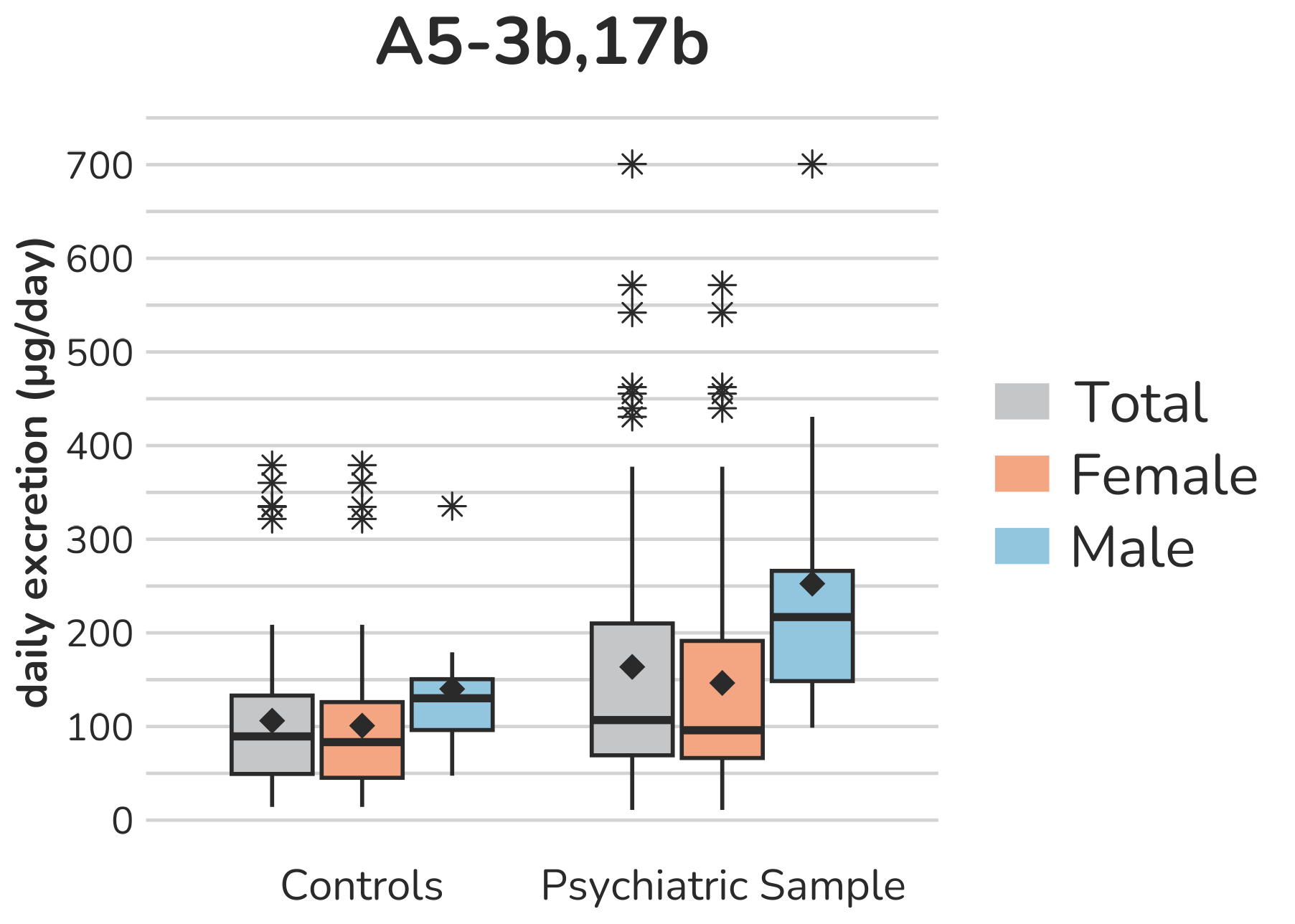

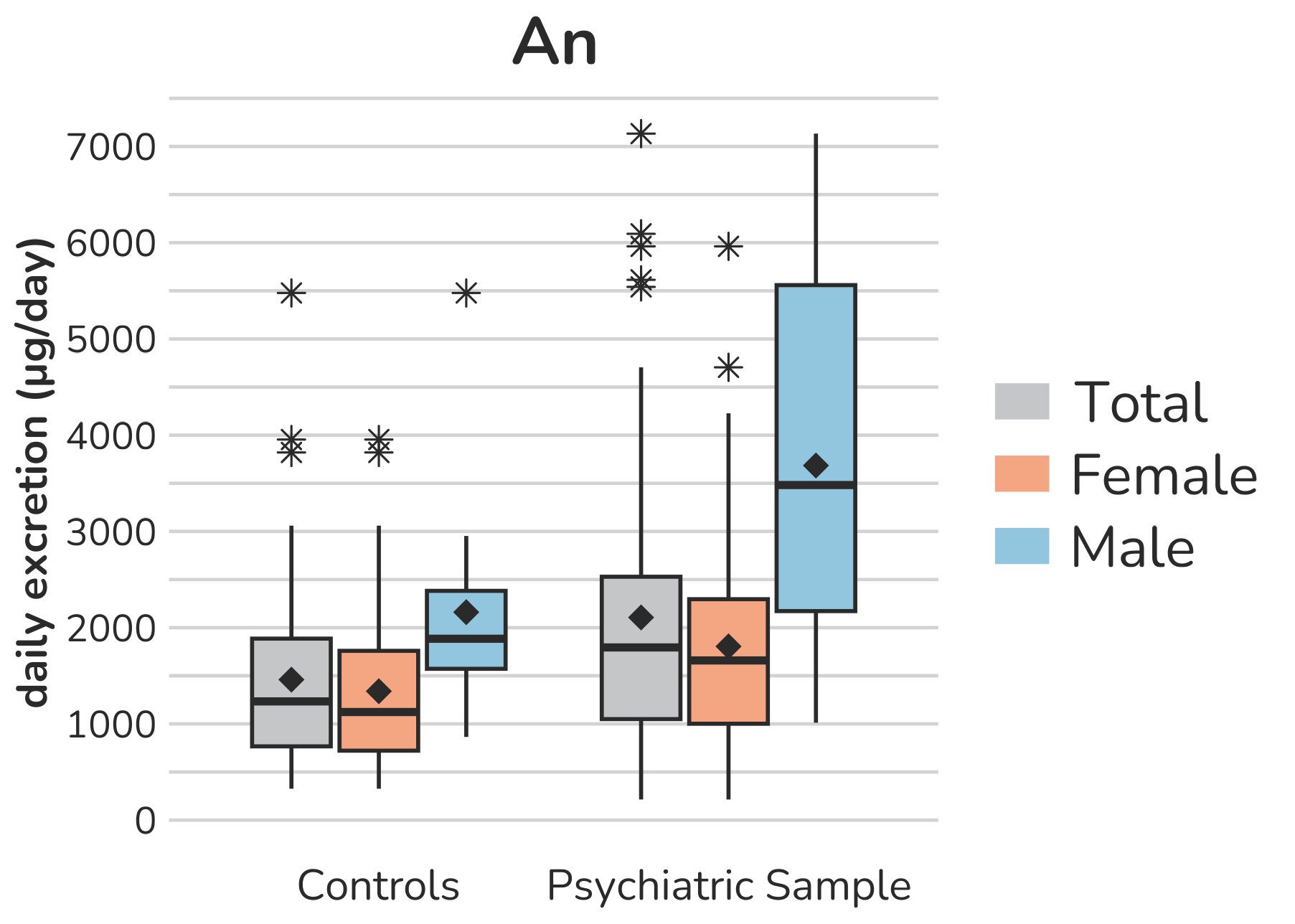

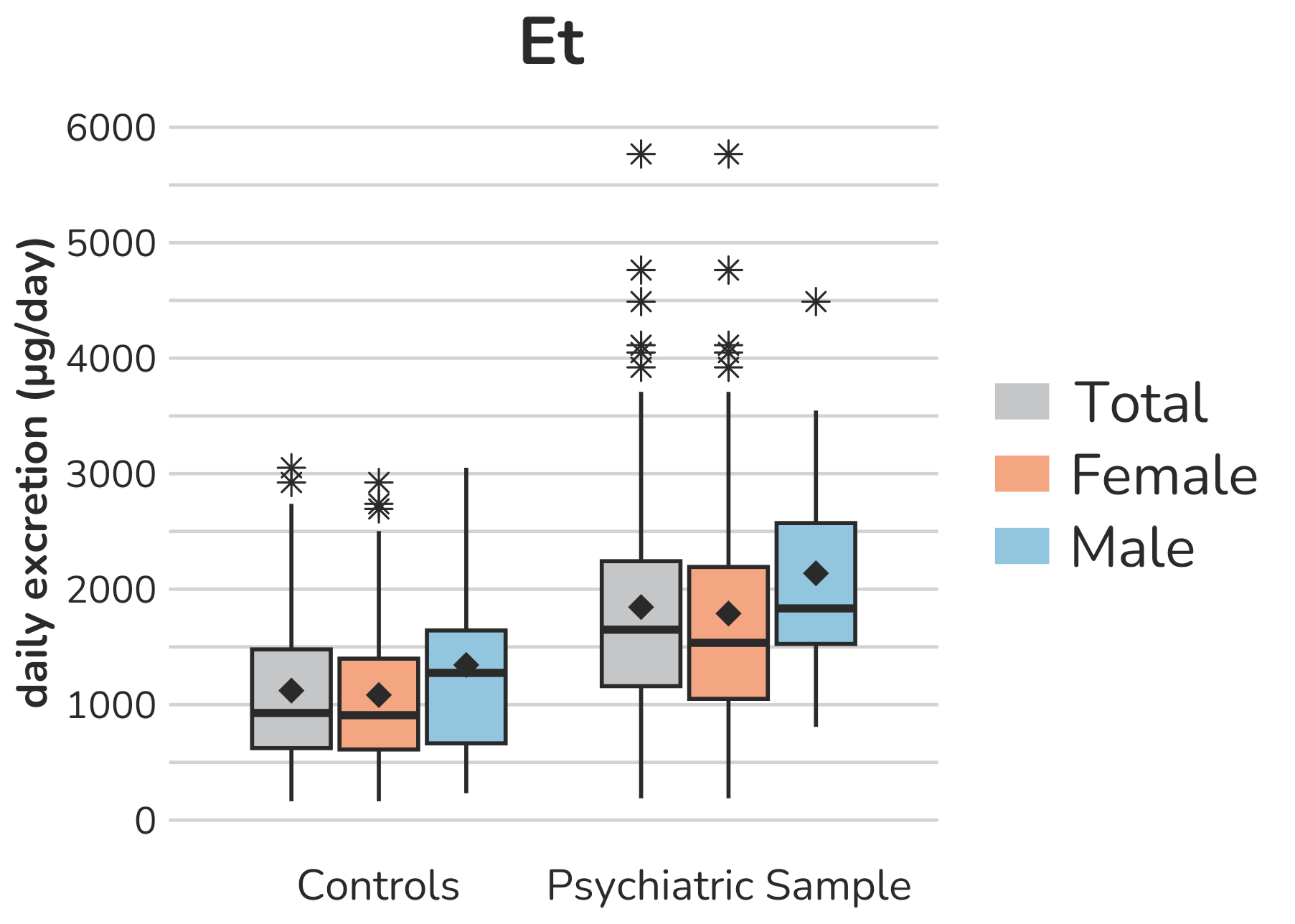

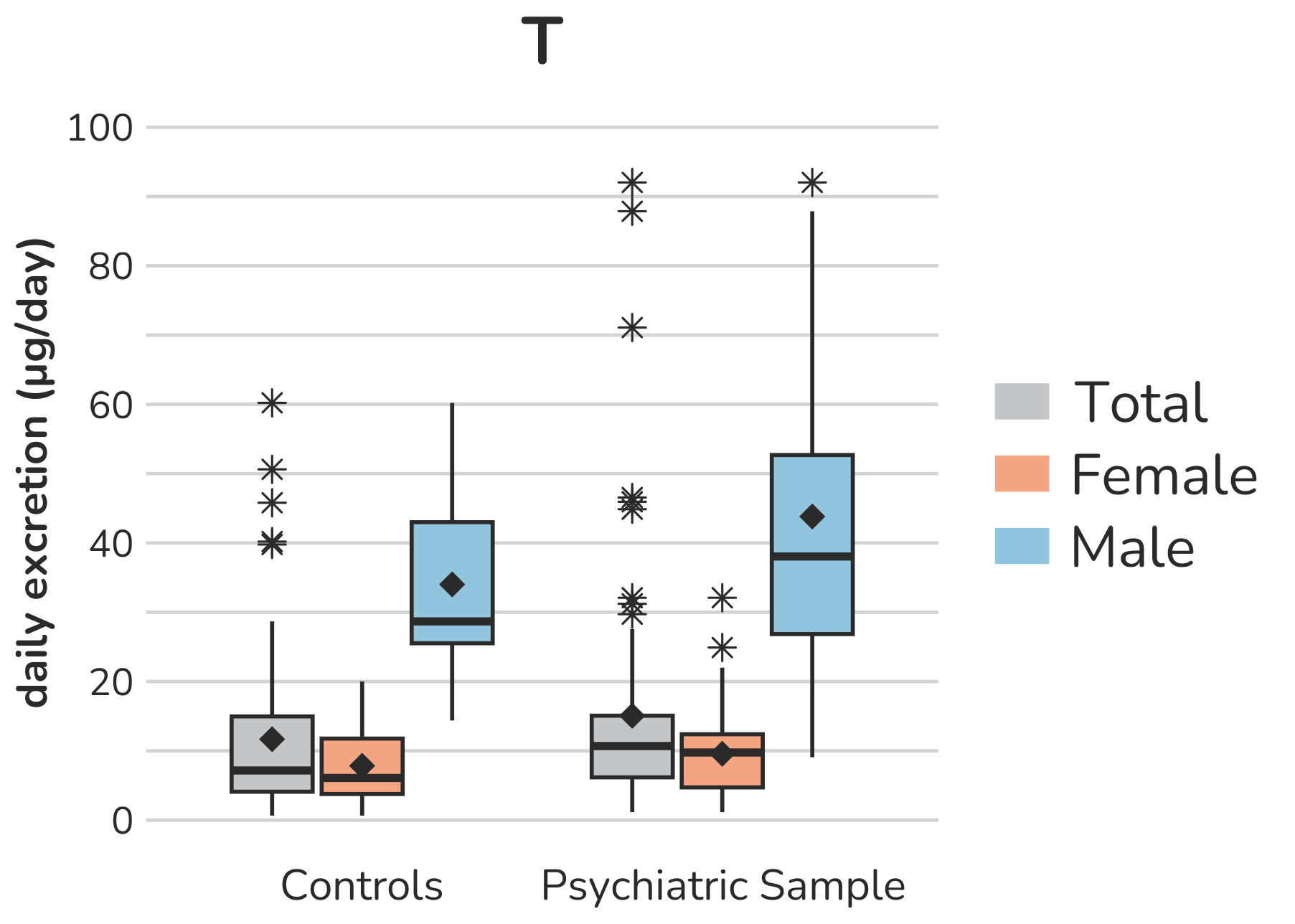

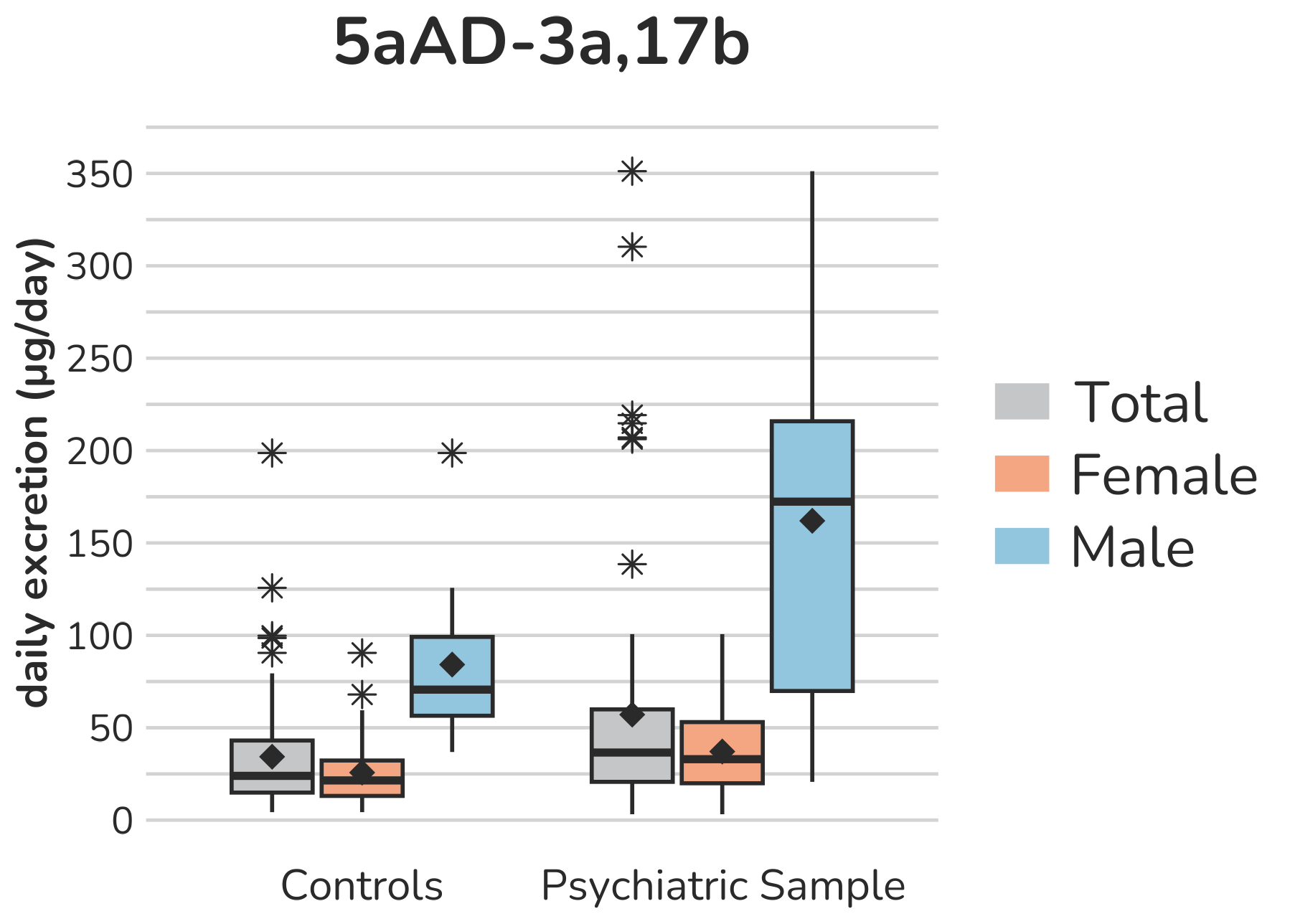

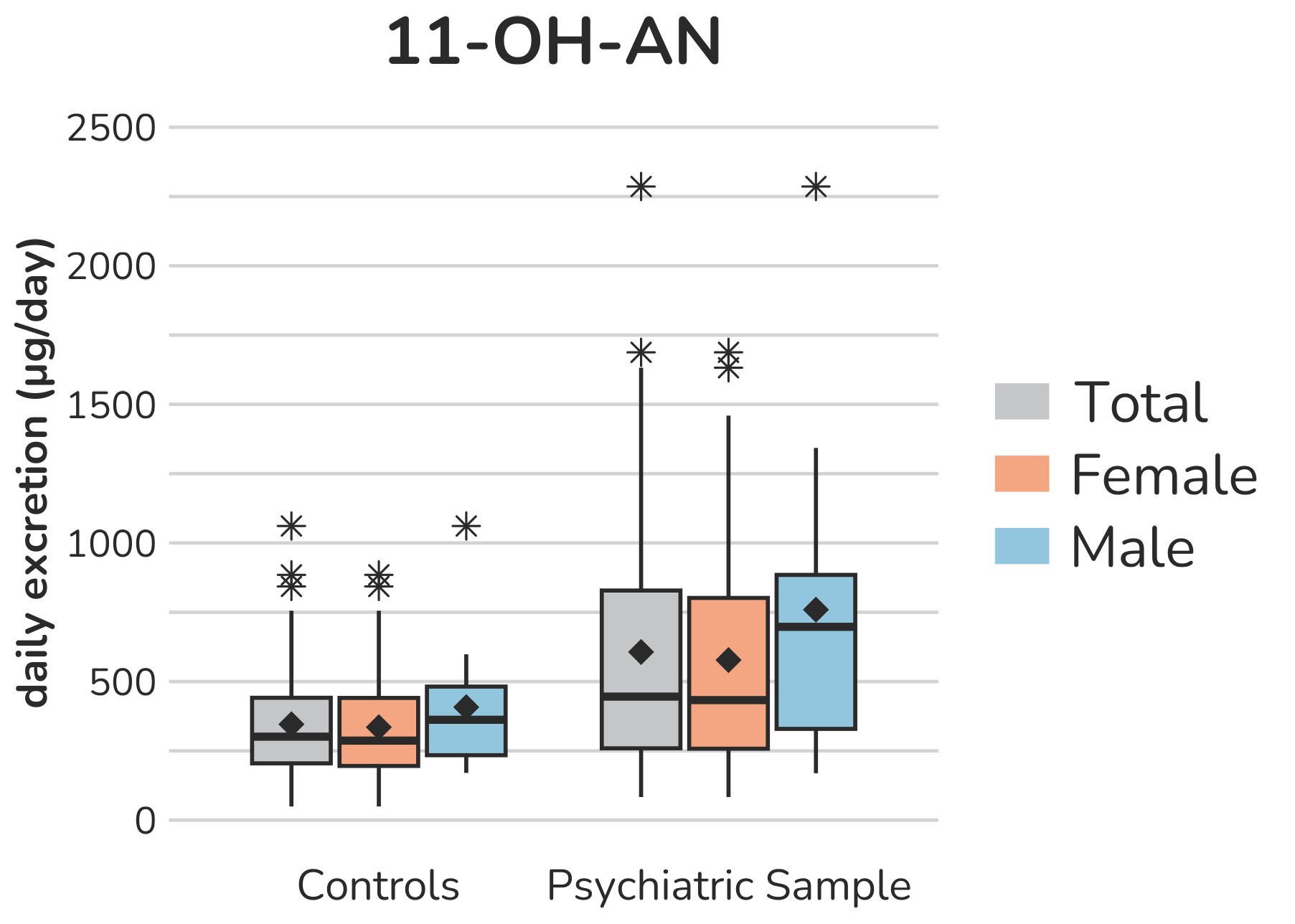

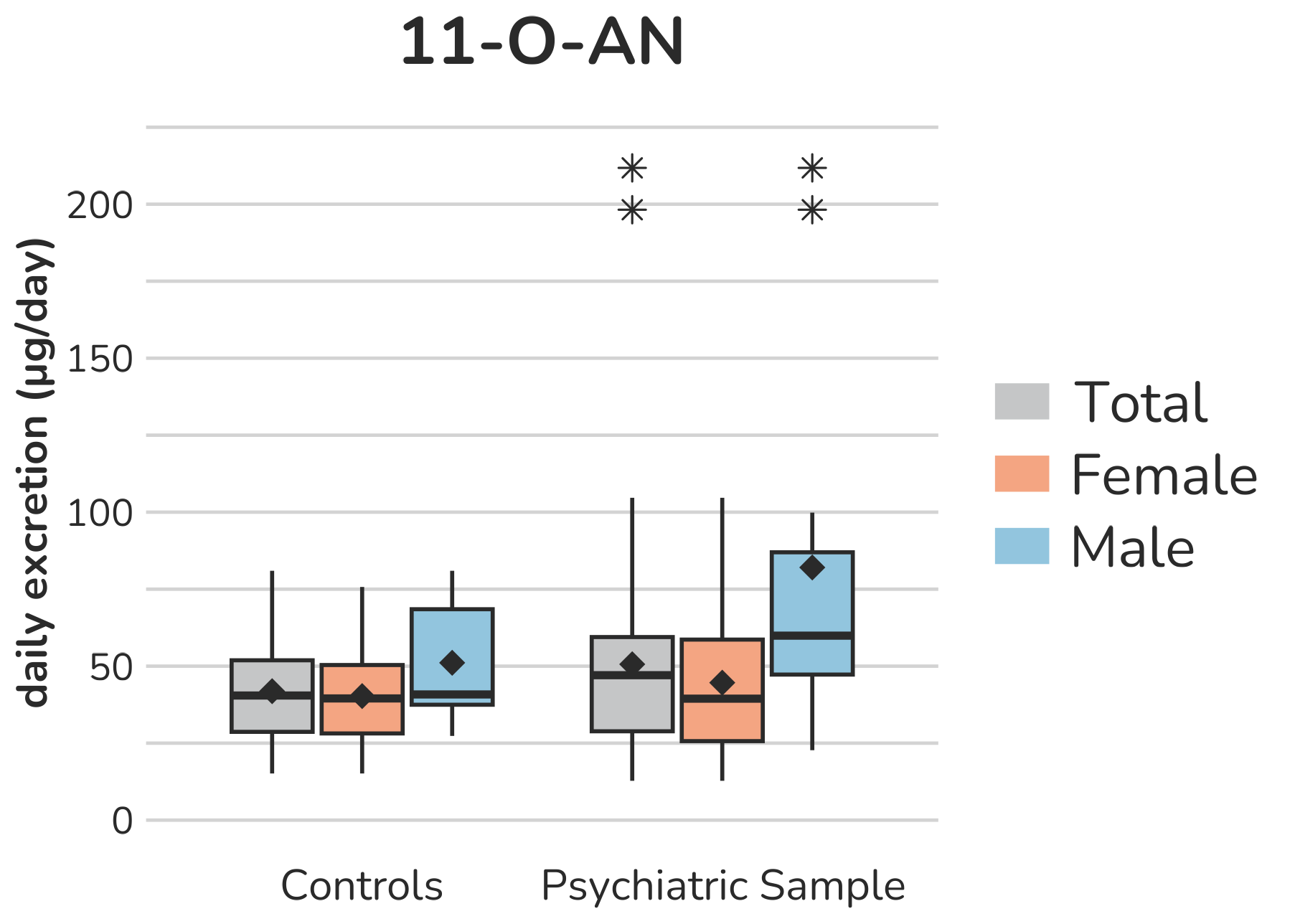

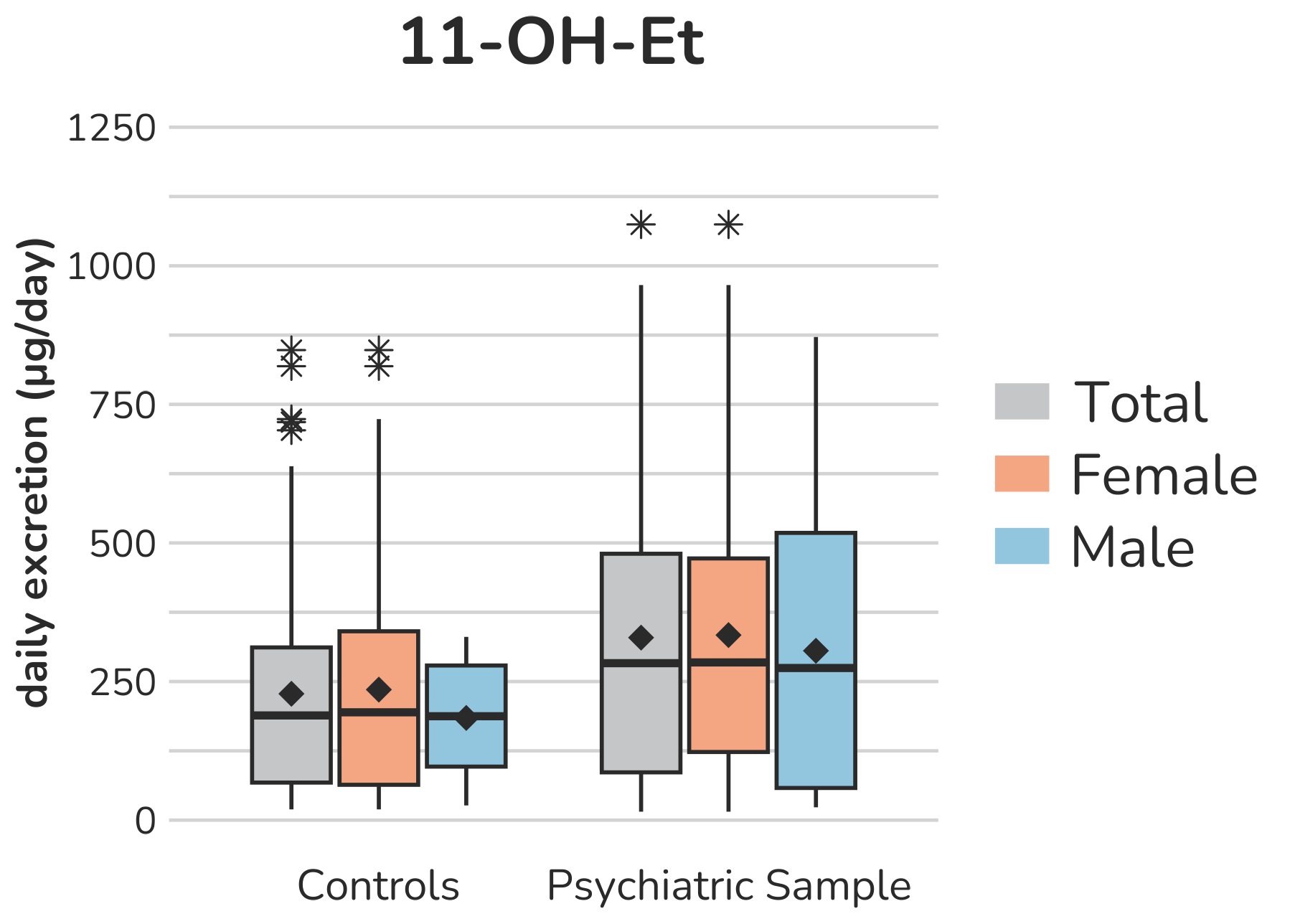

**Supplemental Figure S4 – Boxplots of androgen (C19-steroids) urine metabolites.** Each box gives the 25th and 75th percentile (lower and upper boundary of the box), the median (horizontal line within each box), and the mean (rhombus within each box). The whiskers are defined by 1.5 times the interquartile range. For DHEA, two subjects with a daily excretion of 15,603µg (female, psychiatric sample) and 16,052µg (male, control group) were excluded from the figure to enhance the visualization of the remaining values. For the same reason, for A5-3β,17β the same two subject with a daily excretion of 1474.2µg (female, psychiatric sample) and 1294.9µg (male, control group) are not presented. For 11-OH-Et, one subject (female, psychiatric sample) with a daily excretion of 2381.5µg is not shown. Aside from this, all outliers are displayed (illustrated by *).

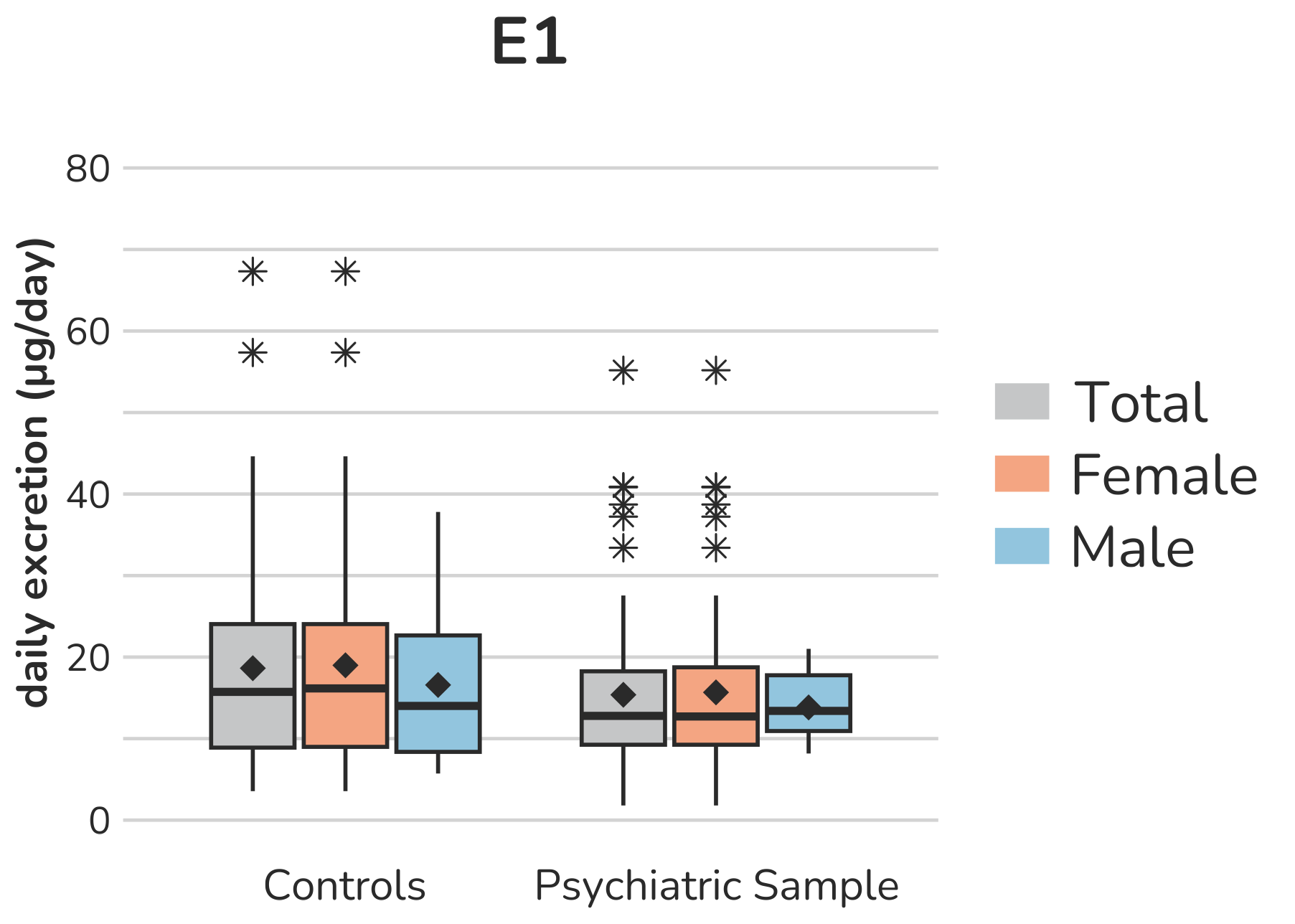

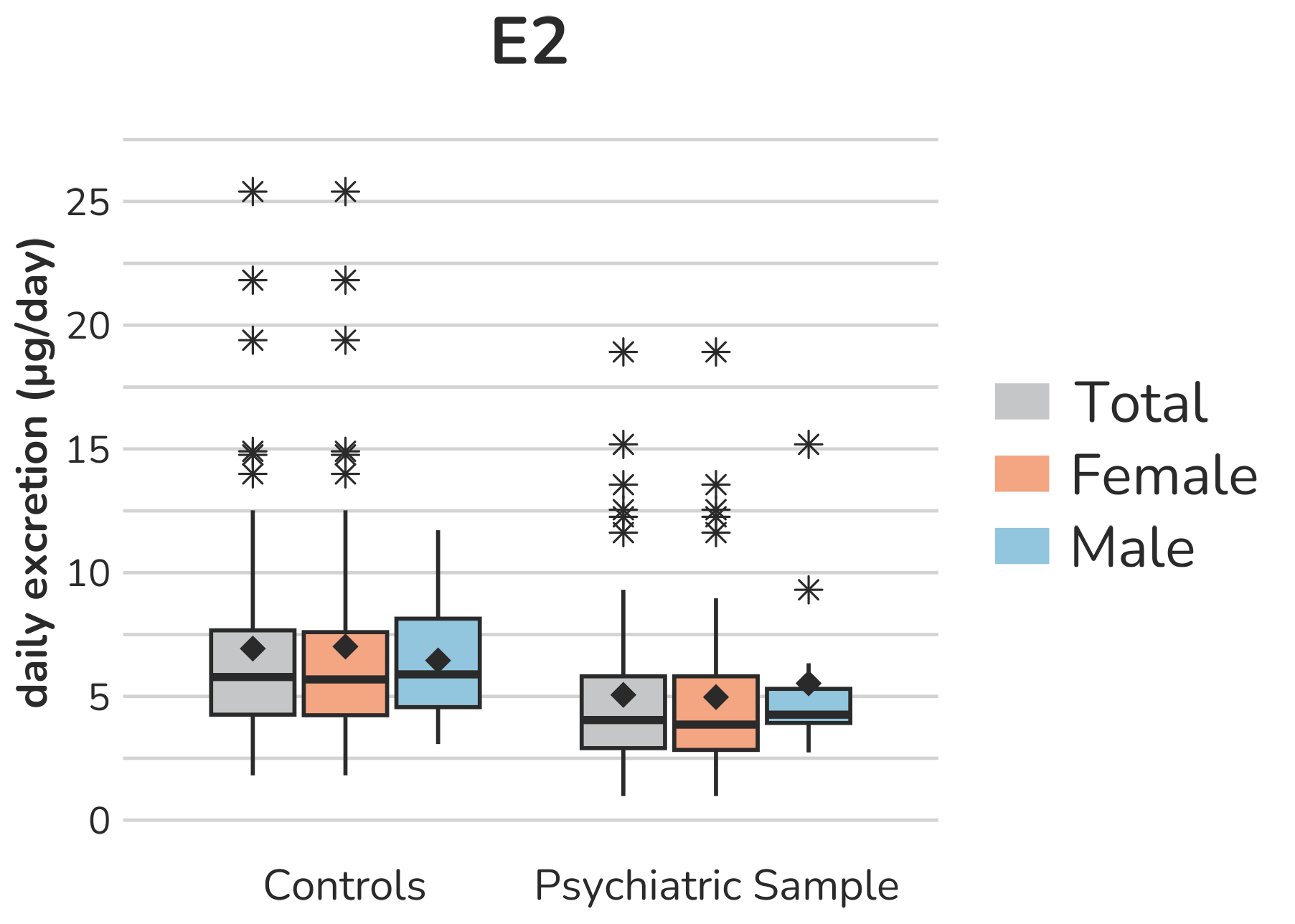

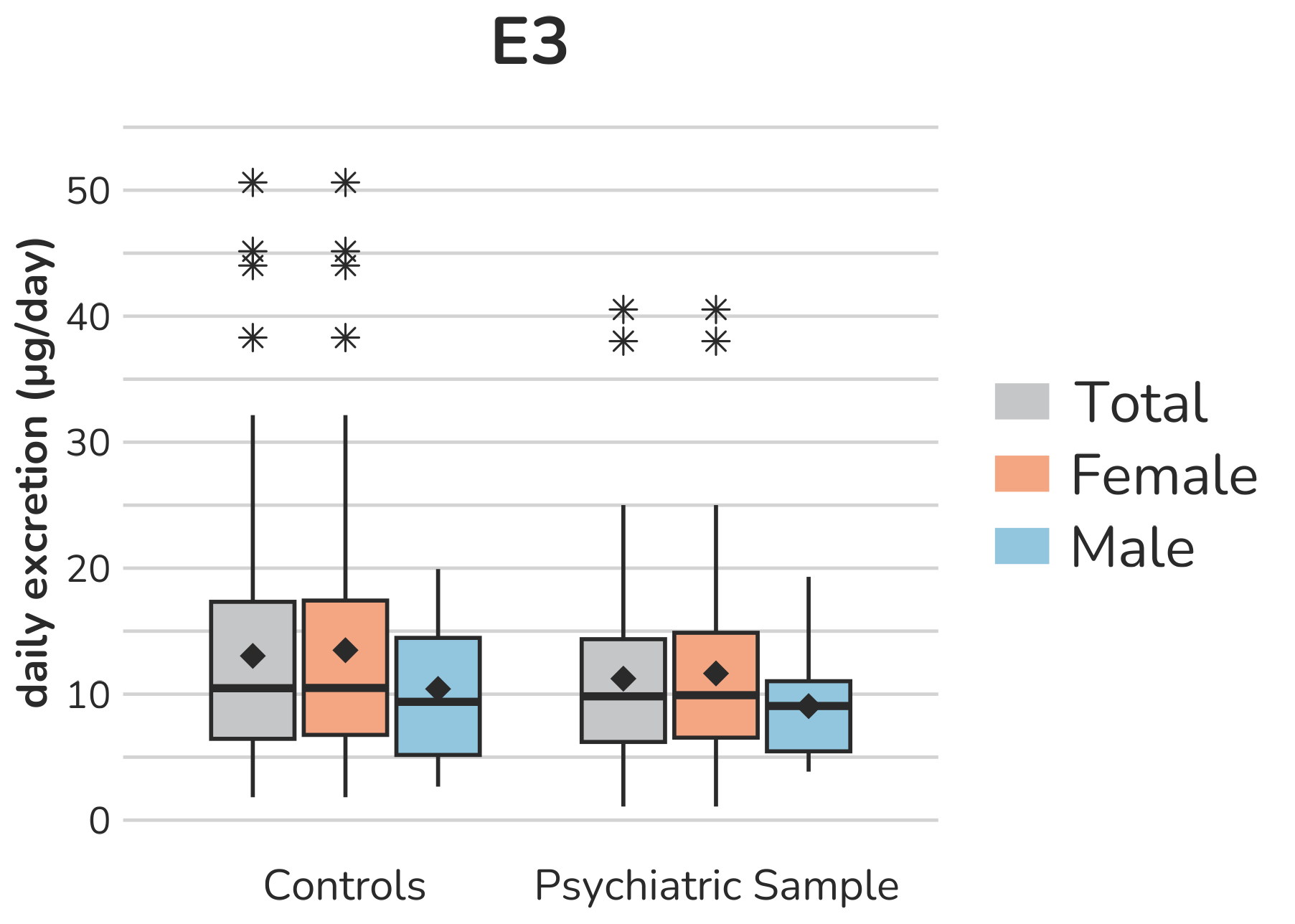

**Supplemental Figure S5 –** **Boxplots of estrogen (C18-steroids) urine metabolites.** Each box gives the 25th and 75th percentile (lower and upper boundary of the box), the median (horizontal line within each box), and the mean (rhombus within each box). The whiskers are defined by 1.5 times the interquartile range. For E1, one female control subject with a daily excretion of 199.1µg is excluded from the figure to enhance the visualization of the remaining values. Aside from this, all outliers are displayed (illustrated by *).

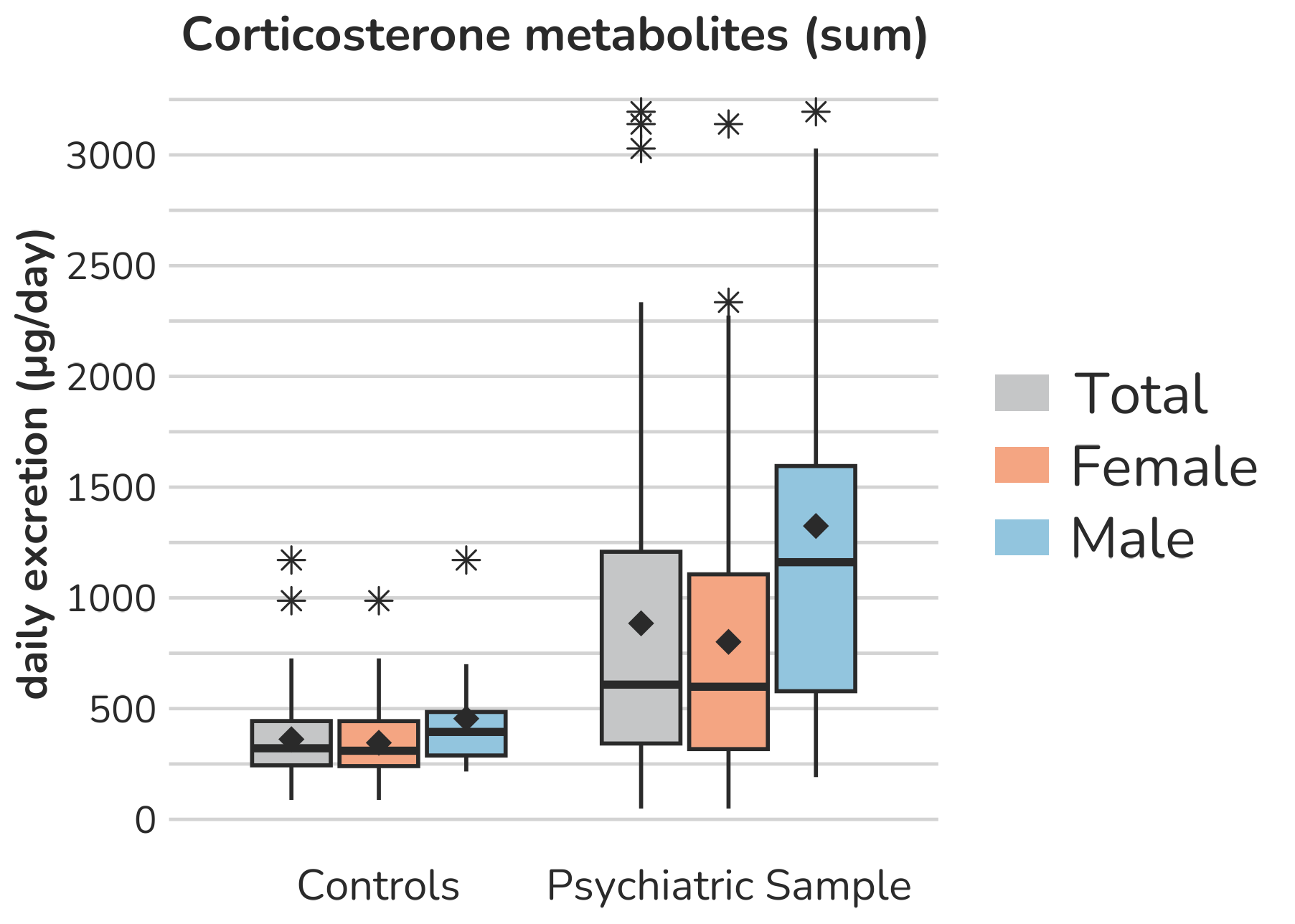

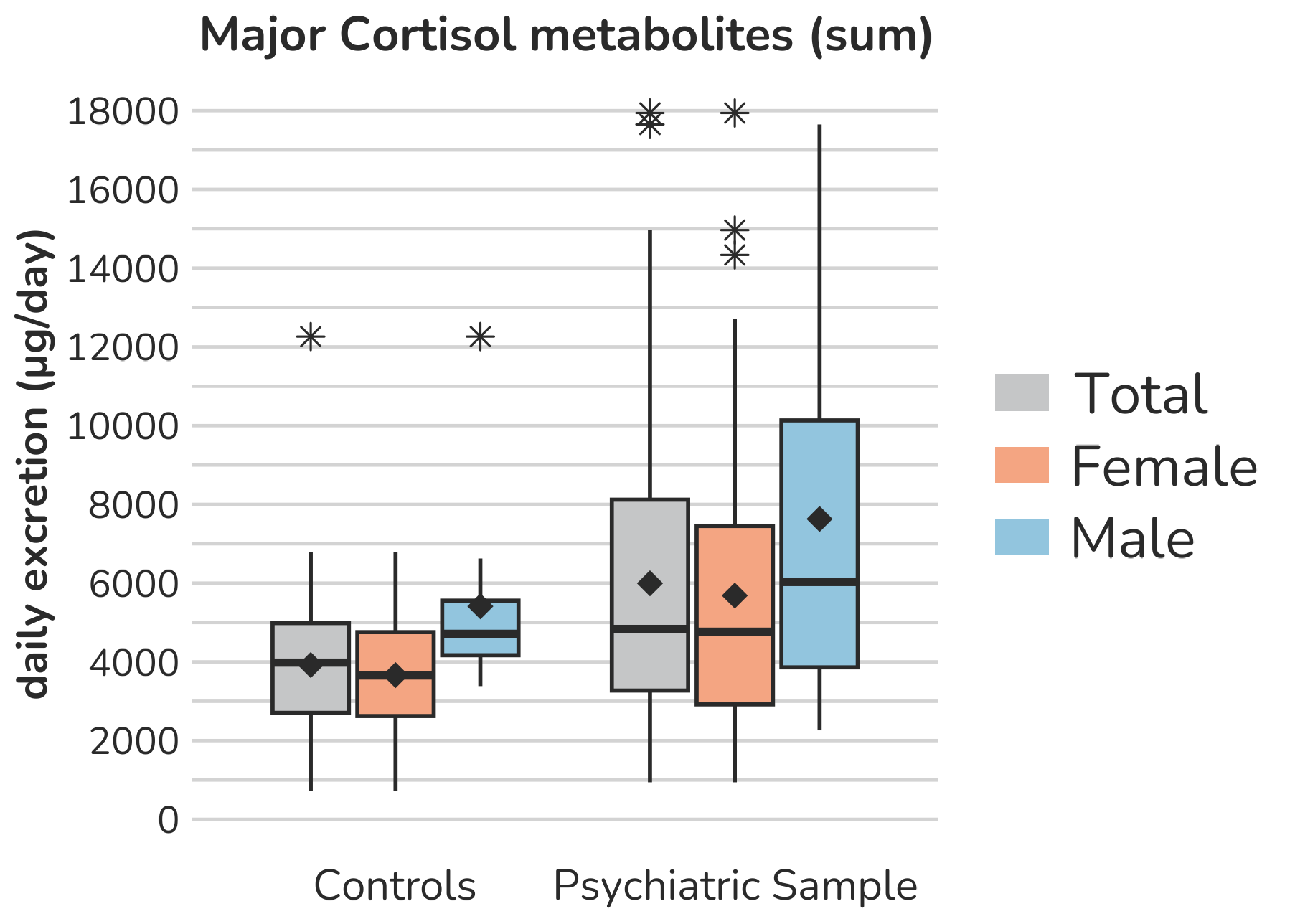

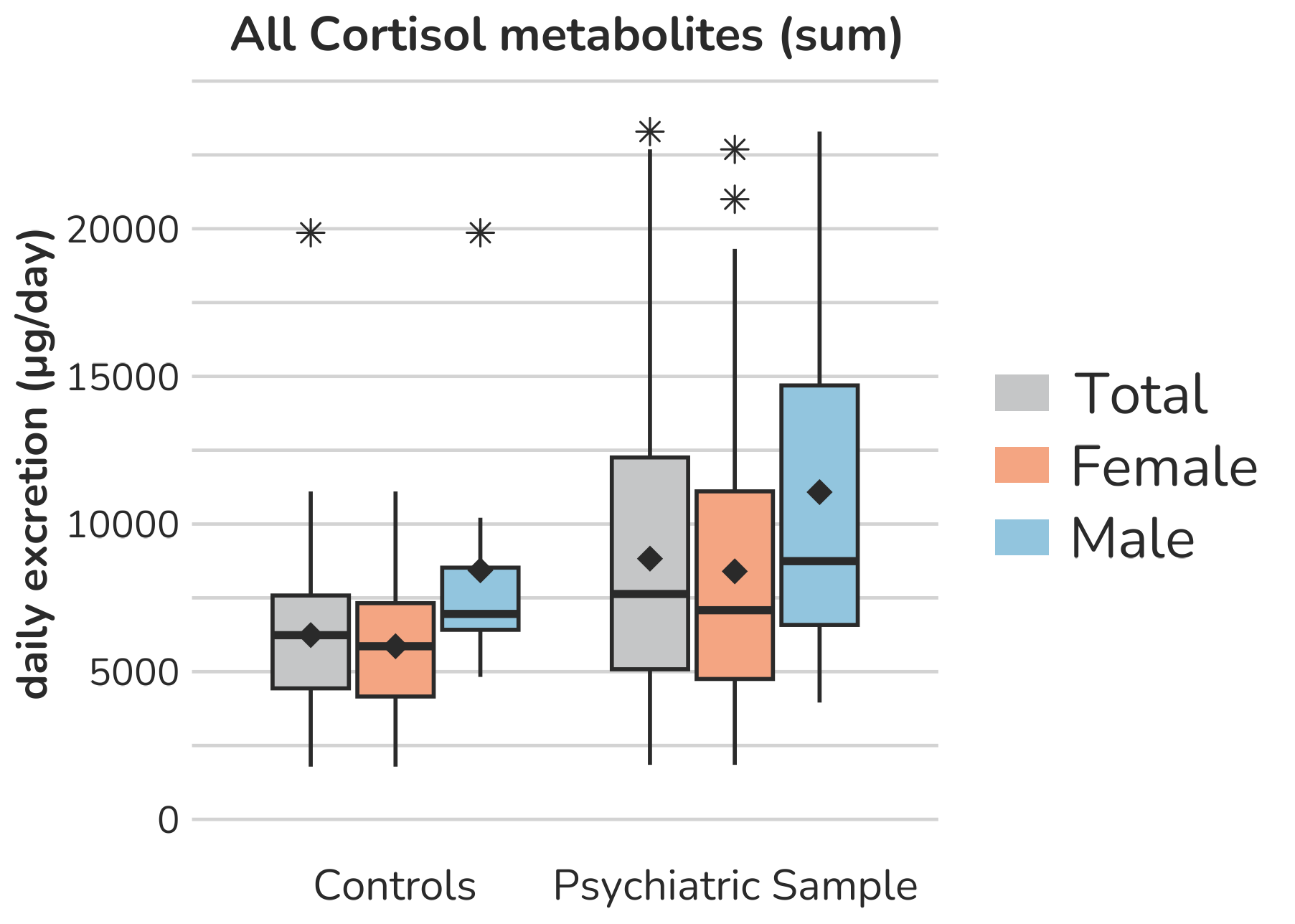

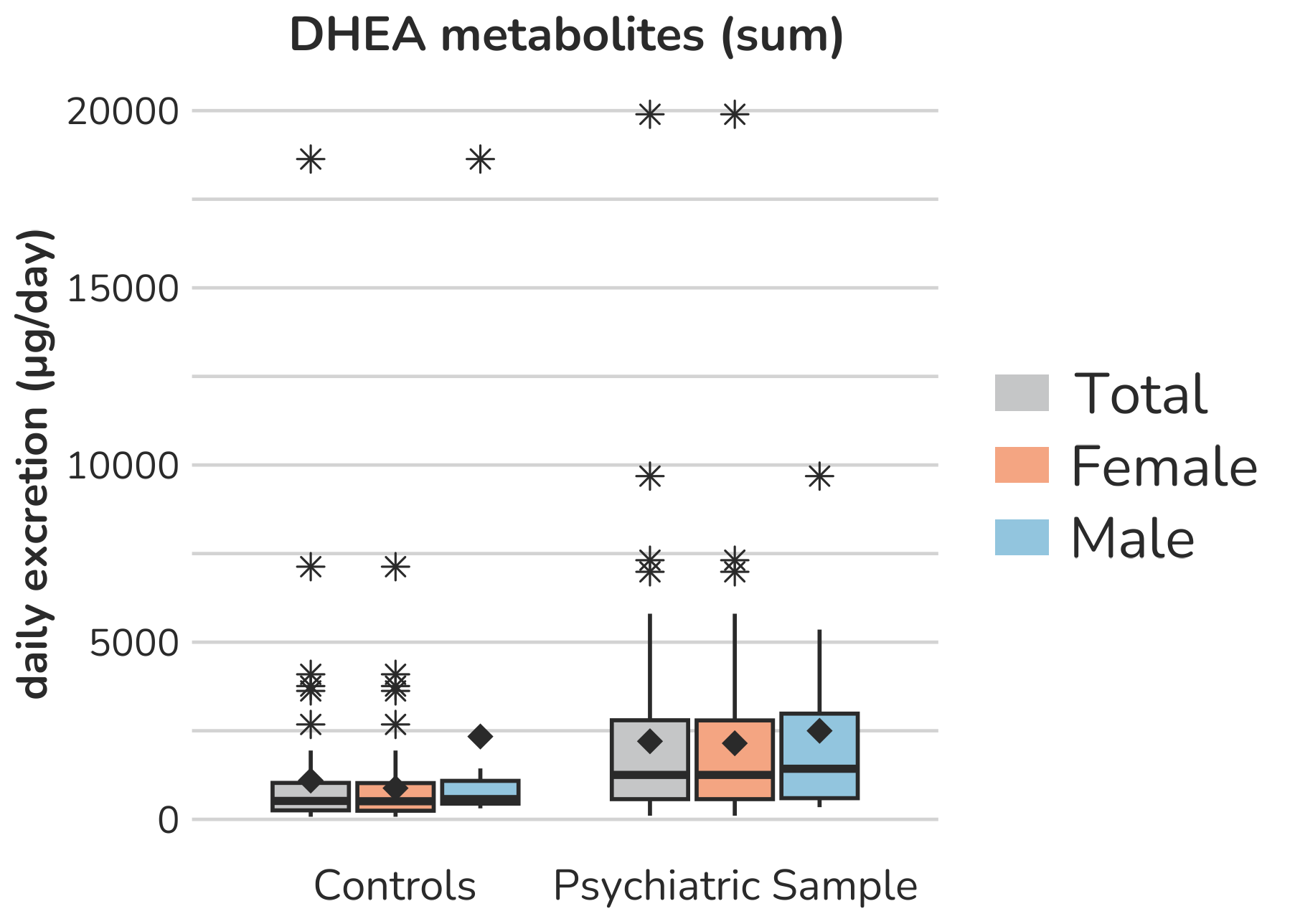

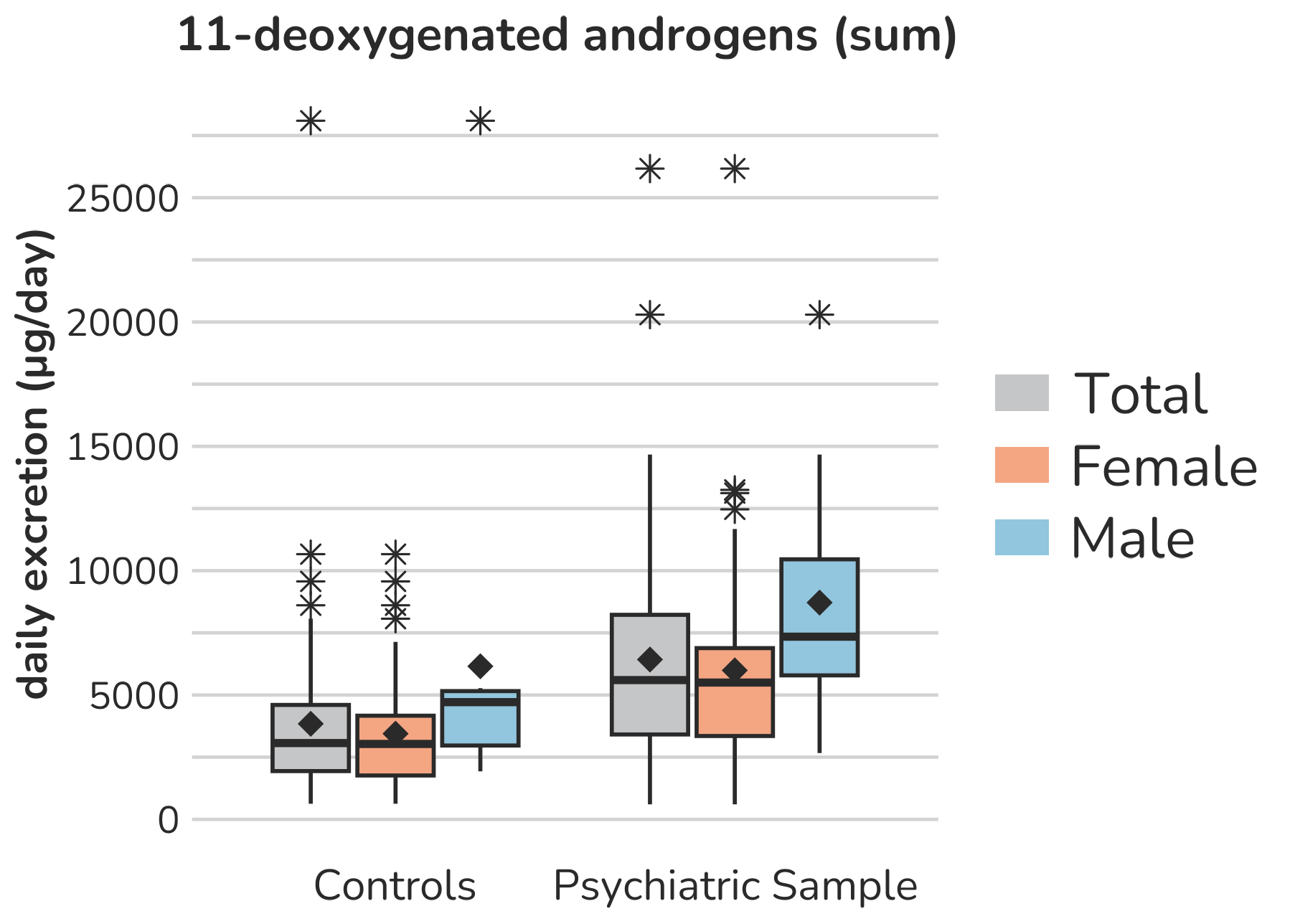

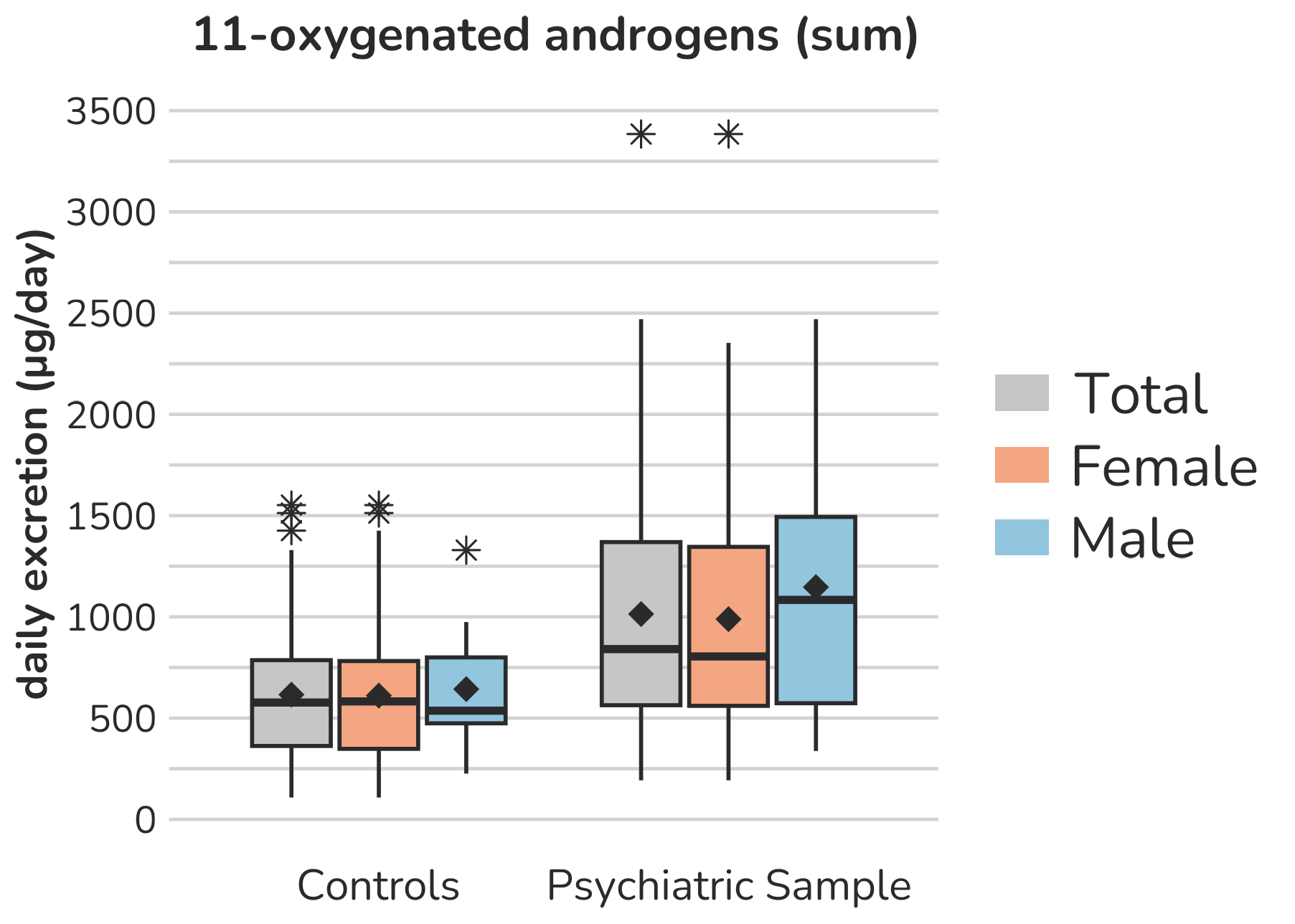

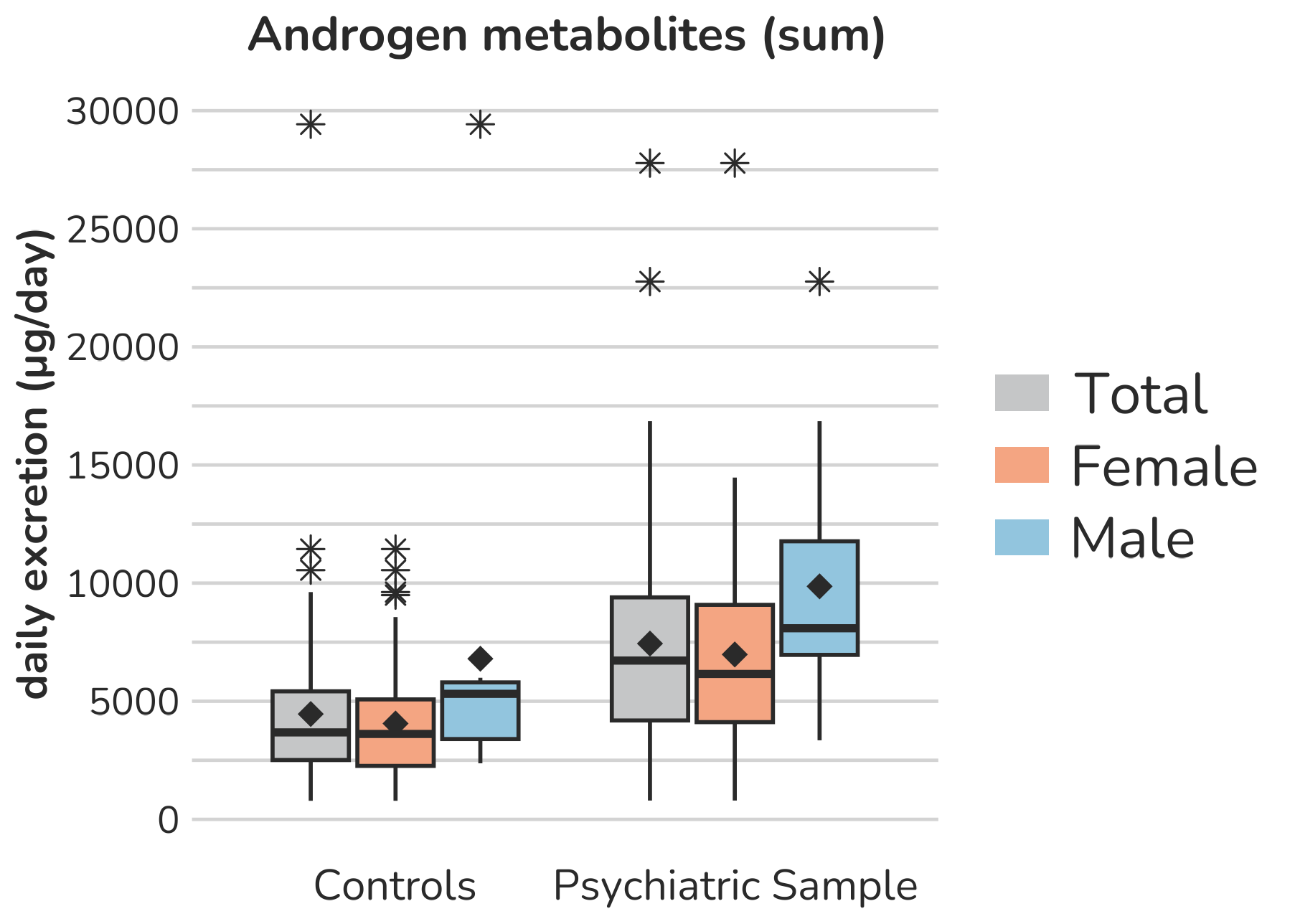

**Supplemental Figure S6 –** **Boxplots of summarized steroid urine metabolites.** Each box gives the 25th and 75th percentile (lower and upper boundary of the box), the median (horizontal line within each box), and the mean (rhombus within each box). The whiskers are defined by 1.5 times the interquartile range. All outliers are presented (illustrated by *).

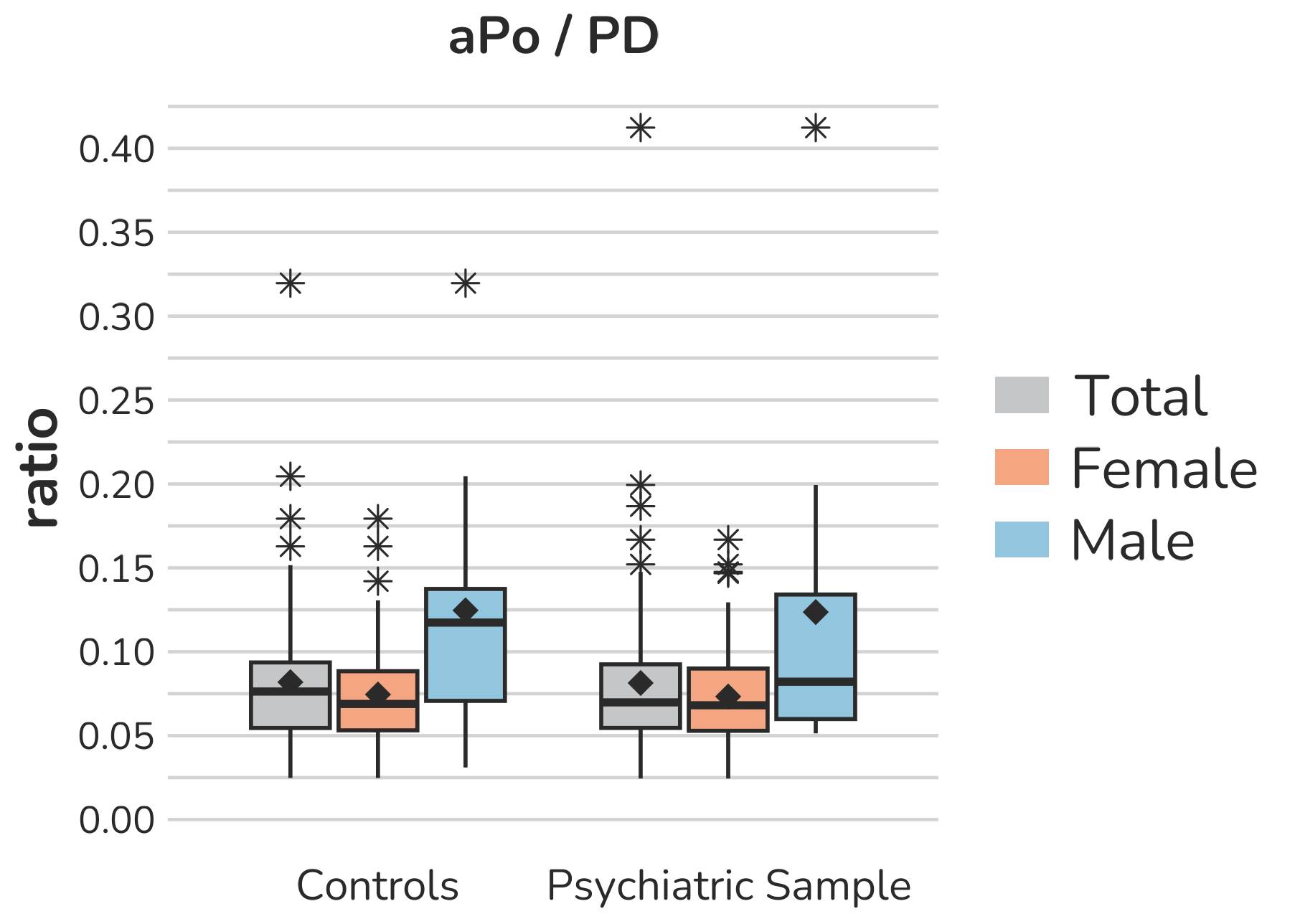

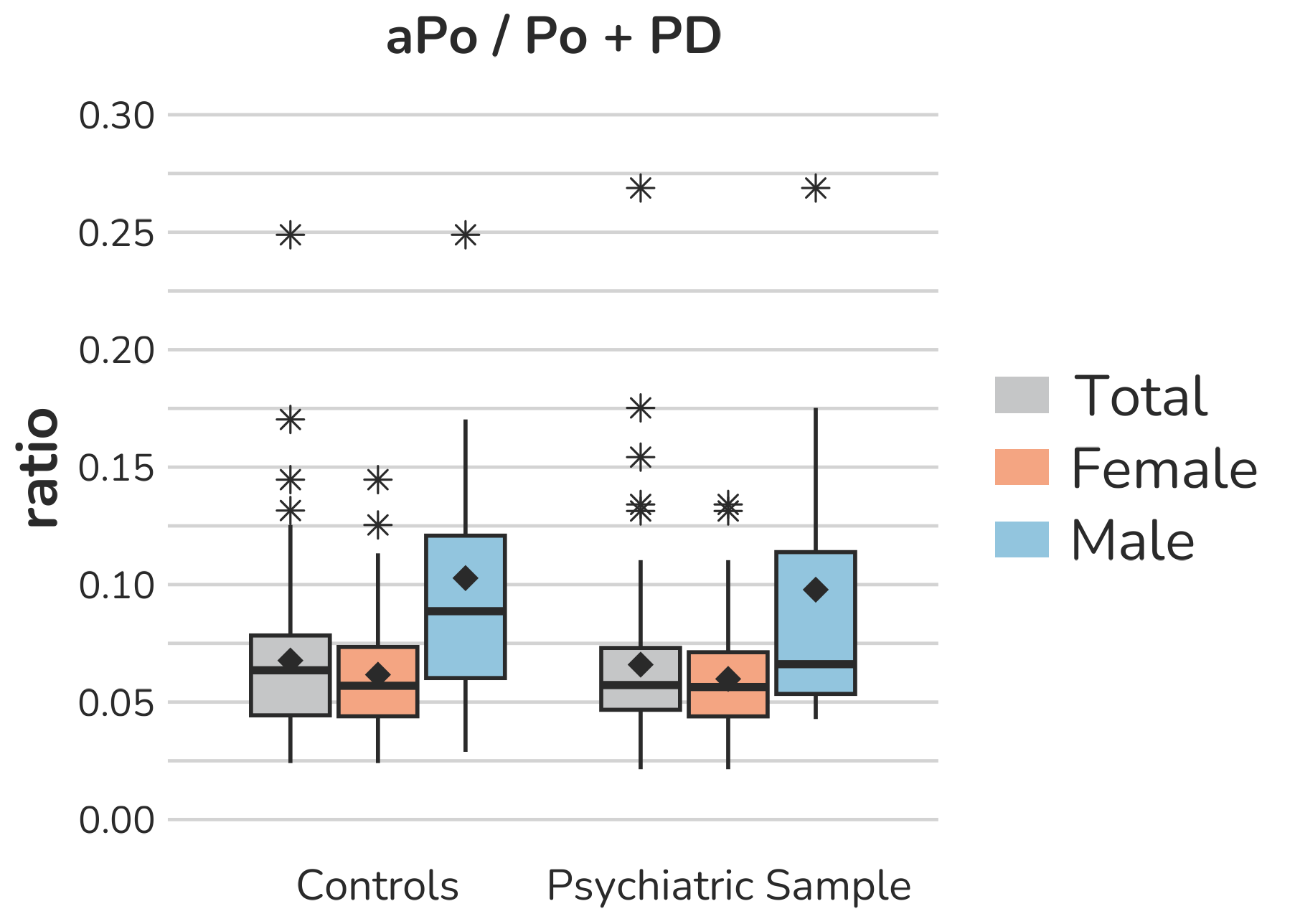

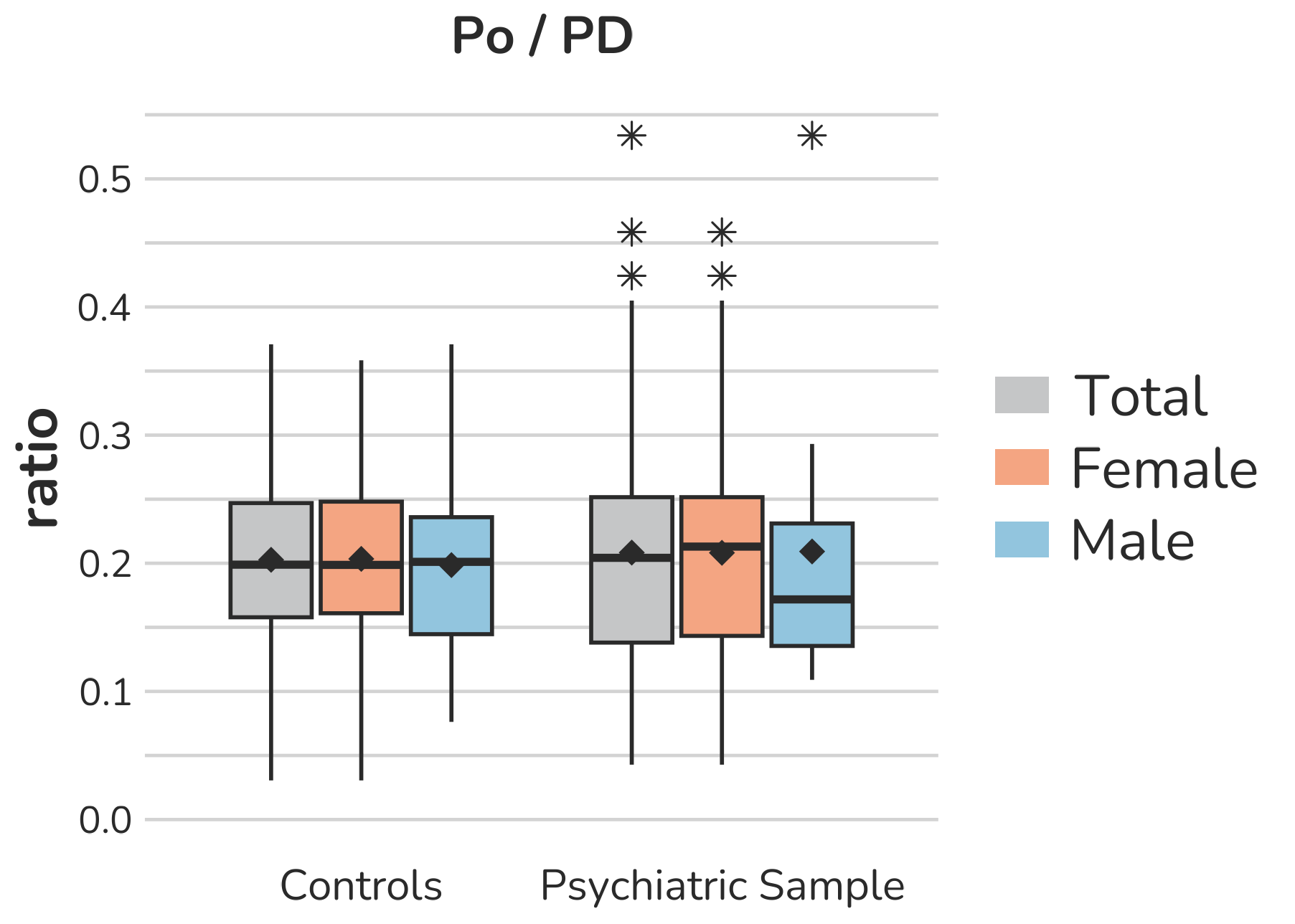

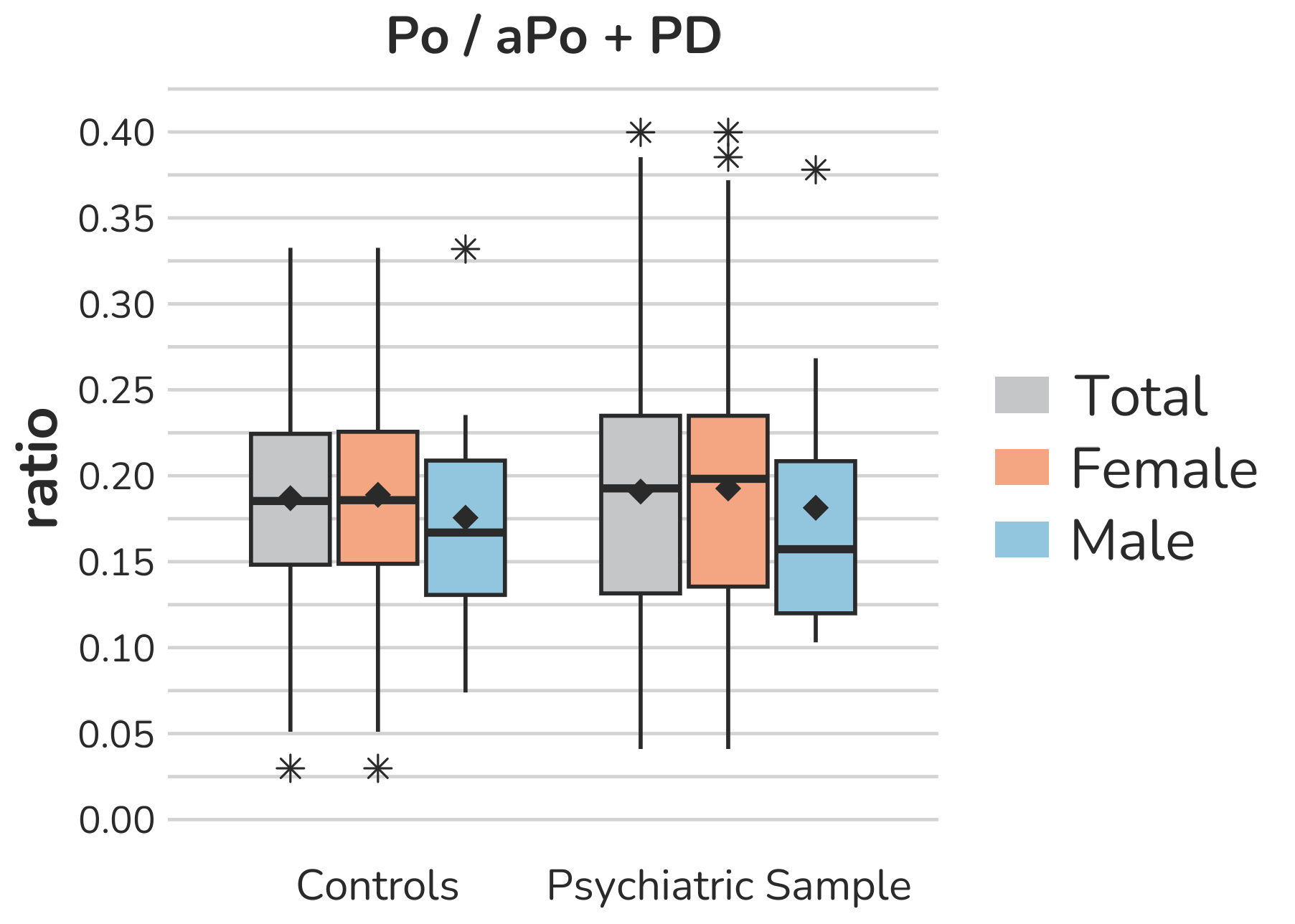

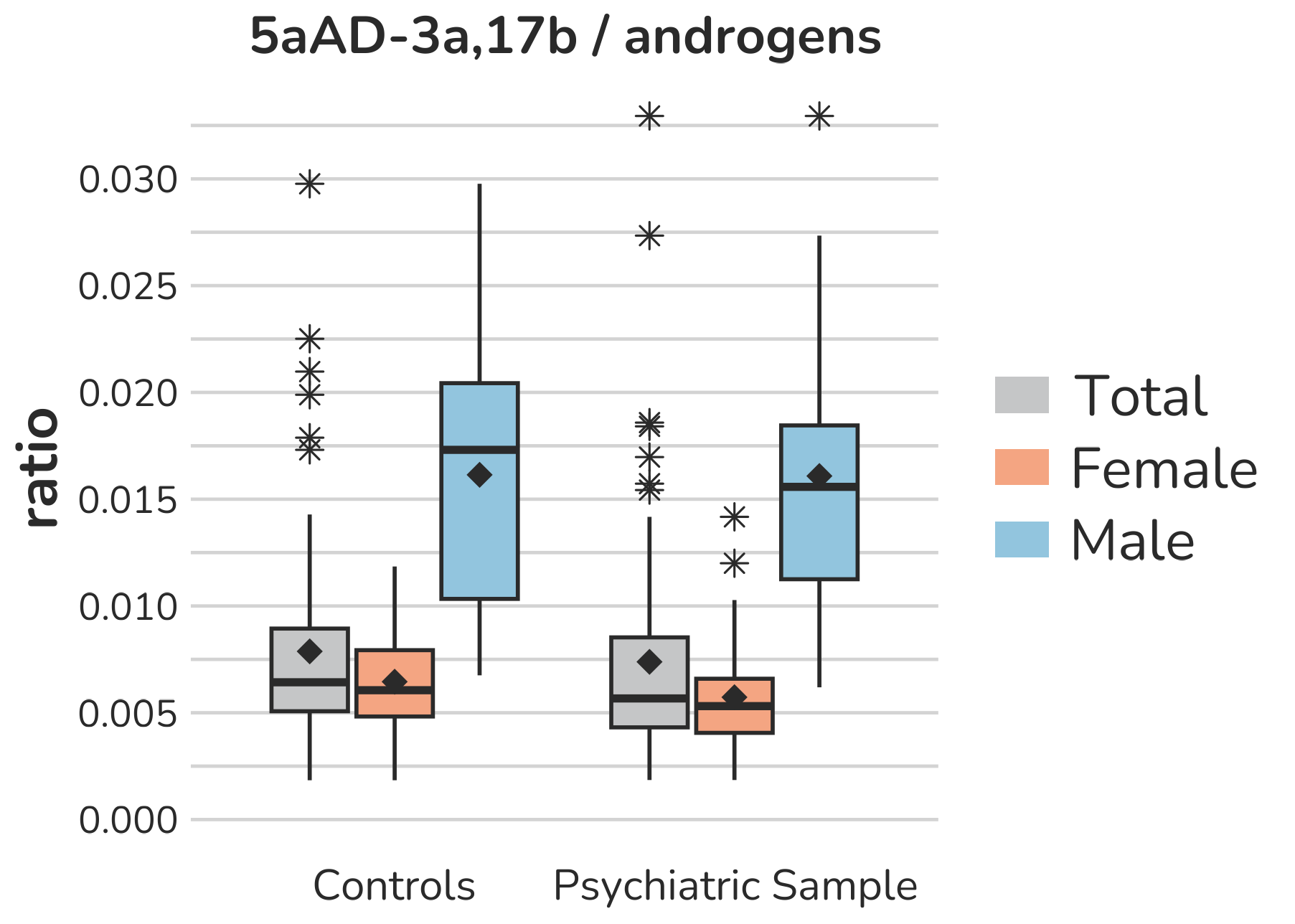

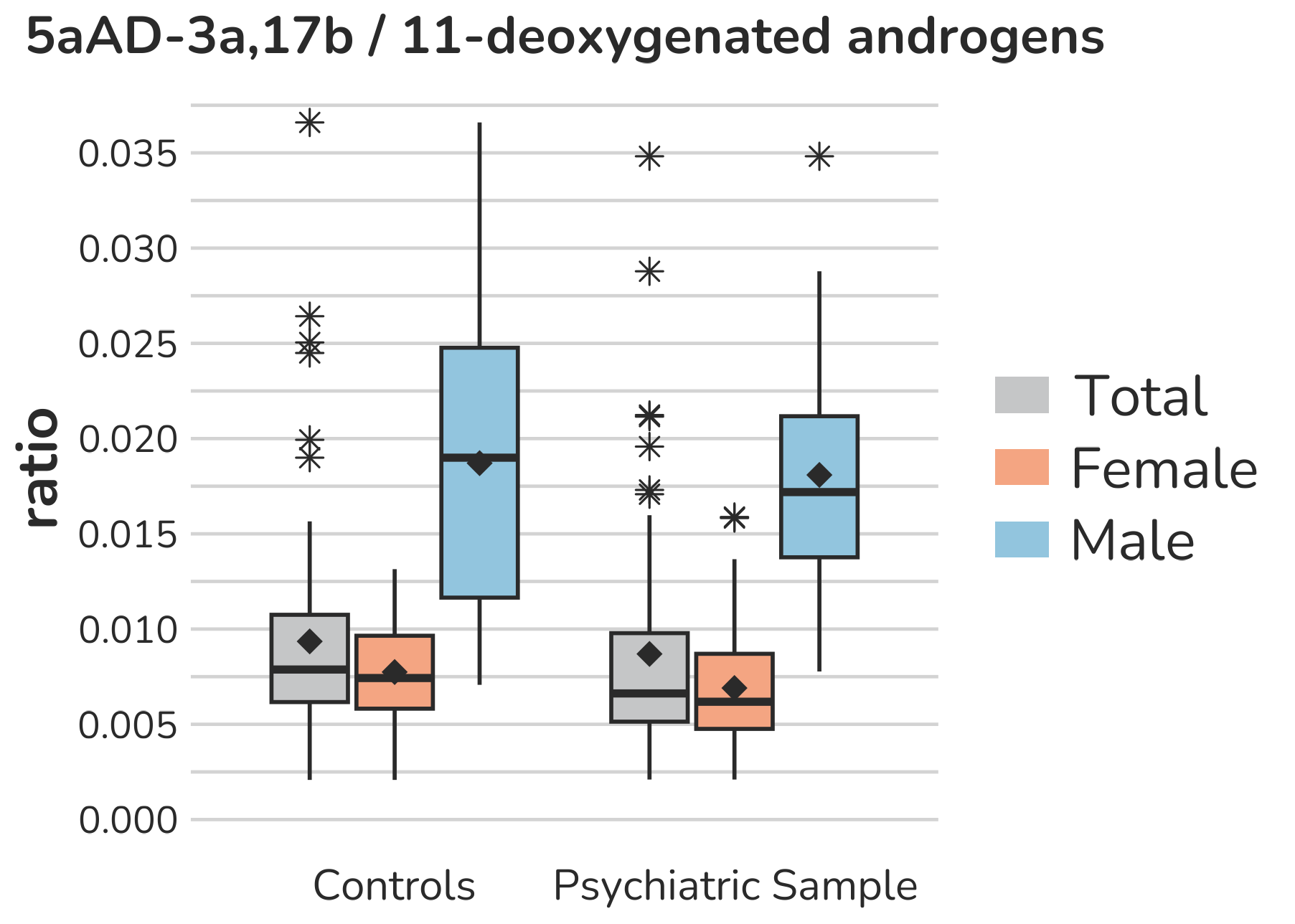
**Supplemental Figure S7 – Boxplots of ratios representing specific ratios as indicators for neuroactive steroid production.** Each box gives the 25th and 75th percentile (lower and upper boundary of the box), the median (horizontal line within each box), and the mean (rhombus within each box). The whiskers are defined by 1.5 times the interquartile range.

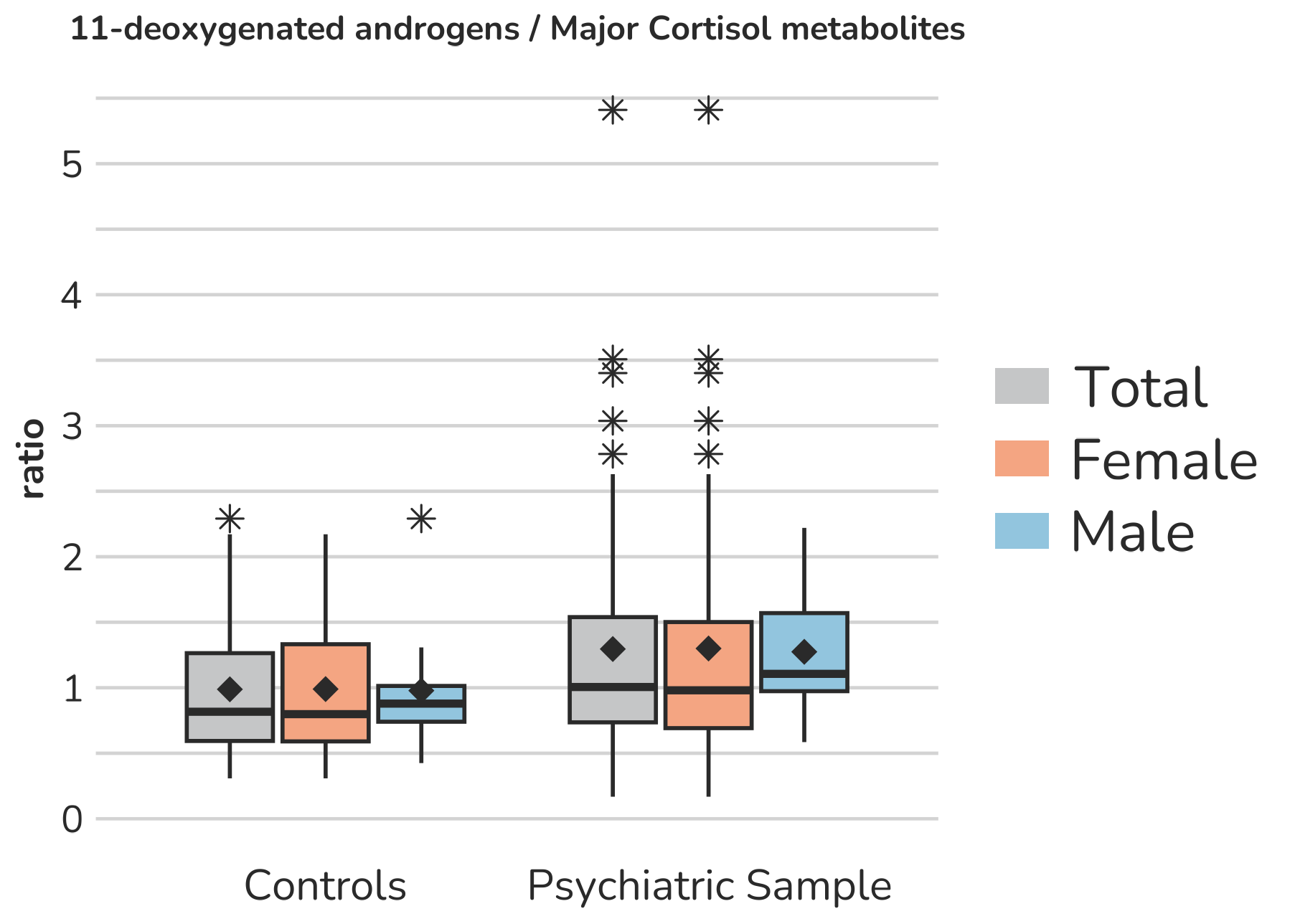

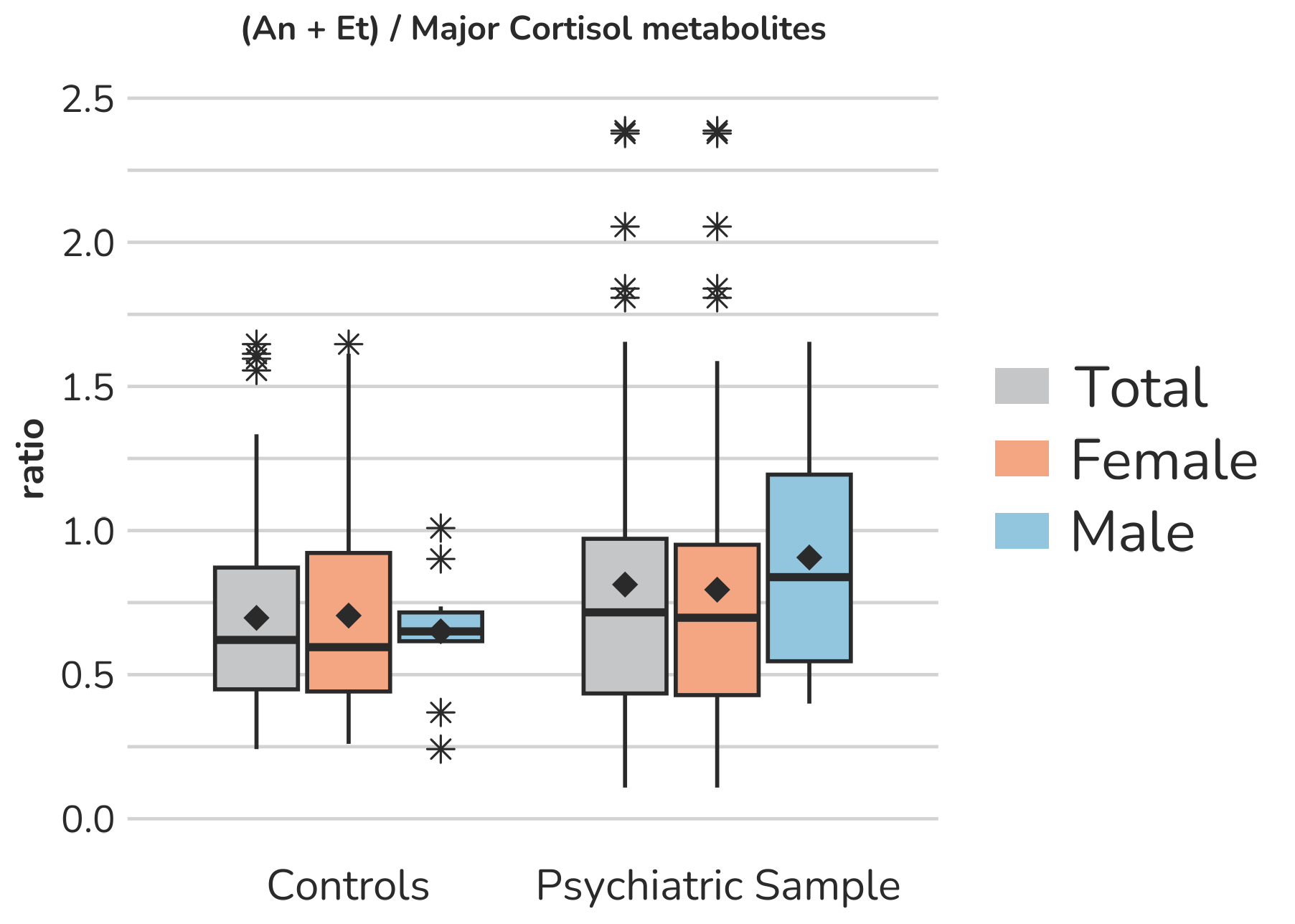

**Supplemental Figure S8 –** **Ratios representing the relative overall androgen production.** Each box gives the 25th and 75th percentile (lower and upper boundary of the box), the median (horizontal line within each box), and the mean (rhombus within each box). The whiskers are defined by 1.5 times the interquartile range.

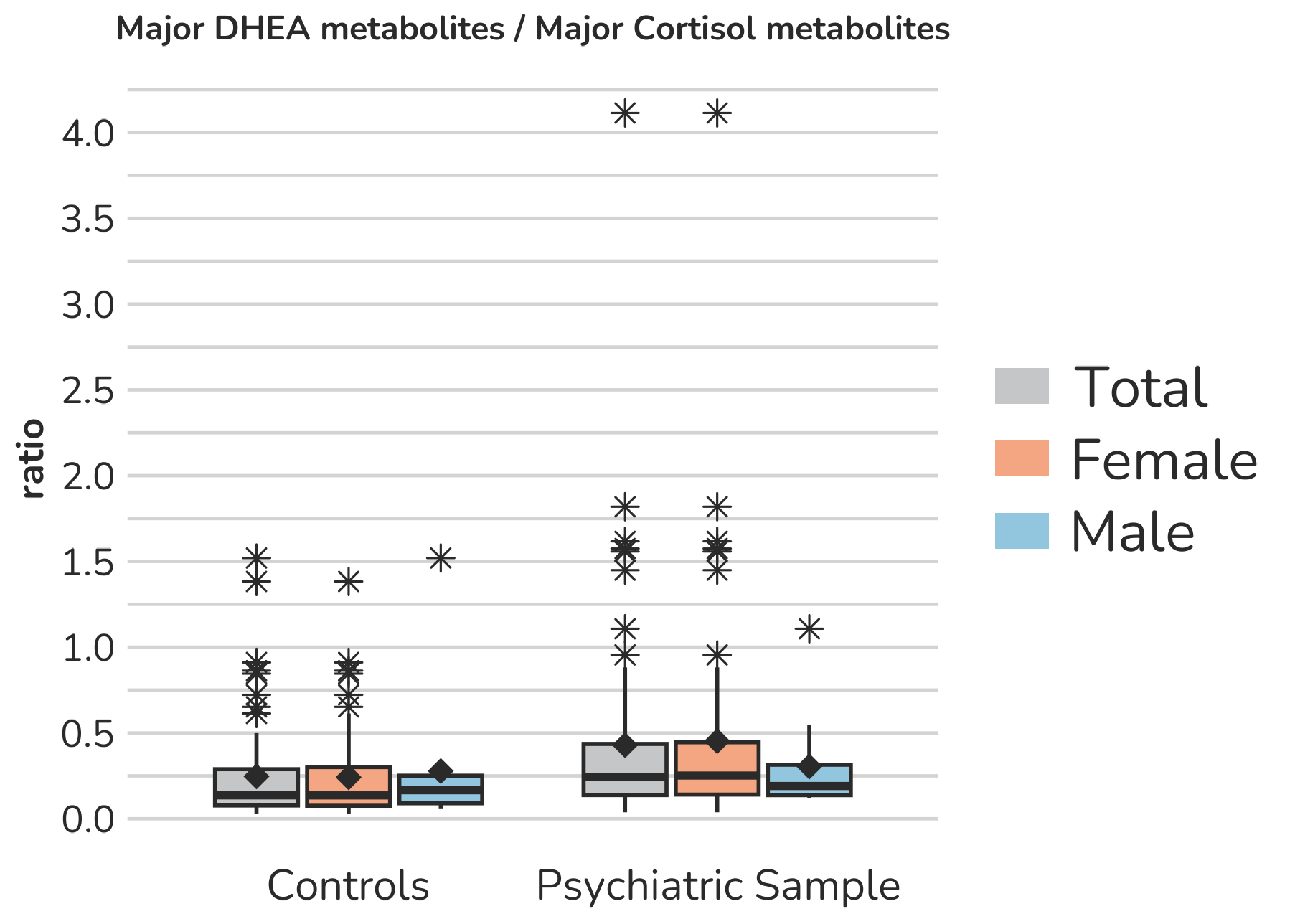

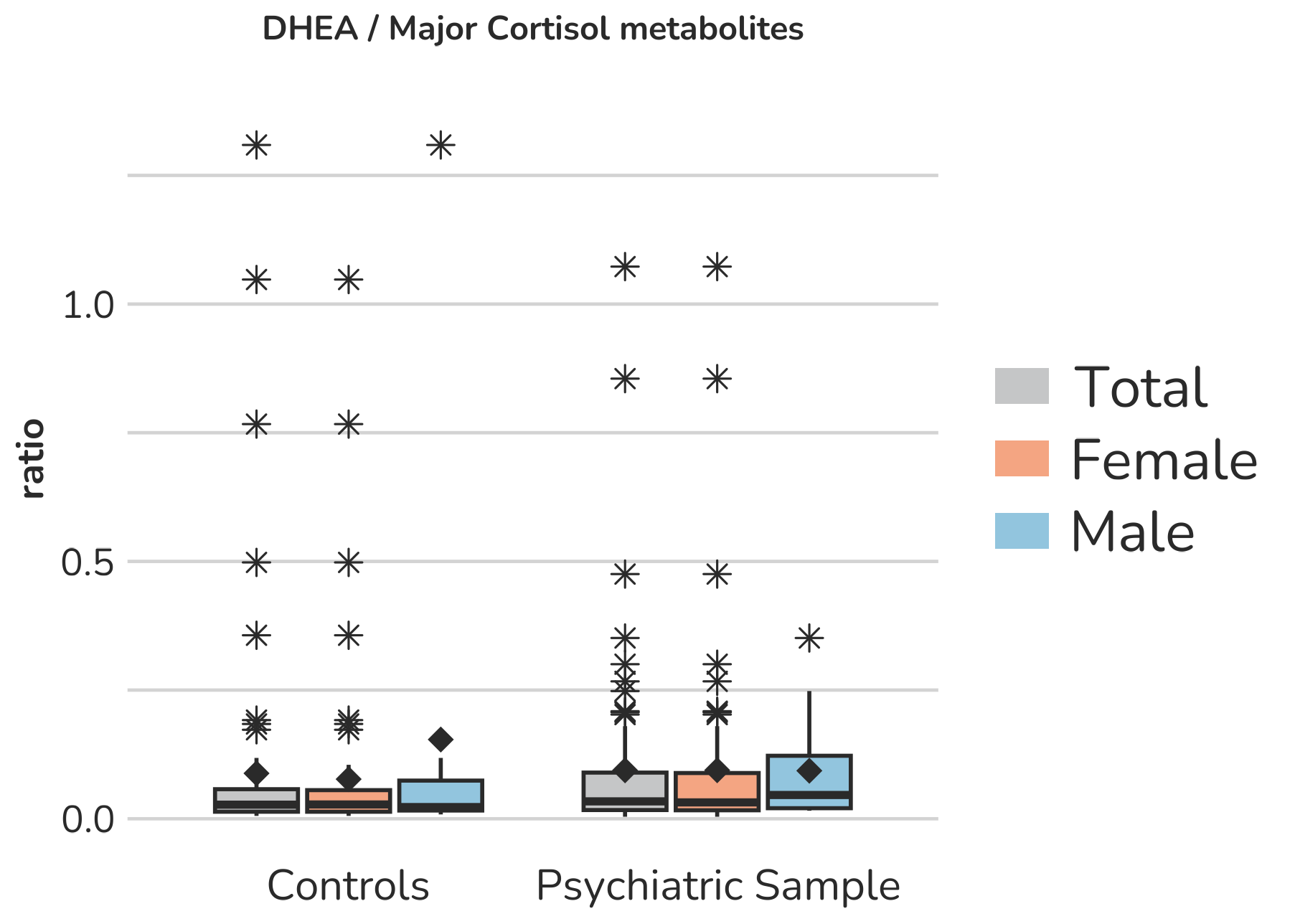

**Supplemental Figure S9 –** **Ratios representing the relative adrenal androgen production.** Each box gives the 25th and 75th percentile (lower and upper boundary of the box), the median (horizontal line within each box), and the mean (rhombus within each box). The whiskers are defined by 1.5 times the interquartile range. For DHEA/Major cortisol metabolites, one female subject of the psychiatric sample with a ratio of 3.2 is excluded from the figure to enhance the visualization of the remaining values. Aside from this, all outliers are displayed (illustrated by *).

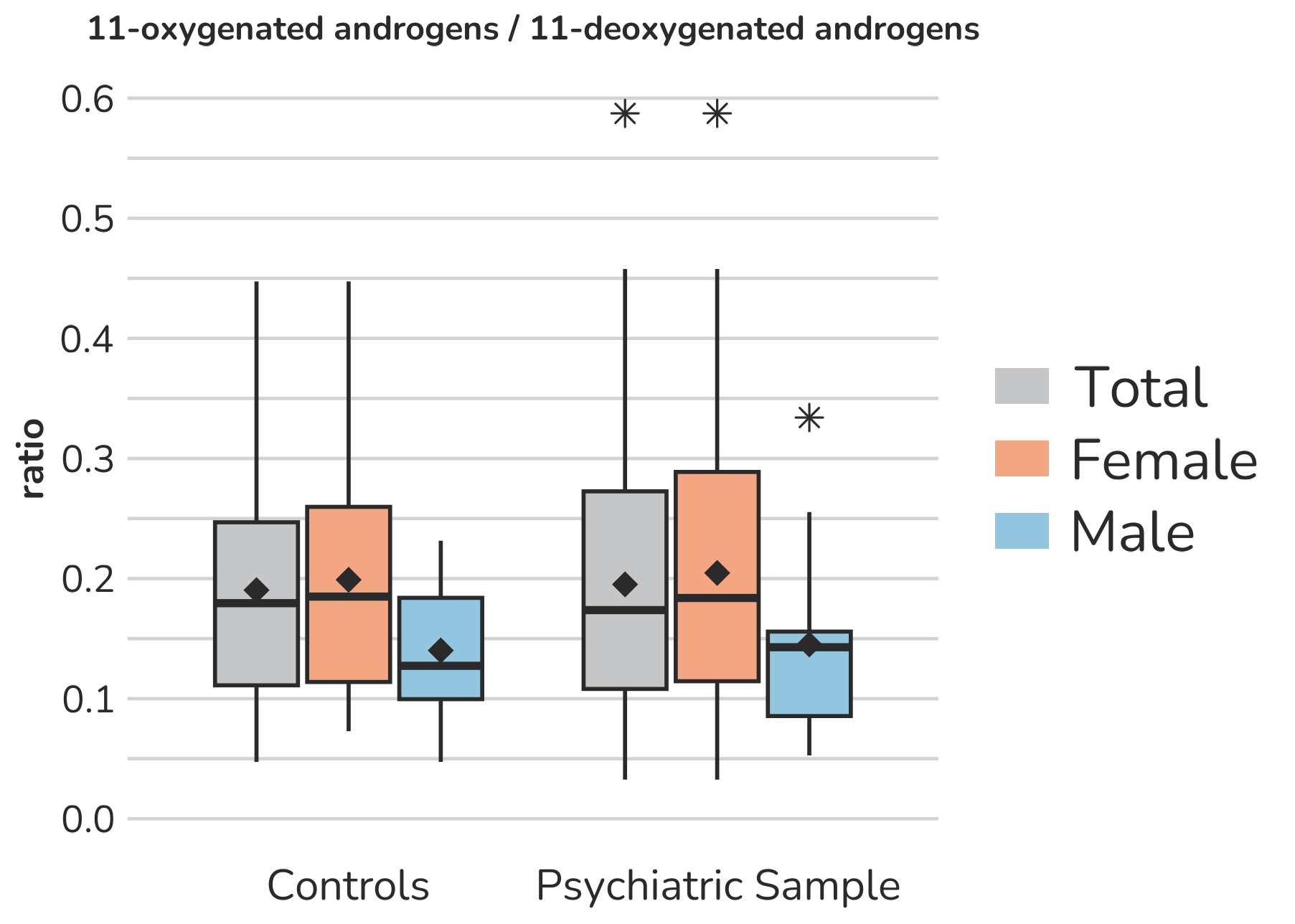

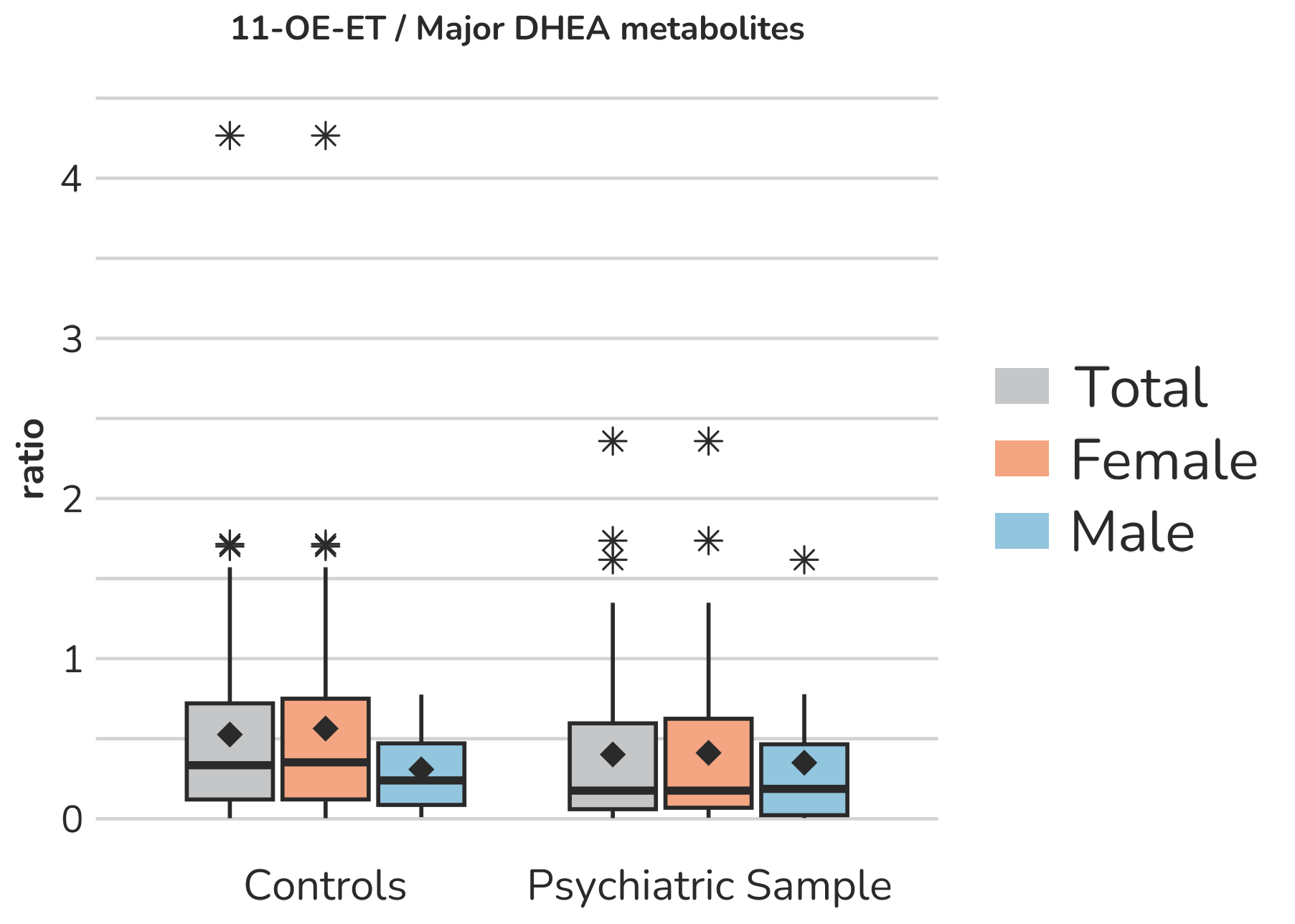

**Supplemental Figure S10 – Boxplots of ratios representing production of** **11-oxygenated androgens in relation to 11-deoxygenated androgens.** Each box gives the 25th and 75th percentile (lower and upper boundary of the box), the median (horizontal line within each box), and the mean (rhombus within each box). The whiskers are defined by 1.5 times the interquartile range. All outliers are presented (illustrated by *).

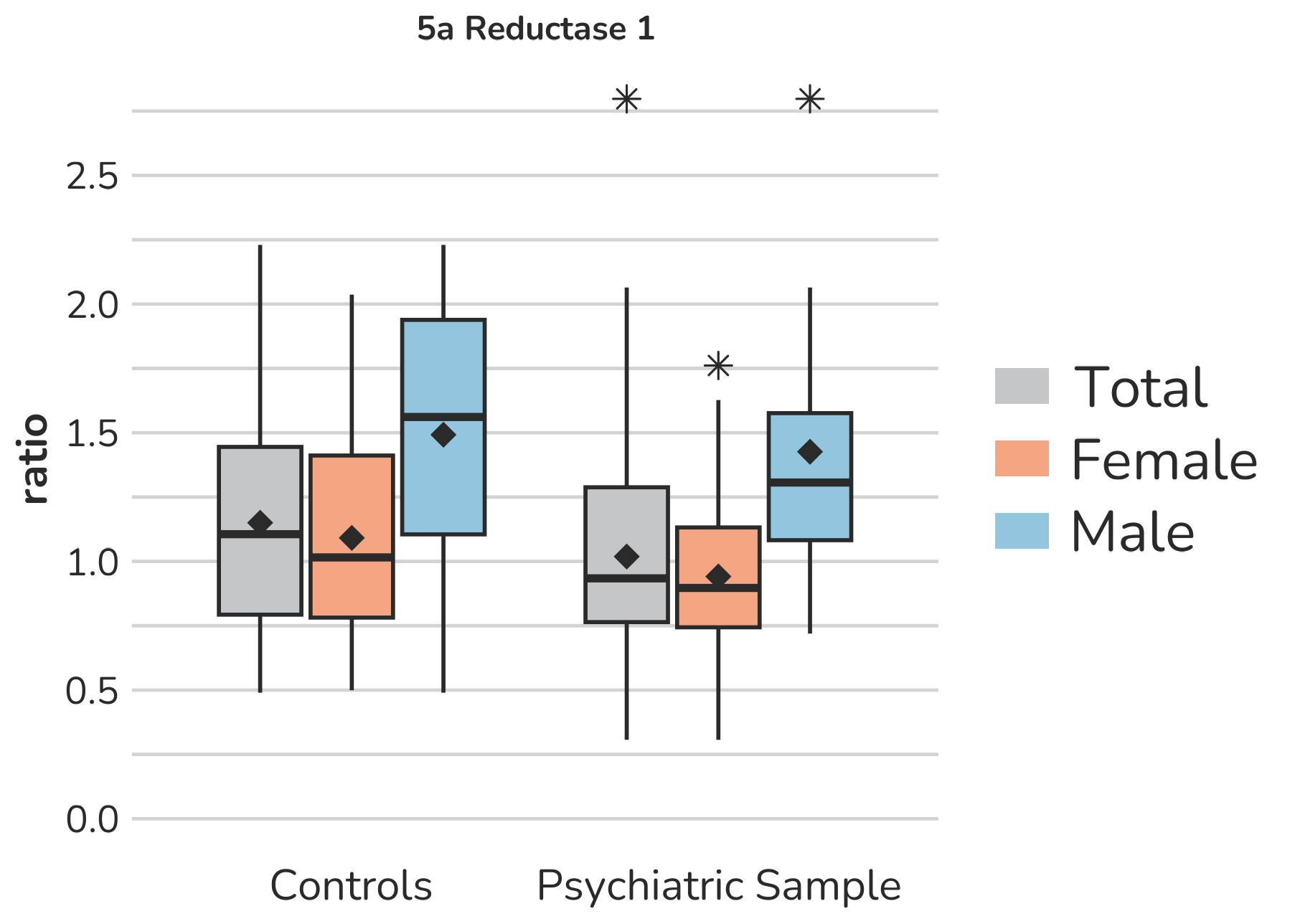

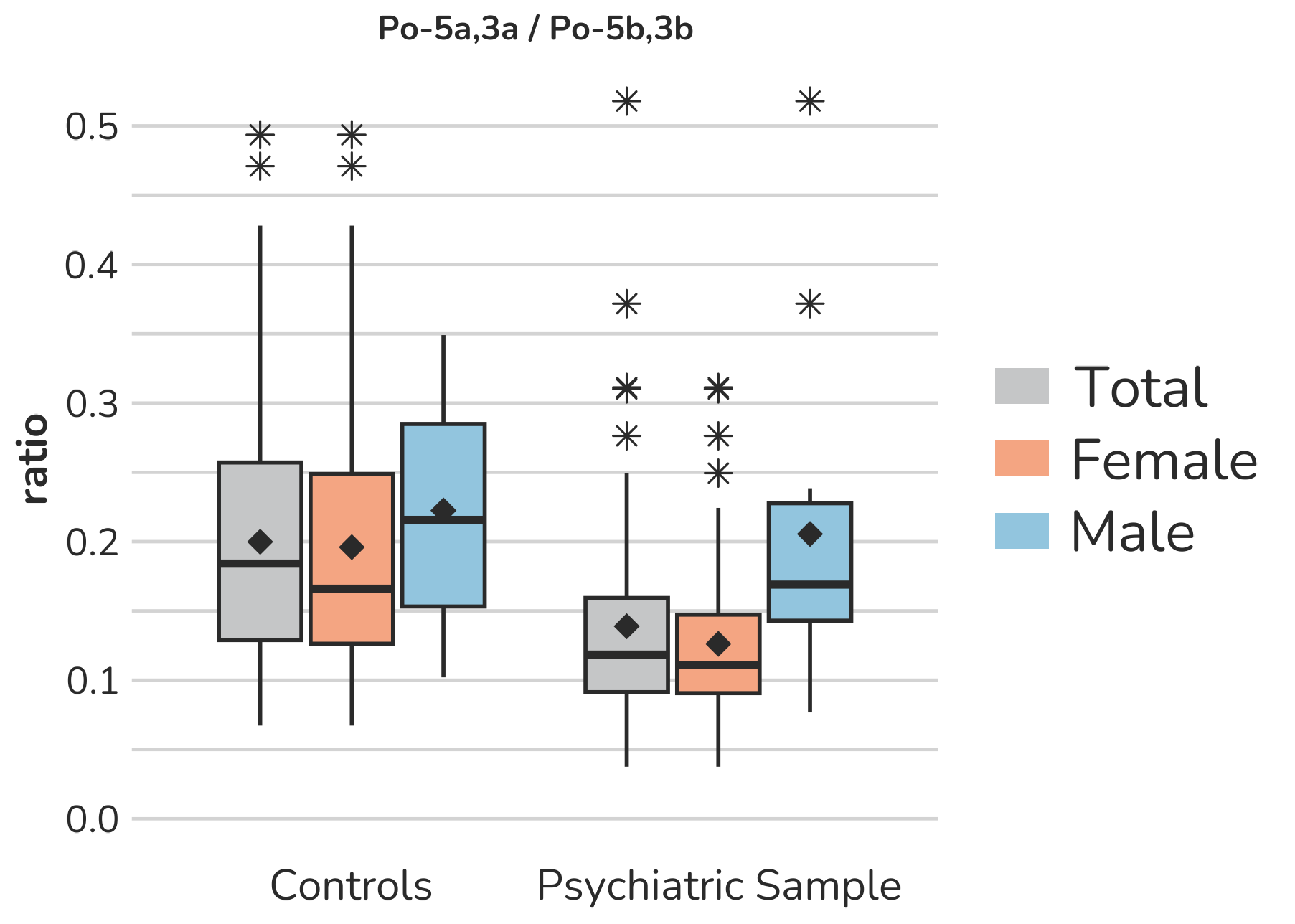

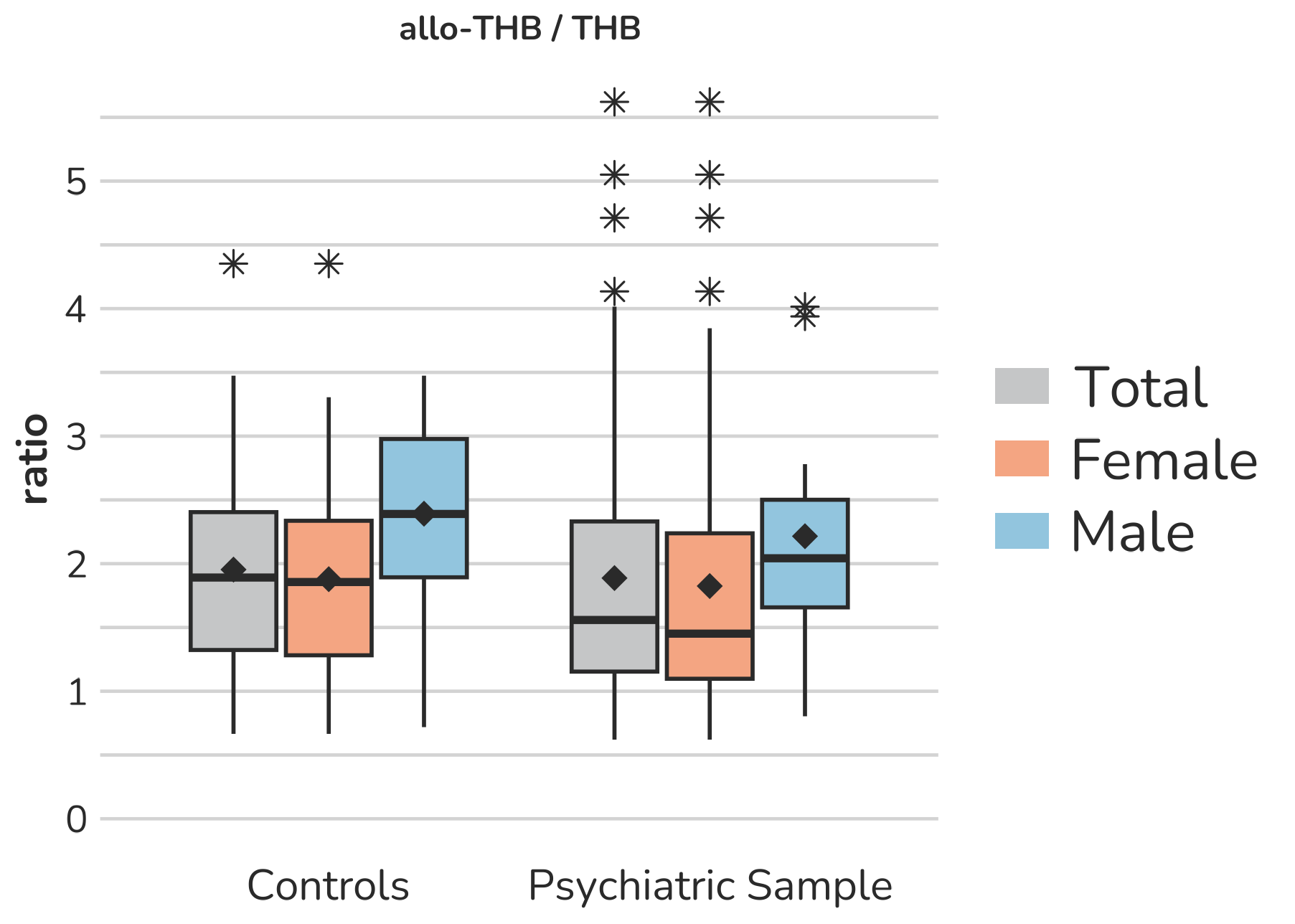

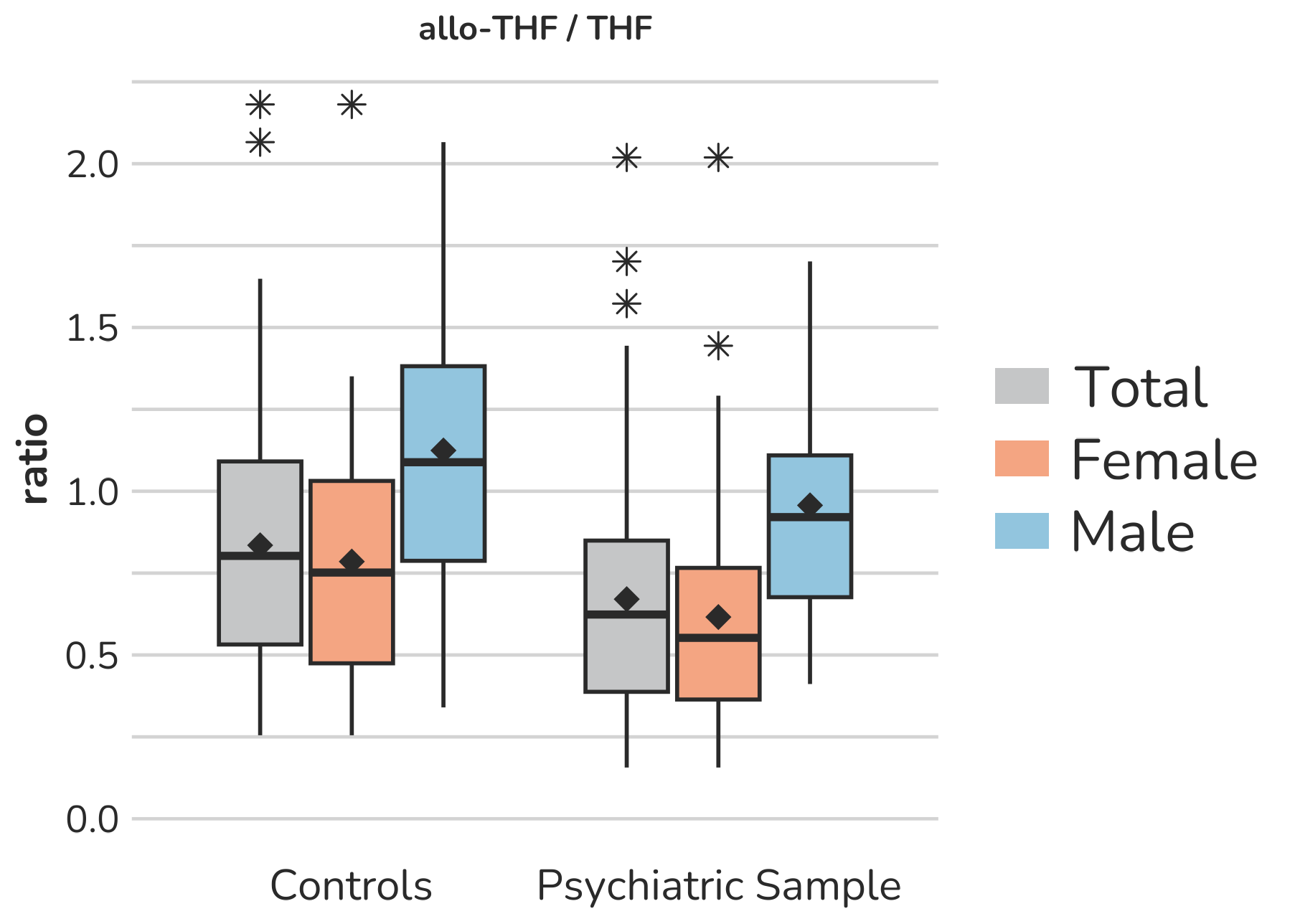

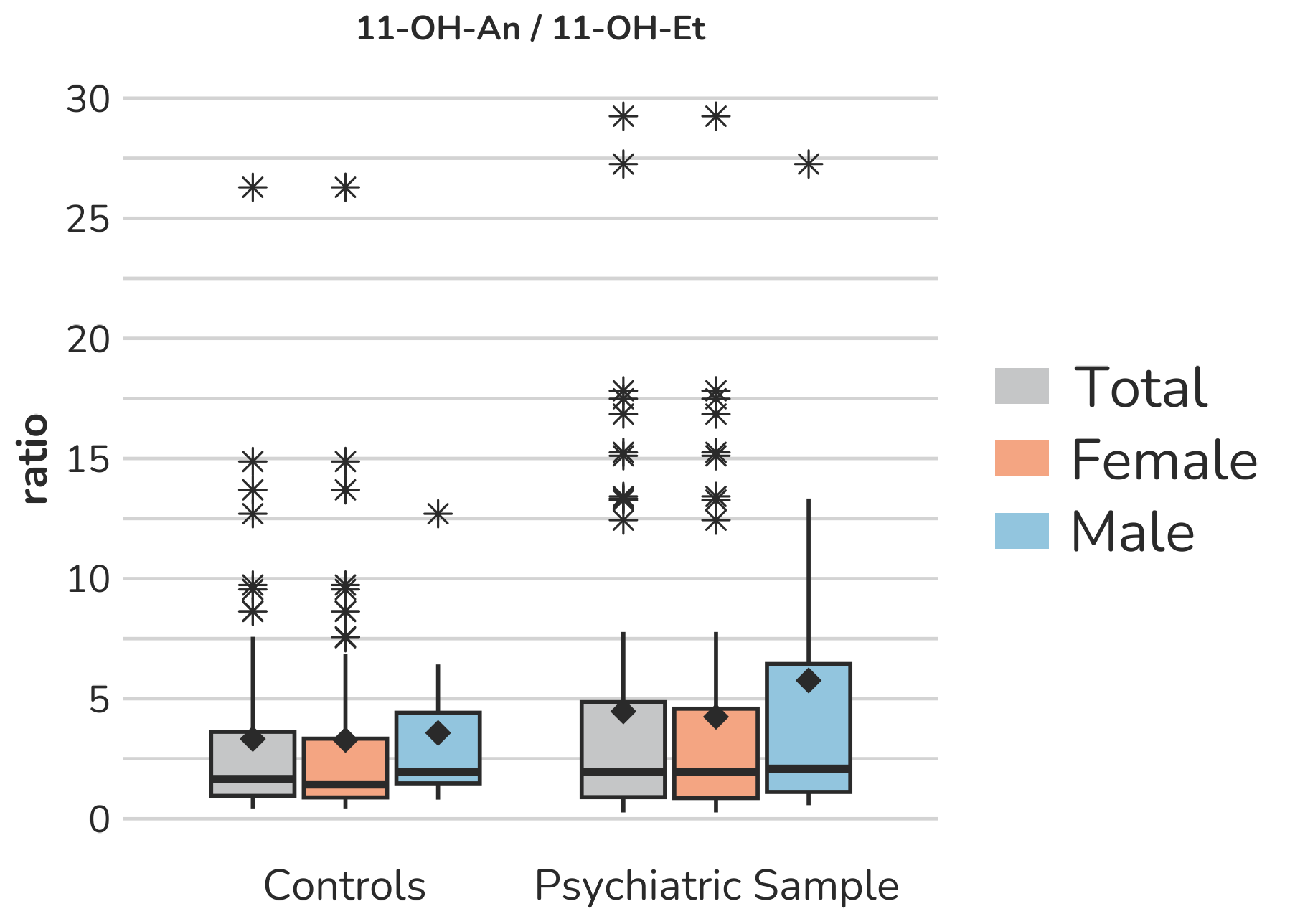

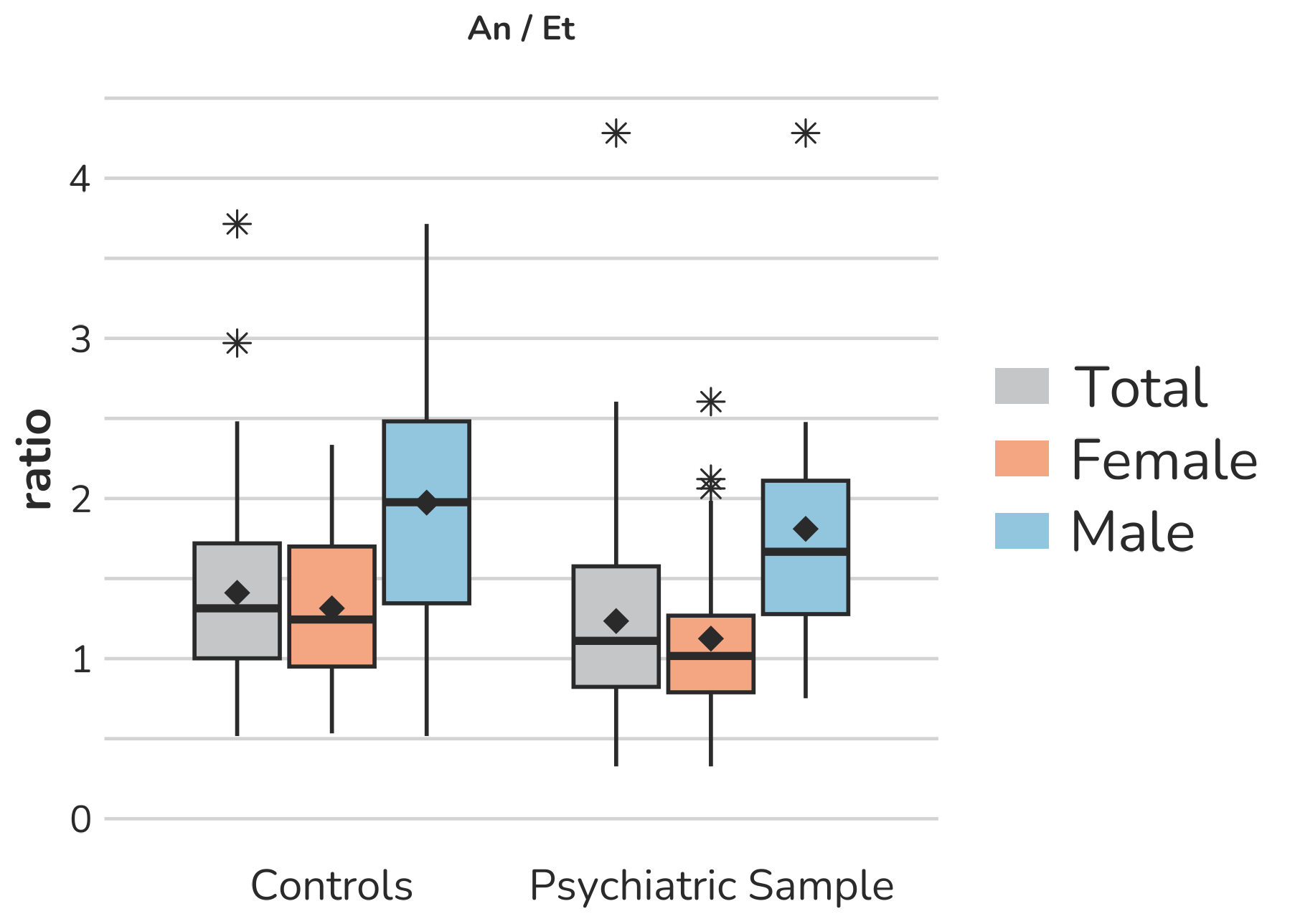

**Supplemental Figure S11 – Boxplots of ratios representing specific ratios as indicators for** **5α-reductase activity.** Each box gives the 25th and 75th percentile (lower and upper boundary of the box), the median (horizontal line within each box), and the mean (rhombus within each box). The whiskers are defined by 1.5 times the interquartile range. For 11-OH-An/11-OH-Et, one male subject of the psychiatric sample with a ratio of 43.0 is excluded from the figure to enhance the visualization of the remaining values. Aside from this, all outliers are displayed (illustrated by *).

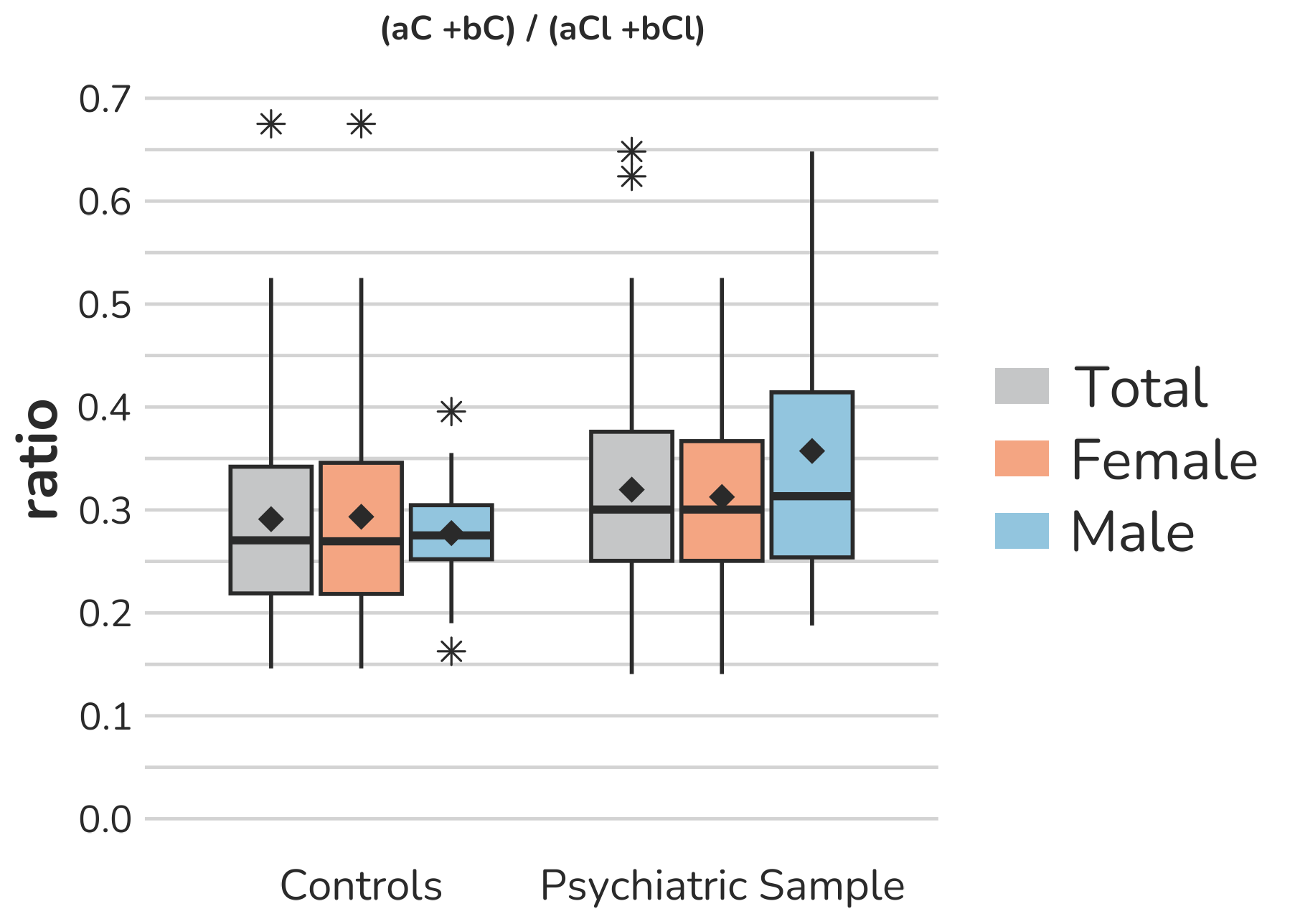

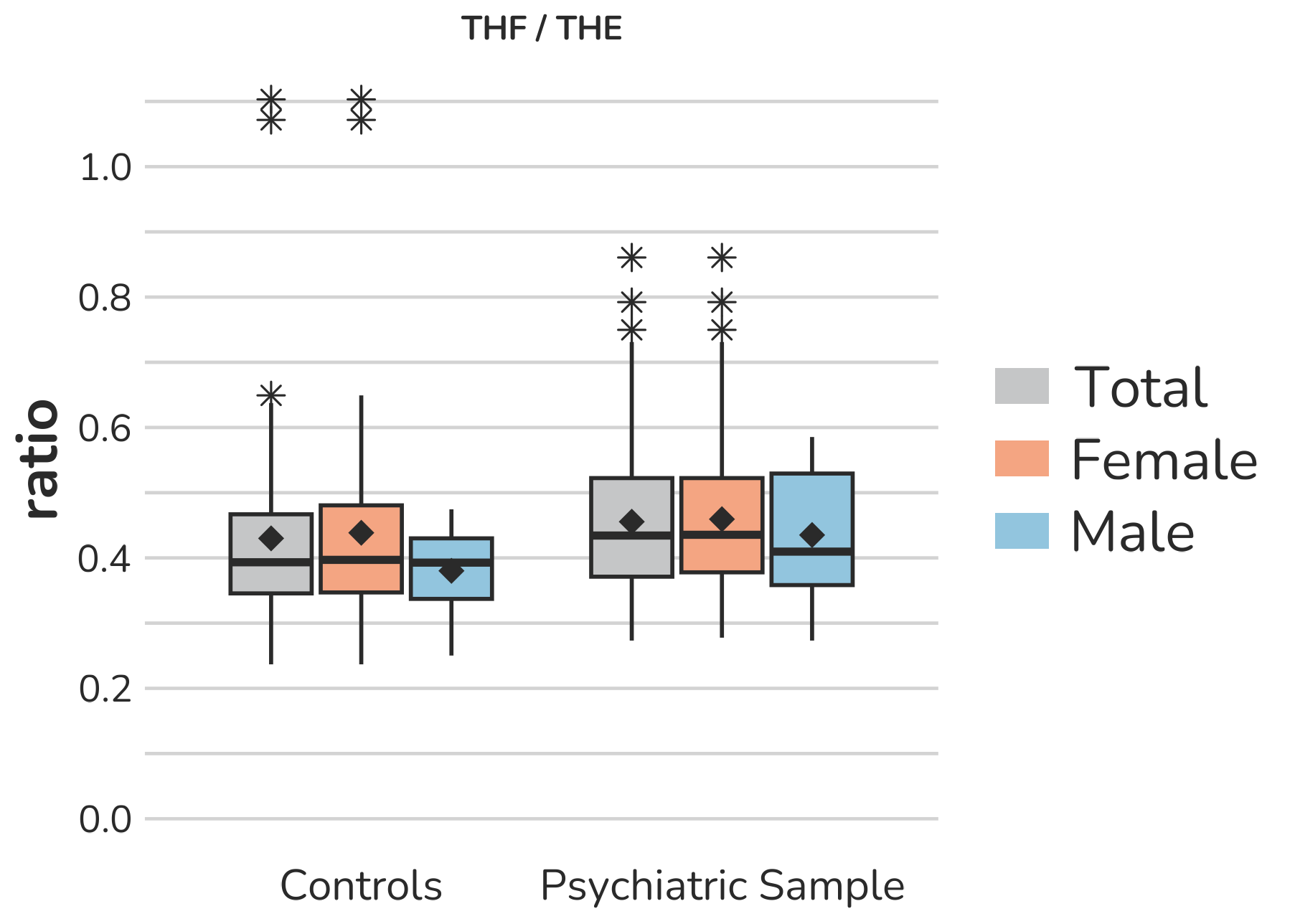

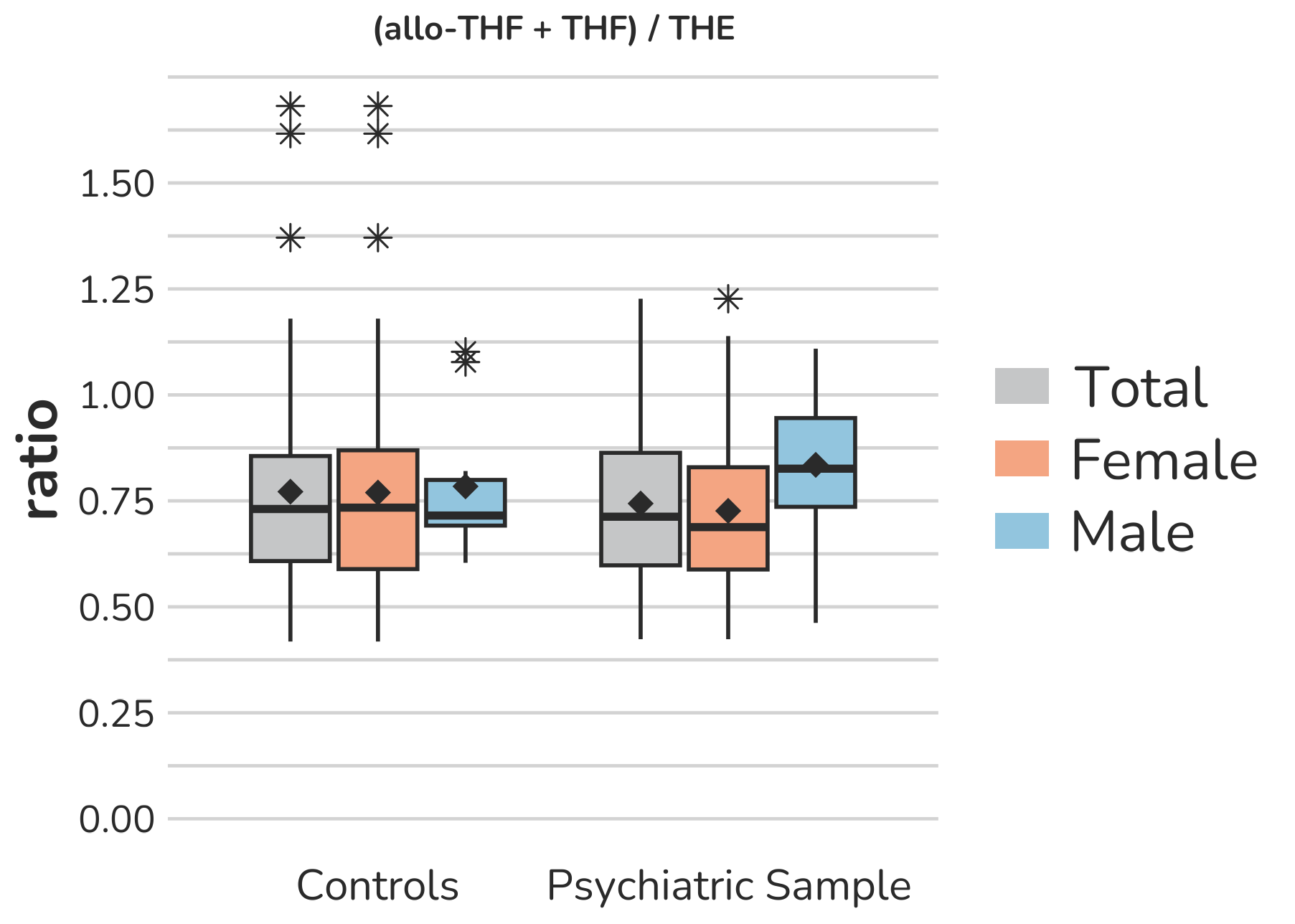

**Supplemental Figure S12 – Boxplots of ratios representing specific ratios as indicators for** **11β-hydroxy-steroiddehydrogenase (11β-HSD) activity.** Each box gives the 25th and 75th percentile (lower and upper boundary of the box), the median (horizontal line within each box), and the mean (rhombus within each box). The whiskers are defined by 1.5 times the interquartile range. All outliers are presented (illustrated by *).

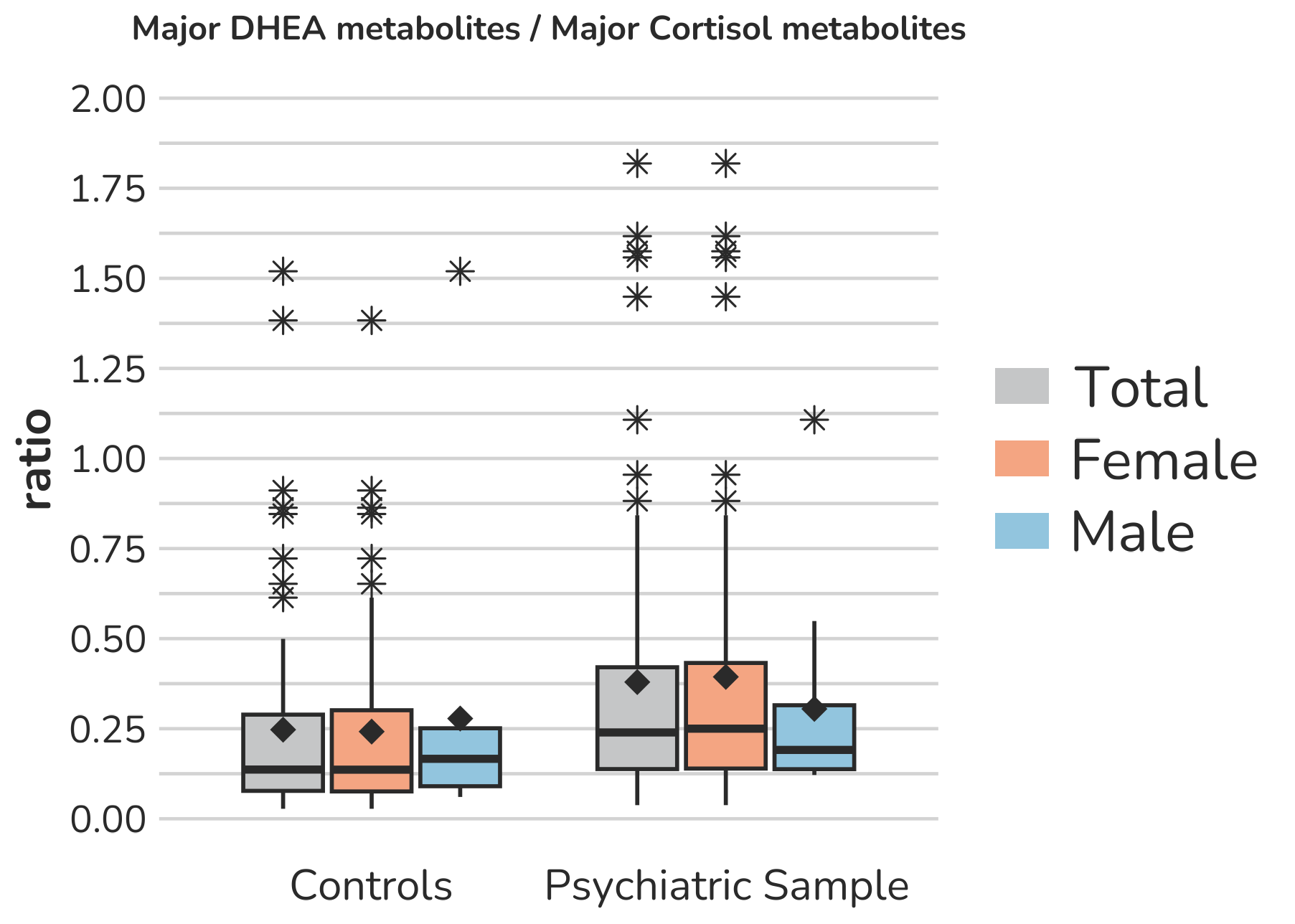

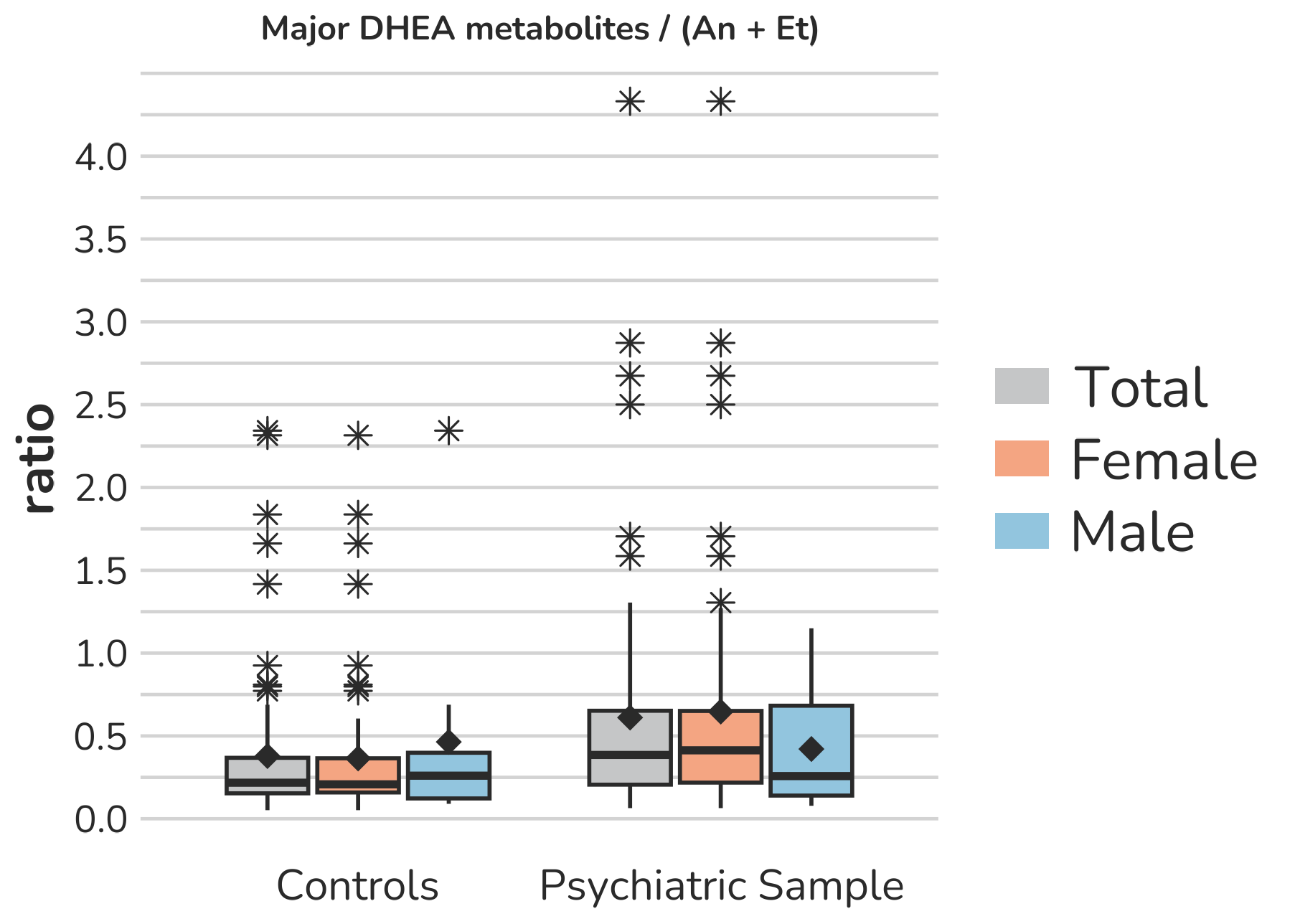

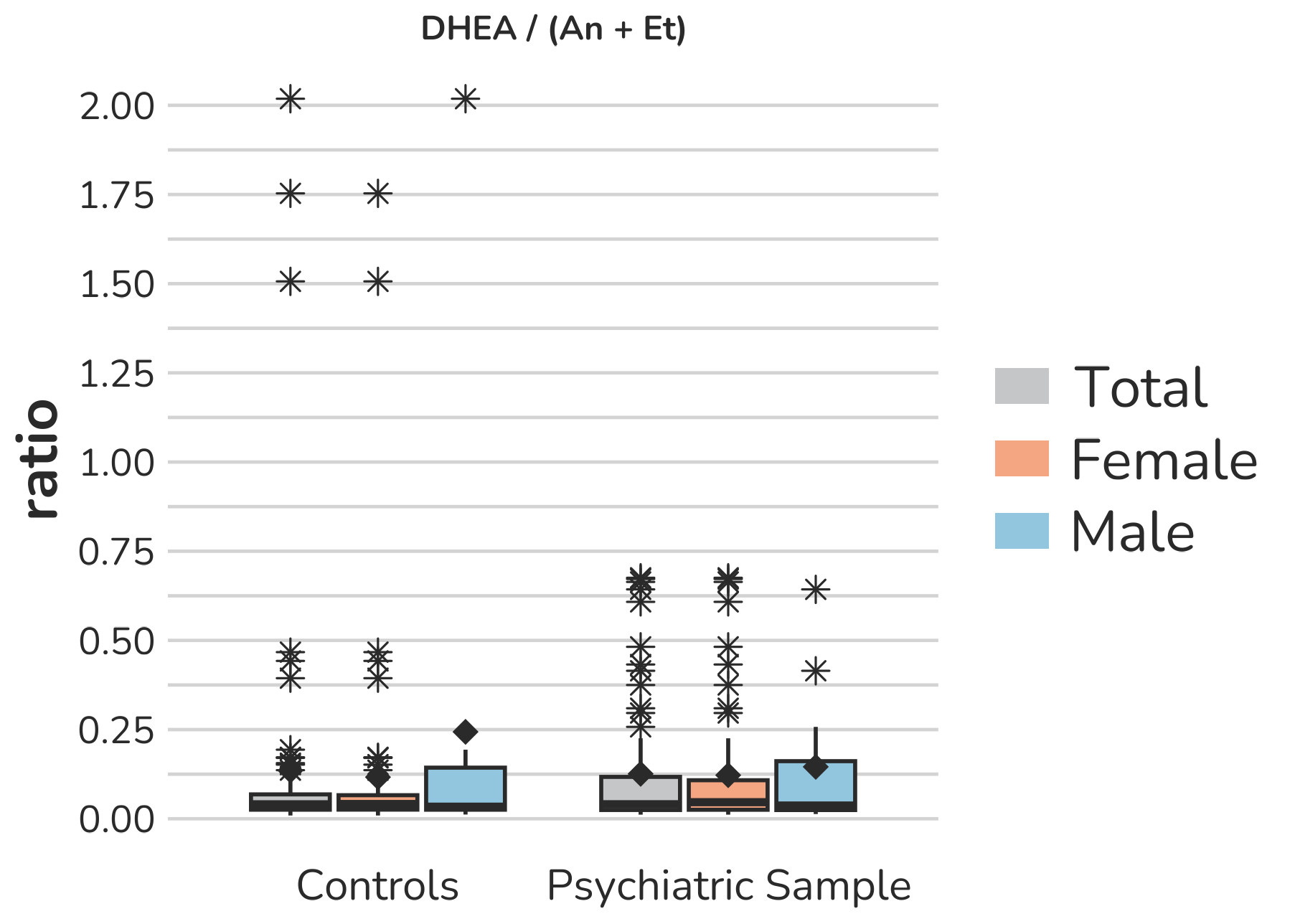

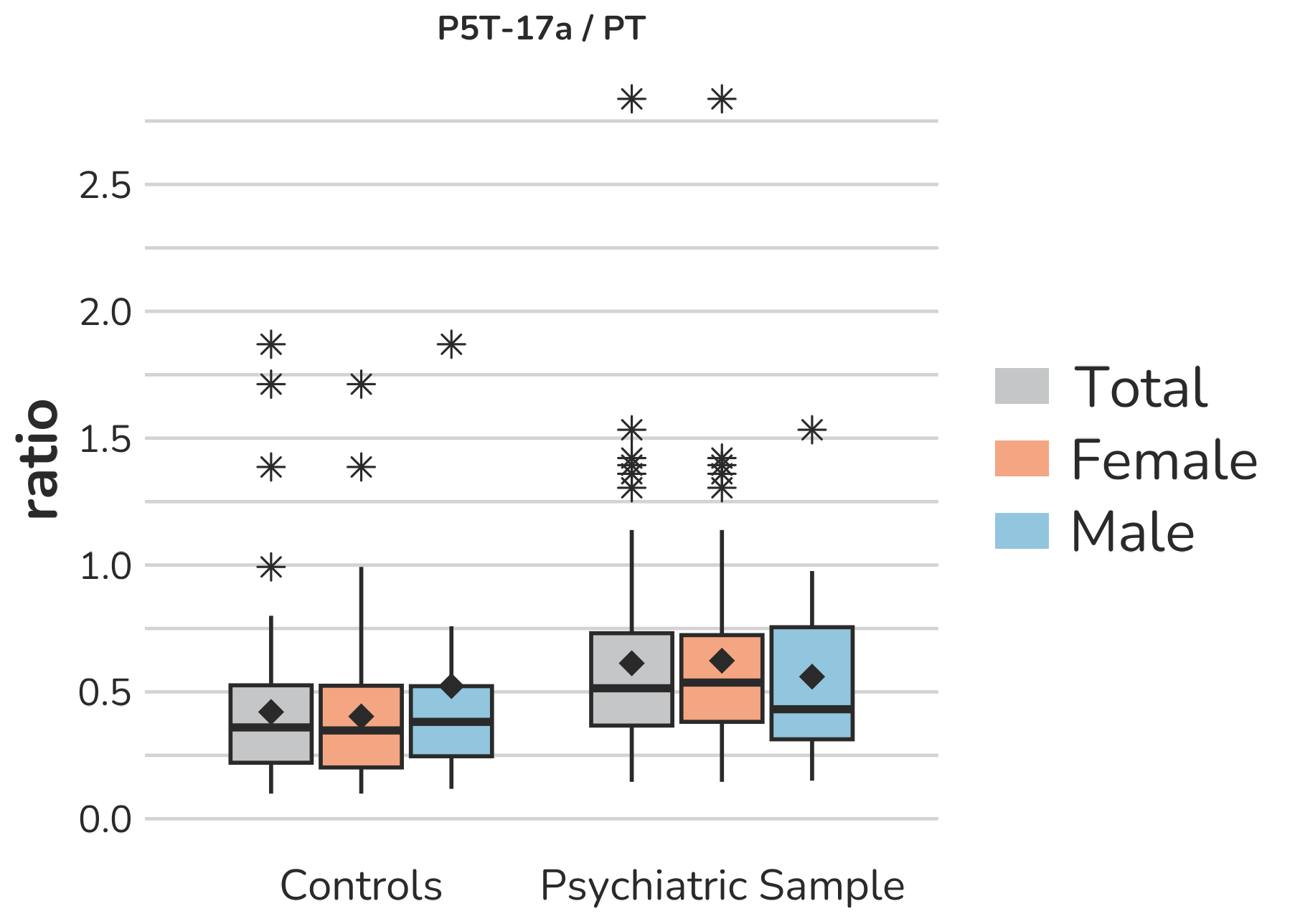

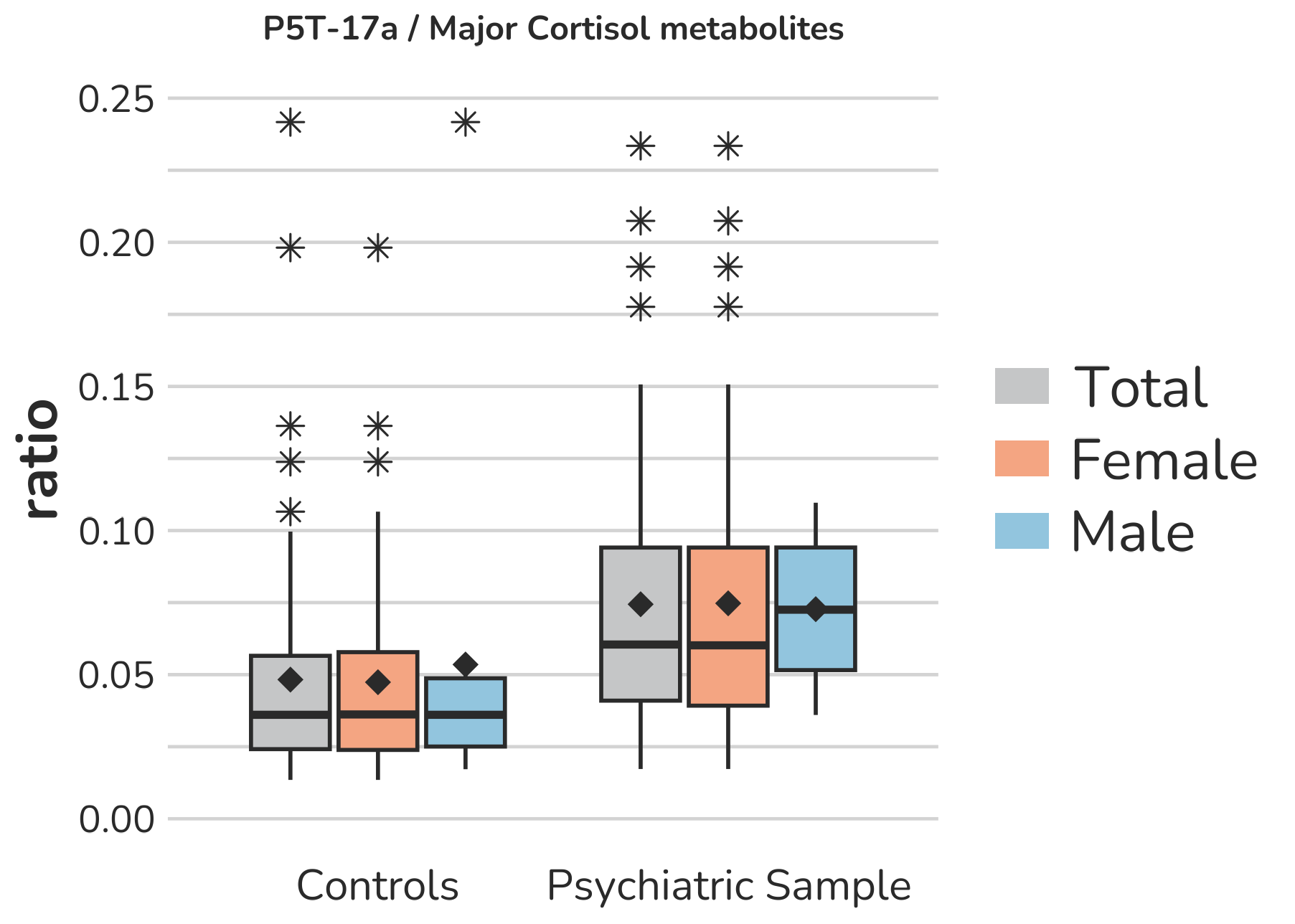

**Supplemental Figure S13 – Boxplots of ratios representing specific ratios as indicators for** **3β-hydroxysteroiddehydrogenase (3β-HSD) activity.** Each box gives the 25th and 75th percentile (lower and upper boundary of the box), the median (horizontal line within each box), and the mean (rhombus within each box). The whiskers are defined by 1.5 times the interquartile range. For the major DHEA metabolites / major Cortisol metabolites and the DHEA / An + Et ratio, one female subject of the psychiatric sample with a ratio of 4.1 and 3.4 is excluded from the figure to enhance the visualization of the remaining values. Aside from this, all outliers are displayed (illustrated by *).

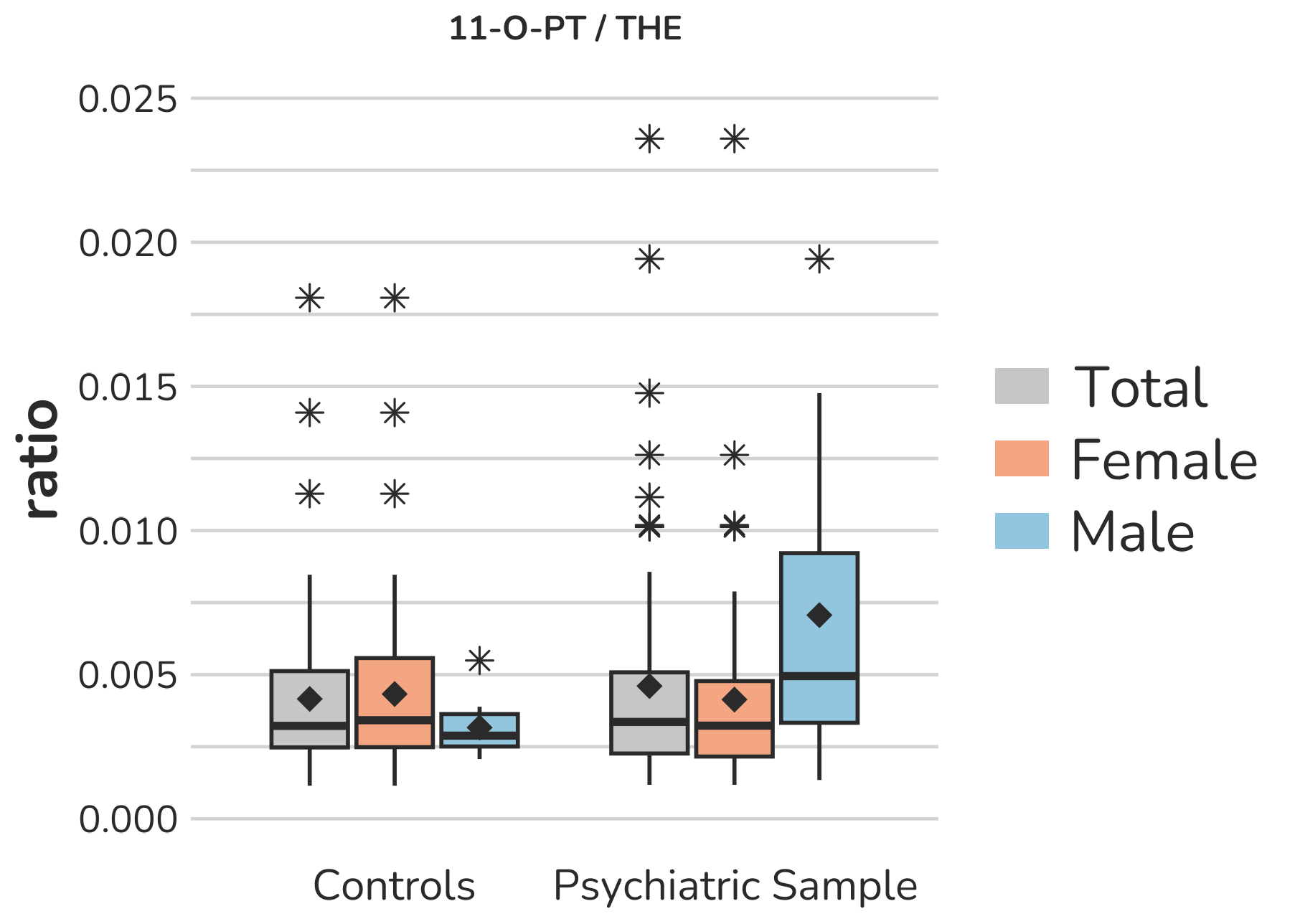

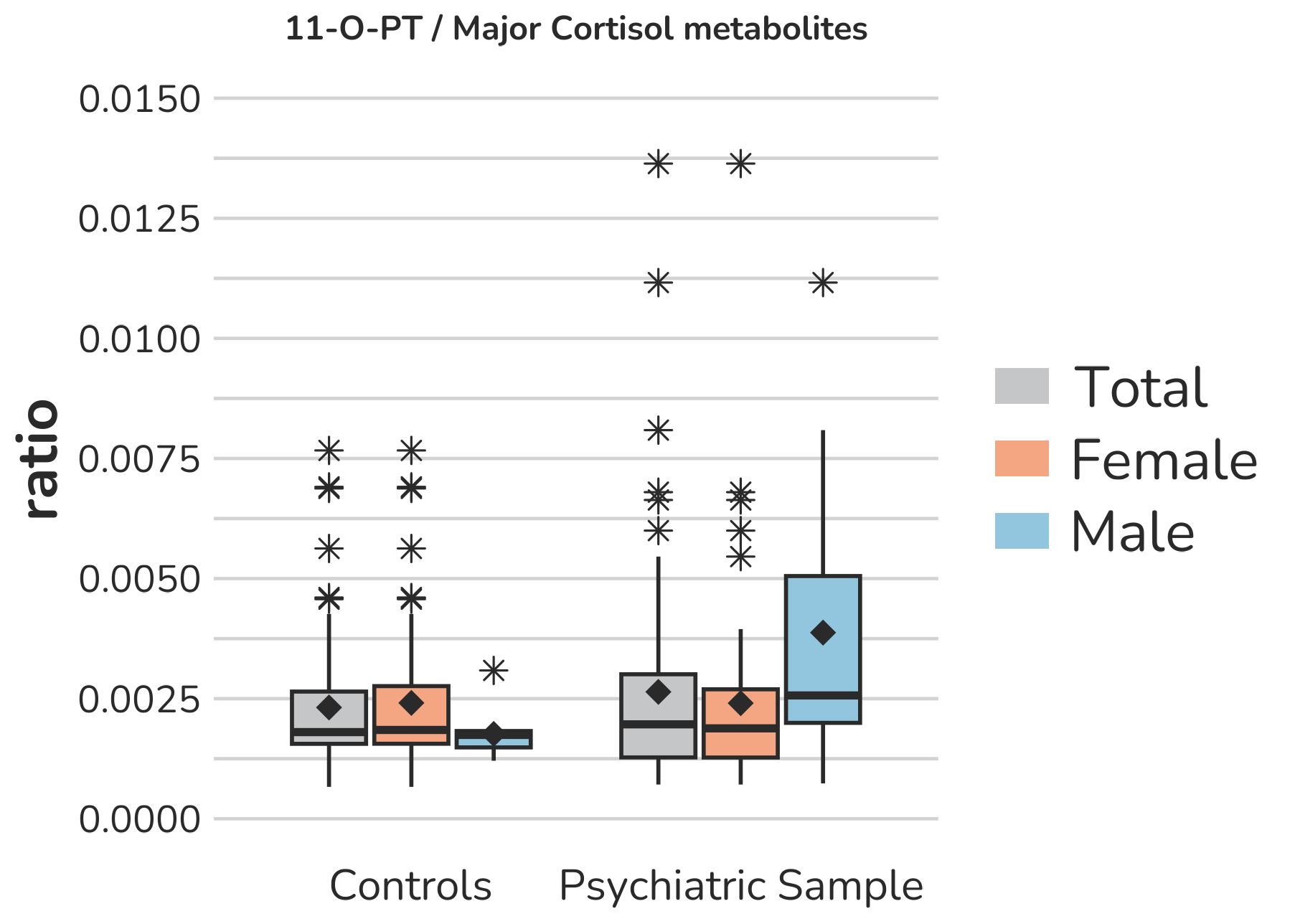

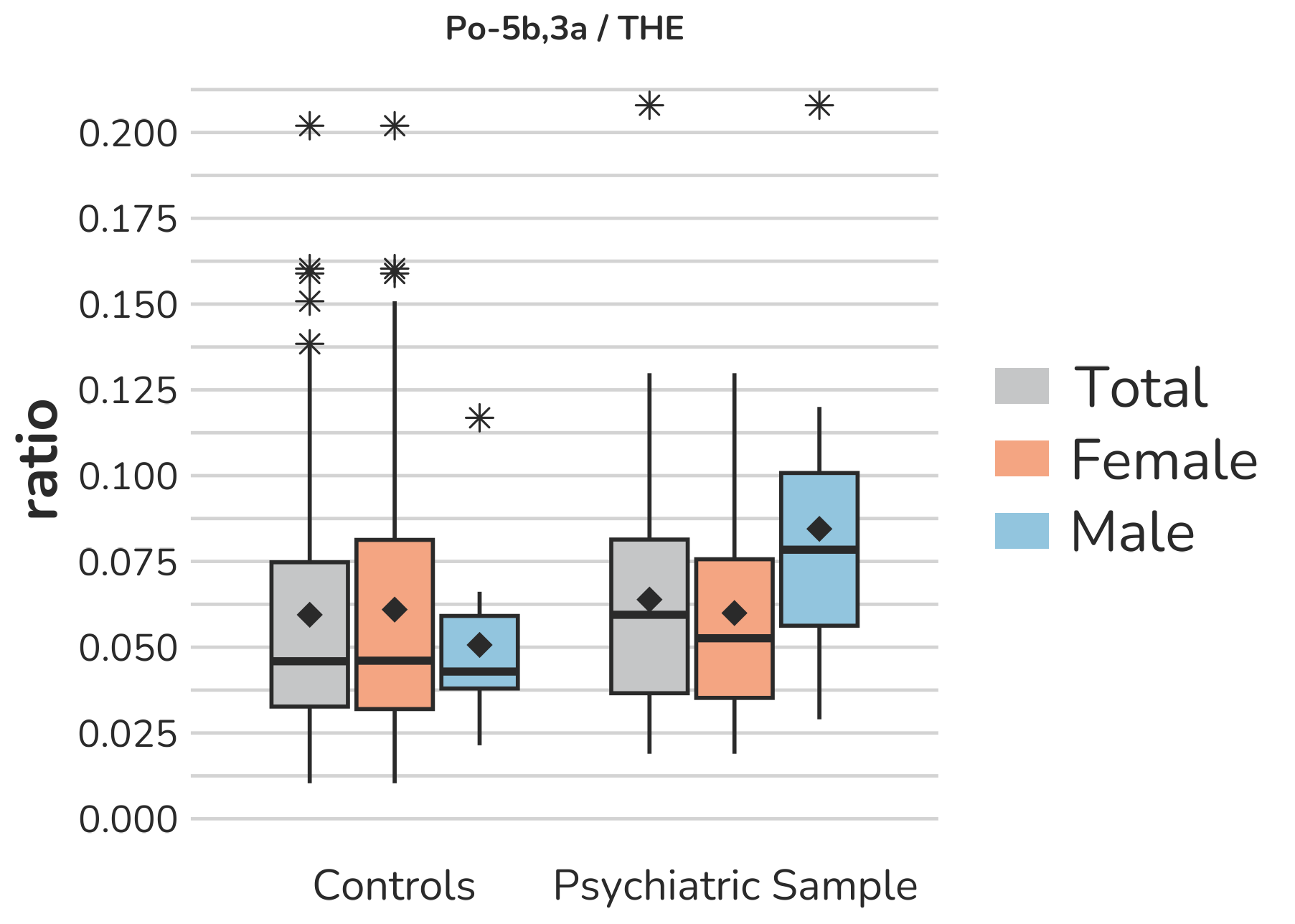

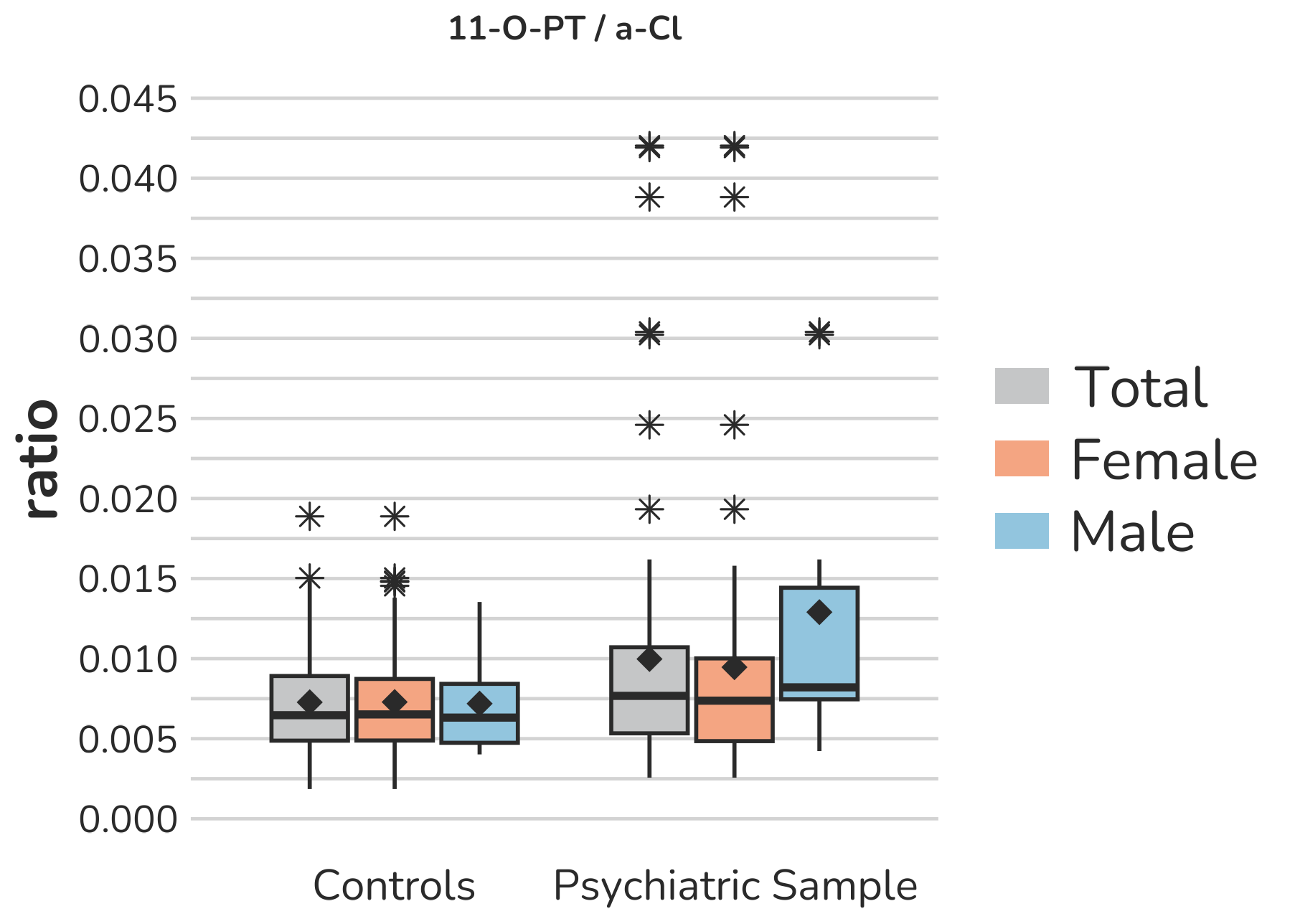

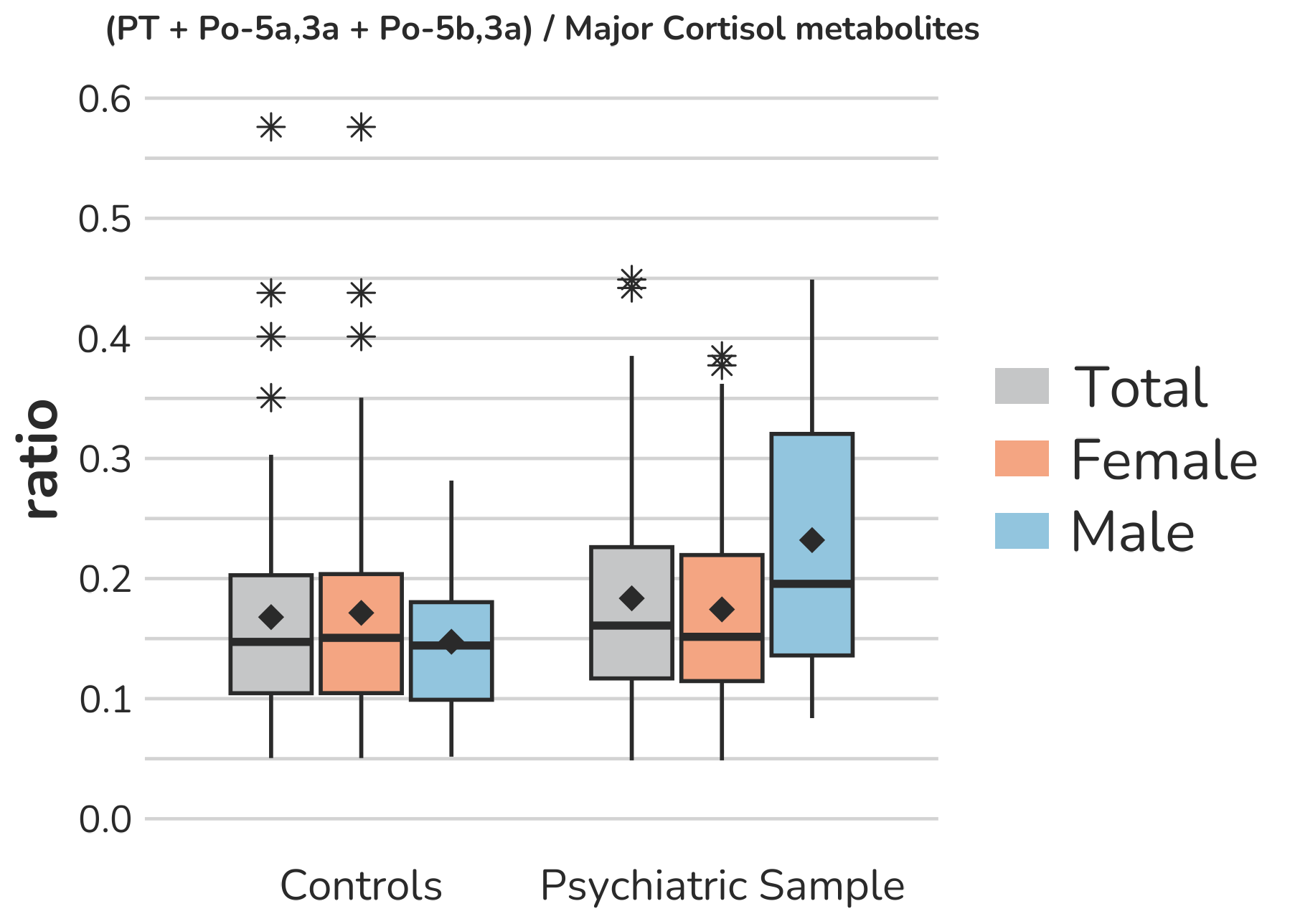

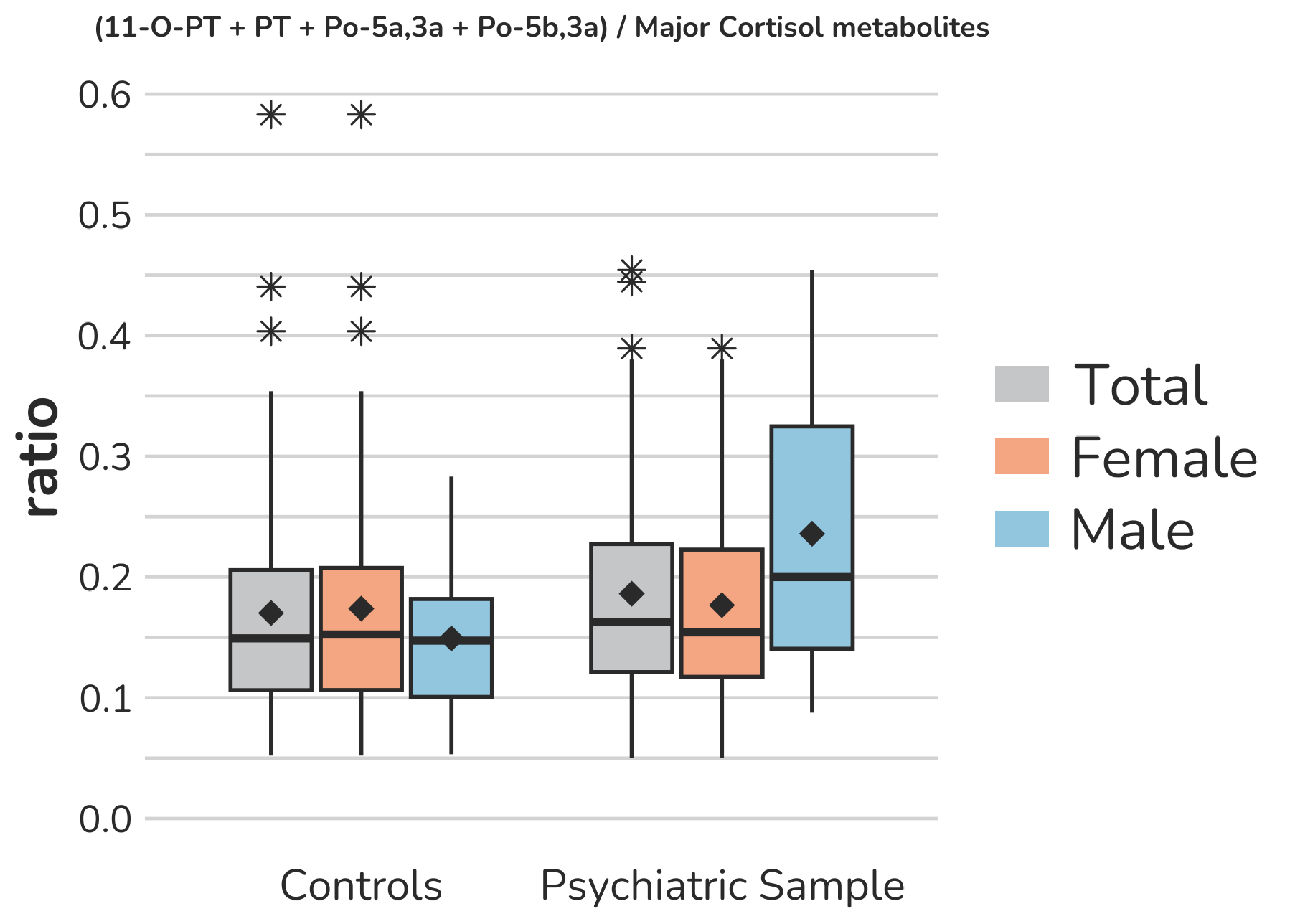

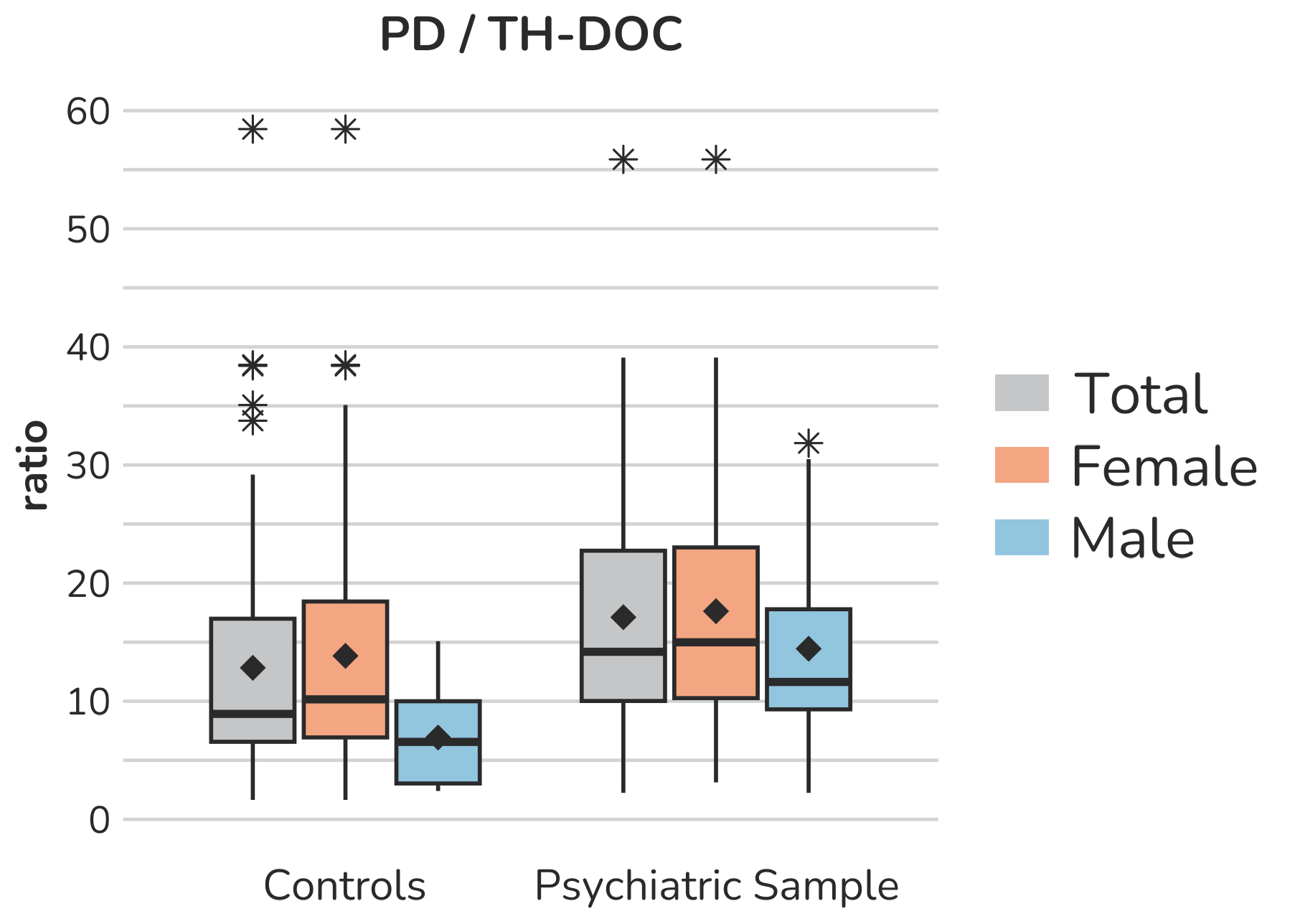

**Supplemental Figure S14 – Boxplots of ratios representing specific ratios as indicators for** **21-hydroxylase-activity.** Each box gives the 25th and 75th percentile (lower and upper boundary of the box), the median (horizontal line within each box), and the mean (rhombus within each box). The whiskers are defined by 1.5 times the interquartile range. For the 11-O-PT / α-Cl ratio, one male subject of the psychiatric sample with a ratio of 0.084 is excluded from the figure to enhance the visualization of the remaining values. Aside from this, all outliers are displayed (illustrated by *).

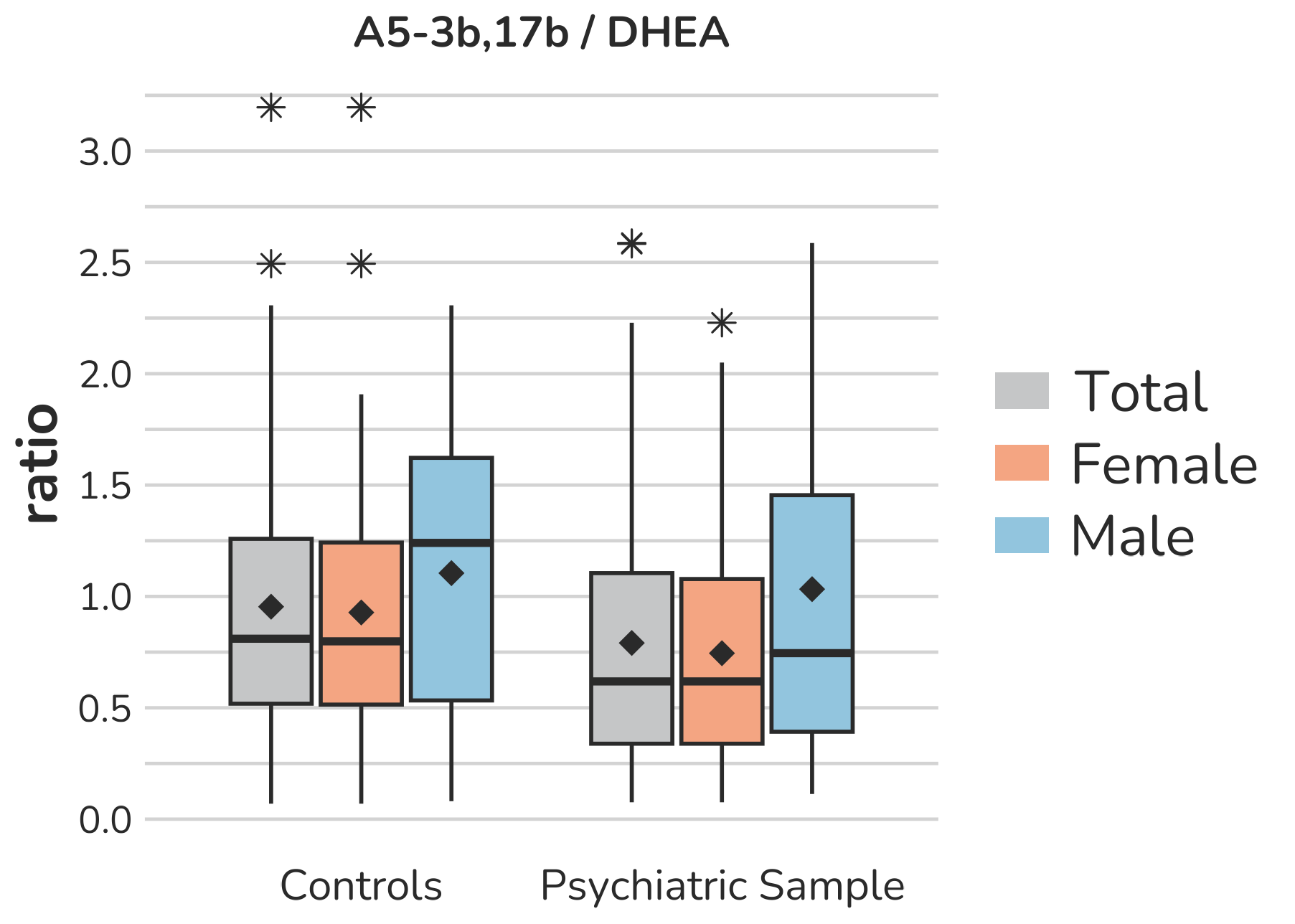

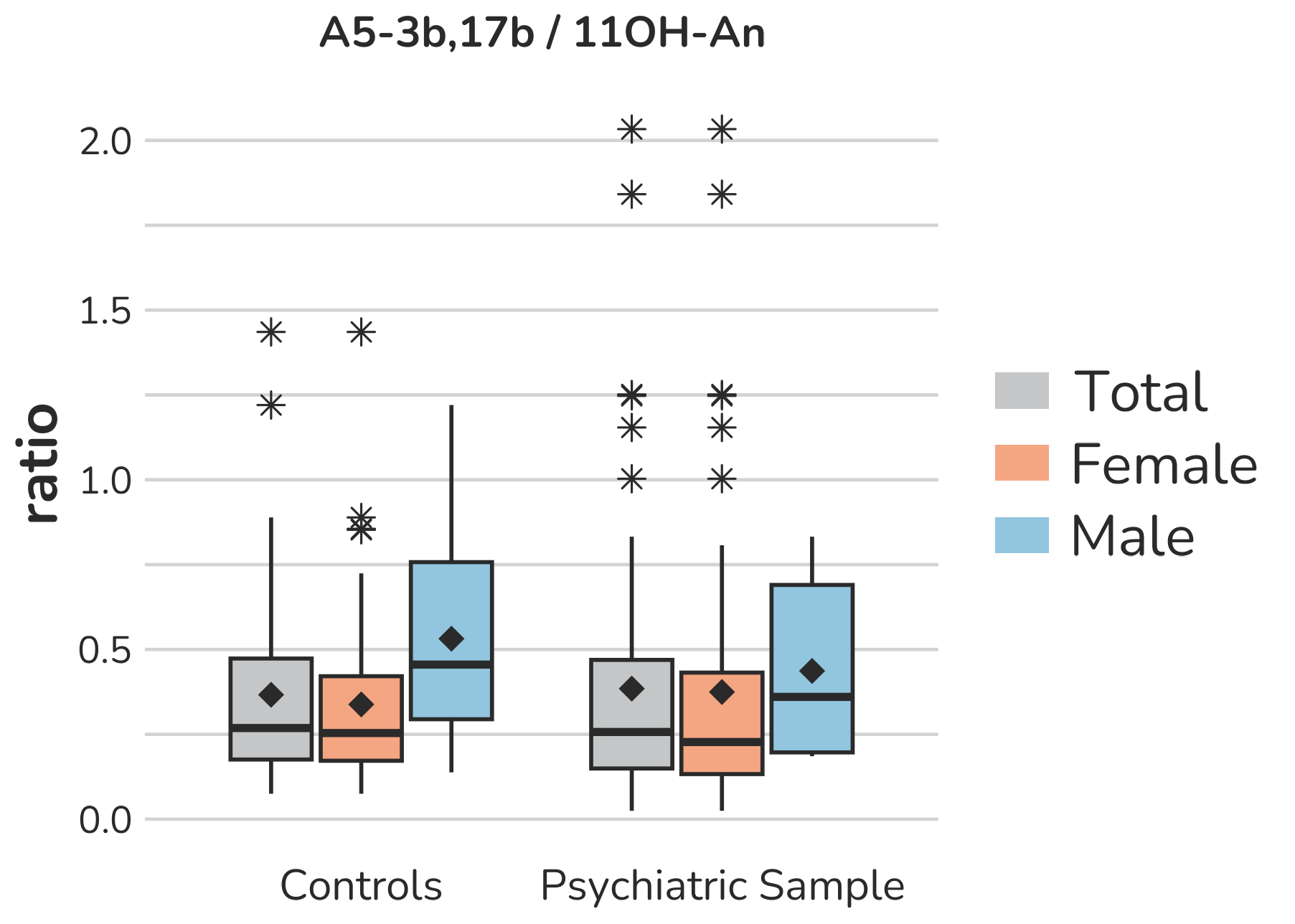

**Supplemental Figure S15 – Boxplots of ratios representing specific ratios as indicators for** **17β-hydroxysteroiddehydrogenase activity.** Each box gives the 25th and 75th percentile (lower and upper boundary of the box), the median (horizontal line within each box), and the mean (rhombus within each box). The whiskers are defined by 1.5 times the interquartile range. All outliers are presented (illustrated by *).

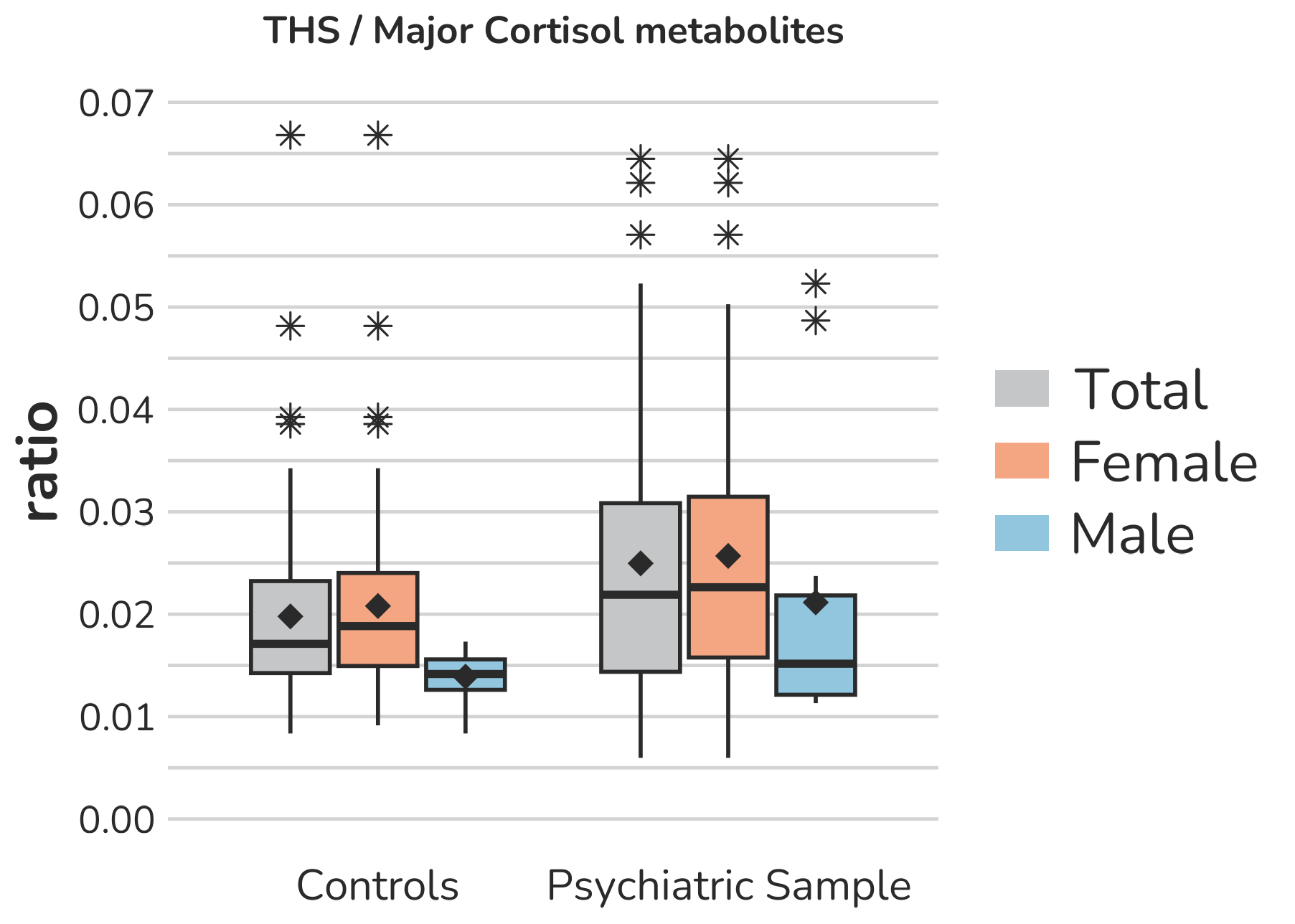

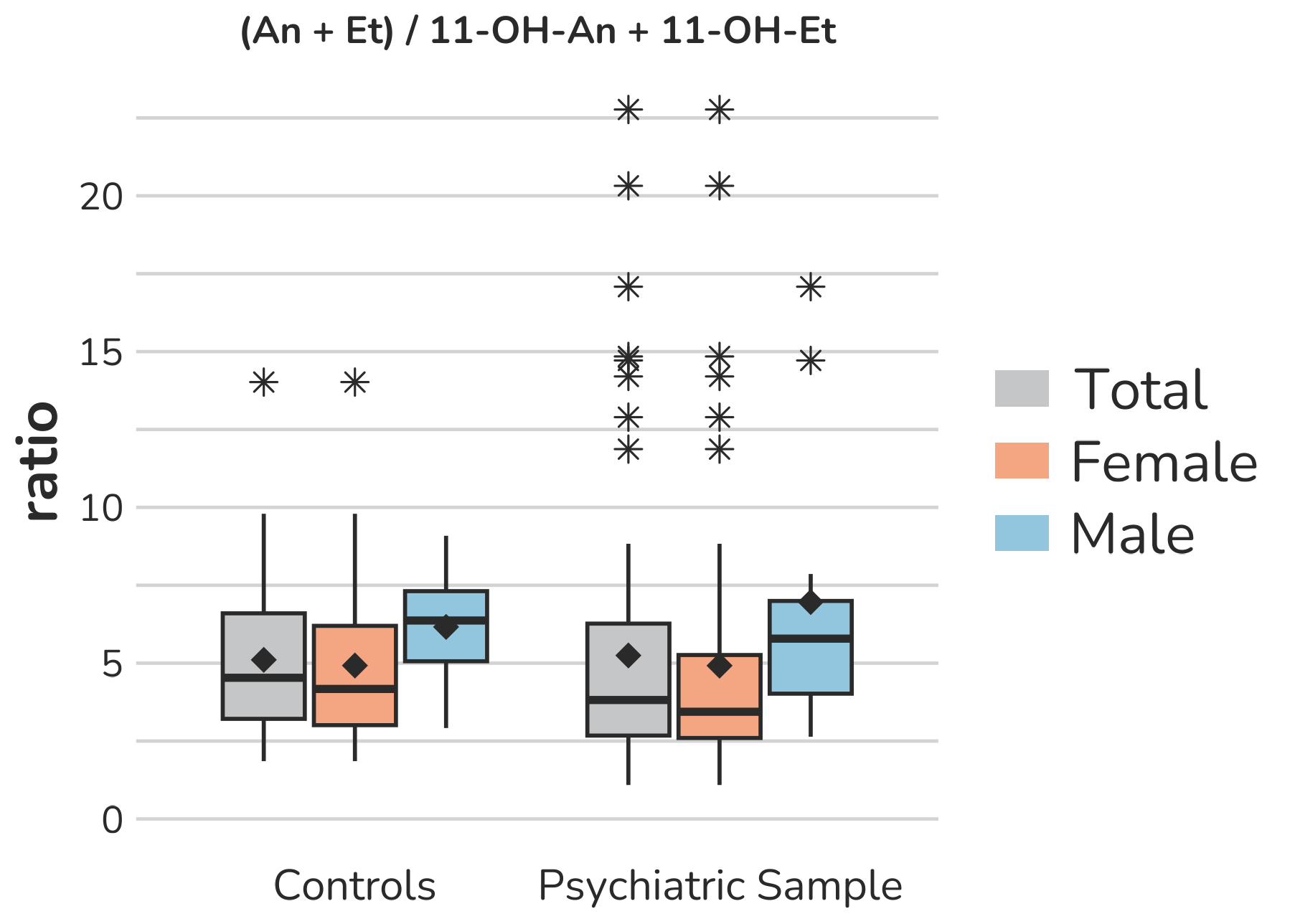

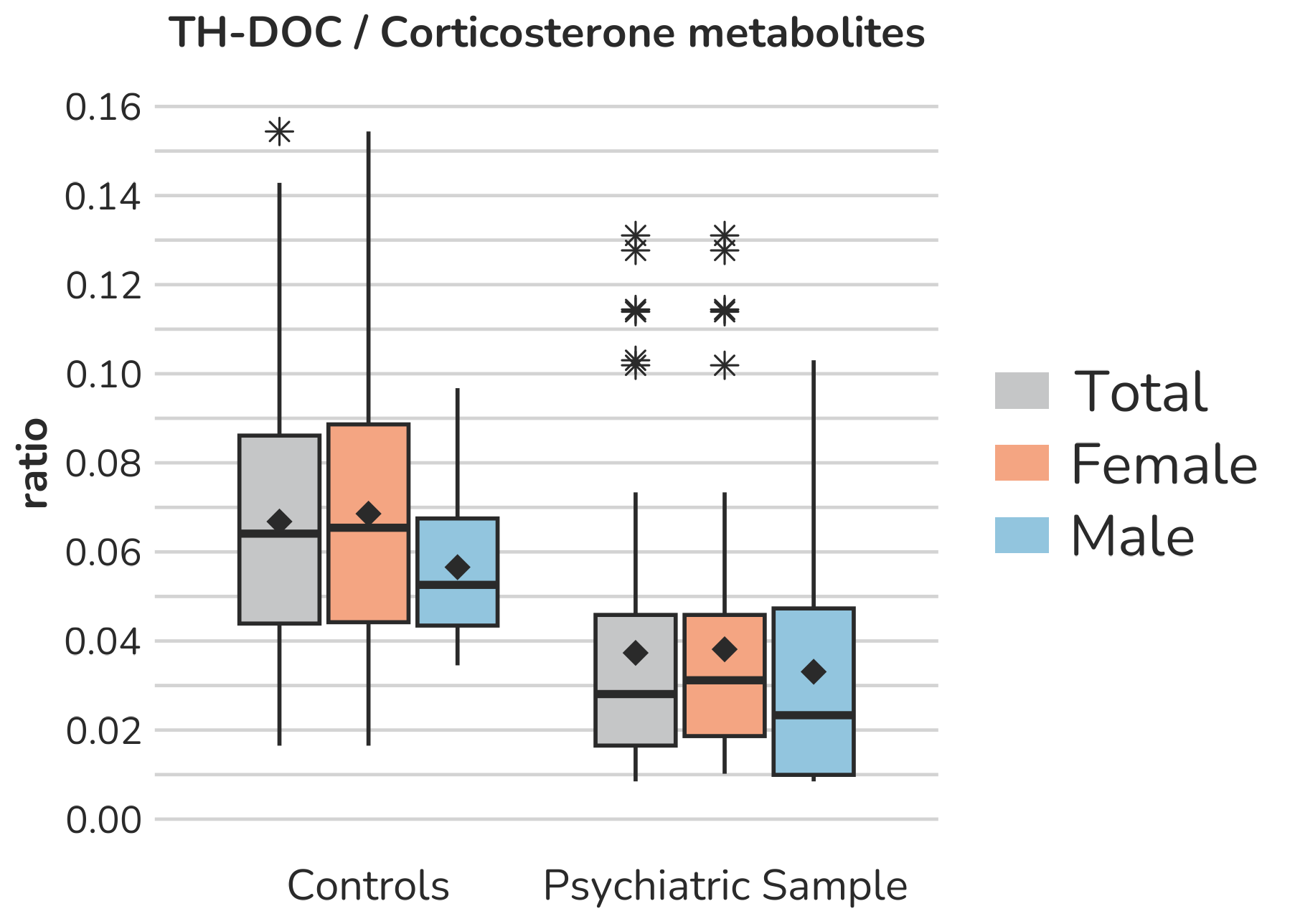

**Supplemental Figure S16 – Boxplots of ratios representing specific ratios as indicators for** **11β-hydroxylase activity.** Each box gives the 25th and 75th percentile (lower and upper boundary of the box), the median (horizontal line within each box), and the mean (rhombus within each box). The whiskers are defined by 1.5 times the interquartile range. All outliers are presented (illustrated by *).

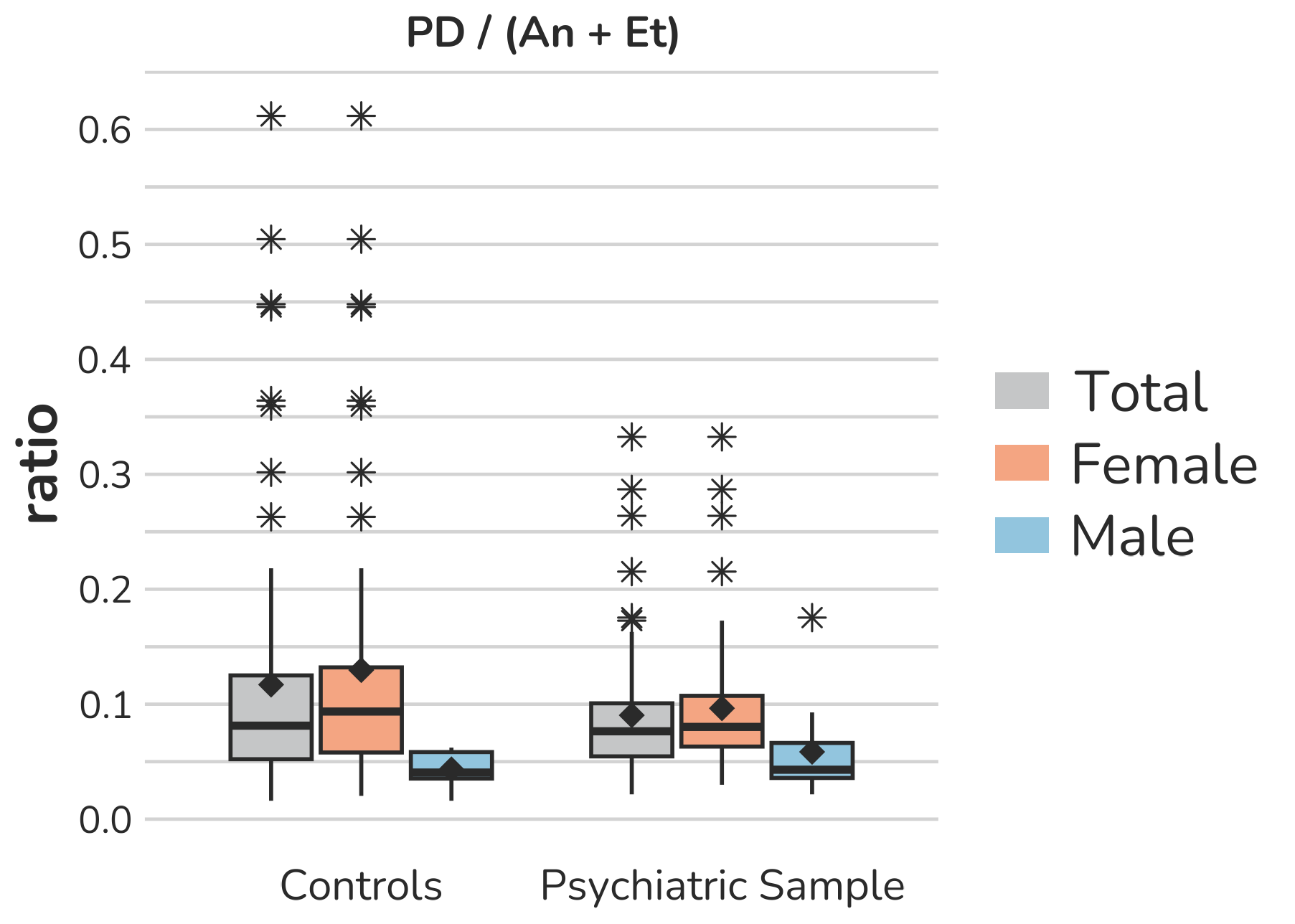

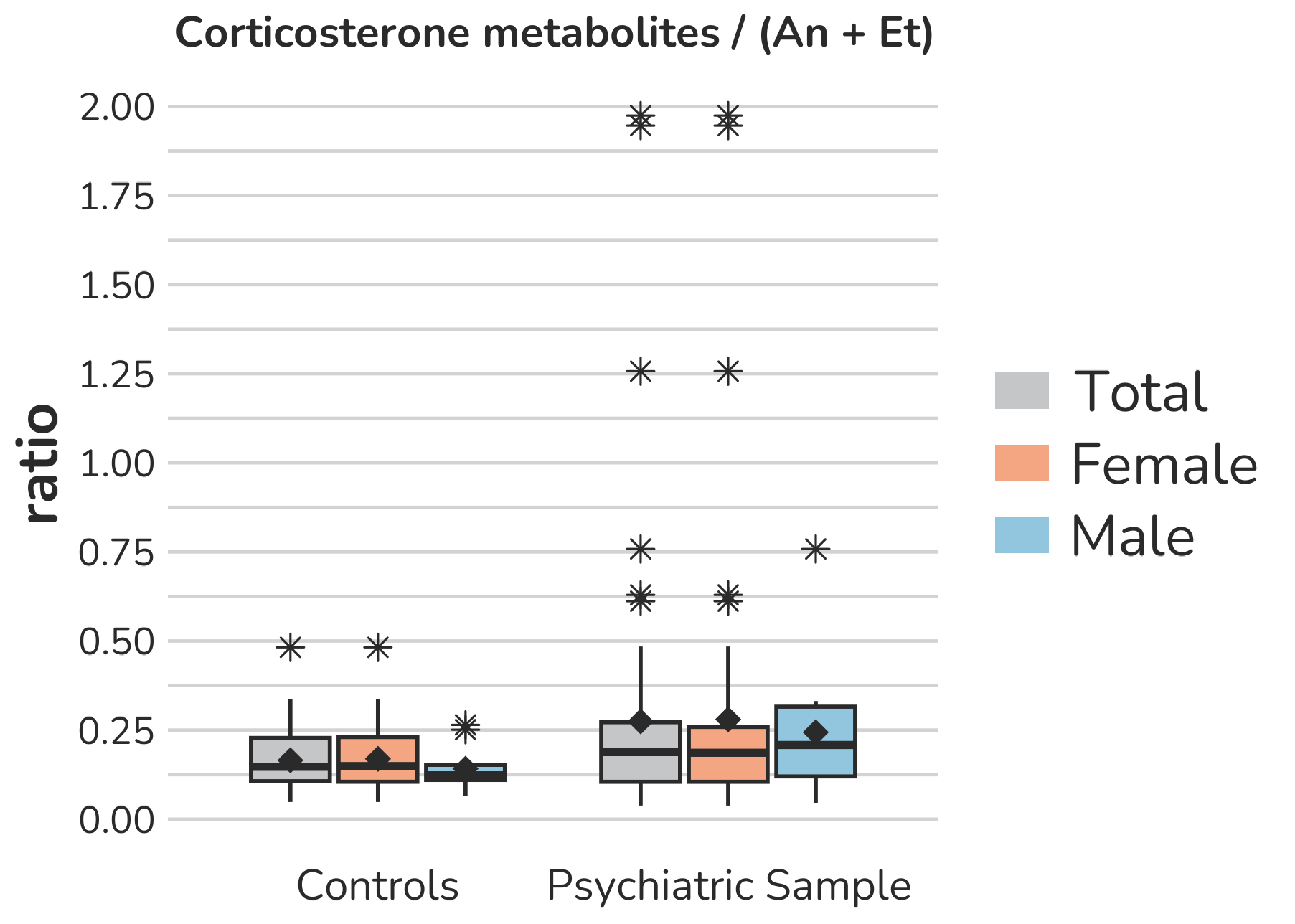

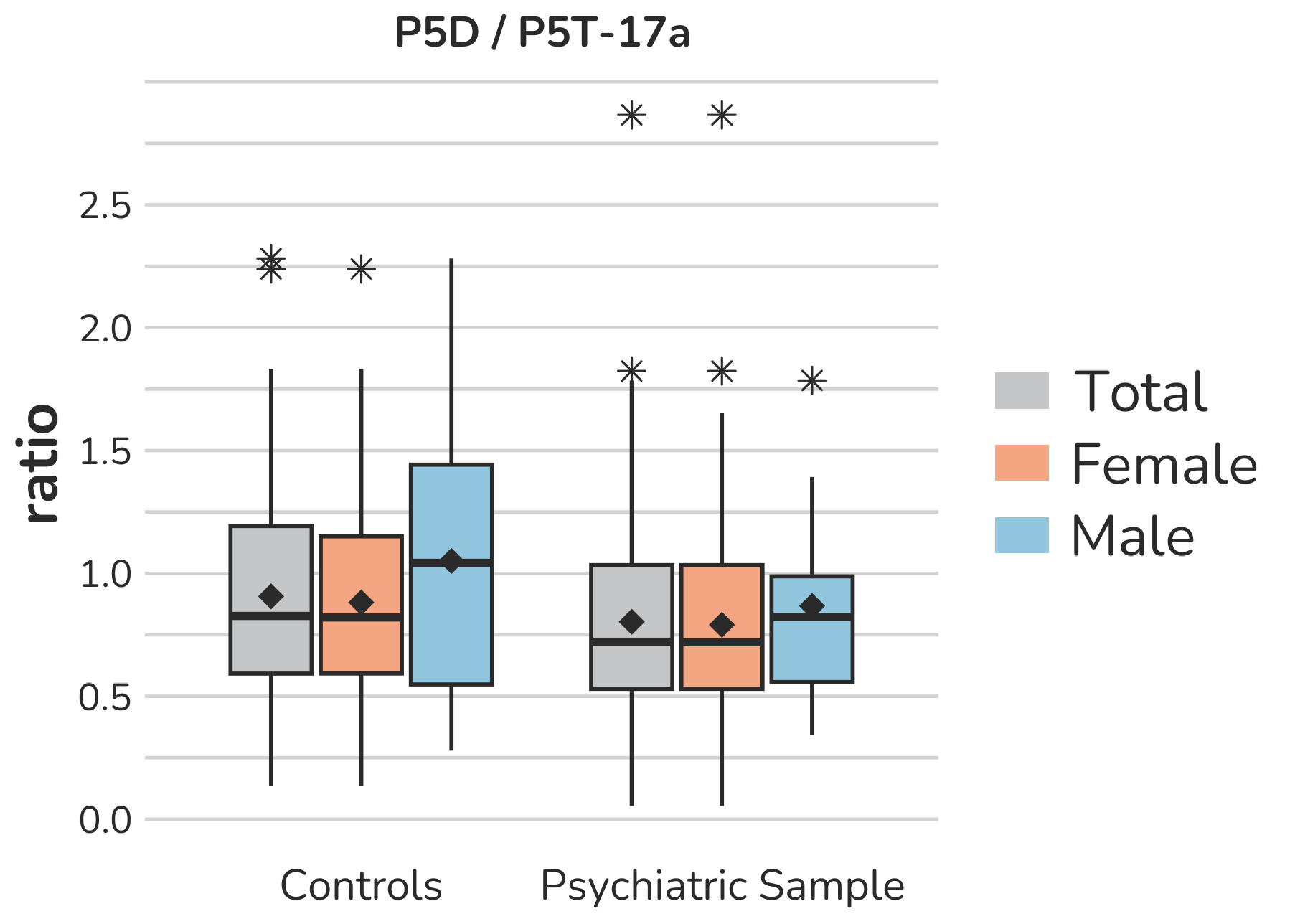

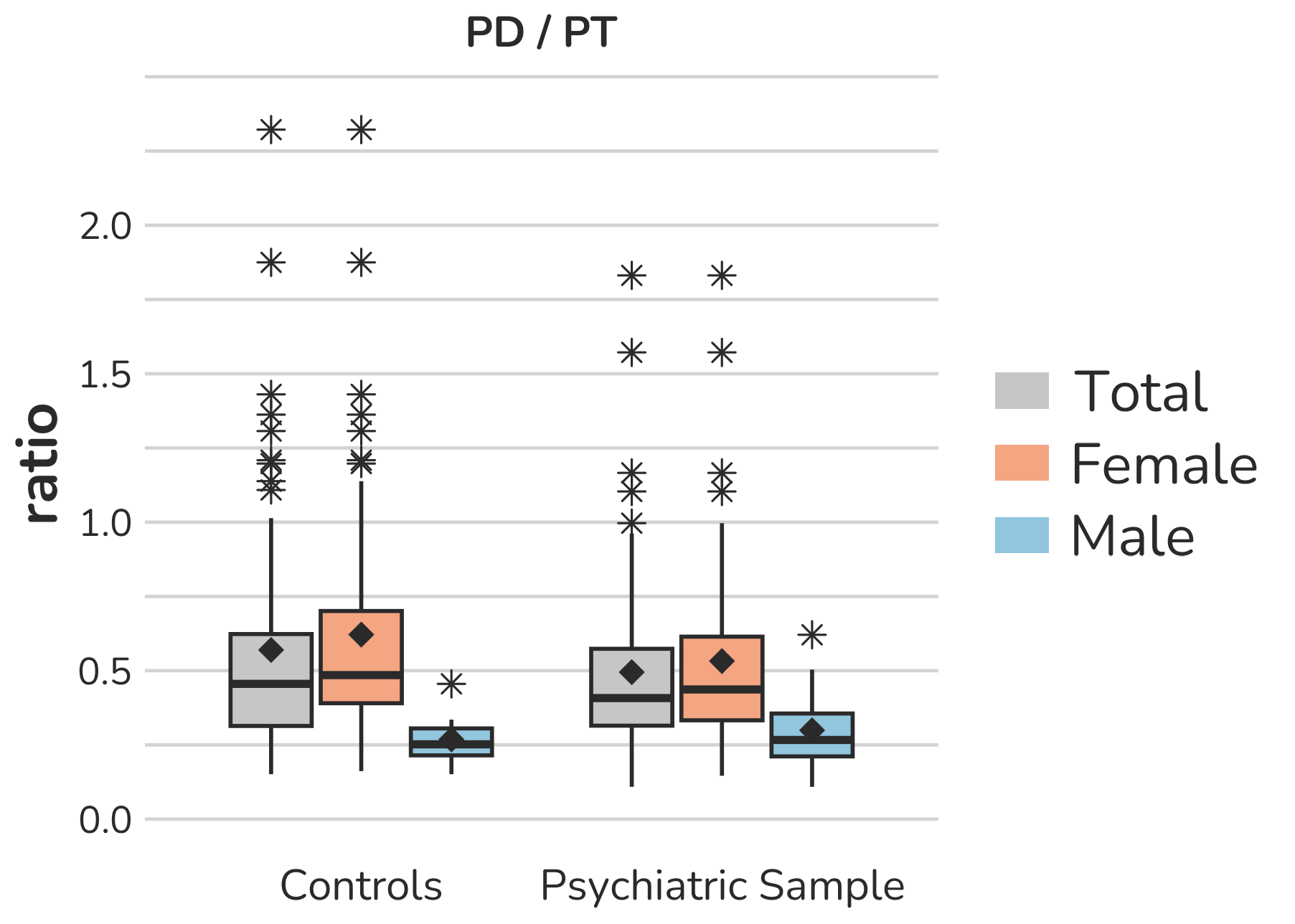

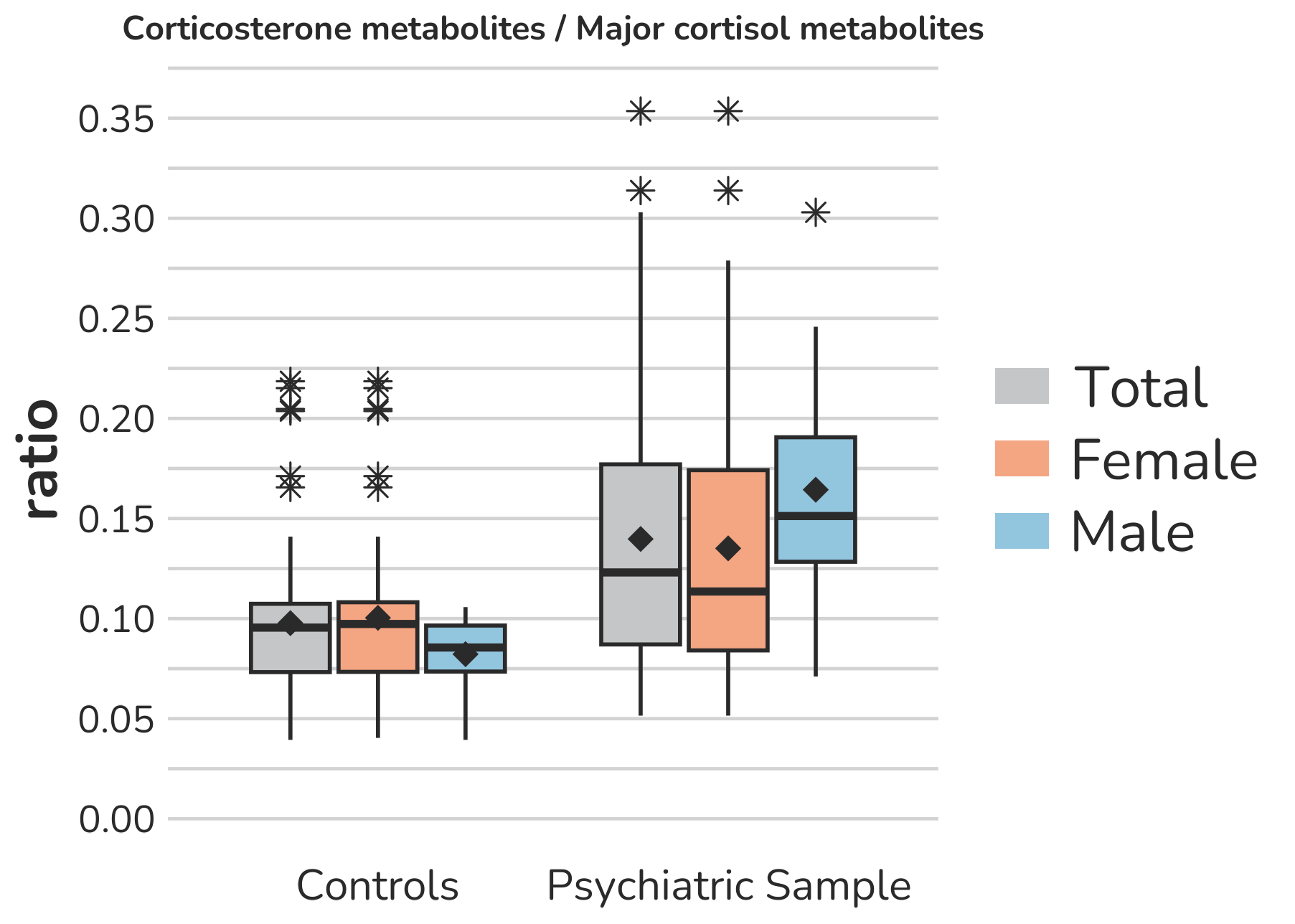

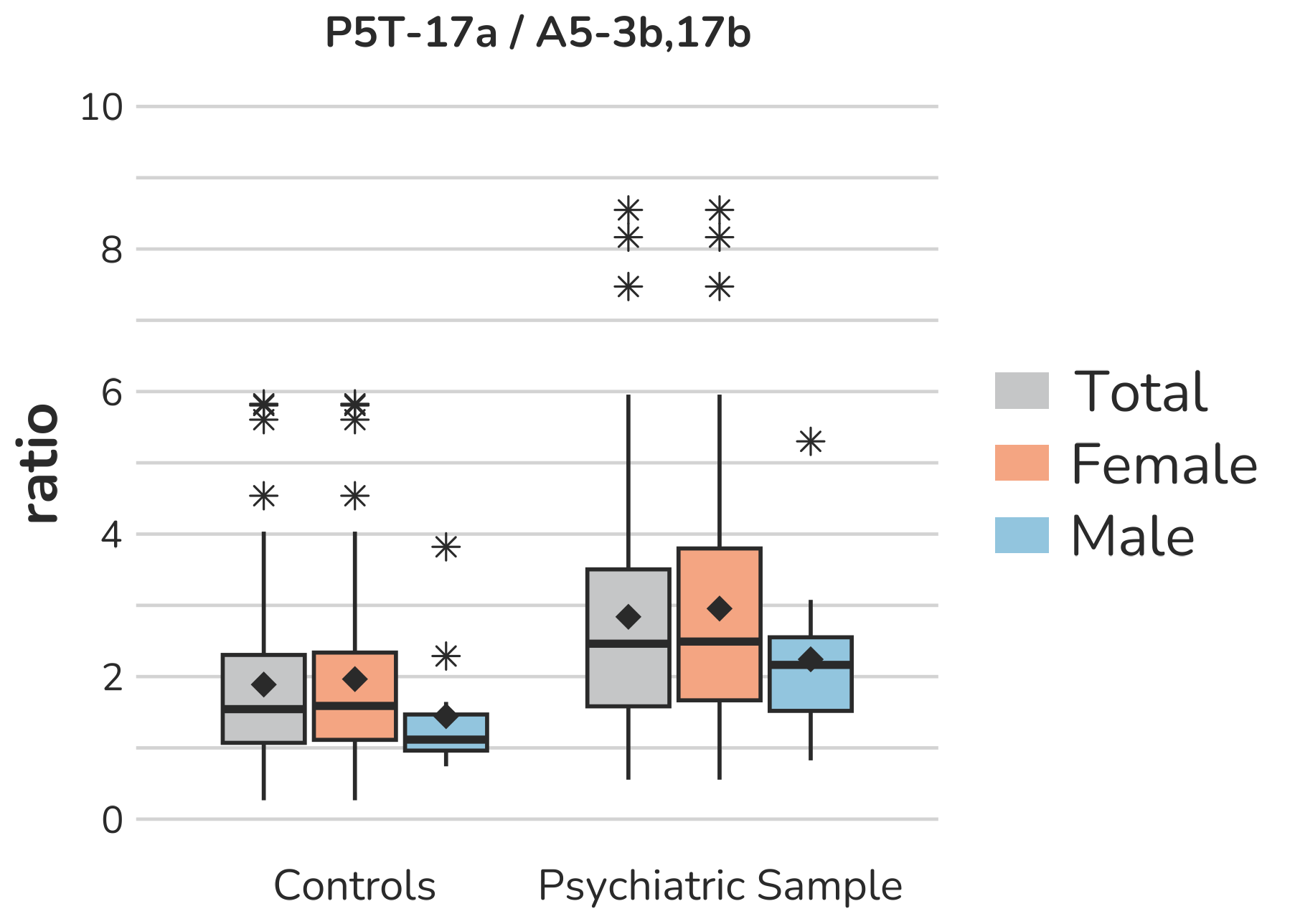

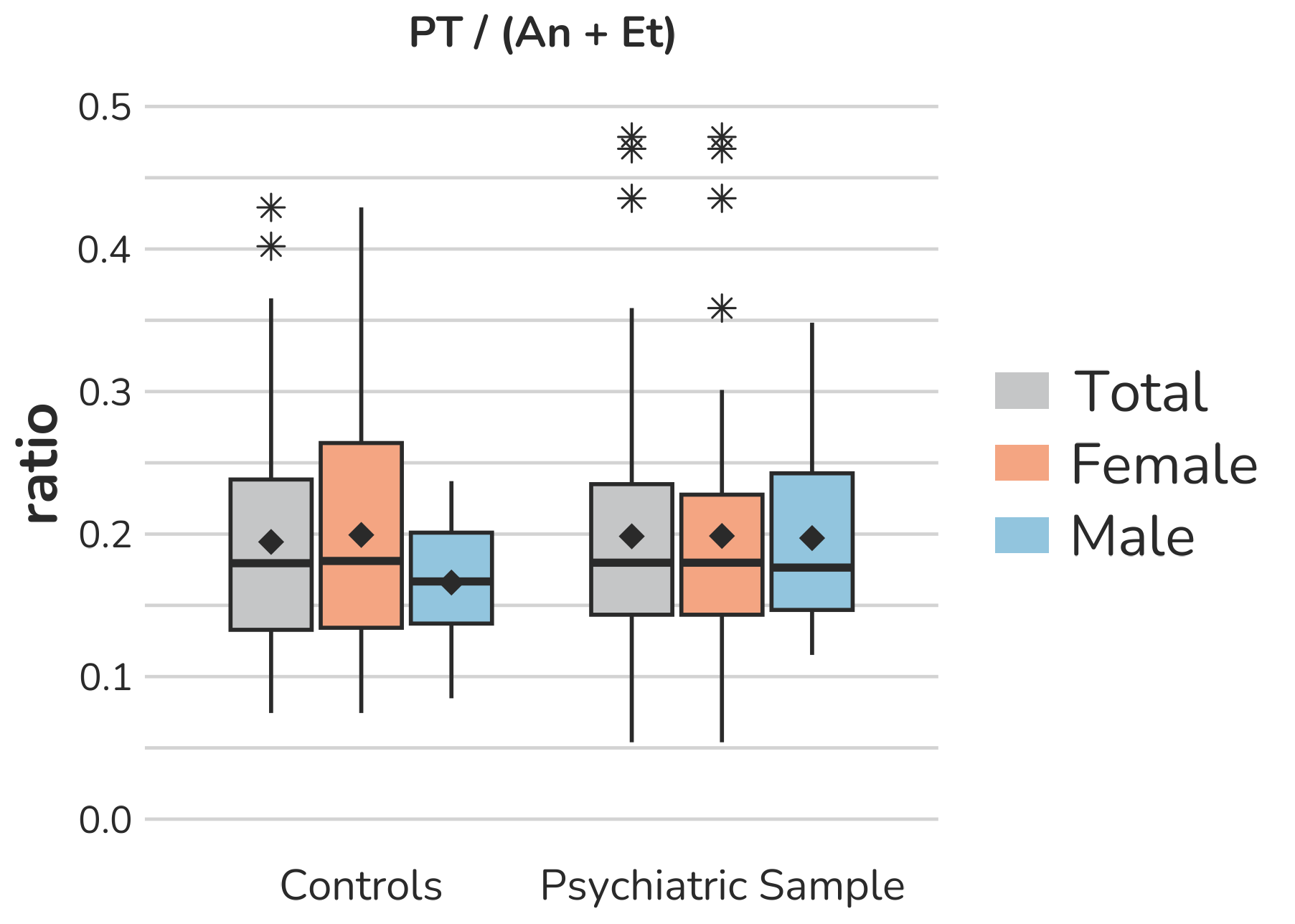

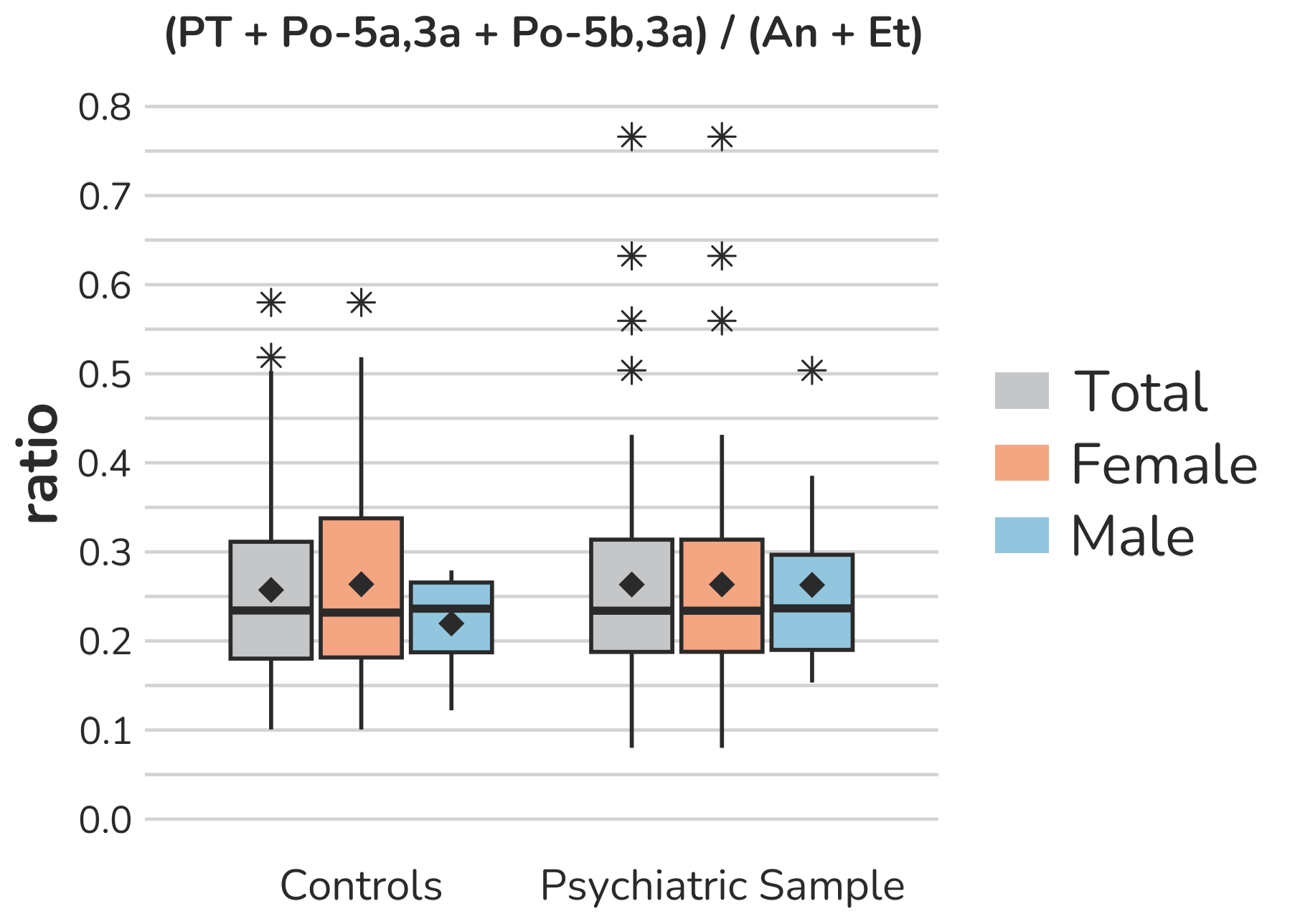

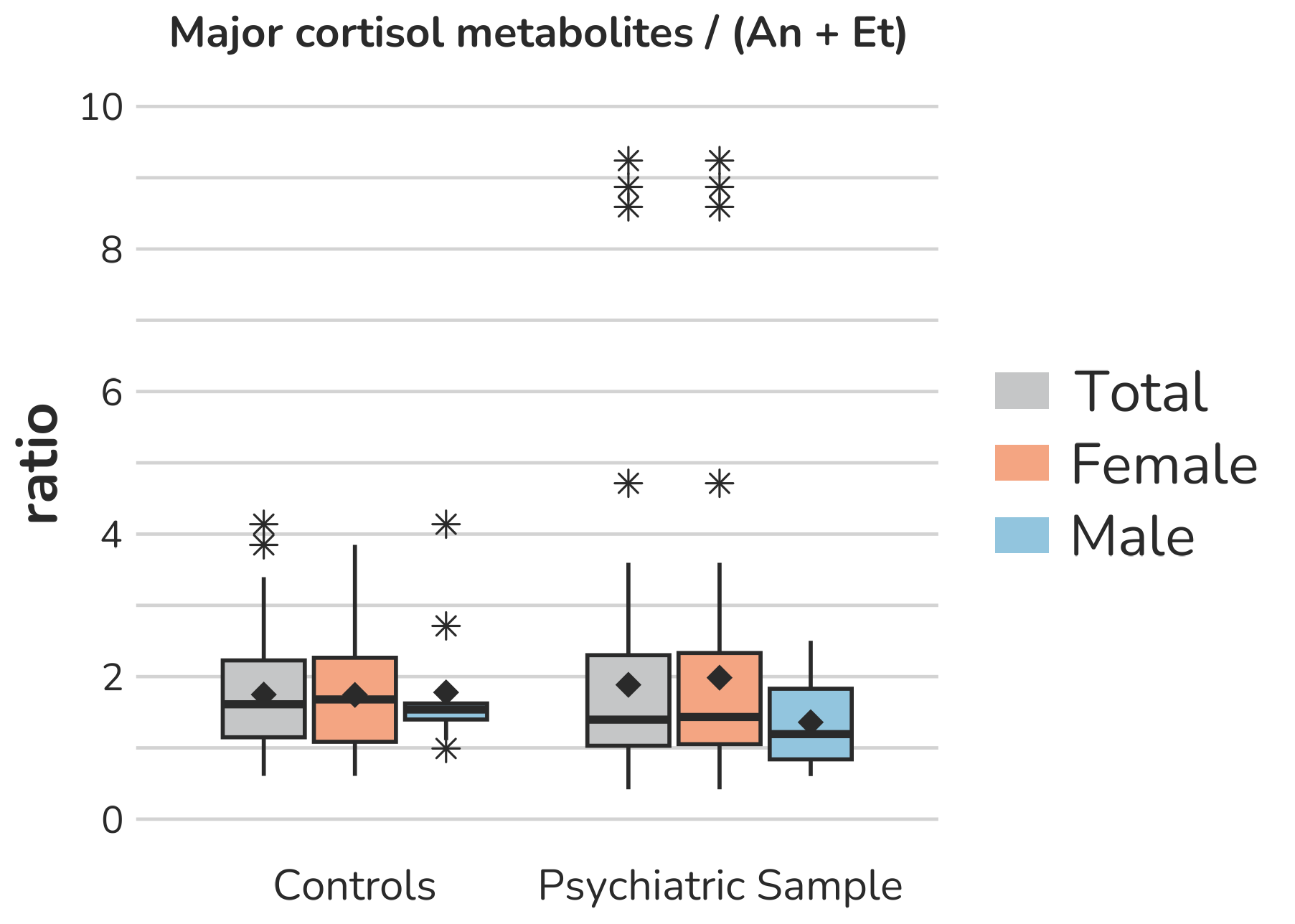

**Supplemental Figure S17 – Boxplots of ratios representing specific ratios as indicators for** **17-hydroxylase/17,20-lyase activity.** Each box gives the 25th and 75th percentile (lower and upper boundary of the box), the median (horizontal line within each box), and the mean (rhombus within each box). The whiskers are defined by 1.5 times the interquartile range. For the P5T-17α / A5-3β,17β, one female subject of the psychiatric sample with a ratio of 20.6 is excluded from the figure to enhance the visualization of the remaining values. Aside from this, all outliers are displayed (illustrated by *).

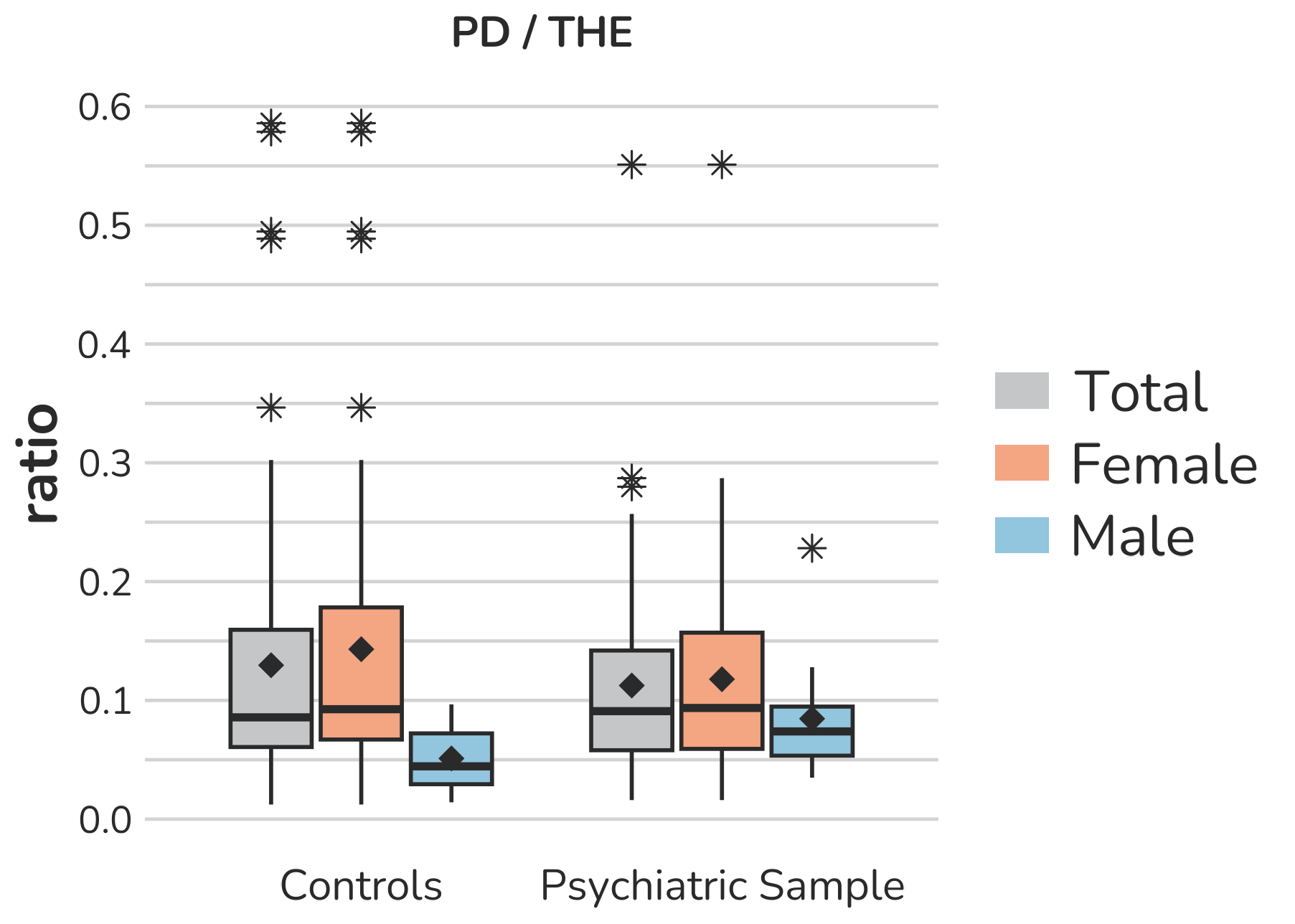

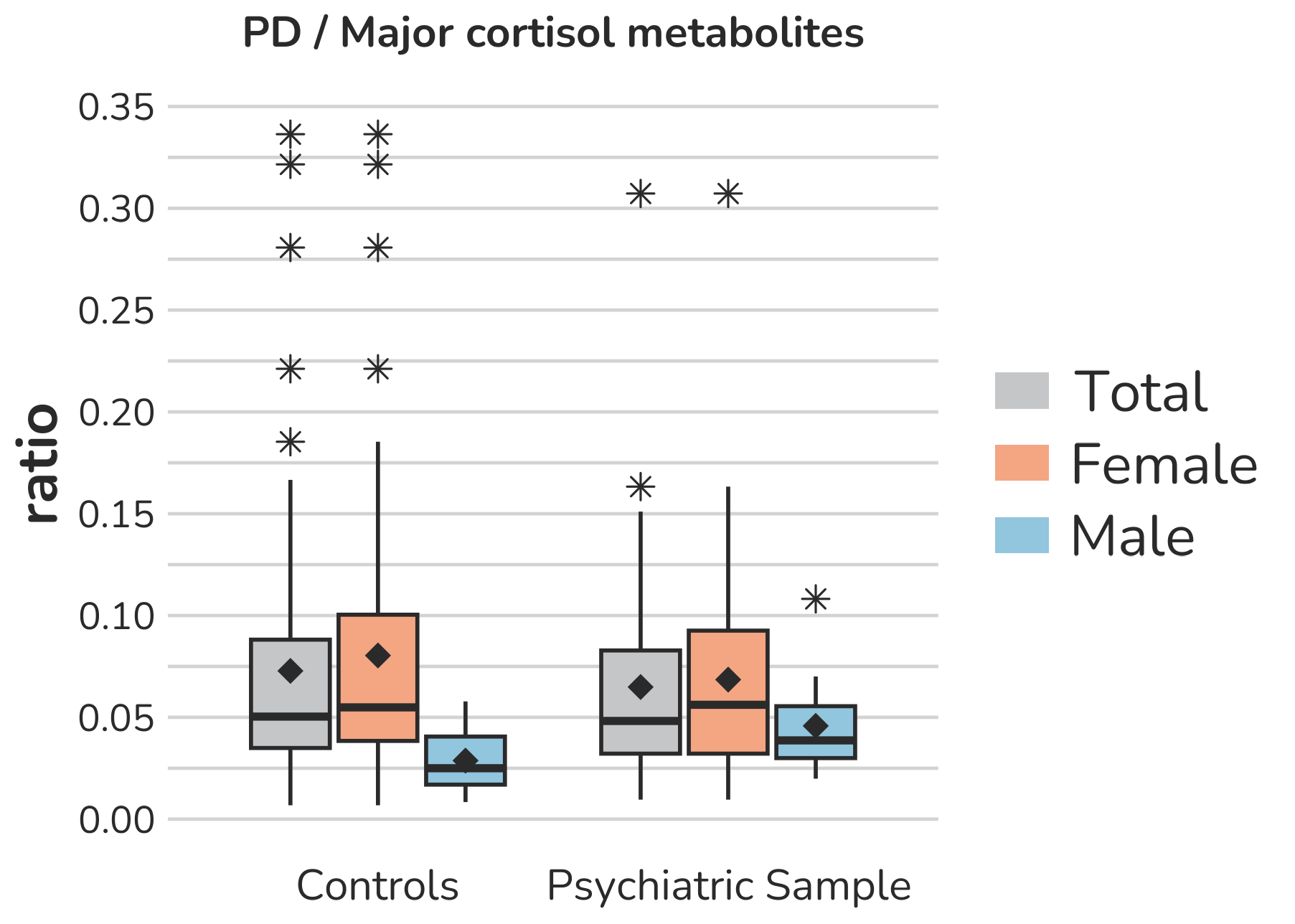

**Supplemental Figure S18 – Boxplots of ratios representing specific ratios as indicators for** **P450-Oxidoreductase (POR) activity.** Each box gives the 25th and 75th percentile (lower and upper boundary of the box), the median (horizontal line within each box), and the mean (rhombus within each box). The whiskers are defined by 1.5 times the interquartile range. All outliers are presented (illustrated by *).

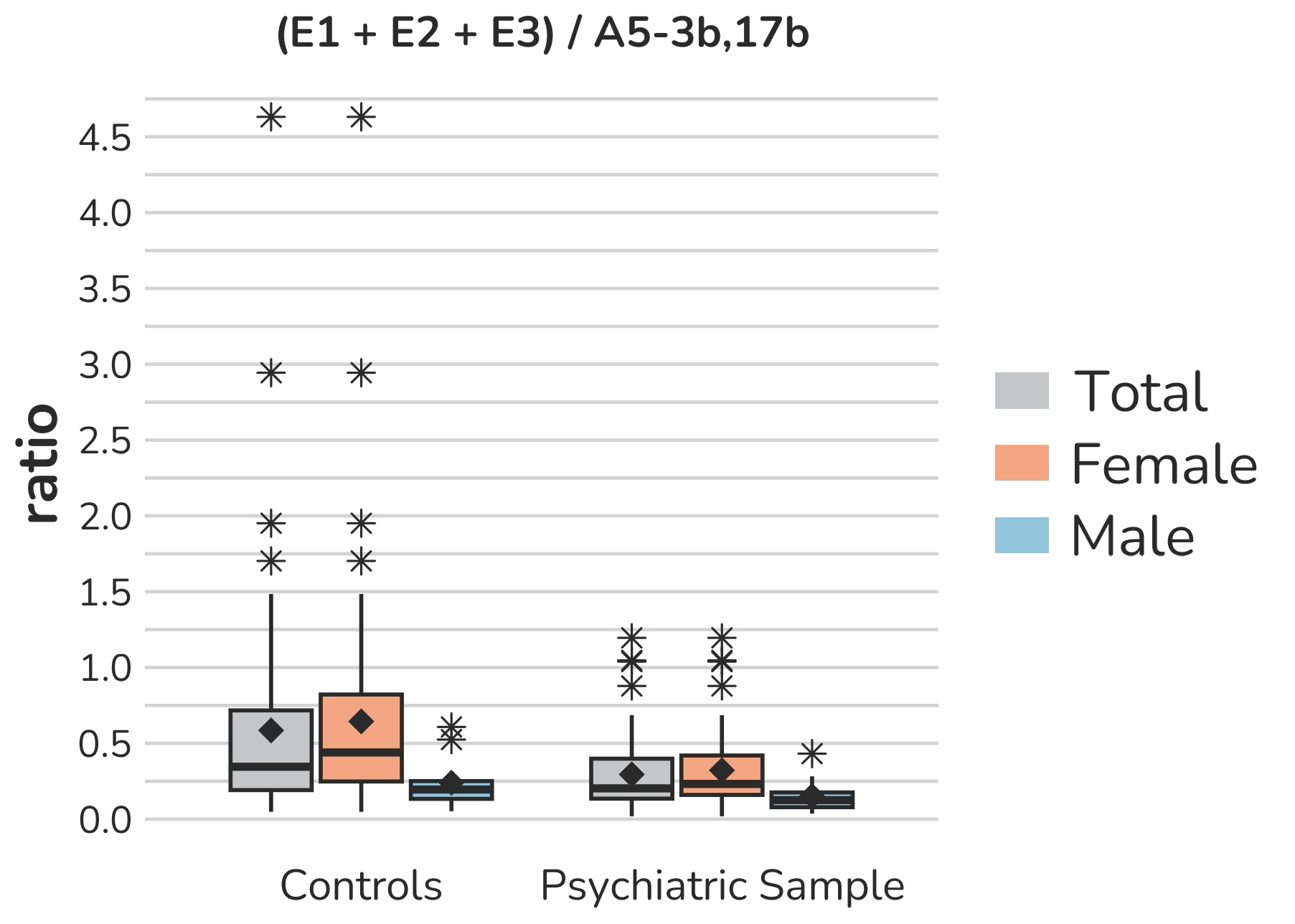

**Supplemental Figure S19 – Boxplots representing the (E1+E2+E3)/** **A5-3β,17β ratio as an indicator for** **aromatase activity.** Each box gives the 25th and 75th percentile (lower and upper boundary of the box), the median (horizontal line within each box), and the mean (rhombus within each box). The whiskers are defined by 1.5 times the interquartile range. All outliers are presented (illustrated by *).

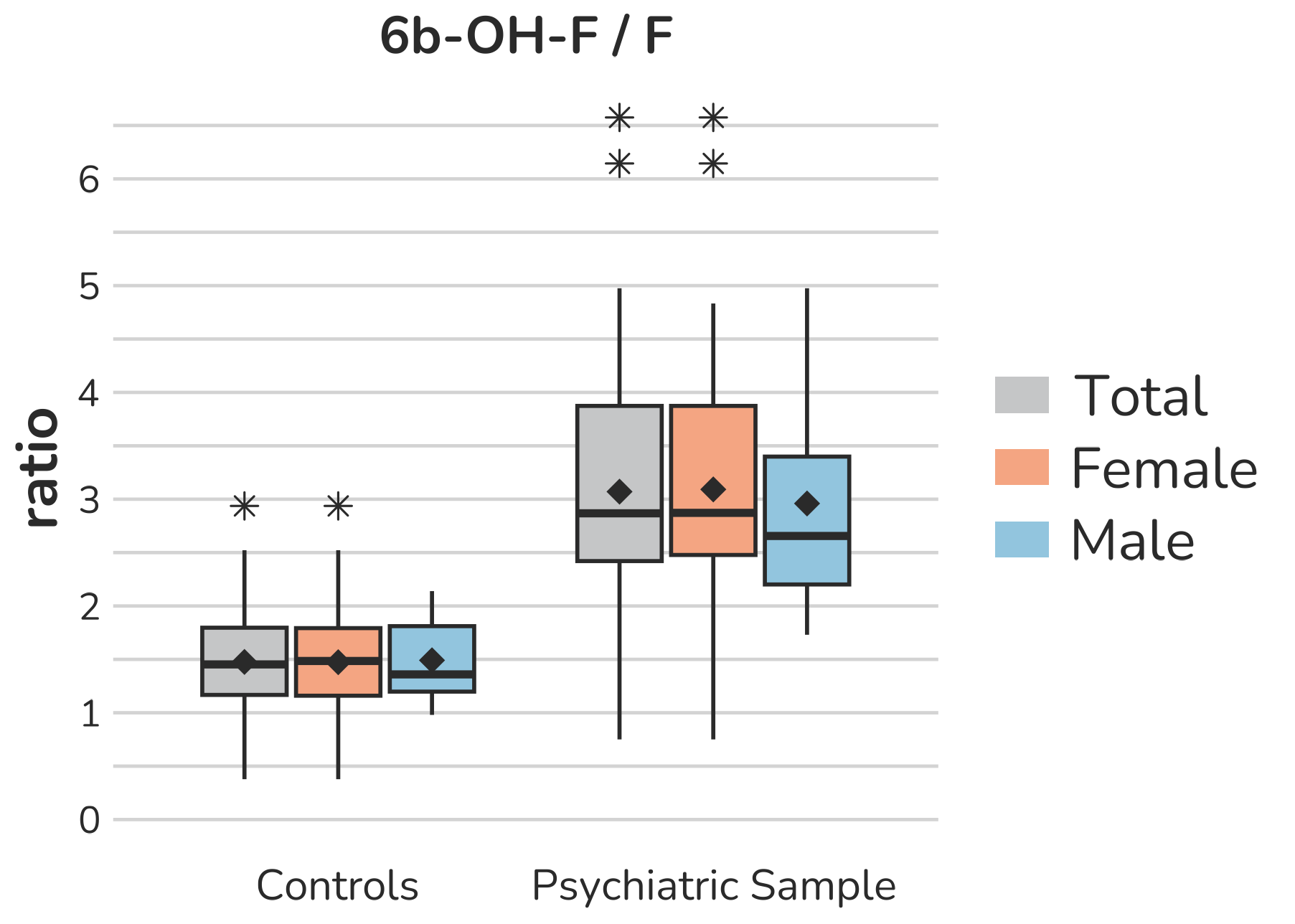

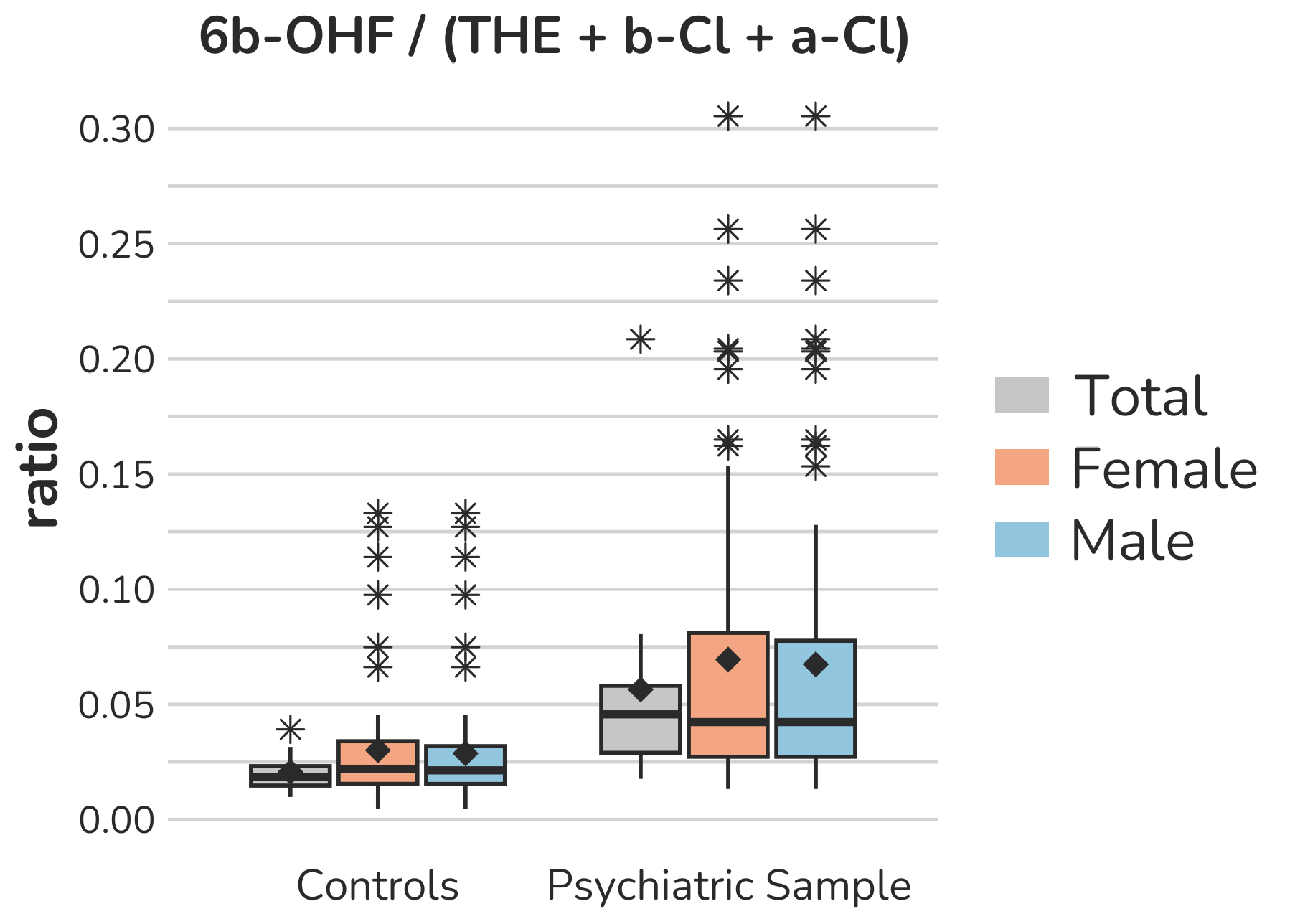

**Supplemental Figure S20 – Boxplots of ratios representing specific ratios as indicators for 6β-Hydroxylase** **(CYP3A4) activity.** Each box gives the 25th and 75th percentile (lower and upper boundary of the box), the median (horizontal line within each box), and the mean (rhombus within each box). The whiskers are defined by 1.5 times the interquartile range. All outliers are presented (illustrated by *).

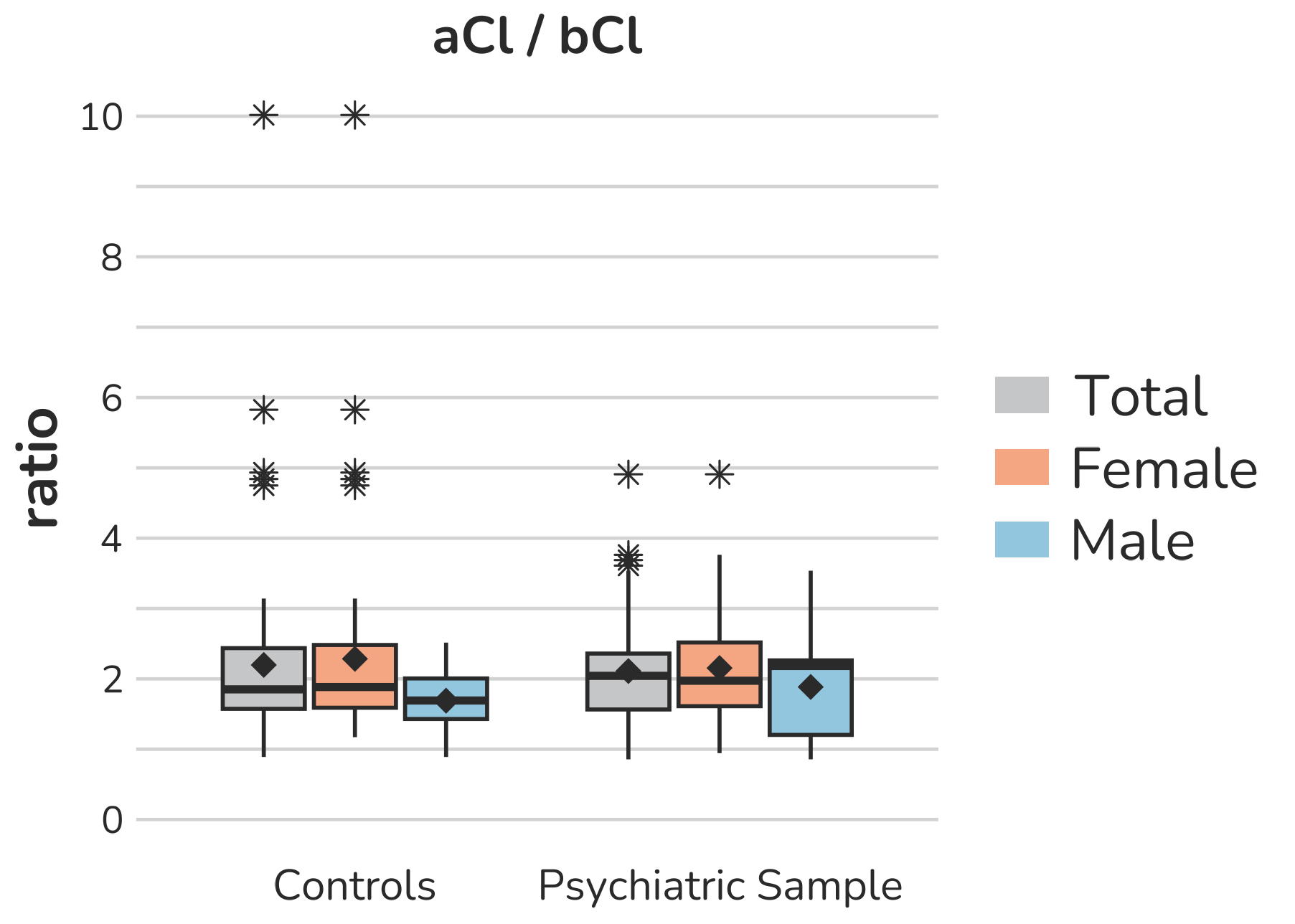

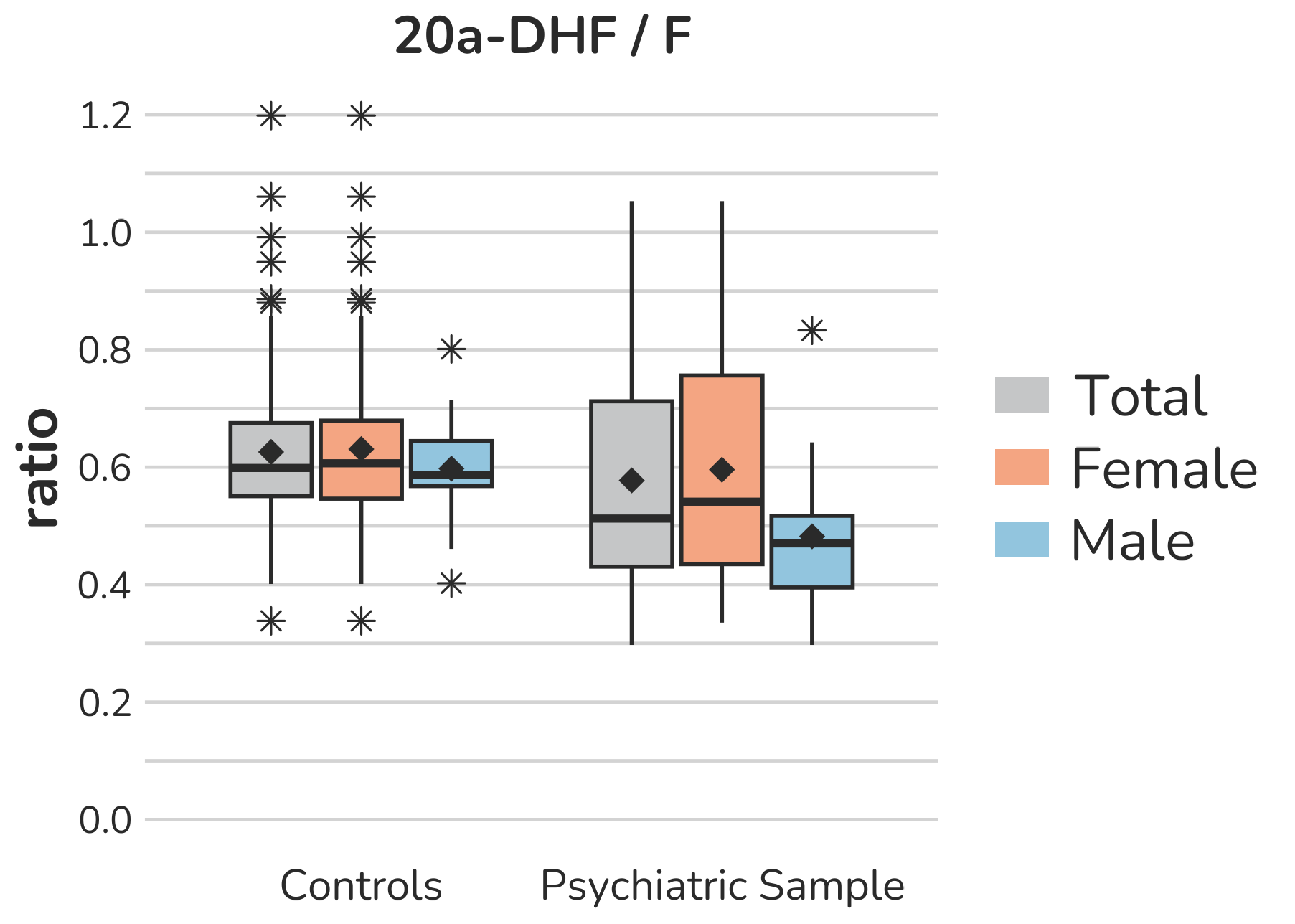

**Supplemental Figure S21 – Boxplots of ratios representing specific ratios as indicators for** **20α-HSD (AKR1C1) activity.** Each box gives the 25th and 75th percentile (lower and upper boundary of the box), the median (horizontal line within each box), and the mean (rhombus within each box). The whiskers are defined by 1.5 times the interquartile range. All outliers are presented (illustrated by *).

**Supplemental Figure S22– Sensitivity analyses: Association of storage time and 6β-OH-F excretion rates.** Sensitivity analyses revealed a significant negative association of storage time (time from sample collection to its analysis) and 6β-OH-F daily excretion rates. Figure S22 illustrates this association, separately for the control group and the psychiatric sample.

**Supplemental Figure S23 – Receiver operatic characteristic (ROC) curves of the performance of selected ratios (to classify between control subjects and the psychiatric sample.** The top ten most influential ratios from the multivariate ROC analyses were further investigated. The ROC curve for the most influential ratio, TH-DOC/corticosterone metabolites, is illustrated in Figure 3. The ROC curves are ordered (A-J) according to their average importance in the multivariate ROC analysis (rank 2nd two 10th). For clarity, ratios were abbreviated according to their order of appearance in Tab. 2 (e.g., 17-hydroxylase 5 = Corticosterone metabolites/Major cortisol metabolites).

**Supplemental Figure S24 –** **Sensitivity analyses for ROC curves of the TH-DOC / Corticosterone ratio as an indicator for 11β-hydroxylase activity. A:** Univariate ROC analysis of the TH-DOC / corticosterone metabolites ratio for the full sample (N = 75 in each group). The optimal cut-off point is indicated by the red dot, with the corresponding cut-off value, specificity, and sensitivity. **B:** Univariate ROC analysis of the restricted sample (excluding individuals using oral contraceptives, on psychotropic medication, or suffering from eating disorders, N = 50 in each group). **C:** Univariate ROC analysis to distinguish between patients with a clinical diagnosis of depression (either by K-SADS or clinical diagnosis) and the matched control group (N = 58 in each group). **D:** Univariate ROC analysis of the sample (N = 54 in each group) after excluding subjects with TH-DOC below the lower limit of detection (LLD). ROC curves B-D did not differ significantly from the full sample (ROC curve A) in multiple DeLong’s tests (B vs. A: p = 0.903; C vs. A p = 0.602; D vs. A p = 0.703).

**Supplemental Figure S25 – Univariate ROC analysis of the 6β-OH-F/F ratio (as an indicator for CYP3A4 activity).** ROC curve of the 6β-OH-F/F ratio demonstrated excellent discriminatory performance (test sample, AUC 0.891, Cohen’s d 1.74), with a sensitivity of 0.75 and a specificity of 0.90 at an optimal cut-off of 2.177. These results were validated in the verification sample (AUC 0.958). However, given the negative association with storage time (see Supplemental Table 5 and Supplemental Figure S22), this finding needs replication.
