## Supplemental Table 1 for "Urinary steroid metabolome shows adrenal, gonadal, and neuroactive steroid dysregulation in adolescents with depression"

| **Supplemental Table S1.** Number of subjects with metabolite concentrations below the lower limit of detection (LLD). | | |
| --- | --- | --- |
|  | N | Proportion |
| **progestagens (C21-steroids)** |  |  |
| P5D | 1 | 0.7% |
| Po | 8 | 5.3% |
| aPo | 13 | 8.7% |
| **mineralocorticoids (C21-steroids)** | |  |
| TH-DOC | 23 | 15.3% |
| **glucocorticoid (C21-steroids)** |  |  |
| F | 2 | 1.3% |
| 20α-DHF | 13 | 8.7% |
| 6β-OH-F | 9 | 6.0% |
| **androgens (C19-steroids)** |  |  |
| T | 73 | 48.7% |
| 5αAD-3α,17β | 2 | 1.3% |
| **estrogens (C18-steroids)** |  |  |
| E1 | 40 | 26.7% |
| E2 | 39 | 26.0% |
| E3 | 28 | 18.7% |

All remaining metabolites yielded values above the lower limit of detection (LLD) in all subjects. The values below the LLD were imputed using the R package zComposition (version 1.5.0-3) based on a log-ratio Expectation-Maximisation (lrEM) algorithm which allows for a robust (towards outliers and skewedness) imputation of left-censored data.
