## Supplemental Table 2 for "Urinary steroid metabolome shows adrenal, gonadal, and neuroactive steroid dysregulation in adolescents with depression"

| **Supplemental Table S2.** Full sample: Group comparisons of steroid excretion rates (µg/24h) of combined and single metabolites. | | | | | | | | | | | | |
| --- | --- | --- | --- | --- | --- | --- | --- | --- | --- | --- | --- | --- |
|  | Controls (N = 75) | | Psychiatric sample (N = 75) | | Unadjusted model  Group effect | | | Adjusted model  Group effect | | | | |
|  | Median | IQR | Median | IQR | d | p_raw_ | estimate | | 95%-CI | t | p_raw_ | p_FDRcorr_ |
| **Combined metabolites** |  |  |  |  |  |  | |  |  |  |  |  |
| Corticosterone metabolites  (allo-THB + THA + THB) | 321.0 | (243.9, 443.8) | 608.4 | (342.4, 1208.2) | 0.90 | <0.001 | | **273.34** | **(175.27, 371.40)** | **5.46** | **<0.001** | **<0.001** |
| Major Cortisol metabolites  (allo-THF + THF + THE) | 3980.9 | (2705.5, 4984.4) | 4836.5 | (3271.9, 8119.4) | 0.54 | 0.001 | | 886.35 | (77.50, 1695.20) | 2.15 | 0.033 | 0.073 |
| All Cortisol metabolites  (allo-THF + THF + THE + α-C + β-C + α-Cl +β-Cl) | 6236.3 | (4435.8, 7580.0) | 7631.1 | (5082.8, 12255.4) | 0.52 | 0.002 | | 1088.79 | (-75.60, 2253.19) | 1.83 | 0.069 | 0.121 |
| Major DHEA metabolites  (DHEA + 16α-OH-DHEA + A5T-16a) | 519.5 | (254.0, 1028.7) | 1253.8 | (569.8, 2796.2) | 0.80 | <0.001 | | **505.08** | **(220.13, 790.03)** | **3.47** | **0.001** | **0.003** |
| 11-deoxygenated androgens  (DHEA + 16α-OH-DHEA + A5T-16a + A5-3β,17α + A5-3β,17β + An + Et) | 3067.2 | (1940.6, 4600.8) | 5600.6 | (3413.1, 8224.5) | 0.87 | <0.001 | | **1651.48** | **(856.27, 2446.70)** | **4.07** | **<0.001** | **<0.001** |
| 11-oxygenated androgens  (11-OH-An + 11-O-An + 11-OH-Et) | 576.8 | (362.4, 786.1) | 840.8 | (563.6, 1369.2) | 0.72 | <0.001 | | **267.57** | **(114.30, 420.85)** | **3.42** | **0.001** | **0.003** |
| Overall androgen metabolites (∑C19):  (11-deoxygenated androgens + 11-oxygenated androgens) | 3680.4 | (2510.8, 5419.0) | 6721.0 | (4185.6, 9395.8) | 0.93 | <0.001 | | **1999.39** | **(1039.06, 2959.72)** | **4.08** | **<0.001** | **<0.001** |
| **progestagens (C21-steroids)** |  |  |  |  |  |  | |  |  |  |  |  |
| P5D | 125.4 | (69.7, 167.7) | 224.3 | (150.5, 333.4) | 0.95 | <0.001 | | **79.68** | **(44.81, 114.55)** | **4.48** | **<0.001** | **<0.001** |
| P5T-17α | 121.3 | (82.1, 206.4) | 327.6 | (191.8, 505.3) | 1.15 | <0.001 | | **141.34** | **(89.58, 193.10)** | **5.35** | **<0.001** | **<0.001** |
| Po | 34.6 | (20.1, 66.4) | 52.4 | (29.3, 88.7) | 0.37 | 0.027 | | 7.94 | (-4.15, 20.02) | 1.29 | 0.200 | 0.318 |
| aPo | 12.5 | (7.8, 26.1) | 18.3 | (11.2, 31.9) | 0.41 | 0.016 | | 2.36 | (-1.47, 6.18) | 1.21 | 0.229 | 0.331 |
| PD | 172.8 | (127.2, 285.4) | 257.8 | (174.2, 433.6) | 0.47 | 0.004 | | 51.14 | (1.64, 100.64) | 2.02 | 0.045 | 0.088 |
| PT | 410.1 | (275.2, 621.9) | 599.7 | (416.5, 894.6) | 0.70 | <0.001 | | **138.47** | **(43.65, 233.28)** | **2.86** | **0.005** | **0.015** |
| Po-5β,3α | 92.9 | (64.7, 174.8) | 157.5 | (104.8, 246.4) | 0.65 | <0.001 | | **44.02** | **(14.98, 73.05)** | **2.97** | **0.003** | **0.012** |
| Po-5α,3α | 18.3 | (11.6, 26.4) | 18.0 | (12.1, 26.6) | 0.08 | 0.608 | | -2.03 | (-5.40, 1.33) | -1.18 | 0.238 | 0.340 |
| **mineralocorticoids (C21-steroids)** | |  |  |  |  |  | |  |  |  |  |  |
| TH-DOC | 18.2 | (15.5, 25.6) | 17.9 | (12.3, 25.6) | -0.18 | 0.251 | | -2.58 | (-5.20, 0.05) | -1.92 | 0.057 | 0.103 |
| allo-THB | 152.2 | (104.9, 212.2) | 310.3 | (139.2, 603.5) | 0.75 | <0.001 | | **132.22** | **(72.75, 191.69)** | **4.36** | **<0.001** | **<0.001** |
| THA | 83.1 | (62.5, 118.8) | 136.8 | (80.6, 277.7) | 0.72 | <0.001 | | **48.55** | **(26.63, 70.46)** | **4.34** | **<0.001** | **<0.001** |
| THB | 81.3 | (58.3, 121.4) | 165.4 | (104.8, 325.9) | 1.09 | <0.001 | | **81.41** | **(55.83, 106.99)** | **6.24** | **<0.001** | **<0.001** |
| **glucocorticoid (C21-steroids)** | |  |  |  |  |  | |  |  |  |  |  |
| 11-O-PT | 6.9 | (5.2, 9.5) | 8.8 | (6.3, 14.6) | 0.49 | 0.003 | | **1.66** | **(0.29, 3.03)** | **2.37** | **0.019** | **0.046** |
| THS | 66.7 | (49.7, 89.0) | 100.1 | (63.7, 178.0) | 0.82 | <0.001 | | **37.69** | **(20.47, 54.92)** | **4.29** | **<0.001** | **<0.001** |
| F | 62.2 | (47.5, 75.0) | 84.2 | (40.0, 137.7) | 0.35 | 0.039 | | **18.37** | **(4.11, 32.63)** | **2.53** | **0.013** | **0.032** |
| 20α-DHF | 35.3 | (26.7, 48.5) | 49.1 | (17.3, 95.3) | 0.14 | 0.372 | | 4.51 | (-5.73, 14.76) | 0.86 | 0.390 | 0.506 |
| 6β-OH-F | 87.2 | (64.8, 116.8) | 246.1 | (115.9, 445.8) | 1.32 | <0.001 | | **155.78** | **(119.79, 191.76)** | **8.49** | **<0.001** | **<0.001** |
| allo-THF | 660.9 | (394.0, 1085.0) | 754.4 | (387.9, 1157.2) | 0.20 | 0.242 | | -19.06 | (-183.76, 145.65) | -0.23 | 0.821 | 0.883 |
| THF | 887.0 | (682.1, 1073.3) | 1252.3 | (720.9, 2158.2) | 0.70 | <0.001 | | **369.01** | **(166.34, 571.67)** | **3.57** | **<0.001** | **0.002** |
| α-C | 135.4 | (111.2, 168.7) | 207.1 | (126.2, 286.1) | 0.58 | <0.001 | | **37.47** | **(7.41, 67.53)** | **2.44** | **0.016** | **0.038** |
| β-C | 302.0 | (207.1, 481.2) | 381.1 | (239.2, 547.0) | 0.26 | 0.104 | | 29.69 | (-44.32, 103.70) | 0.79 | 0.433 | 0.544 |
| THE | 2268.7 | (1539.5, 2794.2) | 2810.2 | (1832.9, 4620.4) | 0.52 | 0.002 | | 568.26 | (56.09, 1080.43) | 2.17 | 0.031 | 0.069 |
| α-Cl | 1068.3 | (826.4, 1398.5) | 1281.2 | (863.5, 1843.3) | 0.35 | 0.043 | | 55.82 | (-128.28, 239.93) | 0.59 | 0.553 | 0.636 |
| β-Cl | 598.5 | (415.9, 777.3) | 639.2 | (453.2, 928.4) | 0.18 | 0.264 | | 15.47 | (-93.96, 124.91) | 0.28 | 0.782 | 0.850 |
| **androgens (C19-steroids)** |  |  |  |  |  |  | |  |  |  |  |  |
| DHEA | 87.4 | (50.1, 202.9) | 166.5 | (77.1, 465.1) | 0.54 | 0.001 | | 46.2 | (-1.94, 94.33) | 1.88 | 0.062 | 0.111 |
| 16α-OH-DHEA | 195.9 | (90.8, 458.4) | 565.7 | (226.1, 1281.3) | 0.82 | <0.001 | | **225.5** | **(103.21, 347.79)** | **3.61** | **<0.001** | **0.002** |
| A5T*-*16α | 190.2 | (108.7, 330.8) | 380.7 | (223.7, 701.8) | 0.90 | <0.001 | | **155.93** | **(73.75, 238.11)** | **3.72** | **<0.001** | **0.001** |
| A5-3β,17α | 37.5 | (27.4, 54.0) | 74.3 | (43.7, 123.2) | 1.06 | <0.001 | | **28.17** | **(16.78, 39.56)** | **4.85** | **<0.001** | **<0.001** |
| A5-3β,17β | 89.5 | (50.2, 133.2) | 107.2 | (71.2, 212.7) | 0.45 | 0.006 | | 15.04 | (-9.10, 39.19) | 1.22 | 0.224 | 0.331 |
| An | 1233.7 | (766.6, 1886.0) | 1794.5 | (1049.7, 2529.1) | 0.54 | 0.001 | | 251.21 | (-59.44, 561.86) | 1.58 | 0.115 | 0.189 |
| Et | 928.2 | (623.5, 1478.3) | 1649.3 | (1160.2, 2242.2) | 0.80 | <0.001 | | **518.33** | **(271.97, 764.70)** | **4.12** | **<0.001** | **<0.001** |
| T | 7.2 | (4.1, 15.0) | 10.7 | (6.2, 15.1) | 0.20 | 0.209 | | 0.88 | (-1.39, 3.15) | 0.76 | 0.449 | 0.554 |
| 5αAD-3α,17β | 24.0 | (14.9, 55.0) | 36.6 | (20.7, 59.9) | 0.49 | 0.003 | | 4.58 | (-2.84, 11.99) | 1.21 | 0.228 | 0.331 |
| 11-OH-An | 301.2 | (204.9, 441.5) | 445.7 | (259.1, 828.4) | 0.65 | <0.001 | | **127.39** | **(34.39, 220.38)** | **2.68** | **0.008** | **0.023** |
| 11-O-An | 40.5 | (28.7, 51.9) | 47.1 | (28.9, 59.4) | 0.18 | 0.262 | | 2.48 | (-4.44, 9.41) | 0.70 | 0.483 | 0.575 |
| 11-OH-Et | 188.9 | (67.8, 311.6) | 283.9 | (88.5, 484.8) | 0.41 | 0.016 | | **92.3** | **(23.54, 161.07)** | **2.63** | **0.009** | **0.025** |
| **estrogens (C18-steroids)** |  |  |  |  |  |  | |  |  |  |  |  |
| E1 | 16.2 | (8.9, 25.1) | 12.8 | (9.2, 18.3) | -0.24 | 0.136 | | -3.08 | (-6.14, -0.02) | -1.97 | 0.051 | 0.094 |
| E2 | 5.8 | (4.3, 7.7) | 4.0 | (2.9, 5.8) | -0.63 | <0.001 | | **-1.86** | **(-2.67, -1.05)** | **-4.51** | **<0.001** | **<0.001** |
| E3 | 10.5 | (6.5, 17.3) | 9.8 | (6.2, 14.4) | -0.12 | 0.488 | | -1.44 | (-3.75, 0.87) | -1.22 | 0.223 | 0.331 |

Overview of the comparison between the psychiatric sample of depressed adolescents and matched controls. The adjusted model refers to the rank-based regression analyses (adjusted for age and z-transformed BMI). The table reports test statistics for the group effect and FDR-corrected p-values (considering all comparisons made for individual metabolites, combined metabolites, and ratios). The unadjusted model refers to group comparisons using Wilcoxon rank-sum tests. P-values (uncorrected for multiple comparisons) and Cohen’s d are given. Supplemental Table S3 provides an overview of the same analyses within the restricted sample and within females and males only. Significant findings (FDR corrected p-value < 0.05) are highlighted in bold.
