## Supplemental Table 3 for "Urinary steroid metabolome shows adrenal, gonadal, and neuroactive steroid dysregulation in adolescents with depression"

| **Supplemental Table S3.**  Sensitivity analysis: Group comparisons of steroid excretion rates (µg/24h) of summarized and single metabolites. | | | | | | | | | | | | |
| --- | --- | --- | --- | --- | --- | --- | --- | --- | --- | --- | --- | --- |
|  | Restricted Sample | | | | Females only | | | | Males only | | | |
|  | Controls: N = 50, Psychiatric Sample: N = 50 | | | | Controls: N = 64, Psychiatric Sample: N = 63 | | | | Controls: N = 11, Psychiatric Sample: N = 12 | | | |
|  | estimate | 95%-CI | t | p_raw_ | estimate | 95%-CI | t | p_raw_ | estimate | 95%-CI | t | p_raw_ |
| **Summarized metabolites** |  |  |  |  |  |  |  |  |  |  |  |  |
| Corticosterone metabolites  (allo-THB + THA + THB) | 240.1 | (109.75, 370.46) | 3.61 | <0.001 | 235.72 | (133.16, 338.28) | 4.50 | <0.001 | 757.51 | (310.24, 1204.77) | 3.32 | 0.004 |
| Major Cortisol metabolites  (allo-THF + THF + THE) | 35.12 | (-934.48, 1004.72) | 0.07 | 0.944 | 901.88 | (20.95, 1782.80) | 2.01 | 0.047 | 1035.09 | (-1897.11, 3967.28) | 0.69 | 0.497 |
| All Cortisol metabolites  (allo-THF + THF + THE + α-C + β-C + α-Cl +β-Cl) | -94.72 | (-1528.01, 1338.56) | -0.13 | 0.897 | 1106.85 | (-143.03, 2356.73) | 1.74 | 0.085 | 1098.41 | (-2793.98, 4990.80) | 0.55 | 0.587 |
| Major DHEA metabolites  (DHEA + 16α-OH-DHEA + A5T-16a) | 295.39 | (-52.09, 642.86) | 1.67 | 0.099 | 506.80 | (199.45, 814.14) | 3.23 | 0.002 | 372.93 | (-683.75, 1429.61) | 0.69 | 0.497 |
| 11-deoxygenated androgens  (DHEA + 16α-OH-DHEA + A5T-16a + A5-3β,17α + A5-3β,17β + An + Et) | 798.62 | (-198.33, 1795.57) | 1.57 | 0.120 | 1459.32 | (606.45, 2312.20) | 3.35 | 0.001 | 2291.61 | (-112.08, 4695.29) | 1.87 | 0.077 |
| 11-oxygenated androgens  (11-OH-An + 11-O-An + 11-OH-Et) | 162.78 | (-22.26, 347.81) | 1.72 | 0.088 | 250.63 | (84.31, 416.95) | 2.95 | 0.004 | 386.61 | (-97.65, 870.88) | 1.56 | 0.134 |
| Overall androgen metabolites (∑C19):  (11-deoxygenated androgens + 11-oxygenated androgens) | 1113.47 | (-21.35, 2248.30) | 1.92 | 0.057 | 1827.4 | (809.84, 2844.96) | 3.52 | 0.001 | 2715.87 | (192.79, 5238.95) | 2.11 | 0.048 |
| **progestagens (C21-steroids)** |  |  |  |  |  |  |  |  |  |  |  |  |
| P5D | 59.51 | (12.05, 106.97) | 2.46 | 0.016 | 68.32 | (33.66, 102.98) | 3.86 | <0.001 | 163.46 | (18.54, 308.38) | 2.21 | 0.040 |
| P5T-17α | 107.98 | (33.65, 182.31) | 2.85 | 0.005 | 129.22 | (74.65, 183.80) | 4.64 | <0.001 | 198.41 | (2.58, 394.23) | 1.99 | 0.062 |
| Po | 0.30 | (-15.00, 15.59) | 0.04 | 0.970 | 5.77 | (-8.11, 19.64) | 0.81 | 0.417 | 15.55 | (-10.20, 41.29) | 1.18 | 0.251 |
| aPo | -0.46 | (-5.89, 4.96) | -0.17 | 0.868 | 1.46 | (-2.61, 5.54) | 0.70 | 0.483 | 11.09 | (-3.12, 25.29) | 1.53 | 0.143 |
| PD | 16.61 | (-42.03, 75.24) | 0.56 | 0.580 | 40.64 | (-15.59, 96.88) | 1.42 | 0.159 | 116.59 | (-10.82, 244.00) | 1.79 | 0.089 |
| PT | 19.05 | (-97.92, 136.03) | 0.32 | 0.750 | 103.54 | (5.63, 201.46) | 2.07 | 0.040 | 405.73 | (117.35, 694.10) | 2.76 | 0.013 |
| Po-5β,3α | 27.18 | (-10.53, 64.89) | 1.41 | 0.161 | 34.03 | (3.53, 64.54) | 2.19 | 0.031 | 111.27 | (9.42, 213.12) | 2.14 | 0.045 |
| Po-5α,3α | -3.82 | (-8.45, 0.81) | -1.62 | 0.109 | -2.83 | (-6.01, 0.35) | -1.74 | 0.084 | 24.47 | (0.87, 48.08) | 2.03 | 0.056 |
| **mineralocorticoids (C21-steroids)** |  |  |  |  |  |  |  |  |  |  |  |  |
| TH-DOC | -4.55 | (-7.69, -1.42) | -2.85 | 0.005 | -2.98 | (-5.86, -0.10) | -2.03 | 0.044 | 1.68 | (-7.72, 11.07) | 0.35 | 0.730 |
| allo-THB | 108.88 | (32.74, 185.02) | 2.8 | 0.006 | 94.22 | (31.86, 156.57) | 2.96 | 0.004 | 444.57 | (196.05, 693.09) | 3.51 | 0.002 |
| THA | 32.17 | (3.05, 61.30) | 2.17 | 0.033 | 43.69 | (20.49, 66.88) | 3.69 | <0.001 | 91.96 | (-35.74, 219.66) | 1.41 | 0.174 |
| THB | 82.27 | (47.08, 117.45) | 4.58 | <0.001 | 71.05 | (44.51, 97.60) | 5.25 | <0.001 | 188.27 | (71.72, 304.82) | 3.17 | 0.005 |
| **glucocorticoid (C21-steroids)** |  |  |  |  |  |  |  |  |  |  |  |  |
| 11-O-PT | 0.39 | (-1.39, 2.17) | 0.43 | 0.670 | 1.07 | (-0.34, 2.49) | 1.49 | 0.140 | 7.55 | (-0.34, 15.43) | 1.88 | 0.076 |
| THS | 36.65 | (12.89, 60.42) | 3.02 | 0.003 | 35.68 | (16.05, 55.32) | 3.56 | 0.001 | 61.72 | (12.65, 110.80) | 2.47 | 0.023 |
| F | 9.59 | (-6.49, 25.66) | 1.17 | 0.245 | 16.83 | (0.64, 33.03) | 2.04 | 0.044 | 28.05 | (-11.23, 67.32) | 1.40 | 0.178 |
| 20α-DHF | 2.94 | (-8.98, 14.86) | 0.48 | 0.630 | 5.50 | (-6.02, 17.02) | 0.94 | 0.351 | 7.24 | (-18.84, 33.31) | 0.54 | 0.593 |
| 6β-OH-F | 115.97 | (76.46, 155.49) | 5.75 | <0.001 | 146.18 | (106.33, 186.04) | 7.19 | <0.001 | 181.57 | (72.27, 290.86) | 3.26 | 0.004 |
| allo-THF | -121.44 | (-316.41, 73.53) | -1.22 | 0.225 | -15.38 | (-167.25, 136.48) | -0.20 | 0.843 | 266.48 | (-447.22, 980.17) | 0.73 | 0.473 |
| THF | 132.03 | (-123.68, 387.74) | 1.01 | 0.314 | 365.16 | (150.73, 579.59) | 3.34 | 0.001 | 291.34 | (-429.94, 1012.62) | 0.79 | 0.438 |
| α-C | 18.00 | (-19.54, 55.54) | 0.94 | 0.350 | 32.33 | (0.95, 63.70) | 2.02 | 0.046 | 80.87 | (-16.28, 178.01) | 1.63 | 0.119 |
| β-C | 5.82 | (-85.08, 96.72) | 0.13 | 0.900 | 45.34 | (-30.51, 121.20) | 1.17 | 0.244 | 19.7 | (-261.10, 300.49) | 0.14 | 0.892 |
| THE | 2.23 | (-551.21, 555.67) | 0.01 | 0.994 | 582.73 | (53.37, 1112.08) | 2.16 | 0.033 | 328.65 | (-1290.51, 1947.80) | 0.40 | 0.695 |
| α-Cl | -129.40 | (-359.58, 100.78) | -1.10 | 0.273 | 66.66 | (-138.07, 271.39) | 0.64 | 0.525 | -49.17 | (-523.76, 425.43) | -0.20 | 0.841 |
| β-Cl | -89.44 | (-210.55, 31.66) | -1.45 | 0.151 | 25.68 | (-84.24, 135.60) | 0.46 | 0.648 | 45.64 | (-222.18, 313.46) | 0.33 | 0.742 |
| **androgens (C19-steroids)** |  |  |  |  |  |  |  |  |  |  |  |  |
| DHEA | 13.85 | (-60.02, 87.72) | 0.37 | 0.714 | 47.35 | (-0.22, 94.92) | 1.95 | 0.053 | 32.98 | (-303.98, 369.94) | 0.19 | 0.850 |
| 16α-OH-DHEA | 142.49 | (-11.14, 296.12) | 1.82 | 0.072 | 227.97 | (97.46, 358.49) | 3.42 | 0.001 | 169.78 | (-199.19, 538.74) | 0.90 | 0.378 |
| A5T*-*16α | 137.36 | (40.14, 234.58) | 2.77 | 0.007 | 159.6 | (70.03, 249.18) | 3.49 | 0.001 | 153.12 | (-83.52, 389.75) | 1.27 | 0.220 |
| A5-3β,17α | 23.40 | (9.29, 37.51) | 3.25 | 0.002 | 23.07 | (11.98, 34.15) | 4.08 | <0.001 | 60.97 | (24.04, 97.91) | 3.24 | 0.004 |
| A5-3β,17β | -4.60 | (-37.76, 28.55) | -0.27 | 0.786 | 9.14 | (-16.03, 34.30) | 0.71 | 0.478 | 54.02 | (-26.95, 135.00) | 1.31 | 0.207 |
| An | -30.80 | (-381.55, 319.95) | -0.17 | 0.864 | 181.00 | (-115.58, 477.58) | 1.20 | 0.234 | 825.00 | (-371.63, 2021.63) | 1.35 | 0.192 |
| Et | 258.57 | (-43.99, 561.13) | 1.68 | 0.097 | 488.74 | (222.35, 755.14) | 3.60 | <0.001 | 587.49 | (-181.21, 1356.19) | 1.50 | 0.151 |
| T | -0.29 | (-3.53, 2.95) | -0.18 | 0.861 | 0.89 | (-0.96, 2.74) | 0.94 | 0.348 | 4.31 | (-14.47, 23.08) | 0.45 | 0.658 |
| 5αAD-3α,17β | 0.06 | (-8.88, 8.99) | 0.01 | 0.990 | 4.79 | (-1.49, 11.07) | 1.49 | 0.138 | 46.8 | (-1.39, 94.98) | 1.90 | 0.072 |
| 11-OH-An | 75.52 | (-31.94, 182.97) | 1.38 | 0.172 | 112.74 | (16.55, 208.93) | 2.30 | 0.023 | 210.65 | (-152.31, 573.60) | 1.14 | 0.269 |
| 11-O-An | -5.10 | (-13.65, 3.45) | -1.17 | 0.245 | 0.44 | (-6.45, 7.32) | 0.12 | 0.902 | 21.92 | (-8.51, 52.35) | 1.41 | 0.174 |
| 11-OH-Et | 44.95 | (-38.56, 128.46) | 1.05 | 0.294 | 91.62 | (12.68, 170.55) | 2.27 | 0.025 | 76.77 | (-123.22, 276.76) | 0.75 | 0.461 |
| **estrogens (C18-steroids)** |  |  |  |  |  |  |  |  |  |  |  |  |
| E1 | -5.78 | (-9.41, -2.15) | -3.12 | 0.002 | -3.99 | (-7.56, -0.42) | -2.19 | 0.030 | -0.84 | (-7.37, 5.69) | -0.25 | 0.804 |
| E2 | -2.41 | (-3.24, -1.57) | -5.64 | <0.001 | -2.01 | (-2.90, -1.13) | -4.45 | <0.001 | -1.06 | (-2.95, 0.82) | -1.11 | 0.281 |
| E3 | -2.43 | (-5.06, 0.20) | -1.81 | 0.073 | -1.59 | (-4.21, 1.03) | -1.19 | 0.237 | -0.8 | (-6.01, 4.41) | -0.30 | 0.767 |

This table provides an overview of the following sensitivity analyses: Analyses within the restricted sample (i.e., excluding patients under psychotropic medication or eating disorders and controls using oral contraceptives as well as their respective matching partners), within females-only, and within males-only. The effect estimates for the group effect (psychiatric sample versus controls), their 95%-confidence intervals, and the t-values und p-values for the adjusted model (rank-based analysis of variance, adjusted for age and z-transformed BMI) are shown. For sensitivity analyses, the p-values were not corrected for multiple comparisons (p_raw_). Supplemental Table S2 provides an overview of the same analyses for the full sample.
