## Supplemental Table 4 for "Urinary steroid metabolome shows adrenal, gonadal, and neuroactive steroid dysregulation in adolescents with depression"

| **Supplemental Table S4.** Sensitivity analysis: Group comparisons of metabolite ratios as measures of enzyme activity. | | | | | | | | | | | | | |
| --- | --- | --- | --- | --- | --- | --- | --- | --- | --- | --- | --- | --- | --- |
|  | | Restricted Sample | | | | Females only | | | | Males only | | | |
|  | | (Controls: N = 50, Psychiatric Sample: N = 50) | | | | (Controls: N = 64, Psychiatric Sample: N = 63) | | | | (Controls: N = 11, Psychiatric Sample: N = 12) | | | |
|  | | estimate | 95%-CI | t | p_raw_ | estimate | 95%-CI | t | p_raw_ | estimate | 95%-CI | t | p_raw_ |
| **Relative neuroactive steroid production** | |  |  |  |  |  |  |  |  |  |  |  |  |
| aPo / PD | | 0.00 | (-0.01, 0.01) | -0.48 | 0.632 | 0.00 | (-0.02, 0.01) | -0.79 | 0.432 | -0.02 | (-0.07, 0.03) | -0.85 | 0.406 |
| aPo / Po + PD | | 0.00 | (-0.01, 0.01) | -0.66 | 0.513 | 0.00 | (-0.01, 0.00) | -0.94 | 0.347 | -0.02 | (-0.06, 0.02) | -0.99 | 0.335 |
| Po / PD | | 0.01 | (-0.03, 0.04) | 0.37 | 0.713 | 0.01 | (-0.03, 0.04) | 0.42 | 0.674 | -0.01 | (-0.09, 0.07) | -0.24 | 0.817 |
| Po / aPo + PD | | 0.01 | (-0.03, 0.04) | 0.34 | 0.735 | 0.01 | (-0.02, 0.04) | 0.49 | 0.628 | -0.01 | (-0.07, 0.06) | -0.19 | 0.853 |
| 5αAD-3α,17β / overall androgen metabolites | | 0.00 | (-0.00, 0.00) | -1.59 | 0.116 | 0.00 | (-0.00, -0.00) | -3.13 | 0.002 | 0.00 | (-0.01, 0.00) | -0.99 | 0.336 |
| 5αAD-3α,17β / 11-deoxygenated androgens | | 0.00 | (-0.00, 0.00) | -1.24 | 0.219 | 0.00 | (-0.00, -0.00) | -2.94 | 0.004 | 0.00 | (-0.01, 0.00) | -0.73 | 0.477 |
| **Relative overall androgen production** | |  |  |  |  |  |  |  |  |  |  |  |  |
|  | 11-deoxygenated androgens / Major Cortisol metabolites | 0.11 | (-0.11, 0.33) | 1 | 0.321 | 0.10 | (-0.09, 0.29) | 1.00 | 0.318 | 0.17 | (-0.18, 0.51) | 0.95 | 0.353 |
|  | (An + Et) / Major Cortisol metabolites | 0.04 | (-0.10, 0.18) | 0.54 | 0.588 | 0.02 | (-0.11, 0.15) | 0.31 | 0.760 | 0.16 | (-0.10, 0.41) | 1.21 | 0.240 |
| **Relative adrenal androgen production** | |  |  |  |  |  |  |  |  |  |  |  |  |
|  | Major DHEA metabolites / Major Cortisol metabolites | 0.05 | (-0.02, 0.12) | 1.39 | 0.169 | 0.06 | (-0.00, 0.13) | 1.84 | 0.068 | 0.04 | (-0.09, 0.17) | 0.57 | 0.578 |
|  | DHEA / Major Cortisol metabolites | 0.00 | (-0.01, 0.01) | 0.03 | 0.976 | 0.00 | (-0.01, 0.01) | 0.72 | 0.471 | 0.00 | (-0.06, 0.06) | 0.06 | 0.951 |
| **Ratio 11-oxygenated / 11-deoxygenated androgens** | |  |  |  |  |  |  |  |  |  |  |  |  |
|  | 11-oxygenated androgens / 11-deoxygenated androgens | 0.02 | (-0.02, 0.06) | 1.11 | 0.268 | 0.03 | (-0.01, 0.06) | 1.50 | 0.137 | 0.03 | (-0.03, 0.08) | 0.87 | 0.393 |
|  | 11-OH-Et / Major DHEA metabolites | -0.01 | (-0.15, 0.13) | -0.11 | 0.912 | -0.02 | (-0.15, 0.11) | -0.29 | 0.770 | 0.05 | (-0.24, 0.33) | 0.33 | 0.745 |
| **5α-reductase activity** | |  |  |  |  |  |  |  |  |  |  |  |  |
|  | (An + 11OH-An + a-THF + a-THB + Po-5α,3α) / (Et + 11OH-Et + THF + THB + Po-5β,3α) | -0.18 | (-0.33, -0.02) | -2.24 | 0.027 | -0.19 | (-0.32, -0.06) | -2.78 | 0.006 | -0.18 | (-0.64, 0.28) | -0.76 | 0.459 |
|  | Po-5α,3α / Po-5β,3α | -0.04 | (-0.07, -0.02) | -3.40 | 0.001 | -0.05 | (-0.08, -0.03) | -4.21 | <0.001 | -0.03 | (-0.12, 0.06) | -0.58 | 0.570 |
|  | allo-THB / THB | -0.31 | (-0.63, 0.01) | -1.93 | 0.057 | -0.28 | (-0.55, -0.01) | -2.03 | 0.044 | -0.38 | (-1.16, 0.39) | -0.97 | 0.345 |
|  | allo-THF / THF | -0.19 | (-0.34, -0.04) | -2.51 | 0.014 | -0.19 | (-0.30, -0.07) | -3.18 | 0.002 | -0.17 | (-0.61, 0.27) | -0.75 | 0.463 |
|  | 11β-OH-An / 11β-OH-Et | 0.11 | (-0.75, 0.97) | 0.25 | 0.800 | -0.01 | (-0.63, 0.60) | -0.04 | 0.969 | 0.42 | (-2.47, 3.31) | 0.28 | 0.78 |
|  | An / Et | -0.25 | (-0.47, -0.02) | -2.15 | 0.034 | -0.28 | (-0.46, -0.10) | -3.08 | 0.003 | -0.38 | (-1.03, 0.26) | -1.17 | 0.258 |
| **11β-hydroxy-steroiddehydrogenase (11β-HSD) activity** | |  |  |  |  |  |  |  |  |  |  |  |  |
|  | (α-C +β-C) / (α-Cl +β-Cl) | 0.05 | (0.01, 0.09) | 2.48 | 0.015 | 0.03 | (-0.00, 0.06) | 1.74 | 0.084 | 0.04 | (-0.04, 0.12) | 0.95 | 0.356 |
|  | THF / THE | 0.03 | (-0.01, 0.07) | 1.29 | 0.202 | 0.01 | (-0.03, 0.05) | 0.38 | 0.702 | 0.04 | (-0.06, 0.14) | 0.79 | 0.442 |
|  | (allo-THF + THF) / THE | -0.02 | (-0.09, 0.05) | -0.62 | 0.537 | -0.05 | (-0.12, 0.01) | -1.59 | 0.114 | 0.05 | (-0.09, 0.20) | 0.74 | 0.471 |
| **3β-hydroxysteroiddehydrogenase (3β-HSD) activity** | |  |  |  |  |  |  |  |  |  |  |  |  |
|  | Major DHEA metabolites / Major Cortisol metabolites | 0.05 | (-0.02, 0.12) | 1.39 | 0.169 | 0.06 | (-0.00, 0.13) | 1.84 | 0.068 | 0.04 | (-0.09, 0.17) | 0.57 | 0.578 |
|  | Major DHEA metabolites / (An + Et) | 0.06 | (-0.04, 0.15) | 1.15 | 0.253 | 0.10 | (0.01, 0.18) | 2.26 | 0.026 | -0.06 | (-0.27, 0.16) | -0.52 | 0.611 |
|  | DHEA / (An + Et) | 0.00 | (-0.02, 0.02) | -0.12 | 0.904 | 0.01 | (-0.01, 0.02) | 0.80 | 0.425 | 0.00 | (-0.12, 0.11) | -0.04 | 0.969 |
|  | P5T-17α / PT | 0.15 | (0.04, 0.26) | 2.58 | 0.012 | 0.15 | (0.06, 0.24) | 3.43 | 0.001 | 0.02 | (-0.26, 0.31) | 0.17 | 0.867 |
|  | P5T-17α / Major Cortisol metabolites | 0.02 | (0.01, 0.03) | 3.32 | 0.001 | 0.02 | (0.01, 0.03) | 3.33 | 0.001 | 0.03 | (0.00, 0.05) | 2.28 | 0.034 |
| **21-hydroxylase-activity** | |  |  |  |  |  |  |  |  |  |  |  |  |
|  | 11-O-PT / THE | 0.00 | (-0.00, 0.00) | 1.13 | 0.260 | 0.00 | (-0.00, 0.00) | -0.44 | 0.664 | 0.00 | (0.00, 0.01) | 3.26 | 0.004 |
|  | 11-O-PT / Major Cortisol metabolites | 0.00 | (-0.00, 0.00) | 1.56 | 0.122 | 0.00 | (-0.00, 0.00) | -0.10 | 0.924 | 0.00 | (0.00, 0.00) | 3.08 | 0.006 |
|  | Po-5β,3α / THE | 0.01 | (-0.00, 0.02) | 1.32 | 0.190 | 0.00 | (-0.01, 0.01) | 0.02 | 0.988 | 0.03 | (0.01, 0.06) | 2.41 | 0.027 |
|  | 11-O-PT / α-Cl | 0.00 | (0.00, 0.00) | 2.38 | 0.019 | 0.00 | (-0.00, 0.00) | 1.84 | 0.068 | 0.01 | (0.00, 0.01) | 2.94 | 0.008 |
|  | (PT + Po-5α,3α + Po-5β,3α) / Major Cortisol metabolites | 0.01 | (-0.02, 0.04) | 0.80 | 0.423 | 0.00 | (-0.02, 0.03) | 0.29 | 0.773 | 0.07 | (-0.03, 0.16) | 1.40 | 0.178 |
|  | (11-O-PT + PT + Po-5α,3α + Po-5β,3α) / Major Cortisol metabolites | 0.01 | (-0.02, 0.04) | 0.81 | 0.418 | 0.00 | (-0.02, 0.03) | 0.27 | 0.787 | 0.07 | (-0.02, 0.16) | 1.46 | 0.162 |
|  | PD / TH-DOC | 3.25 | (0.22, 6.28) | 2.10 | 0.038 | 3.64 | (0.51, 6.78) | 2.28 | 0.025 | 4.68 | (-0.14, 9.50) | 1.90 | 0.072 |
| **17β-hydroxysteroiddehydrogenase (****17β-HSD) activity** | |  |  |  |  |  |  |  |  |  |  |  |  |
|  | A5-3β,17β / DHEA | -0.10 | (-0.33, 0.13) | -0.87 | 0.387 | -0.19 | (-0.38, -0.01) | -2.06 | 0.041 | -0.06 | (-0.72, 0.60) | -0.18 | 0.859 |
|  | A5-3β,17β / 11-OH-An | -0.06 | (-0.15, 0.02) | -1.48 | 0.141 | -0.05 | (-0.13, 0.02) | -1.53 | 0.129 | -0.11 | (-0.36, 0.13) | -0.93 | 0.366 |
| **11β-hydroxylase activity** | |  |  |  |  |  |  |  |  |  |  |  |  |
|  | THS / Major Cortisol metabolites | 0.01 | (0.00, 0.01) | 4.18 | <0.001 | 0.00 | (0.00, 0.01) | 2.41 | 0.017 | 0.01 | (0.00, 0.01) | 2.35 | 0.03 |
|  | (An + Et) / 11-OH-An + 11-OH-Et | -0.85 | (-1.64, -0.07) | -2.14 | 0.035 | -0.94 | (-1.66, -0.21) | -2.52 | 0.013 | -0.26 | (-2.53, 2.01) | -0.23 | 0.824 |
|  | TH-DOC / Corticosterone metabolites | -0.03 | (-0.04, -0.02) | -5.50 | <0.001 | -0.03 | (-0.04, -0.02) | -6.14 | <0.001 | -0.02 | (-0.04, -0.01) | -2.73 | 0.013 |
| **17-hydroxylase/17,20-lyase activity** | |  |  |  |  |  |  |  |  |  |  |  |  |
|  | PD / (An + Et) | 0.00 | (-0.02, 0.02) | 0.27 | 0.784 | -0.01 | (-0.02, 0.01) | -0.79 | 0.429 | 0.01 | (-0.00, 0.03) | 1.56 | 0.134 |
|  | Corticosterone metabolites / (An + Et) | 0.05 | (0.01, 0.09) | 2.40 | 0.018 | 0.03 | (-0.01, 0.07) | 1.54 | 0.125 | 0.12 | (0.05, 0.18) | 3.49 | 0.002 |
|  | P5D / P5T-17α | -0.07 | (-0.26, 0.11) | -0.76 | 0.449 | -0.12 | (-0.28, 0.04) | -1.51 | 0.135 | -0.07 | (-0.46, 0.32) | -0.34 | 0.734 |
|  | PD / PT | 0.00 | (-0.09, 0.09) | 0.02 | 0.987 | -0.05 | (-0.14, 0.03) | -1.30 | 0.195 | 0.03 | (-0.07, 0.13) | 0.55 | 0.587 |
|  | Corticosterone metabolites / Major cortisol metabolites | 0.04 | (0.02, 0.06) | 4.51 | <0.001 | 0.02 | (0.01, 0.04) | 2.79 | 0.006 | 0.10 | (0.06, 0.13) | 5.31 | <0.001 |
|  | P5T-17α / A5-3β,17β | 0.84 | (0.38, 1.31) | 3.57 | 0.001 | 1.00 | (0.56, 1.44) | 4.48 | <0.001 | 0.79 | (0.18, 1.41) | 2.52 | 0.021 |
|  | PT / (An + Et) | 0.00 | (-0.03, 0.03) | -0.07 | 0.941 | 0.00 | (-0.03, 0.03) | -0.03 | 0.973 | 0.03 | (-0.04, 0.10) | 0.87 | 0.397 |
|  | (PT + Po-5α,3α + Po-5β,3α) / (An + Et) | 0.00 | (-0.04, 0.04) | 0.14 | 0.892 | 0.00 | (-0.04, 0.03) | -0.21 | 0.837 | 0.04 | (-0.05, 0.13) | 0.89 | 0.385 |
|  | Major cortisol metabolites / (An + Et) | -0.05 | (-0.36, 0.26) | -0.30 | 0.764 | -0.03 | (-0.34, 0.27) | -0.22 | 0.824 | -0.30 | (-0.88, 0.27) | -1.03 | 0.315 |
| **P450-Oxidoreductase (POR) activity** | |  |  |  |  |  |  |  |  |  |  |  |  |
|  | PD / THE | 0.00 | (-0.02, 0.02) | 0.15 | 0.877 | -0.01 | (-0.03, 0.01) | -0.86 | 0.390 | 0.03 | (-0.01, 0.07) | 1.51 | 0.147 |
|  | PD / Major cortisol metabolites | 0.00 | (-0.01, 0.02) | 0.41 | 0.686 | 0.00 | (-0.02, 0.01) | -0.57 | 0.569 | 0.02 | (-0.00, 0.04) | 1.74 | 0.098 |
| **Aromatase activity** | |  |  |  |  |  |  |  |  |  |  |  |  |
|  | (E1 + E2 + E3) / A5-3β,17β | -0.07 | (-0.15, 0.01) | -1.81 | 0.074 | -0.10 | (-0.19, -0.00) | -2.01 | 0.046 | -0.02 | (-0.11, 0.06) | -0.51 | 0.618 |
| **CYP3A4 activity** | |  |  |  |  |  |  |  |  |  |  |  |  |
|  | 6β-OH-F / F | 1.65 | (1.30, 1.99) | 9.42 | <0.001 | 1.62 | (1.32, 1.92) | 10.57 | <0.001 | 1.24 | (0.54, 1.95) | 3.46 | 0.003 |
|  | 6β-OH-F / THE + α-Cl +β-Cl | 0.02 | (0.02, 0.03) | 6.68 | <0.001 | 0.02 | (0.01, 0.03) | 5.52 | <0.001 | 0.03 | (0.02, 0.05) | 3.66 | 0.002 |
| **20α-HSD (AKR1C1) activity** | |  |  |  |  |  |  |  |  |  |  |  |  |
|  | α-Cl / β-Cl | -0.01 | (-0.25, 0.24) | -0.08 | 0.938 | -0.11 | (-0.35, 0.13) | -0.91 | 0.364 | 0.18 | (-0.38, 0.74) | 0.64 | 0.533 |
|  | 20α-DHF / F | -0.07 | (-0.13, -0.02) | -2.57 | 0.012 | -0.07 | (-0.13, -0.01) | -2.40 | 0.018 | -0.09 | (-0.21, 0.04) | -1.39 | 0.181 |

Overview of the sensitivity analyses, including analyses within the restricted sample (excluding patients on psychotropic medication or eating disorders, and controls using oral contraceptives, as well as their respective matching partners), within females-only, and males-only. The table reports effect estimates for the group effect (psychiatric sample versus controls), their 95%-confidence intervals, the t-values and p-values for the adjusted model (rank-based analysis of variance, adjusted for age and z-transformed BMI) are shown. For sensitivity analyses, p-values were not corrected for multiple comparisons (p_raw_). Table 3 provides an overview of the same analyses within the full sample.
