## Supplemental Table 5 for "Urinary steroid metabolome shows adrenal, gonadal, and neuroactive steroid dysregulation in adolescents with depression"

| **Supplemental Table S5.** Sensitivity analysis: Spearman rank correlation with storage time. | | |
| --- | --- | --- |
|  | Rank correlation | |
|  | 𝜌 | p-value |
| **progestagens (C21-steroids)** |  |  |
| P5D | -0.035 | 0.668 |
| P5T-17α | -0.068 | 0.408 |
| Po | 0.152 | 0.064 |
| aPo | 0.137 | 0.094 |
| PD | 0.080 | 0.330 |
| PT | 0.038 | 0.642 |
| Po-5β,3α | 0.047 | 0.567 |
| Po-5α,3α | 0.097 | 0.236 |
| **mineralocorticoids (C21-steroids)** | |  |
| TH-DOC | 0.142 | 0.082 |
| allo-THB | -0.043 | 0.598 |
| THA | -0.004 | 0.960 |
| THB | -0.017 | 0.832 |
| **glucocorticoid (C21-steroids)** |  |  |
| 11-O-PT | 0.023 | 0.778 |
| THS | -0.058 | 0.478 |
| F | 0.024 | 0.771 |
| 20α-DHF | 0.053 | 0.521 |
| 6β-OH-F | -0.320 | ≤0.001 |
| allo-THF | 0.028 | 0.734 |
| THF | 0.002 | 0.984 |
| α-C | -0.040 | 0.627 |
| β-C | 0.040 | 0.624 |
| THE | -0.048 | 0.556 |
| α-Cl | -0.034 | 0.677 |
| β-Cl | 0.038 | 0.647 |
| **androgens (C19-steroids)** |  |  |
| DHEA | 0.005 | 0.949 |
| 16α-OH-DHEA | -0.017 | 0.836 |
| A5T*-*16α | -0.025 | 0.759 |
| A5-3β,17α | ≤0.000 | 0.996 |
| A5-3β,17β | 0.047 | 0.567 |
| An | 0.074 | 0.369 |
| Et | 0.054 | 0.514 |
| T | 0.074 | 0.370 |
| 5αAD-3α,17β | 0.043 | 0.602 |
| 11-OH-An | -0.001 | 0.993 |
| 11-O-An | -0.067 | 0.412 |
| 11-OH-Et | -0.124 | 0.132 |
| **estrogens (C18-steroids)** |  |  |
| E1 | 0.126 | 0.125 |
| E2 | 0.147 | 0.072 |
| E3 | 0.053 | 0.521 |

Spearman's rank correlation coefficient (two-tailed) between the storage time (in days) and the daily excretion rates of the specified steroids (in µg/24h).
